# Protocol for NAC Attack, a phase-3, multicenter randomized, parallel, double masked, placebo controlled trial evaluating the efficacy and safety of oral N-acetylcysteine (NAC) in patients with retinitis pigmentosa

**DOI:** 10.1101/2025.11.05.25339486

**Authors:** Xiangrong Kong, Folahan Ibukun, Muhammad J. Khan, Gulnar Hafiz, Dagmar Wehling, Kurt Dreger, Mandeep Singh, David Birch, Glenn Jaffe, Fahd Naufal, Elias I. Traboulsi, Jacque Duncan, Rob Hufnagel, Joseph Carroll, Yuchen Lu, Rodrigo Matsui, Lawrence H. Moulton, Barbara Hawkins, Peter A. Campochiaro, the NAC Attack Study Investigative Group

**Affiliations:** Wilmer Eye Institute, School of Medicine, Johns Hopkins University, Baltimore, MD; 2Department of Biostatistics, Bloomberg School of Public Health, Johns Hopkins University, Baltimore, MD; Department of Epidemiology, Bloomberg School of Public Health, Johns Hopkins University, Baltimore, MD; Department of Health Behavior and Society, Bloomberg School of Public Health, Johns Hopkins University, Baltimore, MD; Retina Foundation of the Southwest, Dallas, TX; Department of Ophthalmology, Duke University, Durham, NC; Department of Ophthalmology, University of California San Francisco, San Francisco, CA; Cole Eye Institute, Cleveland Clinic, Cleveland, OH; Center for Integrated Healthcare Research, Kaiser Permanente Hawaii. Honolulu, HI; Department of Ophthalmology and Visual Sciences, Medical College of Wisconsin, Milwaukee, WI; School of Medicine and Dentistry, University of Rochester, Rochester, NY; Fundación de Asistencia Privada Conde de Valenciana I.A. P, CDMX, México.; Department of International Health, Bloomberg School of Public Health, Johns Hopkins University, Baltimore, MD; Department of Neuroscience, School of Medicine, Johns Hopkins University

**Keywords:** Inherited retinal disease, antioxidant, oxidative stress, estimand framework, outcome measure selection

## Abstract

**Background:** Retinitis pigmentosa (RP) is the most common inherited retinal disease. Genetic mutations of many genes have been linked to RP. The mutations cause rod photoreceptor degeneration while sparing cone photoreceptors. However, loss of rod photoreceptors results in oxidative stress leading to cone photoreceptors degeneration, causing constriction of visual fields. In animal models of RP, treatments that reduce oxidative stress, including N-acetylcysteine (NAC), promote cone survival and maintenance of function.

**Methods:** NAC Attack, funded by the US National Institutes of Health, is a multicenter, randomized, double-masked, parallel and placebo-controlled clinical trial testing whether oral NAC can delay disease progression in RP. A total of about 483 RP patients aged 18-65 are recruited from 31 sites in America and Europe, and randomized 2:1 to take NAC 1800 mg bid or placebo for 45 months. Eligible eyes have best-corrected visual acuity (BCVA) of 20/63 or better and the ellipsoid zone (EZ) width on the horizontal fovea optical coherence tomography (OCT) scan between 1500 µm and 8000 µm. The primary outcome is the cumulative loss of EZ, calculated as the area above the curve using EZ width measured every 9 months over 45 months. Secondary outcomes are change in mean macular sensitivity (MMS) measured by microperimetry and change in BCVA best-corrected visual acuity (BCVA) from baseline to M45. The long-term safety and tolerability of oral NAC 1800 mg bid will be assessed based upon the incidence and severity of ocular and systemic adverse events.

**Discussion:** Data from NAC Attack will provide clinical evidence (or lack of) on the role of oxidative stress in human RP. Findings from NAC Attack have the potential to change clinical management of a blinding disease and will lead to a better understanding of basic mechanisms in RP. Moreover, data from NAC Attack will advance our knowledge on safety and drug interaction of NAC, informing studies in other medical areas where NAC is of interest for its potential in treating many medical conditions. Therefore, the trial is of high public health, clinical, and scientific significances.

**Trial registration:** The ClinicalTrials.gov ID for NAC Attack is NCT05537220.

## BACKGROUND

Retinitis pigmentosa (RP) is the most common inherited retinal disease and occurs in roughly 1/4000 individuals. Inheritance pattern may be autosomal recessive, autosomal dominant, X-linked recessive, or simplex (only one family member affected).[1–3] Disease is usually limited to the eye, but in 20-30% of RP patients, retinal degeneration in association with non-ocular disease is part of a syndrome. Usher syndrome, in which there is hearing loss in addition to retinal degeneration, is particularly common accounting for 10-20% of RP cases.[4] A common feature of the many different mutations that cause RP is that they differentially cause damage and death of rod photoreceptors while having little or no effect on cone photoreceptors. In some cases, this is because the disease gene is only expressed in rods, which is the case for *Rhodopsin*.[5] But mutations in some ubiquitously expressed genes differentially affect rods and cause RP. For instance, mutations in one allele of genes that code for pre-mRNA processing factors (PRPFs) cause dominantly inherited RP due to haploinsufficiency, because rod photoreceptors have a specific splicing program needed for ciliogenesis and are less tolerant of reduced levels of PRPFs than other cell types.[6] Thus, the highly specialized structure and abnormally high metabolic demands of rod photoreceptors may make them more susceptible than other cells to damage and death from reduced function of a variety of gene products underlying the surprising number and types of RP disease genes.

The loss of rod photoreceptors results in poor vision in dim illumination (night blindness), but does not initially affect activities of daily life such as reading or driving. However, after most of the rod photoreceptors are eliminated, cone photoreceptors begin to die. The drop out of cones usually begins in the mid-periphery of the retina and extends peripherally and centrally. At this stage, patients have mid-peripheral scotomata that coalesce into a ring which gradually enlarges and patients may complain that things or people in the environment may disappear and pop into view as images pass through the scotomata. As cone cell death continues to progress more centrally, patients become aware of constriction of visual fields. As the visual fields constrict, the level of disability increases but visual acuity is maintained until late.

Rod and cone photoreceptors are the only cell types in the outer retina and rods greatly out number cones with a ratio of 95:5. After rods die from a mutation, oxygen consumption in the outer retina is markedly reduced, but oxygen supply from the choroid is unchanged so that the remaining cones are exposed to very high levels of tissue oxygen.[7] The high tissue oxygen results in generation of superoxide radicals[8] initiating a cascade of production of reactive oxygen species (ROS) and reactive nitrogen species (RNS) that cause progressive damage to cones.[9, 10] In animal models of RP, reduction of oxidative stress by administration of antioxidants [11, 12] or altered expression of components of the antioxidant defense system,[13, 14] promotes cone survival and maintenance of cone function. The hostile environment of the outer retina in RP induces metabolic abnormalities in cones that may also contribute to cone cell death,[15, 16] but it is clear that oxidative damage plays an important role.[17]

N-acetylcysteine (NAC) is a derivative of L-cysteine that neutralizes ROS and RNS itself, but is also converted to cysteine which is used to biosynthesize glutathione, a major component of the endogenous antioxidant defense system.[18] When given in a timely manner, NAC is a life-saving treatment for acetaminophen overdose because it prevents severe oxidative damage from aldehydes formed in the liver during metabolic breakdown of acetaminophen.[19, 20] Orally administered NAC slows cone cell death and promotes maintenance of function in an animal model of RP.[21] The FIGHT-RP study was a dose-ranging study in 30 patients with RP given oral NAC in doses ranging from 600mg bid to 1800mg tid. The study showed that good intraocular levels of NAC were obtained with doses of 1200 mg bid or higher, a maximum tolerated dose of 1800 mg bid and there were statistically significant improvements of BCVA and macular sensitivity during the 24 weeks of treatment. These encouraging results after 24 weeks treatment with NAC raised the question of whether long term treatment with NAC could slow progression of retinal degeneration in patients with RP. The NAC Attack clinical trial was designed to answer that question

NAC Attack is a double-blinded, randomized, placebo-controlled parallel clinical trial to evaluate the efficacy and safety of oral NAC 1800mg bid in delaying disease progression in RP. The primary outcome measure is the cumulative loss of ellipsoid zone (EZ) width measured on horizontal spectral domain-optical coherence tomography (SD-OCT) scans through the fovea obtained every 9 months for 45 months. Secondary outcomes are functional outcome measures: 1) change in BCVA between baseline and month (M) 45 measured by the protocol of the Early Treatment Diabetic Retinopathy Study (ETDRS) protocol[22], and 2) change in mean macular sensitivity measured by microperimetry between baseline and M45.

Recent significant advances in gene therapies would only apply to very specific genotypes in IRDs. NAC Attack, on the other hand, tests a pharmacological approach that has the potential to promote cone survival and that does not depend on the knowledge of specific genetic variants. If proven efficacious, the trial will lead to a general therapy benefitting patients with a clinical diagnosis of RP, irrespective of the identification of pathogenic variant(s). Thus, NAC Attack has the potential to impact the clinical management of RP. It will also provide clinical evidence (or lack of evidence) elucidating the role of oxidative stress in cone cell death in RP, leading to a better understanding of the molecular mechanism of photoreceptor degeneration in human RP. Moreover, data from NAC Attack will advance our knowledge on the safety and drug interaction of NAC and inform studies in other medical areas where NAC is of interest for its potential in treating many medical conditions.

## METHODS AND DESIGN

The NAC Attack trial is registered at ClinicalTrials.gov (NCT05537220). The SPIRIT checklist for reporting clinical trial information is also available as Supplemental Material [S1 checklist]. [23] The complete protocol is available as Supplemental Material [S2 protocol]. Patient orienting videos can be found at the trial website at https://www.hopkinsmedicine.org/wilmer/research/nac-attack. The study was approved by the following competent authorities and Institutional Review Board (IRB) or Research Ethics Committees: in the United States (US)-Johns Hopkins University single IRB (sIRB) with reliance agreements of the local IRBs of participating sites in the USA and the US Federal Drug and Food Administration; in Austria, Germany and the Netherlands: the European Medicines Agency; in Canada: Health Canada and Research Ethics Board of McGill University Health Center; in Switzerland: Swissmedic and Swissthics; and in the United Kingdom: the Medicines and Healthcare products Regulatory Agency and the Health Research Authority.

### AIM OF THE STUDY

The aim of the study is to determine if orally administered NAC, 1800 mg bid, can slow the progression of retinal degeneration in patients with RP and has acceptable safety and tolerability.

### DESIGN AND SETTING OF THE STUDY

NAC Attack is an international, multicenter, randomized, double-masked, parallel, and placebo-controlled trial. It is funded by the National Institutes of Health (NIH) of the US through cooperative grants awarded to study Principal Investigators at the Johns Hopkins University (JHU), Duke University, and University of California San Francisco. The Study Leadership include the Study Chairman (JHU), Coordinating Center Director (JHU) and NEI Representatives. About 483 eligible patients with RP are randomized in a ratio of 2:1 to receive NAC effervescent tablets 1800mg bid or matching placebo control and are followed at their enrolling sites for 45 months. A list of participating sites can be found at ClinicalTrials.gov (NCT05537220). Figure 1 shows the Spirit schedule of enrollment, interventions and assessments. Figure 2 shows the Schematic of NAC Attack Study Visits including in-clinic study visits and protocol specified virtual visits with site Investigator and phone calls with site Coordinator.

**Figure 1.**
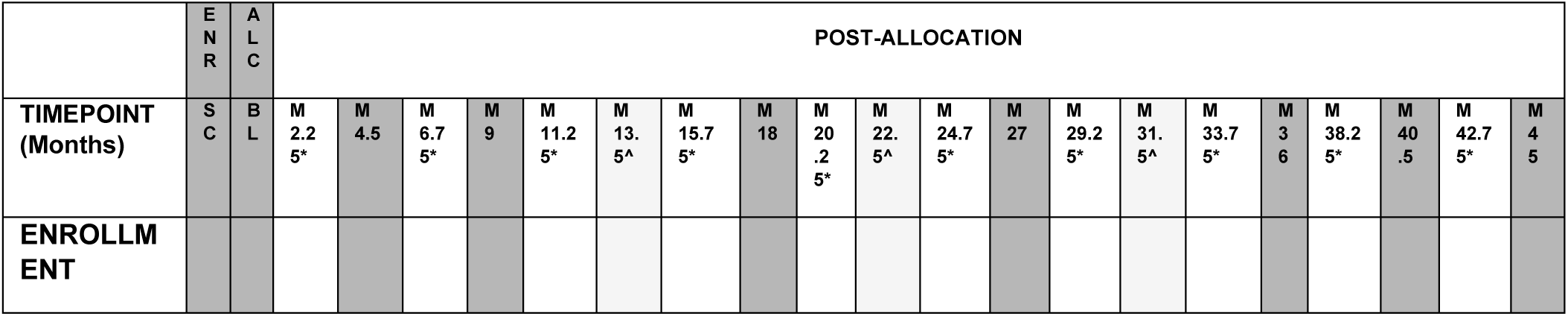
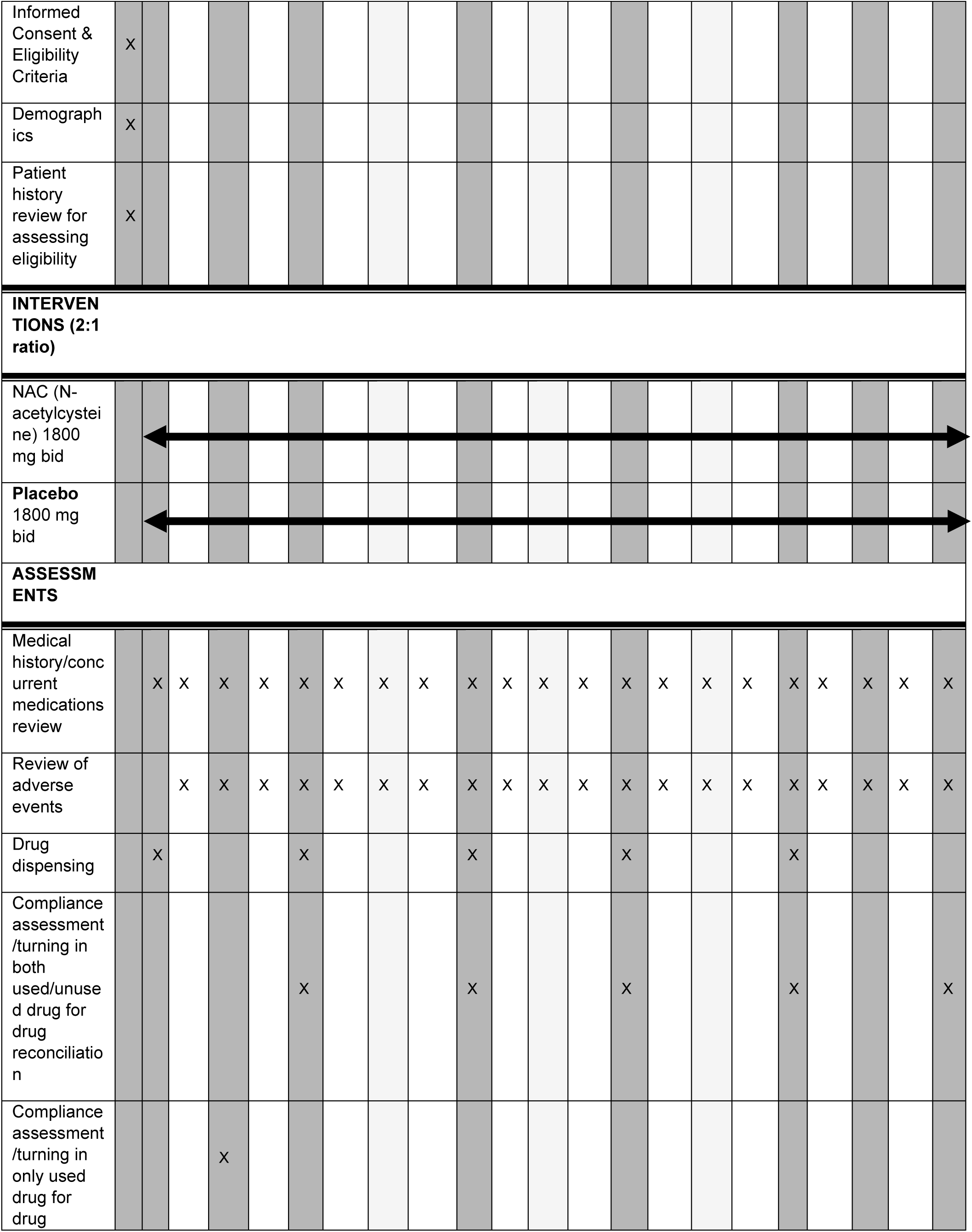
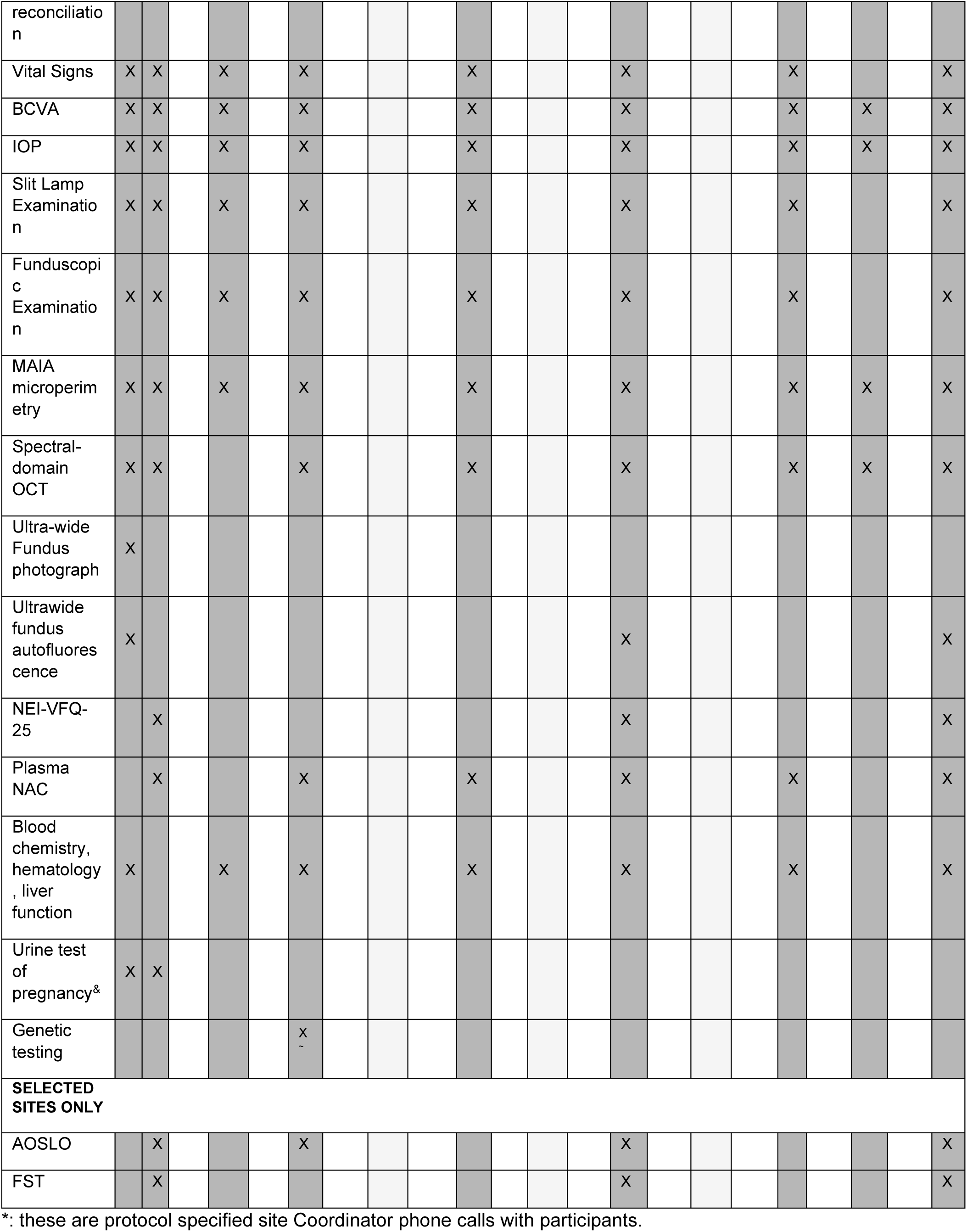
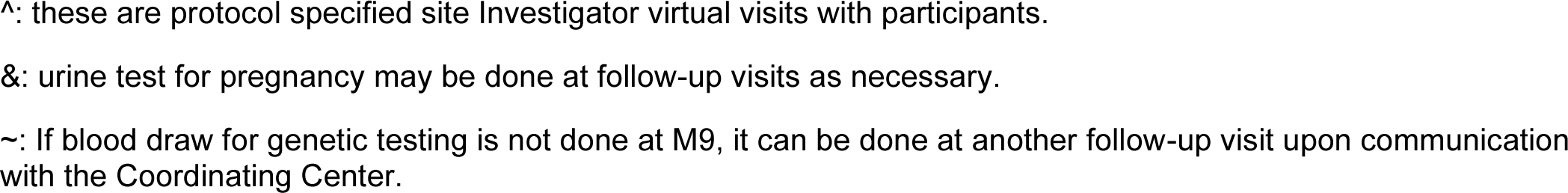
SPIRIT schedule of enrollment, interventions, and assessments of NAC Attack. ENR: Enrollment; ALC: Allocation. SC: Screening visit. BL: Baseline visit including randomization BCVA, best-corrected visual acuity; SD-OCT, spectral domain-optical coherence tomography of the retina; FST, full-field stimulus threshold; NEI-VFQ, National Eye Institute Visual Function Questionnaire; AOSLO. Adaptive optics scanning laser ophthalmoscopy.

**Figure 2.**
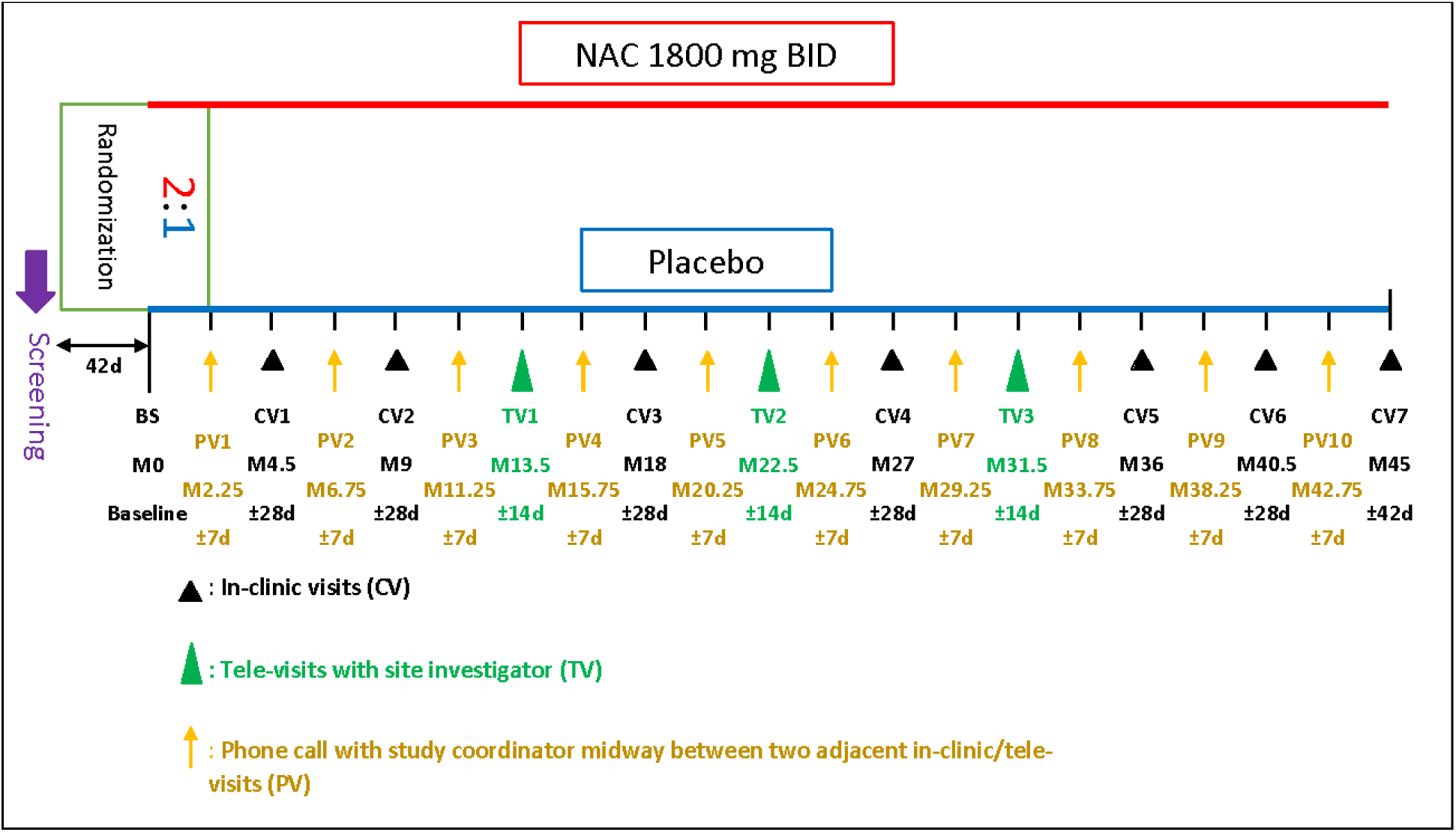
Schematic of NAC Attack Study Visits. Each participant will be followed for 45 months after randomization. Study visits including in-clinic visits, tele-visits with site Investigator, and phone calls with site Coordinator. BS: Baseline. d: days.

### STUDY ORGANIZATIONAL STRUCTURE

#### Study Sites and Other Entities

The study is funded by the National Eye Institute (NEI) of the NIH through 4 UG1 cooperative agreement grants, including a grant funding Study Chair’s Office (UG1EY033286, Wilmer Eye Institute of the School of Medicine of Johns Hopkins University, Maryland, USA), a Coordinating Center grant (UG1EY033293, Wilmer Eye Institute of the School of Medicine of Johns Hopkins University, Maryland, USA), and resource center grants for the OCT Reading Center (UG1EY033287 Duke Eye Center of

Duke University, North Carolina, USA), and the AOSLO Reading Center (UG1EY033292 Department of Ophthalmology, University of California San Francisco and Department of Ophthalmology and Visual Sciences, Medical University of Wisconsin, USA). Patients are enrolled from 31 Clinical Sites of 7 countries (**Figure *3***), including 24 sites in the US, 1 site each in Canada, Austria, Germany, the United Kingdom and Switzerland, and 2 sites in the Netherlands.

**Figure 3.**
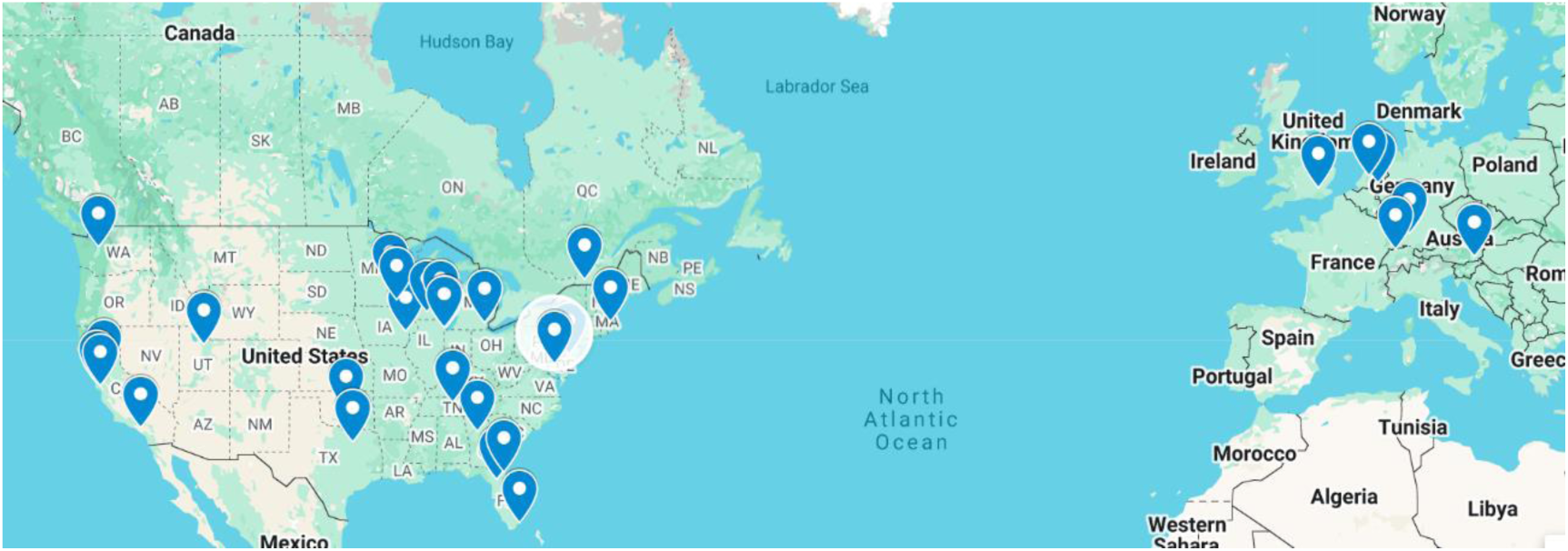
Locations of the NAC Attack clinical sites. The list of clinical sites is presented in Supplemental Material (S3)-List of NAC Attack clinical and AOSLO imaging sites.

Other trial entities include the study medication provider Zambon SpA, Italy providing both active and placebo; study drug secondary packaging, labeling and distribution center which is a GMP certified vendor specialized in global clinical supply chain services; and 2 accredited laboratory vendors, one to provide whole genome sequencing and retinal dystrophy genetic testing and the other one to provide plasma NAC level measurements for study participants.

#### Study Standing Committees

The Executive Committee is the study leadership includes Study Chair, Director of the Coordinating Center, and Program Officials of the NEI. The Steering and Quality Assurance Committee (SQAC) is composed by a subset of investigators, clinical coordinators, and representatives from other trial entities. The SQAC committee periodically reviews study progresses and serves as the primary source of advice to the study leadership regarding scientific, policy, administrative and logistical issues encountered during the conduct of the trial.

The Study Investigative Group consists of all trial personnel at all participating Clinical Sites,

Study Chairman’s office, Coordinating Center, resource centers, and other entities.

The Data and Safety Monitoring Committee (DSMC) has 7 members with a broad range of expertise including design and management of large clinical trials, biostatics, clinical aspects of RP and other inherited retinal degenerations, and bioethics. They are responsible for monitoring the ethical conduct of the NAC Attack Trial and for monitoring the accumulating trial data for evidence of adverse and beneficial effects of NAC, the study drug under evaluation. The DSMC is appointed by the NEI and provide recommendations to study leadership and the NEI.

### CHARACTERISTICS OF PARTICIPANTS

#### Inclusion and Exclusion Criteria

**General Inclusion Criteria**

- Ability and willingness to provide informed consent
- Age ≥ 18 and ≤65 years at time of signing Informed Consent Form
- Ability and willingness to comply with the study protocol and to participate in all study visits and assessments in the investigator’s judgement
- For candidates of childbearing potential: willingness to use a method of contraception
- Agreement not to take supplements other than vitamin A

**Ocular Inclusion Criteria**

- Both eyes must exhibit the RP phenotype with evidence of loss of night vision, gradual constriction of visual fields, and maintenance of visual acuity;
- Gradable EZ on a horizontal SD-OCT scan through the fovea center with width ≤ 8000 µm and ≥1500 µm and with well-defined truncation at both the nasal and temporal sides;
- BCVA ≥ ETDRS letter score of 61 (20/60, [6/18] Snellen equivalent);
- Sufficiently clear ocular media and adequate pupillary dilation to allow good quality images sufficient for analysis and grading by central reading center.

**General Exclusion Criteria**

- Active cancer within the past 12 months, except for appropriately treated carcinoma in situ of the cervix, non-melanoma skin carcinoma, or prostate cancer with Gleason score ≤ 6 and stable prostate specific antigen for > 12 months
- Renal failure requiring renal transplant, hemodialysis, peritoneal dialysis, or anticipated to require hemodialysis or peritoneal dialysis during the study
- History of thrombocytopenia not due to a reversible cause or other blood dyscrasia
- Uncontrolled blood pressure (defined as systolic > 180 and/or diastolic > 100 mmHg while at rest) at screening. If a patient’s initial measurement exceeds these values, a second reading may be taken 30 or more minutes later. If the patient’s blood pressure must be controlled by antihypertensive medication, the patient may become eligible if medication is taken continuously for at least 30 days.
- History of other disease, physical examination finding, or clinical laboratory finding giving reasonable suspicion that oral NAC may be contraindicated or that follow up may be jeopardized
- Cerebrovascular accident or myocardial infarction within 6 months of screening
- Patients taking slow-release formulations of nitrates
- Participation in an investigational study that involves treatment with any drug or device within 6 months of screening
- Three relatives already enrolled in study
- Pregnant or breast feeding females. Women of childbearing potential who have not had tubal ligation must have a urine pregnancy test at screening.
- Known history of allergy to NAC
- Having taken NAC in any form in the past 4 months
- Phenylketonuria
- Fructose intolerance
- Glucose-galactose malabsorption
- Sucrase-isomaltase insufficiency
- Any major abnormal findings on blood chemistry, hematology, and renal function lab tests that in the opinion of the Site Investigator and/or the Study Chair makes the candidate not suitable to participate in the trial
- HIV or hepatitis B infection
- Severe angina that requires frequent administration of nitrates

**Ocular Exclusion Criteria**

- Evidence of cone-rod dystrophy or pattern dystrophy including focal areas of atrophy or pigmentary changes in the central macula
- Cystoid spaces involving the fovea substantially reducing vision
- Glaucoma or other optic nerve disease causing visual field loss or reduced visual acuity
- Intra ocular pressure >27 mm Hg from two measurements. If a patient’s initial measurement exceeds 27 mm Hg, a second reading must be taken.
- Any retinal disease other than RP causing reduction in visual field or visual acuity
- Any prior macular laser photocoagulation
- Intraocular surgery within 3 months prior to screening
- High myopia with spherical equivalent refractive error > 8 diopters. If an eye has had cataract surgery or refractive surgery, a pre-operative refractive error spherical equivalent > 8 diopters is an exclusion
- Any concurrent ocular condition that might affect interpretation of results
- History of uveitis in either eye

### RECRUITMENT AND CONSENT

Potentially eligible patients are identified by chart review of RP patients previously seen at the study site, referred to the site by an outside physician for consideration for the trial, or by patients’ self-reaching out to the site. Patients deemed as potentially eligible are invited to an onsite screening visit. At this visit, after written informed consent, patients undergo study screening procedures. Screening data are recorded on study specified case reporting forms and entered into the study data management system based on the JHU REDCap (Research Electronic Data Capture) platform managed by the Coordinating Center. Images and test results are uploaded to portals of the Study Reading Center at Duke University and Coordinating Center at JHU.

Eligibility is determined by the Study Reading Center and the Study Chair’s Office with final confirmation by the Coordinating Center. If deemed as eligible, the patient candidate is scheduled for study Baseline Visit within 42 days of the screening date (**Figure 2**).

### RANDOMIZATION

The randomization schedule was generated using permuted block method by Coordinating Center and incorporated into the study REDCap database before study enrollment started. At Baseline visit, if eligibility is further confirmed by enrolling site investigator, patient candidate is randomized in a ratio of 2:1 to NAC 1800 mg bid or placebo. Site Coordinator uses the Randomization function in the study REDCap database to obtain a letter code encoding the treatment group assigned to the candidate. The cover of each study medication package also has a letter code. Site Coordinator will dispense medication packages that have the same letter code as the letter code to which the participant is assigned through REDCap. The treatment group letter encodes information about the content of the study medication (active drug or placebo) which is known only to selected personnel at the Coordinating Center.

### INTERVENTIONS

Participants are randomized 2:1 to active drug NAC 1800 mg bid or placebo, and instructed to take 2 doses during a day for 45 months. Each dose is taken by dissolving 3 effervescent tablets and drinking the solution.

Brand name of the active drug is Fluimucil 600 mg effervescent tablets. Placebo tablets have identical size, appearance, and ingredients except NAC. Both active and placebo are provided by Zambon SpA, Italy and manufactured in the plant at Cadempino, Switzerland. The study contracted Drug Labeling and Distribution Center (DLDC) provides secondary packaging to repackage 21 blister cards from Zambon into a tri-fold package which is the primary packaging and contains one week of supply. A multi-language booklet label following regulatory requirements from all involved regulatory agencies are affixed to each tri-fold package. The label cover page has a barcode unique to the tri-fold package and encodes the identifying number of this package. The label also is printed with a letter code that can identify content in the package. The linkage information between the letter code and study drug is securely kept and only accessible to DLDC and Coordinating Center unmasked personnel. Once randomized at Baseline and at M9. M18. M27 and M36 visits, participants are dispensed with packages for the next 9 months to use.

### OUTCOMES AND ESTIMANDS

**<u>Primary Outcome Measure and Estimand</u>**: the cumulative loss of ellipsoid zone (EZ) width between baseline and M45, calculated as the area above the curve (AAC) (***Figure 4***). EZ width is measured on the centermost horizontal SD-OCT scans through the fovea obtained at baseline, M9, M18, M27, M36, M40, and M45 and graded by the Study Reading Center.

**Figure 4.**
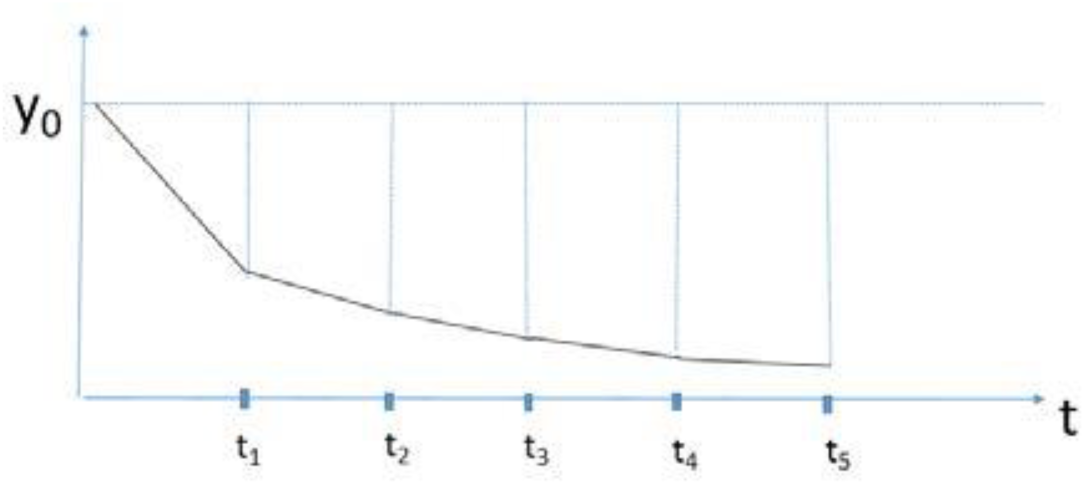
Estimating the Area Above Curve with outcome measurements observed at pre-specified follow-up visits. The AAC can be empirically and non-parametrically estimated as the sum of all the trapezoids determined by the measurements at all visits.

**<u>Secondary Outcome Measures and Estimands:</u>** 1) change between baseline and M45 in mean macular sensitivity measured by MAIA microperimetry, and 2) change between baseline and M45 in best corrected visual acuity (BCVA) measured by Early Treatment Diabetic Retinopathy Study (ETDRS) protocol.

**Exploratory Efficacy Objective**: to assess the relative efficacy in eyes of participants taking NAC 1800 mg bid compared with eyes of participants taking placebo on the basis of the following endpoints: 1) cumulative loss between baseline and M45 of EZ area assessed as the AAC, 2) change from baseline mean macular sensitivity measured by microperimetry at M4.5, M9, M18, M27, M36 and M40.5, 3) change from baseline BCVA at M4.5, M9, M18, M27, M36 and M40.5, 4) change from baseline in cone spacing, regularity, and reflectivity measured by adaptive optics-scanning laser ophthalmoscopy (AOSLO) at M9, M27, and M45 at selected sites, 5) proportion of eyes with ≥ 5 loci improved from baseline by ≥ 6 decibels (dB) on microperimetry testing at M4.5, M9, M18, M27, M36, M40.5 and M45, 6) proportion of eyes with ≥ 5 loci decreased from baseline by ≥ 6 dB measured by microperimetry at M4.5, M9, M18, M27, M36, M40.5 and M45, and 7) exploratory subgroup analysis to assess the treatment efficacy compared to placebo in subgroups defined by sex, race, inheritance mode, genotype, age of symptom onset, smoking status, and by history of oral supplement consumption, and 8) change from baseline to M27 and M45 in patient reported outcome assessed using NEI-VFQ 25.

**Safety Objective**: to evaluate the safety and tolerability of oral NAC 1800 mg bid in patients with RP based upon the Incidence and severity of ocular adverse events (AEs) and the incidence and severity of non-ocular AEs.

**<u>Pharmacokinetic objective:</u>** to measure plasma NAC levels at each visit in all participants to correlate plasma NAC levels with efficacy endpoints. The plasma NAC levels will also provide an assessment of compliance.

**<u>Genetic and Pharmacogenomic Objectives</u>**: 1) examine the relationship between disease causing mutations and rate of decrease in EZ width over time in placebo participants, and 2) examine the effect of disease variants and modifier variants on treatment effect. Participants will be divided into high-responders and low-responders and their exomes will be compared (a) for patterns in disease genes and (b) in an unbiased fashion regardless of their disease gene to determine if certain disease genes or unbiased-selected genes have higher variant burden in high responders versus low-responders.

### SAMPLE SIZE DETERMINATION

#### Original Sample Size Calculation

Sample size is determined for the treatment effect as measured by the primary outcome measure of EZ-width on the OCT fovea scan. The variable for assessing the treatment effect is the AAC during the M45 follow-up. The summary statistic for the treatment effect is the difference in the AAC between the intervention arm and the control arm. The AAC variable calculated is essentially a linear combination of the repeated measurements for an eye. Thus, for each study eye, the repeated measurements are reduced to a scalar statistic-the AAC. The framework of generalized estimating equation (GEE) with linear model can be used to estimate the difference between the intervention and control arms while accounting for the between-eye correlation on the AAC variable. PASS 2019 the GEE Test for Two Groups (Continuous Outcome) was used to do the sample size calculations.

##### Sample Size Re-estimation and Final Sample Size

Sample size calculations are always based on assumptions of relevant parameters. How well these assumptions apply to the NAC Attack study populations is unknown as a priori. To ensure enough study power regarding the primary outcome measure, during enrollment period, the study sample size would be re-estimated based on refined parameters estimated using baseline data of available NAC Attack study participants and longitudinal data from other studies of RP ongoing at the Wilmer Eye Institute.

The sample size re-estimation would be based on masked data only. This ensures the study type-I error is not inflated. The re-estimation also needs to be conducted relatively early during the enrollment period, allowing time to arrange necessary logistics before closure of the originally planned enrollment period, should study sample size be increased.

After about 100 trial participants were enrolled and randomized by the end of 2024, their data were used to refine the parameters needed in sample size calculation. Assumptions in the original sample size calculation on the parameters such as the proportion of participants with one eye enrolled and the between-eye correlation of the AAC of EZ-width were deemed as reasonable, but the observed study withdrawal rate was about two times higher than the originally assumed rate. Therefore, the Data Safety Monitoring Committee (DSMC) deemed it sensible to refine the assumption of this parameter in sample size determination.

Further considering enrollment feasibility and logistics capacity, the final study sample size was decided to be 483. Assuming the between-eye correlation of AAC is 0.7, alpha=0.049, 86% of participants have both eyes enrolled, this sample size can yield 85% and 90% power if the effect size comparing the active and placebo arms is Cohen’s D of 31% and 34%, respectively.

### TRIAL CLINICAL PROCEDURES

In-clinic study visits at the Screening, Baseline and follow-up visits may include the following procedures with patients per the schedule of procedures in **Figure 1**: measuring vital signs, intraocular pressure, BCVA testing using the ETDRS protocol, MAIA microperimetry testing, slit lamp and funduscopic examinations, SD-OCT imaging, ultra-wide field fundus photography, ultra-wide field fundus autofluorescence (FAF), blood draws for chemistry, hematology and liver function tests at local labs, blood draw for whole genome sequencing by study central lab, blood draws for plasma NAC measurements by study central lab urine collection for pregnancy testing, and study medication reconciliation and dispensing. The last procedure at the Baseline Visit after randomization is training participants about taking, recording dosing date and time, and caring of their study medication. Participants also take their first dose under supervision by site study Coordinator. At selected sites, patients also undergo full field stimulus testing at the Baseline, M27 and M45 visits.

Six study sites participate in the AOSLO sub-study. During the informed consent process at these sites, patient candidates are given the option to participate or not in the AOSLO sub-study. Participants who consent to participate will be assessed for AOSLO sub-study eligibility by their enrolling site investigator. The assessment data are provided to the AOSLO Reading Center to determine AOSLO sub-study eligibility: if eligible, AOSLO baseline visit should be scheduled before the NAC Attack baseline visit so that AOSLO baseline imaging is acquired before study medication initiation. AOSLO Reading Center determines the final eligibility for AOSLO imaging follow-up based on quality of the imaging acquired at the AOSLO baseline visit.

Follow-up visits include tele-visits at M13.5, M22.5, and M31.5 with site Investigator and telephone calls with site Coordinator in-between in-clinic visits and/or tele-visits (**Figure 2**). The purpose of a tele-visit with Investigator or phone call with Coordinator is to maintain contact with participants and keep them engaged in the study. A brief case reporting form is completed to document the tele-visit or phone call, including medication compliance assessment and any adverse events (AEs) reported.

In addition, a short message service (SMS) text-based system set by the Coordinating Center is used to provide a SMS reminder to take medication on a daily basis. Through the SMS, once a week, participants are also asked to report how many doses were missed over the past week. Participants can opt in or out to receive the daily text reminder or weekly text survey at any time during their study participation.

### SAFETY CONSIDERATIONS

Participants have a medical history and specific questioning to elicit any AEs at each study visit after baseline. Ocular safety is assessed by measurement of BCVA, IOP measurement, slit lamp examination and indirect ophthalmoscopy at in-clinic visits as specified **Figure 1**. Systemic safety is assessed by medical history, vital signs, and blood chemistry, hematology, and liver function tests. All participants are instructed to contact their Clinical Site at any time if they have any health-related concerns. If warranted, participants will be asked to return to their Clinical Site as soon as possible for an unscheduled safety assessment visit. In addition, tele-visits and study coordinator phone calls are conducted in between in-clinic visits to inquire about AEs and compliance.

Site Investigator and Coordinator documents the medical name of an AE on a study CRF and enter the AE description into study database. Site Investigator also grades severity of an event and attributes causality to study drug of the event. The Study Chair’s Office review all AEs, including grading AE severity and determining causality to the study. Moreover, a Medical Monitor and the DSMC monitor safety throughout the trial.

Serious adverse events (SAEs) and AEs of Special Interest as defined in study Protocol and pregnancies of female participants during their study participation require site Investigator’s immediate reporting to the Coordinating Center to allow Coordinating Center to take appropriate measures to address potential new risks and if justified to report to IRB or ethics committees and regulatory agencies with their required timelines.

### DATA MANAGEMENT PLAN

Data collected about trial participants include data collected on CRFs and electronic files from imaging or tests. Standardized CRFs are developed by study Coordinating Center and Chair’s Office and used by all sites. During a patient visit, data are first recorded on paper CRFs. Site staff then enter CRF data on the corresponding study database in REDCap. A scanned copy of CRFs is uploaded to JHOneDrive based storage location. Coordinating Center routinely checks REDCap data entry accuracy by comparing to the scanned source CRFs. Electronic data files are uploaded to Coordinating Center OneDrive. The REDCap platform and JHOneDrive are administered by the Johns Hopkins University Information Technology department and are compliant to HIPAA and relevant international standards on data security. Data quality assurance procedures are described in study Protocol and Manual of Procedures. OCT and AOSLO imaging files are submitted by site staff to the Study OCT Reading Center and AOSLO Reading Center, respectively, through each Reading Center’s FDA CFR Part 11 compliant data transmission portal. The Reading Centers routinely submit completed image grading data to the Coordinating Center’s JHOneDrive.

### DESIGN AND STATISTICAL ANALYSES PLAN

The NAC Attack study was designed following FDA’s Guidance regarding ICH E9 (R1) Statistical Principles for Clinical Trials: Addendum: Estimands and Sensitivity Analysis in Clinical Trials. The ICH E9(R1) addendum aims “to improve the planning, design, analysis and interpretation of clinical trials” by presenting a structured framework, the estimand framework, to link trial objectives with trial design and tools for estimation and hypothesis testing. ^[24]^ At time of developing of the NAC Attack study design, the estimand framework required clear delineations on 4 attributes of the trial: A: Population; B: Variable; C: Intercurrent event; and D: Population-level summary for the variable.

#### Estimand-framework Attribute A: Study Population

The NAC Attack study population is patients with RP and enrolled in NAC Attack. The study inclusion and exclusion criteria reflect characteristics of RP patients represented in NAC Attack.

#### Estimand-framework Attribute B: Primary Endpoint

##### EZ-width as the primary outcome measure

The primary outcome measure of NAC Attack is the width of the EZ on a horizontal OCT scan through the foveal center. Regulatory agencies prefer functional over anatomic or structural endpoints. [25] However, RP is a slow progressing disease and using functional outcomes such as visual field loss in RP treatment trials may be impractical and expensive as many years would be needed to show efficacy. The BCVA has been the gold standard endpoint in ophthalmic trials because of its strong correlation with ability to function in activities of daily life, [26, 27] but in RP, BCVA is excellent until late in the disease and is expected to show no substantial decline for decades in the RP patient population that we are interested in studying. [25] Macular sensitivity measured by microperimetry may provide a useful functional outcome in inherited retinal diseases.[28] However, in RP, it is noted that some patients may not be able to perform microperimetry test reliably as identified by a consistently high false-positive response rate.[29, 30] Therefore, using a microperimetry based parameter as a primary outcome measure would restrict patient enrollment criteria.

Additionally, visual function measurements are subject to influences of confounding pathologies in addition to cone function and survival. Patients with RP often develop posterior subcapsular cataract and cystoid macula edema (CME) which have a negative impact on visual function. Different clinical sites may have different practices in managing cataract and CME. Thus, factors such as progression of lens opacity, cataract surgery, and fluctuations in CME may confound the estimation of the effect of the intervention on visual function measurements of BCVA or macular sensitivity outcomes.

The EZ represents the ellipsoid region of the inner segments of photoreceptors and is recognized as a well-defined bright line on a SD-OCT scan. The EZ width is measured as the horizontal extent of the EZ on a SD-OCT scan through the foveal center and provides a measurement of remaining photoreceptors with intact inner and outer segments in the horizontal meridian through the fovea. Thus, the EZ-width is an anatomic measure that reflects the integrity of photoreceptors. It is selected as the primary outcome measure for NAC Attack with the following considerations:

- EZ width correlates with visual function outcomes and has been accepted by the FDA as a surrogate endpoint in the context of geographic atrophy due to AMD.[28]
- EZ width can be reliably measured in patients with RP.[31] In the FIGHT-RP study, the intra class correlation coefficient was 0.98 baseline EZ width measurements graded for two baseline images by different graders.
- Natural history studies in patients with RP have shown that loss of EZ width is detectable during one to a few years of follow-up.[32–34]
- The FDA has agreed that assessment of loss of EZ is an appropriate outcome measure for evaluating treatment effect in RP.

The area of EZ can be measured on enface SD-OCT images, and reflects the preservation of the EZ of the entire macula; whereas foveal-centered EZ-width is a measurement of the EZ on the horizontal meridian through the foveal center. However, numerically, published studies suggest that these two variables are highly correlated and that EZ-width measurement has stronger structure-function correlations than EZ-area, and using EZ-width is statistically more efficient and incurs less stringent enrollment criteria. EZ-width on the horizontal fovea scan therefore is determined to be the primary outcome measure in NAC Attack.

##### Primary Endpoint Derived from the Primary Outcome of EZ-width

The primary outcome of NAC Attack is the EZ-width on the fovea horizontal OCT scan. Because it is unknown whether the loss of EZ-width over 45 months would follow a linear trajectory, the rate of EZ width loss per year would not be an appropriate parameter as “rate” implies linear change over time. An alternative variable is the change of EZ-width from the baseline to the M45 follow-up. However, the FDA recommended that the primary outcome measure should be assessed at a minimum of 5 follow-up time points with 2 adjacent time points at least 9 months apart and that the efficacy should reflect separation of EZ loss curves for the intervention versus the placebo groups. Therefore, the derived variable of Area Above the Curve (AAC) which can be interpreted as the cumulative loss of EZ over the M45 of follow-up was selected as the primary endpoint. **Figure 4** illustrates the estimation of the AAC. Operationally, to estimate the AAC, the area is non-parametrically estimated as the sum of trapezoids in **Figure 4**. Although EZ loss is expected to be a degenerative process, it is unknown whether EZ width at a follow-up visit may be greater than that at baseline due to measurement error or due to intervention effect. A general formula without assuming how EZ width changes over time is derived and presented below **(Figure 5**, Protocol Addendum 2024-10-01).

**Figure 5.**
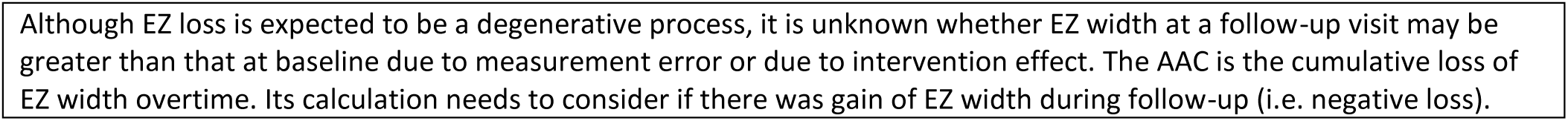
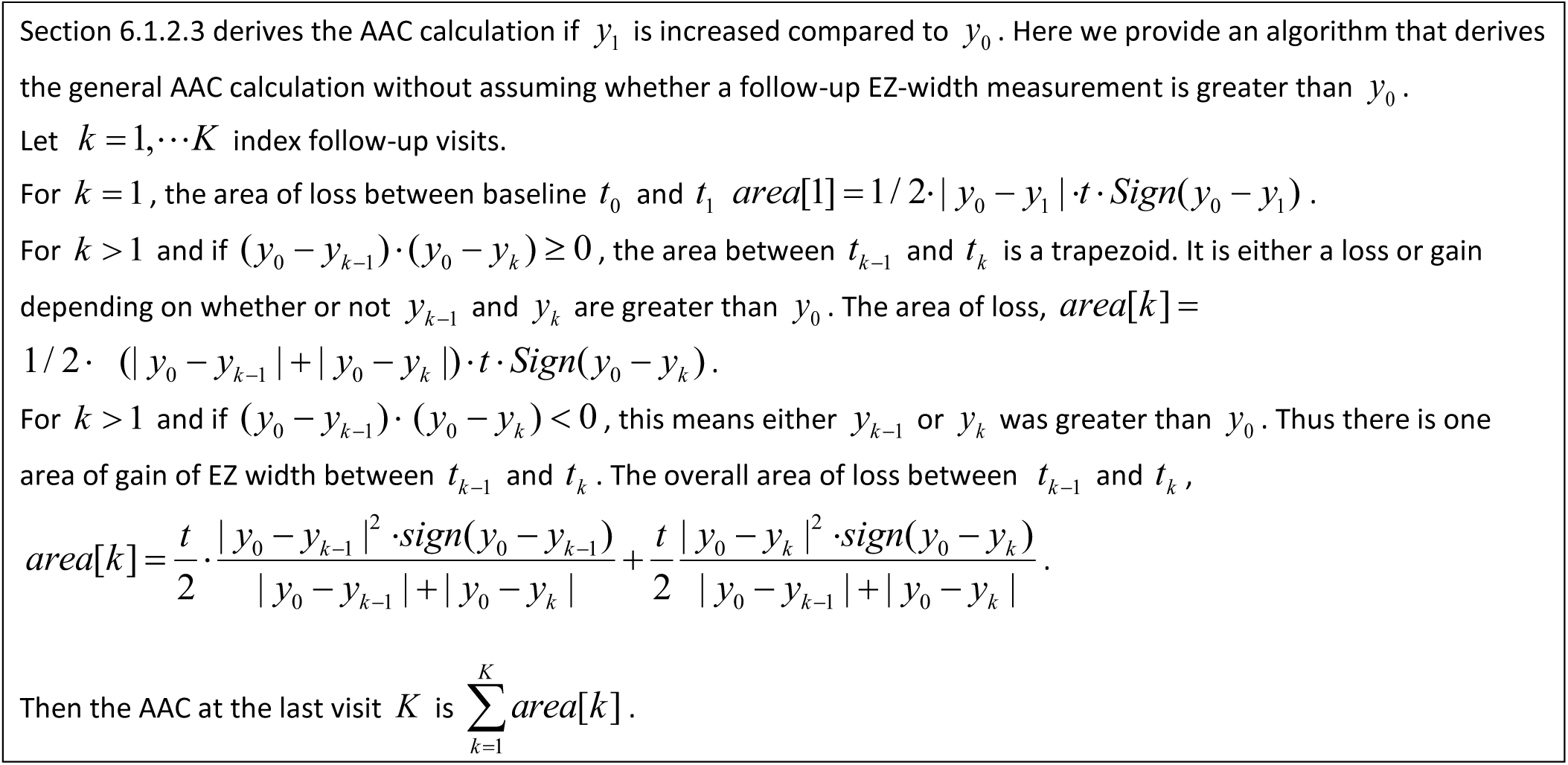
Algorithm to calculate the primary estimand of Area Above the Curve, i.e. cumulative loss of EZ-width.

#### Estimand-framework Attribute C: Intercurrent Events

“Intercurrent events” refers to “Events occurring after treatment initiation that affect either the interpretation or the existence of the measurements associated with the clinical question of interest.” The ICH E9(R1) recommends to clearly describe the envisioned intercurrent events and the planned strategies to account for the intercurrent events at the design stage.

Possible intercurrent events in NAC Attack:

- Participant taking his/her assigned medication but not at the prescribed level or frequency, i.e. sub-optimal medication adherence. “Treatment policy strategy” where the observed values for the primary and secondary outcome measures will be used irrespective of the medication adherence measures. The “Treatment policy strategy” reflects the well-known ICH E9 Intention-To-Treat (ITT) principle. [24] Additionally, the treatment effect under the “Hypothetical strategy” is also of interest. Supplementary analysis will be conducted to estimate the treatment effect under hypothetical strategy, i.e. using the Per-Protocol Set. [35]
- Participants randomized to the control arm self-taking NAC supplement. The measurements of plasma NAC using samples obtained during in-clinic visits can help identify incompliance of control arm participants. “Hypothetical strategy” will be used to account for intercurrent event of this kind of incompliance in the control group.
- Intercurrent event due to pregnancy. Primary analysis accounting for this type of intercurrent event will use the “Hypothetical strategy” which is the scenario that pregnancy had not occurred and the participant had stayed on study drug. Supplementary analysis will also be conducted to estimate the treatment effect under “Treatment policy strategy”. Additionally, sensitivity analysis will be conducted by excluding the participants who experience pregnancy during the study follow-up.
- For intercurrent event due to study treatment discontinuation for managing AEs, the “Treatment policy strategy” i.e. ICH E9 Intention-To-Treat (ITT) principle will be used for data analysis.

#### Estimand-framework Attribute D: Population-Level Summary for the Primary Endpoint of Area Under the Curve

The difference between the AAC in the intervention group and the AAC in the control group over the 45 months follow-up period is the population-level summary for the variable for the primary outcome in NAC Attack.

#### Secondary Endpoints

The ICH E9 (R1) specifies that estimands for secondary trial objectives (i.e. related to secondary variables) that may support regulatory decisions should also be defined and specified explicitly. NAC Attack secondary outcome measures include macular mean sensitivity (MMS) obtained from the MAIA microperimetry test and BCVA. The corresponding estimands are the change between baseline and M45 of BCVA and MMS, respectively. The study population, intercurrent events, and population-level summary are similar to those for the primary endpoint. Analysis for the secondary endpoints will follow the same principles for the primary outcome measure.

#### Principles for Primary Analysis

The primary analysis will follow the Intent-to-treat principle and use the Full-Analysis-Set including all randomized participants. Construction of the Full-Analysis-Set handles missing data (if the EZ-width measurement for the last study visit at M45 is unavailable) and intercurrent events. AAC is essentially an eye-level summary measure of the repeated measurements over time. Most participants will have both eyes enrolled as study eyes, and thus analysis needs to account for the clustered data nature. The statistical modeling framework will be linear regression with GEE to account for the between-eye correlation. SAS PROC GENMOD will be used for the analysis. The “Compound Symmetry” option will be used for modeling the working correlation matrix (dimension is 2x2) and the empirical standard error estimates will be used. Wald-test will be used for hypothesis testing.

To improve precision and efficiency and following the FDA’s “Adjusting for Covariates in Randomized Clinical Trials for Drugs and Biologics with Continuous Outcomes Guidance for Industry” (April 2019), the primary analysis model will adjust for baseline covariates that introduce imbalance by chance between the groups as well as prognostic factors known to be strongly associated with disease progression. Because the study does not use an equal allocation, the baseline covariates that have chance imbalance must be determined after study unmasking. Any baseline covariate having a p-value ^≤ 0.1^ for comparing the intervention and control groups will be included in the primary analysis model.

Sensitivity analysis will also be conducted to examine the robustness of the primary analysis results. Sensitivity analysis will be conducted using the tipping point analysis based on multiple imputations.[36, 37]

#### Supplementary Analysis

Supplementary analysis will be conducted by taking into account of medication adherence and placebo control group participants incompliance. The Per-Protocol Set [35] (PPS) will be used in such analysis. As noted in the ICH E9 R(1) Addendum, the PPS based results may be subject to bias, for reasons such as that medication adherence pattern is different between the treatment arms or that control group participants of certain characteristics that are related to disease progression may be more likely to self-taking a supplement formulation of NAC. If the results from the primary and per-protocol analyses differ substantially, further exploratory analyses will be performed to evaluate the factors that have contributed to the differences.

To estimate the “hypothetical strategy” estimand which provides an estimate of the biological effect of the treatment and account for the intercurrent event of suboptimal adherence to study medication dosing or frequency (i.e. an intercurrent event), additional analysis with the PPS dataset will apply causal inference methods such as propensity score based matching or inverse probability weighting methods. [38]

#### Hypothesis Testing with the Primary and Secondary Outcomes, Multiplicity and Disjunctive Power

The primary outcome measure is EZ-width which is an anatomic parameter reflecting the integrity of photoreceptors. It is selected as the primary outcome measure for sample size considerations and because of the limitations of visual function outcomes in RP. The secondary outcomes of BCVA and MMS are both visual function outcomes and are clinically important. EZ-width has been shown to be correlated with visual functions in RP.[33, 34, 39, 40] It has also been shown that there can be sensitivity response outside of the reflective band of the EZ.[41, 42] Therefore, the 3 endpoints are correlated but provide assessments of different aspect of the disease process. Thus, the hypotheses testing of these 3 endpoints are deemed of equal scientific importance and will not be tested sequentially following a pre-specified order (normally determined by their levels of importance).

Following the FDA “Multiple Endpoints in Clinical Trials Clinical Trials Guidance for Industry” (Draft Guidance, January 2017), multiplicity needs to be accounted for to allow findings of significant treatment effects at the individual endpoint level while controlling the Type I error rate (*α*) of the trial.Assumptions are considered as met for using stepwise semiparametric procedures with a data-driven hypothesis ordering. Specifically, the step-up Hochberg method will be used. The procedure is presented in **Figure 6** detail in Protocol 6.5.3.1.

**Figure 6.**
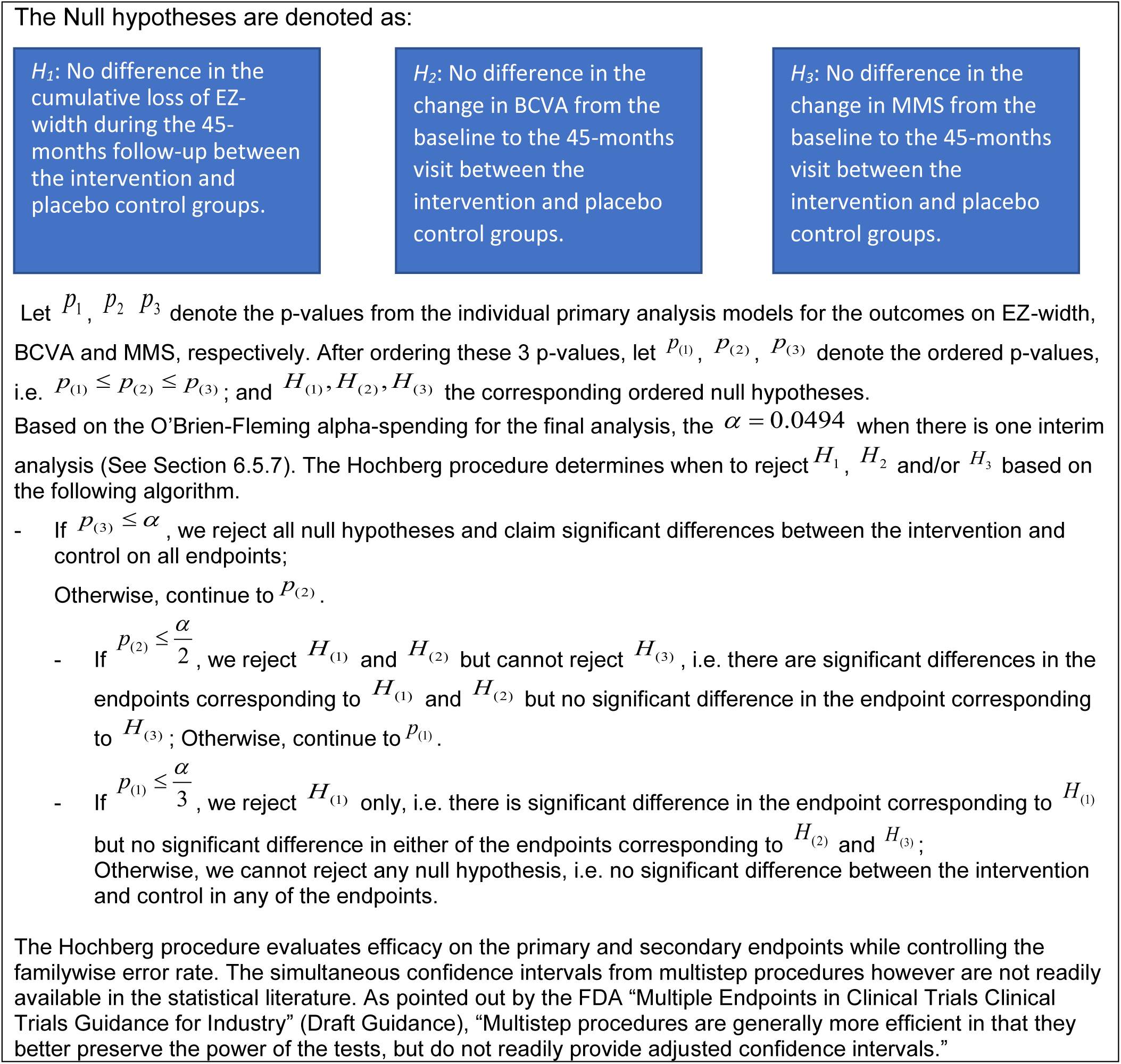
Hypothesis testing of the primary and secondary outcomes using Hochberg method.

Disjunctive power is the power to detect at least one effect from all the outcome measures in the hypothesis testing.[43] The study sample size was determined based on power and sample size calculations of the primary outcome measure and considering resources and feasibility. Simulations were conducted to assess the disjunctive power of the study hypothesis testing plan given the effective study sample size (Protocol Addendum 2025-02-20).

#### Interim Analysis

NAC Attack will have one interim analysis and the final analysis. The interim analysis will be conducted after all participants complete their 27-months follow-up and will use all data up to the 27-months visit. The decision of an interim analysis at 27-months is based on 2 considerations: first, RP progression is slow in the disease natural history (patients lose vision over decades). Thus it is thought that at least 2 years would be needed to have a likelihood to observe some treatment effect that can suggest long-term efficacy of NAC on delaying disease progression. Second, the study treatment NAC has known a safety profile for its FDA approved usages (such as treating acetaminophen overdose) which is short-term use. There are no data on the long-term safety of NAC. Therefore, it is important to have long-term use safety data.

The focus of the interim analysis is to evaluate safety and efficacy. The interim analysis will not seek to re-estimate sample size or change study population because of the logistical complexity in an international multicenter trial. The interim analysis will also not test for futility because given the great unmet need in RP management and the slow progressing nature of the disease, any type II error will be consequential as it will be undesirable to discard a promising treatment with good safety profile by erroneously stopping the trial early. [44]

The efficacy evaluation at the interim analysis will use the O’Brien-Fleming alpha spending rule[44] and the decision rule will be based only on the efficacy of the primary objective of significant reduction in the cumulative loss of the EZ-width (i.e. AAC over the 27-months) in the intervention group compared to the placebo group. The significance level uses O’Brien-Fleming boundary of 0.0054. [44]

If efficacy is determined, the randomized trial will stop but NAC Attack will continue to follow all participants until all participants complete their M45 visits. Each control group participant will be provided with NAC at no cost for another 12 months. As soon as interim efficacy is determined, the study Drug Manufacturer and Drug Labeling and Distribution Center will be notified to prepare for the batch allowing for the control group participants to receive NAC for 12 months.

### PROCEDURES REGARDING BIOLOGICAL SAMPLES

NAC Attack requires blood sample collection at most in-clinic visits. The schedule of blood draw can be found in **Figure 1**. Blood samples will be used for 3 purposes listed below. All blood samples collected for NAC Attack study purpose will be destroyed by the end of the study.

- Genetic testing to identify pathogenic variants for RP and whole genome (exome) sequencing.

- Test Frequency: only once using the blood sample collected at one study visit.
- Specimen type: whole blood.
- Lab: Study central laboratory contracted by the Coordinating Center.
- Testing for routine blood chemistry, hematology, and liver function to monitor safety.

- Test Frequency: at most in-clinic visits per **Figure 1**.
- Specimen type: whole blood.
- Lab: Clinical Site’s local diagnostic lab.
- Testing for plasma level of NAC.

- Test Frequency: at most in-clinic visits per **Figure 1**.
- Specimen type: plasma.
- Lab: Study central laboratory contracted by the Coordinating Center.

### ASSESSMENT OF MEDICATION ADHERENCE

Compliance with taking of study medication will be assessed in several ways: 1) reconciling of study drug remaining in returned drug packets at in-clinic visits, 2) participant self-report of medication adherence in the past week at each study visit including virtual visits and coordinator phone calls. 3) weekly SMS texting surveys, and 4) plasma NAC levels measured using blood drawn at select in-clinic visits. In addition, participants are required to keep drug diary about each dose taken.

### EMERGENCY UNMASKING

Site investigator is responsible for the medical care of trial participants. Under emergency situations when site investigator feels a participant has an SAE that necessities immediate unmasking of the participant’s treatment group to manage the event, site investigator may contact the Coordinating

Center to receive the unmasked treatment group information. Coordinating Center unmasked staff will provide the information to site investigator in a timely manner. This process should be recorded in the Unmasking Form. Communication during this process between the Coordinating Center and the clinical site will ensure masking remain in place for all other participants.

### SITE DOCUMENTATION AND MONITORING

#### Study Documentation

The investigator must maintain adequate and accurate records to enable the conduct of the study to be fully documented, including, but not limited to, the protocol, protocol amendments, Informed Consent Forms, and documentation of IRB and regulatory approval. In addition, at the end of the study, the investigator will receive patient study data, including an audit trail containing a complete record of all changes to data.

#### Protocol Deviations

The Investigator should document and explain any protocol deviations. The Investigator should promptly report any deviations that might have an impact on patient safety and data integrity to the Coordinating Center and to the IRB in accordance with established IRB policies and procedures.

Protocol deviations may also be identified during Coordinating Center’s site monitoring visits or quarterly conference call with clinical sites.

The Coordinating Center will review all protocol deviations and assess whether any represent a serious breach of Good Clinical Practice guidelines and require reporting to health authorities.

#### Management of Study Quality Through Training and Certification

The Coordinating Center implements a NAC Attack Training and Certification Program to ensure standardized training on all protocols and data collection procedures, along with certification of NAC Attack personnel. The NAC Attack Steering and Quality Assurance Committee oversees the implementation of the training program. The Coordinating Center is responsible for finalizing certification requirements, developing the on-line system for completing knowledge assessments, providing customized feed-back, issuing certification certificates, and maintaining the electronic and paper roster of all certified study personnel.

In brief, all members of the Investigative Group must complete a General Knowledge Assessment about NAC Attack that requires knowledge of basic design facts (e.g. duration and frequency of participant follow-up, the primary and secondary outcome measures), AE and IRB reporting guidelines and timelines. In addition, individuals are required to complete role-specific assessments.

#### Monitoring and Quality Assurance

The Coordinating Center has primary responsibility for assuring the data collected and reported in the study are consistently of high quality. Some major quality assurance features of the study include:

- Standard data collection forms and procedures;
- MASKED outcome assessments at the Clinical Sites;
- Central masked grading of OCTs;
- Checking data entered into the study database by the Coordinating Center by comparing to the scanned copy of source forms submitted by Clinical Sites.
- Central, computer driven data editing for missing, invalid, and suspect responses;
- On-Site and Virtual monitoring visits to all centers;
- Specific data analyses to identify incorrect or fraudulent data collection processes;
- Certification of clinic staff and of imaging equipment;
- Quarterly conference calls with each clinical site;
- Regular meetings of the SQAC to review methods and discuss problems.

### PUBLIC AND PATIENT INVOLVEMENT

Several patient groups were involved in the trial planning in various ways. The USHER 2020 Foundation assisted in the design of study medication secondary packaging and study patient facing materials, including information about the study and study medication which are given to study participants after they are randomized; public study website, and education videos about study participation. They also provided study participants with roller duffle bags to facilitate participants transport of study medication from clinic to home. All documents and materials given to participants were approved by IRB or ethics committees. Retina UK facilitated the study in obtaining approval from UK’s Medicines and Healthcare products Regulatory Agency.

To keep study participants engaged throughout the trial, annually the study Investigative Group Meeting will include a Zoom session where study patients are invited to join.

## DISCUSSION

### DESIGN CONSIDERATIONS

The NAC Attack Trial aims to determine if oral NAC 1800 mg bid can slow progression of retinal degeneration in patients with RP. The rate of EZ loss in RP is slow and therefore a long follow-up period is needed to observe a statistically significant and clinically meaningful NAC treatment effect.

The FDA recommended measurement of cumulative change in 5 EZ measurements with minimum time between measurements of 9 months. This requires a total follow-up period of 45 months which determined the study duration. To reduce study cost and burden on participants and clinical sites, data collection focus on testing essential for assessment of primary and secondary outcomes and patient safety, and in-clinic study visits are done infrequently at every 9 months, plus a visit at M4.5 for monitoring safety and study drug adherence soon after study drug initiation and a visit at M40.5 to allow more chance of collecting data on outcome measures. Despite such infrequent visits, patient oversight and engagement are maintained with frequent protocol specified Coordinator phone calls, tele-visits with investigators, and SMS text reminders and surveys. Patient attitudes and acceptability of these design features were considered during protocol development.

Dose selection of NAC Attack was informed by data from the FIGHT RP study.[29] In the FIGHT RP study, the maximum tolerated dose of NAC was 1800 mg bid and the third cohort showed significant improvement in mean macular sensitivity during treatment with 1800 mg bid NAC for 3 months followed by 1800 mg tid for 3 months and a significant effect was not seen with lower doses.[45] Selection of a dose of 1800 mg bid in the phase-3 trial was also supported by locus-level analysis of macular sensitivity changes which demonstrated that the percentage of loci that decreased ≥ 6 dB was significantly less in the third cohort.[46]

NAC does not target any specific disease-causing genetic variant, but is meant to reduce oxidative damage and promote cone survival. This is in contrast to interventions aimed at correcting or mitigating the effects of the pathogenic genetic variant in patients with RP which would only apply to a small subgroup of RP patients who have the targeted pathogenic genetic variant. These intervention approaches are meant to promote rod photoreceptor survival and function, and thus should be instituted early in the disease before there is substantial rod photoreceptor cell death. Treatment with NAC potentially applies to all patients with RP, and knowing the pathologic genetic variant is not a requirement for participation in NAC Attack.

However, the study population is restricted to the stage of RP in which photoreceptor degeneration is measurable by SD-OCT. To minimize the restriction of SD-OCT imaging window, the scan setting of 30°X 25° is used rather than the common 20°x20° setting.[47] Thus study eye eligibility includes having an EZ on the horizontal foveal OCT scan that has well-defined truncation at both the nasal and temporal sides as visualized by SD-OCT using a 25°X 30° window, which has an upper limit of about 8000 µm. The study population also requires exclusion of late-stage patients who have limited preserved EZ because more centrally constricted EZ is known to be associated with slower EZ loss and the FIGHT RP study observed that those late stage patients who had a limited measurable EZ-width continued to have functional decline during the NAC treatment period. Moreover, a mathematical modeling study computationally predicted that under mutation-induced rod degeneration, treatments with antioxidants or trophic factors can significantly delay degeneration of photoreceptors and the earlier the treatment, the greater delay in the onset of blindness in patients with RP. [48] This suggests that there may be a larger NAC treatment effect for early stage versus late-stage RP patients. Based upon these considerations, it has been decided that only RP patients with EZ width ≥1500 µm will be eligible for the study.

### STUDY PLANNING PHASE AND SITE SELECTION

Evidence of availability of patient population and qualifications of clinical centers was necessary in supporting the grant applications to NEI. During the trial planning phase (2019-2021), the Coordinating Center designed and organized a multicenter retrospective chart review study “NAC Attack Zero”. Following the data collection protocol provided by the Coordinating Center, potential clinical sites voluntarily identified at least 20 potentially eligible patients from medical charts or their database of RP patients and entered de-identified data regarding demographics, genetic testing information, latest known visual acuity and EZ widths into the NAC Attack Zero project database in REDCap. Sites also provided information on their facilities, equipment, and personnel resources

The data were summarized and together with information on clinical sites facility, equipment, and relevant personnel resources were presented in the grant applications, demonstrating feasibility of patient enrollment, capability of and commitment from these clinical sites. The list of 20 identified patients later provides a starting point for inviting patients for screening, allowing rapid study recruitment.

### ELIGIBILITY DETERMINATION AND RECRUITMENT STRATEGY

Pre-screening of RP patients is encouraged with assistance provided by the Chair’s Office, Coordinating Center, and OCT Reading Center (OCTRC) to reduce the likelihood of screen failures. After a screening visit, the site enrolling Investigator preliminarily determine patient eligibility. Data collected from the screening visit procedures are further reviewed by the Study Chair’s Office to determine RP (or not) diagnosis, and OCT images are reviewed and graded by the OCTRC to determine EZ related eligibility criteria. The Coordinating Center subsequently synthesize eligibility determination results from the Chair’s Office and OCTRC and notify the enrolling site about the final eligibility of the patient.

## STATUS AND TIMELINE OF THE STUDY

The study received funding from the NEI in March 2022. Patient screening started in October 2023 using approved study Protocol Version 2.0 Version Date 2023-07-13, and as of June 2025 the study is approximately 90% enrolled. It is projected that enrollment (randomization) may be completed by August of 2025. Participant follow-up will be completed in 45 months after the last patient is randomized.

The end of study is defined as one year after the date when the last study participant completes the final study visit. This end of study is defined to allow completion of scheduled data analyses, scientific publication of the main study findings and appropriate archiving of study materials.

### Supplemental Materials

*Supplemental Material 1: Spirit Checklist*

*Supplemental Material 2: Full Study Protocol*

*Supplemental Material 3: List of active NAC Attack clinical and AOSLO imaging sites.*

*Supplemental Material 4: The NAC Attack Study Investigative Group*

## DECLARATIONS

### ETHICS APPROVAL AND CONSENT TO PARTICIPATE

This study is conducted in full conformance with the ICH E6 guideline for Good Clinical Practice and the principles of the Declaration of Helsinki, as well as the laws and regulations of the country in which the research is conducted, whichever affords the greater protection to the individual. The study complies with the requirements of the ICH E2A guideline (Clinical Safety Data Management: Definitions and Standards for Expedited Reporting).

The study is approved by the U.S. Food and Drug Administrating (FDA), the European Medicines Agency (EMA) under E.U. Clinical Trial Regulation (No 536/2014), UK Health Research Authority (HRA) and Medicines and Healthcare products Regulatory Agency (MHRA), Swissmedic, and by Health Canada under Natural Health Products Regulations.

The sIRB (Single Institutional Review Board) of the Johns Hopkins University (JHU) School of Medicine with reliance from local IRBs of participating clinical sites reviews and monitors the study conduct in the US sites. For each non-US study site, the study is approved and overseen by the local institutional or national Research Ethics Committee or Research Ethics Board.

Informed Consent Form (ICF) was developed by the study leadership following JHU sIRB requirements and applies to sites in the US. Each non-US site working with the Coordinating Center adapted the JHU ICF into local language and complying to local format and ethics requirements. The ICF must be signed and dated by the patient before his or her participation in the study. The case history or clinical records for each patient shall document the informed consent process and that written informed consent was obtained prior to participation in the study.

The Study maintains confidentiality standards by coding each patient enrolled in the study through assignment of a unique study identification number. This means that patient names are only known to the study team members of the clinical site where the patient is enrolled. Patient names or medical record numbers will not be included in data sets, images, test results or specimens transmitted to the Coordinating Center, the Reading Centers, or study laboratories. Patient medical information obtained by this study is confidential and may be disclosed to third parties only as permitted by the ICF (or separate authorization for use and disclosure of personal health information) signed by the patient, unless permitted or required by law.

### CONSENT FOR PUBLICAITON

Regardless of the outcome of the trial, the study leadership are dedicated to openly providing information on the trial to healthcare professionals and to the public, at scientific congresses, in clinical trial registries of the U.S. National Institutes of Health and the European Medicines Agency, and in peer-reviewed journals. The Study will comply with all require-ments for publication of study results. Study data may be shared with others who are not participating in this study, and redacted clinical study reports and other summary reports will be provided upon request.

Study publications are defined as those that use data, documents, or other information collected during the course of the Study. Publication of study results will be governed by the policies and procedures in the study protocol (Protocol Section 11.2).

### AVAILABILITY OF DATA AND MATERIALS

Study data may be shared with researchers, government agencies, companies, or other groups that are not participating in this study to advance science and public health, or for analysis, development, and commercialization of products to treat and diagnose disease. The study data sharing will follow the NIH’s Data Sharing Policy published in the NIH Guide on February 26, 2003. In accord with the NIH guidelines, a summary, deidentified NAC Attack data set will be made available through submission of the dataset to a government or other health research database. In accord with the NIH guidelines, a summary, de-identified NAC Attack data set will be made available through submission of the dataset to a government or other health research database. We will share individual-participant level data (IPD) together with their associated data dictionaries. The data will be made available 1 year after publication of the primary findings of the study, in a de-identified format. In addition to the IPD data set, the researchers will share the trial protocol including the statistical analysis plan, data collections forms, and analytic codes used in the main reports of the trial. Sharing of the genetic data will follow NIH’s GDS Policy (https://grants.nih.gov/grants/guide/notice-files/NOT-OD-14-124.html). Within 12 months after the study closure, genomic data will be submitted to the NIH maintained Database of Genotype and Phenotype (dbGaP) as Controlled-access data.

The rights and privacy of people who participated in the Study will be protected at all times by stripping all identifiers including study IDs that could lead to disclosing the identity of individual research participants from the data. This commitment to privacy-protected data sharing will be incorporated in all levels of data sharing activities. In addition, redacted clinical study reports, retinal images, and other summary reports may be provided to researchers upon approval of their requests by the study leadership. The requesting researchers will be required to sign a data use agreement before they are given access to study reports or images.

### COMPETING INTERESTS

The authors declare that they have no competing interests.

### FUNDING

The NAC Attack trial is funded by the following grants from the NEI of the NIH:

UG1EY033286, PI- Campochiaro, Wilmer Eye Institute of the School of Medicine of Johns Hopkins University, Maryland, USA.

UG1EY033293, PI – Kong, Wilmer Eye Institute of the School of Medicine of Johns Hopkins University, Maryland, USA

UG1EY033287, PIs – Jaffe, Farsiu. Duke Eye Center of Duke University, North Carolina, USA

UG1EY033292, PIs- Duncan, Department of Ophthalmology, University of California San Francisco; Carroll, Department of Ophthalmology and Visual Sciences, Medical University of Wisconsin, USA.

The Manual of Procedures(MOP) and Protocol of the NAC Attack trial were developed under funding of R34EY031429 (PIs: Kong/Campochiaro) awarded by NEI.

The funding agency played no role in the design of the study and collection, analysis, and interpretation of data and in writing the manuscript.

### AUTHOR’S CONTRIBUTIONS

XK led to receive the funding for trial planning and developed the MOP and Protocol with review and edits from PAC. XK, PAC, JD, JC, and GJ obtained funding for trial conduct. XK, PAC, MS, MJK, DW, GH, FI, FN, YL, KD, and BH developed study procedures and data collection forms. KD developed technical infrastructure to ensure implementation of all procedures. JD and JC developed all procedures and data collection forms related to the AOLSO sub-study with input from KD. GJ led to development of the OCT Reading Center related procedures and manuals. DB provided significant input on the FST testing procedures. EIT provided significant input on preliminary data collection leading to the NAC Attack funding. XK and PAC designed the study. XK developed the statistical design and analysis plan. LHM oversaw and provided important comments on the statistics plan. RH developed genetics data analysis plan. RM led ethical and regulatory approvals in Mexico (though the study eventually did not enroll patients in Mexico). XK led ethical and regulatory approvals in countries with active trial activities. The NAC Attack Study Investigative Group provided input on the Protocol and study procedures. The complete list of the Study Investigative Group is available in Supplemental Material 4.

## Supporting information

Supplemental 1 SPIRIT checklist

Supplemental 2 Protocol and Protocol addendums

Supplemental 3 list of sites

Supplemental 4 List of Study Group 7.21.2025

## Data Availability

Not applicable because the manuscript does not report data.

## ACKNOWLEDGEMENTS

We sincerely thank Zambon SpA, Italy for donating study drug including both active drug and placebo, and thank the Data and Safety Monitoring Committee, all study site personnel, all current and future study participants, and patient advocacy groups including USHER 2020 Foundation and Retina UK for their efforts, support of and dedication to the study. We thank the National Eye Institute (NEI) for funding the study and for providing funding for the planning phase (R34EY031429, PIs: Kong and Campochiaro) which allowed us to develop the design, logistics plan and Manual of Procedures. The planning phase was also made possible by the bridge funding (PI: Kong) provided by the unrestricted grant from Research to Prevent Blindness to the Wilmer Eye Institute. We are deeply grateful for the guidance from the NEI program officials since the early conception of the study. We also greatly appreciate the advice and support from the Wilmer Eye Institute leadership as well as research administration support in managing the subawards from the Johns Hopkins School of Medicine and the Wilmer Eye Institute.

## Notes

### Competing Interest Statement

The authors have declared no competing interest.

### Clinical Trial

NCT05537220

### Funding Statement

National Eye Institute:
1. R34EY031429 Dr Xiangrong Kong
2. UG1EY033286 Dr. Peter A. Campochiaro, MD
3. UG1EY033293 Dr Xiangrong Kong
4. UG1EY033287 Dr. Glenn Jaffe, MD
5. UG1EY033292 Dr. Jacque Duncan, MD

### Author Declarations

The study was approved by the following competent authorities and Institutional Review Board (IRB) or Research Ethics Committees: in the United States (US)- Johns Hopkins University single IRB (sIRB) with reliance agreements of the local IRBs of participating sites in the USA and the US Federal Drug and Food Administration; in Austria, Germany and the Netherlands: the European Medicines Agency; in Canada: Health Canada and Research Ethics Board of McGill University Health Center; in Switzerland: Swissmedic and Swissthics; and in the United Kingdom: the Medicines and Healthcare products Regulatory Agency and the Health Research Authority.

## References

1. Bunker CH, Berson EL, Bromley WC, Hayes RP, Roderick TH. Prevalence of retinitis pigmentosa in Maine. Am J Ophthalmol. 1984;97:357–65.

2. Haim M. Epidemiology of retinitis pigmentosa in Denmark. Acta Ophalmol Scand Suppl. 2002;233:1–34.

3. Rivolta C, Sharon D, DeAngelis MM, Dryja TP. Retinitis pigmentosa and allied diseases: numerous diseases, genes, and inheritance patterns. Hum Mol Genet. 2002;11:1219–27.

4. Boughman JA, Vernon M, Shaver KA. Usher syndrome: definition and estimated of prevalence from two high-risk populations. J Chronic Dis. 1983;36:595–603.

5. Dryja TP, McGee tL, Reichel E, Hahn LB, Cowley GS, Yandell DW, et al. A point mutation of the rhodopsin gene in one form of retinitis pigmentosa. Nature. 1990;343:364–6.

6. Daiger SP, Sullivan LS, Bowne SJ. Genes and mutations causing retinitis pigmentosa. Clin Genet. 2013;84:132–41.

7. Yu DY, Cringle SJ, Su EN, Yu PK. Intraretinal oxygen levels before and after photoreceptor loss in the RCS rat. Invest Ophthalmol Vis Sci. 2000;41:3999–4006.

8. Usui S, Oveson BC, Lee SY, Jo YJ, Yoshida T, Miki A, et al. NADPH oxidase plays a central role in cone cell death in retinitis pigmentosa J Neurochem 2009;110 1028–37.

9. Shen J, Yan X, Dong A, Petters RM, Peng Y-W, Wong F, et al. Oxidative damage is a potential cause of cone cell death in retinitis pigmentosa. J Cell Physiol. 2005;203(3):457–64.

10. Komeima K, Usui S, Shen J, Rogers BS, Campochiaro PA. Blockade of neuronal nitric oxide synthase reduces cone cell death in a model of retinitis pigmentosa. Free Radic Biol Med. 2008;45:905–12.

11. Komeima K, Rogers BS, Lu L, Campochiaro PA. Antioxidants reduce cone cell death in a model of retinitis pigmentosa. Proc Natl Acad Sci USA. 2006;103(38):11300–5.

12. Komeima K, Rogers BS, Campochiaro PA. Antioxidants slow photoreceptor cell death in mouse models of retinitis pigmentosa. J Cell Physiol. 2007;213(3):809–15.

13. Usui S, Komeima K, Lee SY, Jo Y-J, Ueno S, Rogers BS, et al. Increased expression of catalase and superoxide dismutase 2 reduces cone cell death in retinitis pigmentosa. Molec Ther. 2009;17:778–86.

14. Usui S, Oveson BC, Iwase T, Lu L, Lee SY, Jo YJ, et al. Overexpression of SOD in retina: need for increase in H_2_O_2_-detoxifying enzyme in same cellular compartment. Free Radic Biol Med. 2011;51:1347–54.

15. Petit L, Ma S, Cipi J, Cheng SY, Zeiger M, Hay N, et al. Aerobic glycolysis is essential for normal rod function and controls secondary cone death in retinitis pigmentosa. Cell Rep. 2018;23(9):2629–42.

16. Ait-Ali N, Fridlich R, Millet-Puel G, Clerin E, Delalande F, Jaillard C, et al. Rod-derived cone viability factor promotes cone survival by stimulating aerobic glycolysis. Cell. 2015;161:817–32.

17. Campochiaro PA, Mir TA. The mechanism of cone cell death in retinitis pigmentosa. Prog Retin Eye Res. 2018;62(1):24–37.

18. Aldini G, Altomare A, Baron G, Vistoli G, Carini M, Borsani L, et al. N-Acetylcysteine as an antioxidant and disulphide breaking agent: the reasons why. Free Radic Res. 2018;52(7):751–62.

19. Prescott LF, Park J, Ballantyne A, Adriaenssens P, Proudfoot AT. Treatment of paracetamol (acetaminophen) poisoning with N-acetylcysteine. Lancet. 1977;2:432–4.

20. Smilkstein MJ, Knapp GL, Kulig KW, Rumack BH. Efficacy of oral N-acetylcysteine in the treatment of acetominophen overdose. Analysis of the national multicenter study (1976-1985). N Eng J Med. 1988;319:1557–62.

21. Lee SY, Usui S, Zafar AB, Oveson BC, Jo YJ, Lu L, et al. N-acetylcysteine promotes long term survival of cones in a model of retinitis pigmentosa. j Cell Physiol. 2011;226:1843–9

22. Group ETDRSR. Photocoagulation for diabetic macular edema. Early Treatment Diabetic Retinopathy Study report number 1. Arch Ophthalmol. 1985;103(12):1796–806.

23. Chan AW, Tetzlaff JM, Gotzsche PC, Altman DG, Mann H, Berlin JA, et al. SPIRIT 2013 explanation and elaboration: guidance for protocols of clinical trials. BMJ. 2013;346:e7586. Epub 2013/01/11. doi: 10.1136/bmj.e7586. PubMed PMID: 23303884; PubMed Central PMCID: PMCPMC3541470.

24. FDA. E9(R1) Statistical Principles for Clinical Trials: Addendum: Estimands and Sensitivity Analysis in Clinical Trials. https://www.fda.gov/regulatory-information/search-fda-guidance-documents/e9r1-statistical-principles-clinical-trials-addendum-estimands-and-sensitivity-analysis-clinical. 2019.

25. O’Neal T, Luther E. Retinitis Pigmentosa. [Updated 2019 Apr 10]. Treasure Island (FL): StatPearls Publishing; 2019. Available from: https://www.ncbi.nlm.nih.gov/books/NBK519518/.

26. Sadda SR, Chakravarthy U, Birch DG, Staurenghi G, Henry EC, Brittain C. Clinical Endpoints for the Study of Geographic Atrophy Secondary to Age-Related Macular Degeneration. Retina. 2016;36(10):1806–22. doi: 10.1097/IAE.0000000000001283. PubMed PMID: 27652913; PubMed Central PMCID: PMCPMC5384792.

27. Linder B, Dill H, Hirmer A, Brocher J, Lee GP, Mathavan S, et al. Systemic splicing factor deficiency causes tissue-specific defects: a zebrafish model for retinitis pigmentosa. Human molecular genetics. 2011;20(2):368–77. doi: 10.1093/hmg/ddq473. PubMed PMID: 21051334.

28. Csaky K, Ferris F, 3rd, Chew EY, Nair P, Cheetham JK, Duncan JL. Report From the NEI/FDA Endpoints Workshop on Age-Related Macular Degeneration and Inherited Retinal Diseases. Investigative ophthalmology & visual science. 2017;58(9):3456–63. doi: 10.1167/iovs.17-22339. PubMed PMID: 28702674; PubMed Central PMCID: PMC5961066.

29. Campochiaro PA, Iftikhar M, Hafiz G, Akhlaq A, Tsai G, Wehling D, et al. Oral N-acetylcysteine improves cone function in retinitis pigmentosa patients in phase I trial. J Clin Invest. 2020;130(3):1527–41. Epub 2019/12/06. doi: 10.1172/JCI132990. PubMed PMID: 31805012; PubMed Central PMCID: PMCPMC7269599.

30. Birch DG, Cheng P, Duncan JL, Ayala AR, Maguire MG, Audo I, et al. The RUSH2A Study: Best-Corrected Visual Acuity, Full-Field Electroretinography Amplitudes, and Full-Field Stimulus Thresholds at Baseline. Translational vision science & technology. 2020;9(11):9. Epub 2020/11/03. doi: 10.1167/tvst.9.11.9. PubMed PMID: 33133772; PubMed Central PMCID: PMCPMC7552938.

31. Ramachandran R, C XC, Lee D, B CE, Locke KG, D GB, et al. Reliability of a Manual Procedure for Marking the EZ Endpoint Location in Patients with Retinitis Pigmentosa. Translational vision science & technology. 2016;5(3):6. Epub 2016/05/27. doi: 10.1167/tvst.5.3.6. PubMed PMID: 27226930; PubMed Central PMCID: PMCPMC4874452.

32. Sujirakul T, Lin MK, Duong J, Wei Y, Lopez-Pintado S, Tsang SH. Multimodal Imaging of Central Retinal Disease Progression in a 2-Year Mean Follow-up of Retinitis Pigmentosa. American journal of ophthalmology. 2015;160(4):786–98 e4. Epub 2015/07/15. doi: 10.1016/j.ajo.2015.06.032. PubMed PMID: 26164827; PubMed Central PMCID: PMCPMC4754981.

33. Iftikhar M, Kherani S, Kaur R, Lemus M, Nefalar A, Usmani B, et al. Progression of Retinitis Pigmentosa as Measured on Microperimetry: The PREP-1 Study. Ophthalmol Retina. 2018;2(5):502–7. doi: 10.1016/j.oret.2017.09.008. PubMed PMID: 31047333.

34. Tee JJL, Carroll J, Webster AR, Michaelides M. Quantitative Analysis of Retinal Structure Using Spectral-Domain Optical Coherence Tomography in RPGR-Associated Retinopathy. American journal of ophthalmology. 2017;178:18–26. Epub 2017/03/23. doi: 10.1016/j.ajo.2017.03.012. PubMed PMID: 28322733; PubMed Central PMCID: PMCPMC5451208.

35. FDA. Guidance for Industry E9 Statistical Principles for Clinical Trials. http://www.fda.gov/cder/guidance/index.htm or http://www.fda.gov/cber/guidelines.htm;. 1998.

36. Yang Y. Sensitivity Analysis in Multiple Imputation for Missing Data.. 2014.

37. Permutt T. Sensitivity analysis for missing data in regulatory submissions. Statistics in medicine. 2016;35(17):2876–9. doi: 10.1002/sim.6753. PubMed PMID: WOS:000379983000003.

38. Austin PC. An Introduction to Propensity Score Methods for Reducing the Effects of Confounding in Observational Studies. Multivar Behav Res. 2011;46(3):399–424. doi: Pii 938470000 10.1080/00273171.2011.568786. PubMed PMID: WOS:000291533400002.

39. Tee JJL, Yang Y, Kalitzeos A, Webster A, Bainbridge J, Michaelides M. Natural History Study of Retinal Structure, Progression, and Symmetry Using Ellipzoid Zone Metrics in RPGR-Associated Retinopathy. American journal of ophthalmology. 2019;198:111–23. doi: 10.1016/j.ajo.2018.10.003. PubMed PMID: 30312579; PubMed Central PMCID: PMC6355316.

40. Tee JJL, Yang Y, Kalitzeos A, Webster A, Bainbridge J, Weleber RG, et al. Characterization of Visual Function, Interocular Variability and Progression Using Static Perimetry-Derived Metrics in RPGR-Associated Retinopathy. Investigative ophthalmology & visual science. 2018;59(6):2422–36. Epub 2018/05/31. doi: 10.1167/iovs.17-23739. PubMed PMID: 29847648; PubMed Central PMCID: PMCPMC5947973.

41. Birch DG, Fish GE. Rod ERGs in retinitis pigmentosa and cone-rod degeneration. Investigative ophthalmology & visual science. 1987;28(1):140–50. PubMed PMID: 3804644.

42. Birch DG, Wen Y, Locke K, Hood DC. Rod sensitivity, cone sensitivity, and photoreceptor layer thickness in retinal degenerative diseases. Investigative ophthalmology & visual science. 2011;52(10):7141–7. Epub 20110909. doi: 10.1167/iovs.11-7509. PubMed PMID: 21810977; PubMed Central PMCID: PMCPMC3207717.

43. Vickerstaff V, Omar RZ, Ambler G. Methods to adjust for multiple comparisons in the analysis and sample size calculation of randomised controlled trials with multiple primary outcomes. BMC Med Res Methodol. 2019;19(1):129. Epub 2019/06/23. doi: 10.1186/s12874-019-0754-4. PubMed PMID: 31226934; PubMed Central PMCID: PMCPMC6588937.

44. Piantadosi S. Clinical trials : a methodologic perspective 2005.

45. Campochiaro PA, Iftikhar M, Hafiz G, Akhlaq A, Tsai G, Wehling D, et al. Oral N-acetylcysteine improves cone function in retinitis pigmentosa patients in phase 1 trial. J Clin Invest. 2019;pii: 132990. doi: 10.1172/JCI132990.

46. Kong X, Hafiz G, Wehling D, Akhlaq A, Campochiaro PA. Locus level changes in macular sensitivity in patient with retinitis pigmentosa treated with oral N-acetylcysteine. Am J Ophthalmol. 2020;Aug; S0002-9394(20)30421-9. doi: 10.1016/j.ajo.2020.08.002.

47. Kong X, Choy K, Khan M, Lu Y, Wehling D, Campochiaro P, et al. Reproducibility of ellipsoid zone (EZ) parameters measured under different OCT scan settings in retinitis pigmentosa (RP). ARVO Annual Meeting; New Orleans, Louisiana, USA 2023.

48. Roberts PA, Gaffney EA, Whiteley JP, Luthert PJ, Foss AJE, Byrne HM. Predictive Mathematical Models for the Spread and Treatment of Hyperoxia-induced Photoreceptor Degeneration in Retinitis Pigmentosa. Invest Ophthalmol Vis Sci. 2018;59(3):1238–49. doi: 10.1167/iovs.17-23177. PubMed PMID: 29625444.

