## Supplemental 1 SPIRIT checklist for "Protocol for NAC Attack, a phase-3, multicenter randomized, parallel, double masked, placebo controlled trial evaluating the efficacy and safety of oral N-acetylcysteine (NAC) in patients with retinitis pigmentosa"

### SPIRIT checklist for the NAC Attack Trial

All the page and Chapter/section numbers in the checklist refer to the page numbers in the complete study protocol in Supporting Information file S1 Protocol: Full Study Protocol

| | | Reporting Item | Page and Line Number | Reason if not applicable |
|---|---|---|---|---|
| **Administrative information** | | | | |
| Title | #1 | Descriptive title identifying the study design, population, interventions, and, if applicable, trial acronym | 1 | |
| Trial registration | #2a | Trial identifier and registry name. If not yet registered, name of intended registry | 1 | |
| Trial registration: data set | #2b | All items from the World Health Organization Trial Registration Data Set | | Not registered with WHO Trial Registration |
| Protocol version | #3 | Date and version identifier | Page1 -146 | |
| Funding | #4 | Sources and types of financial, material, and other support | Page 130 Section 10.7.1 | |
| Roles and responsibilities: contributorship | #5a | Names, affiliations, and roles of protocol contributors | | |
| Roles and responsibilities: sponsor contact information | #5b | Name and contact information for the trial sponsor | Page 1 | |

| | | | |
|---|---|---|---|
| Roles and responsibilities: sponsor and funder | #5c | Role of study sponsor and funders, if any, in study design; collection, management, analysis, and interpretation of data; writing of the report; and the decision to submit the report for publication, including whether they will have ultimate authority over any of these activities | |
| Roles and responsibilities: committees | #5d | Composition, roles, and responsibilities of the coordinating centre, steering committee, endpoint adjudication committee, data management team, and other individuals or groups overseeing the trial, if applicable (see Item 21a for data monitoring committee) | Page 128-132 Chapter 10 |
| **Introduction** | | | |
| Background and rationale | #6a | Description of research question and justification for undertaking the trial, including summary of relevant studies (published and unpublished) examining benefits and harms for each intervention | Page 23-26 Chapter. 1<br>Page 30-37 Chapter 3 |
| Background and rationale: choice of comparators | #6b | Explanation for choice of comparators | Page 37 section 3.3.7 |

| | | | | |
|---|---|---|---|---|
| Objectives | #7 | Specific objectives or hypotheses | Page 28-29 Chapter 2 | |
| Trial design | #8 | Description of trial design including type of trial (eg, parallel group, crossover, factorial, single group), allocation ratio, and framework (eg, superiority, equivalence, non-inferiority, exploratory) | Page 30-37 Chapter 3 | |
| **Methods: Participants, interventions, and outcomes** | | | | |
| Study setting | #9 | Description of study settings (eg, community clinic, academic hospital) and list of countries where data will be collected. Reference to where list of study sites can be obtained | N/A | Not in the protocol. Available on study website and on ClinicalTrial.gov trial registration page. |
| Eligibility criteria | #10 | Inclusion and exclusion criteria for participants. If applicable, eligibility criteria for study centres and individuals who will perform the interventions (eg, surgeons, psychotherapists) | Page 38 Section 4.1 | |
| Interventions: description | #11a | Interventions for each group with sufficient detail to allow replication, including how and when they will be administered | Page 42 Section 4.4 | |
| Interventions: modifications | #11b | Criteria for discontinuing or modifying allocated | Page 60 Section 5.1.2 | |

| | | | |
|---|---|---|---|
| | | interventions for a given trial participant (eg, drug dose change in response to harms, participant request, or improving / worsening disease) | |
| Interventions: adherance | #11c | Strategies to improve adherence to intervention protocols, and any procedures for monitoring adherence (eg, drug tablet return; laboratory tests) | Page 46 Section 4.8.6<br><br>Page 54-57 Section 4.14 |
| Interventions: concomitant care | #11d | Relevant concomitant care and interventions that are permitted or prohibited during the trial | Page 43 Section 4.7 |
| Outcomes | #12 | Primary, secondary, and other outcomes, including the specific measurement variable (eg, systolic blood pressure), analysis metric (eg, change from baseline, final value, time to event), method of aggregation (eg, median, proportion), and time point for each outcome. Explanation of the clinical relevance of chosen efficacy and harm outcomes is strongly recommended | Page 77-88 |
| Participant timeline | #13 | Time schedule of enrolment, interventions (including any run-ins and washouts), assessments, and visits for | Page 16 Figure 1 |

| | | | |
|---|---|---|---|
| | | participants. A schematic diagram is highly recommended (see Figure) | |
| Sample size | #14 | Estimated number of participants needed to achieve study objectives and how it was determined, including clinical and statistical assumptions supporting any sample size calculations | Page 88-99 |
| Recruitment | #15 | Strategies for achieving adequate participant enrolment to reach target sample size | Page 44 |
| **Methods: Assignment of interventions (for controlled trials)** | | | |
| Allocation: sequence generation | #16a | Method of generating the allocation sequence (eg, computer-generated random numbers), and list of any factors for stratification. To reduce predictability of a random sequence, details of any planned restriction (eg, blocking) should be provided in a separate document that is unavailable to those who enrol participants or assign interventions | Page 31 Section 3.1.3<br><br>Page 40 Section 4.2 |
| Allocation concealment mechanism | #16b | Mechanism of implementing the allocation sequence (eg, central telephone; sequentially numbered, opaque, sealed envelopes), describing any steps to | Page 40 Section 4.2 |

| | | | |
|---|---|---|---|
| | | conceal the sequence until interventions are assigned | |
| Allocation: implementation | #16c | Who will generate the allocation sequence, who will enrol participants, and who will assign participants to interventions | Page 31 Section 3.1.3<br><br>Page 40 Section 4.2 |
| Blinding (masking) | #17a | Who will be blinded after assignment to interventions (eg, trial participants, care providers, outcome assessors, data analysts), and how | Page 40 Section 4.2 |
| Blinding (masking): emergency unblinding | #17b | If blinded, circumstances under which unblinding is permissible, and procedure for revealing a participant's allocated intervention during the trial | Protocol Addendum Section 4.12.6 |
| **Methods: Data collection, management, and analysis** | | | |
| Data collection plan | #18a | Plans for assessment and collection of outcome, baseline, and other trial data, including any related processes to promote data quality (eg, duplicate measurements, training of assessors) and a description of study instruments (eg, questionnaires, laboratory tests) along with their reliability and validity, if known. Reference to where data collection forms can be found, if not in the protocol | Page 112 Chapter 7 |

| | | | |
|---|---|---|---|
| Data collection plan: retention | #18b | Plans to promote participant retention and complete follow-up, including list of any outcome data to be collected for participants who discontinue or deviate from intervention protocols | Page 56 Section 4.14.6<br>Page 117 Section 7.5 |
| Data management | #19 | Plans for data entry, coding, security, and storage, including any related processes to promote data quality (eg, double data entry; range checks for data values). Reference to where details of data management procedures can be found, if not in the protocol | Page 112-117 |
| Statistics: outcomes | #20a | Statistical methods for analysing primary and secondary outcomes. Reference to where other details of the statistical analysis plan can be found, if not in the protocol | Page 97 |
| Statistics: additional analyses | #20b | Methods for any additional analyses (eg, subgroup and adjusted analyses) | Page 104-107 |
| Statistics: analysis population and missing data | #20c | Definition of analysis population relating to protocol non-adherence (eg, as randomised analysis), and any statistical methods to handle missing data (eg, multiple imputation) | Page 84 |

| | | | |
|---|---|---|---|
| **Methods: Monitoring** | | | |
| Data monitoring: formal committee | #21a | Composition of data monitoring committee (DMC); summary of its role and reporting structure; statement of whether it is independent from the sponsor and competing interests; and reference to where further details about its charter can be found, if not in the protocol. Alternatively, an explanation of why a DMC is not needed | Page 132 |
| Data monitoring: interim analysis | #21b | Description of any interim analyses and stopping guidelines, including who will have access to these interim results and make the final decision to terminate the trial | Page 106 |
| Harms | #22 | Plans for collecting, assessing, reporting, and managing solicited and spontaneously reported adverse events and other unintended effects of trial interventions or trial conduct | Page 59-77 |
| Auditing | #23 | Frequency and procedures for auditing trial conduct, if any, and whether the process will be independent from investigators and the sponsor | Page 125 |
| **Ethics and dissemination** | | | |

| | | | |
|---|---|---|---|
| Research ethics approval | #24 | Plans for seeking research ethics committee / institutional review board (REC / IRB) approval | Page 118 |
| Protocol amendments | #25 | Plans for communicating important protocol modifications (eg, changes to eligibility criteria, outcomes, analyses) to relevant parties (eg, investigators, REC / IRBs, trial participants, trial registries, journals, regulators) | Page 126 |
| Consent or assent | #26a | Who will obtain informed consent or assent from potential trial participants or authorised surrogates, and how (see Item 32) | Page 118 |
| Consent or assent: ancillary studies | #26b | Additional consent provisions for collection and use of participant data and biological specimens in ancillary studies, if applicable | Page 118 |
| Confidentiality | #27 | How personal information about potential and enrolled participants will be collected, shared, and maintained in order to protect confidentiality before, during, and after the trial | Page 120 |
| Declaration of interests | #28 | Financial and other competing interests for principal investigators for the overall trial and each study site | Page 123 |

| | | | | |
|---|---|---|---|---|
| Data access | #29 | Statement of who will have access to the final trial dataset, and disclosure of contractual agreements that limit such access for investigators | Page 138 | |
| Ancillary and post trial care | #30 | Provisions, if any, for ancillary and post-trial care, and for compensation to those who suffer harm from trial participation | N/A | |
| Dissemination policy: trial results | #31a | Plans for investigators and sponsor to communicate trial results to participants, healthcare professionals, the public, and other relevant groups (eg, via publication, reporting in results databases, or other data sharing arrangements), including any publication restrictions | Page 134 | |
| Dissemination policy: authorship | #31b | Authorship eligibility guidelines and any intended use of professional writers | Page 134 | |
| Dissemination policy: reproducible research | #31c | Plans, if any, for granting public access to the full protocol, participant-level dataset, and statistical code | Page 138 | |
| **Appendices** | | | | |
| Informed consent materials | #32 | Model consent form and other related documentation given | | confidential |

| | | | |
|---|---|---|---|
| | | to participants and authorised surrogates | |
| Biological specimens | #33 | Plans for collection, laboratory evaluation, and storage of biological specimens for genetic or molecular analysis in the current trial and for future use in ancillary studies, if applicable | Page 49 |
