## Supplemental 2 Protocol and Protocol addendums for "Protocol for NAC Attack, a phase-3, multicenter randomized, parallel, double masked, placebo controlled trial evaluating the efficacy and safety of oral N-acetylcysteine (NAC) in patients with retinitis pigmentosa"

Supporting Material-2 NAC Attack Study Protocol and Protocol Addendums

- Protocol V2.0 2023-07-13
- Protocol Addendum 2024-01-01 to Protocol V2.0
- Protocol Addendum 2024-10-01 to Protocol V2.0
- Protocol Addendum 2025-02-20 to Protocol V2.0

### PROTOCOL

**Title:** NAC Attack, A Phase III, Multicenter, Randomized, Parallel, Double Masked, Placebo-Controlled Study Evaluating the Efficacy and Safety of Oral N-Acetylcysteine in Patients with Retinitis Pigmentosa

**Short Title:** Oral N-acetylcysteine for Retinitis Pigmentosa (NAC Attack)

**FDA IND Number:** IND133525

**ClinicalTrials.gov Identifier:** NCT05537220

**EU CT Number:** 2022-501438-46-00

**Sponsor Protocol Number:** JHU-IRB00337490

**Test Product:** N-acetylcysteine effervescent tablets (Fluimucil)

**Funder:** National Eye Institute of the National Institutes of Health, United States Department of Health and Human Services

**Sponsor:**

| | |
|---|---|
| NAC Attack Study Chair<br>Peter A. Campochiaro MD<br>Eccles Professor of Ophthalmology and Neuroscience<br>815 Maumenee<br>The Wilmer Eye Institute<br>600 N. Wolfe Street<br>Baltimore, MD 21287<br>Johns Hopkins University<br><br>Phone number: 1-410-955-5106 | Coordinating Center Director<br>Xiangrong Kong, PhD,<br>Associate Professor of Ophthalmology, Biostatistics and Epidemiology<br>155b Woods<br>The Wilmer Eye Institute<br>600 N. Wolfe Street<br>Baltimore, MD 21287<br>Johns Hopkins University<br><br>Phone number: 1-410-614-4087 |

***The NAC Attack clinical trial is to be conducted in compliance with this protocol, with the Good Clinical Practices, and with EU Clinical Trial Regulation (No 536/2014).***

4.3.2. Study Medication Manufacture and Marketing Authorizations (MA) in Europe... 41

4.14.4. Quality Assurance via Clinical Site Monitoring ... 56
4.14.5. Ensuring Participant Study Medication Compliance ... 56
4.14.6. Clinical Site Measures to Prevent Drop-outs and Missed Visits ... 56
4.14.7. Quality Assurance Methods Ensuring Data Integrity and Quality ... 57
4.15. Treatment, Patient, Study, and Site Discontinuation ... 57
4.15.1. Permanent Study Treatment Discontinuation ... 57
4.15.2. Study Treatment Dose Modification ... 57
4.15.3. Participant Discontinuation from Study ... 58
4.15.4. Study Discontinuation ... 58
4.15.5. Site Discontinuation ... 58
**5. Assessment of Safety** ... 59
5.1. Overview of Safety Plan ... 59
5.1.1. Identified Risks Associated with Oral NAC in Patients with RP ... 59
5.1.1.1. Drug-related AEs in the FIGHT RP study ... 59
5.1.1.2. Gastrointestinal (GI) Symptoms ... 60
5.1.1.3. Exacerbation of Hypertension ... 60
5.1.1.4. Caution Using Cough Suppressant in Subjects with Upper Airway Congestion ... 60
5.1.1.5. Alteration of Laboratory Tests ... 60
5.1.2. Management of Participants Who Experience AEs ... 60
5.1.2.1. Dose Modifications ... 60
5.1.2.2. Study Treatment Interruption ... 61
5.1.2.3. Recommended Management of Exacerbation of Hypertension ... 61
5.1.3. Emergency Medical Contact ... 61
5.2. Overview of Adverse Events Definitions ... 61
5.2.1. Adverse Events Definition ... 61
5.2.1.1. Diagnosis versus Signs and Symptoms ... 62
5.2.1.2. AEs that are Secondary to Other Events ... 62
5.2.1.3. Persistent or Recurrent AEs ... 62
5.2.1.4. Abnormal Laboratory Values ... 63
5.2.1.5. Abnormal Vital Sign Values ... 63
5.2.1.6. Abnormal Liver Function Tests ... 64
5.2.1.7. Deaths ... 64
5.2.1.8. Preexisting Medical Conditions ... 64

6.1.2.3. NAC Attack Primary Endpoint Derived from the Primary Outcome of EZ-width 80
6.1.3. Estimand-framework Attribute C: Possible Intercurrent Events in NAC Attack ... 82
6.3.1.3. Sample Size Calculation Assumptions: Proportion of Participants with Only One Eye Eligible89

10.5. Clinical Sites ... 129
10.6. Study Resource Centers ... 129
10.6.1. OCT Image Reading Center ... 129
10.6.2. AOSLO Image Reading Center ... 130
10.7. Other Trial Stakeholders ... 130
10.7.1. National Eye Institute ... 130
10.7.2. Drug Manufacturer ... 131
10.7.3. Drug Labeling and Distribution Center ... 131
10.7.4. Other Centers/Laboratories/Collaborators ... 131
10.8. Standing Committees ... 131
10.8.1. Executive Committee ... 131
10.8.2. Steering and Quality Assurance Committee ... 132
10.8.3. Investigative Group ... 132
10.8.4. Data and Safety Monitoring Committee (DSMC) ... 132
**11. Study Policies: Publicity, Publication, Ancillary Studies, and Prompt Reporting to IRB** 134
11.1. Publicity Policy ... 134
11.2. Publication Policy ... 134
11.2.1. Authorship ... 134
11.2.2. Manuscript Writing Teams ... 134
11.2.3. Manuscript Pre-Submission Review ... 135
11.3. Site's Local Use of Data from NAC Attack Participants ... 135
11.4. Acknowledgements ... 135
11.5. Clinical Trials Registration and Results Submission ... 135
11.6. Ancillary Studies ... 135
11.6.1. Definition of a NAC Attack Ancillary Study ... 136
11.6.2. Reasons for Requirement of Approval ... 136
11.6.4. Publication of Ancillary NAC Attack Results ... 136
11.7. Prompt Reporting to Coordinating Center and IRB ... 136
11.7.1. Reportable Events Requiring Prompt Reporting to IRB ... 137
11.8. Data and Resource Sharing ... 138
11.8.3. AOSLO Image and Data Sharing ... 140
11.8.3.1. Protocol Sharing Plan ... 140
11.8.3.2. Image and Data Sharing Plan ... 140

**References** .......... 142

### Protocol Acceptance Form

**Title:** NAC Attack, A Phase III, Multicenter, Randomized, Parallel, Double Masked, Placebo-Controlled Study Evaluating the Efficacy and Safety of Oral N-Acetylcysteine in Patients with Retinitis Pigmentosa

**Short Title:** al N-acetylcysteine for Retinitis Pigmentosa (NAC Attack)

**ClinicalTrials.gov Identifier:** NCT05537220

**FDA IND Number:** IND133525

**EU CT Number:** 2022-501438-46-00

**Sponsor Protocol Number:** JHU-IRB00337490

**Test Product:** N-acetylcysteine effervescent tablets (Fluimucil)

**I agree to conduct the study in accordance with the current protocol at the location below and my institution and I permit and cooperate with clinical trial related monitoring, audits and regulatory inspections, including provision of direct access to source data and documents.**

Clinical Site Name: City, State, Country

________________________________ ________________

________________________________

Site Principal Investigator's Name (print)

________________________________ ________________

Site Principal Investigator's Signature Date

Please retain the signed original of this form for your study files. Please return a copy of the signed form to the Coordinating Center.

### Protocol Synopsis

**Title:** NAC Attack, A Phase-3, Multicenter, Randomized, Double Masked, Placebo-Controlled Study evaluating the Efficacy and Safety of Oral N-Acetylcysteine in Patients with Retinitis Pigmentosa

**FDA IND Number:** IND133525
**ClinicalTrials.gov Identifier:** NCT05537220
**EU CT Number:** 2022-501438-46-00
**Sponsor Protocol Number:** JHU-IRB00337490
**Intervention Product:** N-acetylcysteine (NAC) effervescent tablets (Fluimucil)
**Objectives:** to evaluate the efficacy and safety of oral NAC in patients with retinitis pigmentosa (RP).

**Trial Design Summary**

| | |
|---|---|
| Main Eligibility Criteria | • Age ≥ 18 and ≤ 65 years<br>• Both eyes exhibit the RP phenotype with evidence of loss of night vision, gradual constriction of visual fields, and maintenance of visual acuity;<br>• BCVA ≥ 61 Early Treatment Diabetic Retinopathy Study (ETDRS) letter score (20/60 [6/18] or better Snellen equivalent)<br>• Gradable ellipsoid zone (EZ) width on the horizontal fovea SD-OCT scan is < 8000 µm and ≥ 1500 µm<br>• No evidence of cone-rod dystrophy or pattern dystrophy including focal areas of atrophy or pigmentary changes in the central macula<br>• Not having taken NAC in any form in the past 4 months |
| Treatment groups | Intervention of 1800 mg bid of NAC effervescent tablets vs. placebo. |
| Randomization | • Stratified randomization by Clinical Site.<br>• Randomization unit is by person. Both eyes will be enrolled if eligible.<br>• 2:1 randomization ratio of intervention vs. placebo. |
| Total Sample Size | Approximately 438, enrolled from approximately 30 Clinical Sites in the US, Canada, Mexico and Europe. |
| Primary Efficacy Objective | To determine if the progressive loss in EZ width measured as the cumulative loss of EZ (calculated as the area above the curve) between baseline and month (M) 45 is significantly less in eyes of participants taking NAC 1800 mg bid compared with that in eyes of participants taking placebo. |
| Secondary Efficacy Objectives | To assess the relative efficacy in eyes of participants taking NAC 1800 mg bid compared with eyes of participants taking placebo on the basis of the following endpoints:<br>• Change in mean macular sensitivity measured by microperimetry (MP) from baseline to M45.<br>• Change in best-corrected visual acuity (BCVA) measured by ETDRS protocol from baseline to M45. |
| Safety Objective | To evaluate the long-term safety and tolerability of oral NAC 1800mg bid for 45 months on the basis of the following endpoints:<br>• Incidence and severity of ocular adverse events (AEs)<br>• Incidence and severity of non-ocular AEs |

| | |
|---|---|
| Participant Follow-up Duration and Schedule | Total follow-up duration: 45 months<br>• In-clinic visits: M4.5, 9, 18, 27, 36, 40.5 and 45.<br>• Tele-visits with Investigator: M13.5, 22.5, and 31.5.<br>• Phone calls with Coordinator: M2.25, 6.75, 11.25, 15.75, 20.25, 24.75, 29.25, 33.75, 38.25, 42.75, i.e. every 2.25 months after an in-clinic or tele-visit. |
| Statistical Analysis of the Primary Efficacy Endpoint | The estimand of cumulative loss of EZ over 45 months of follow-up is operationally calculated as the sum of the Areas Above the observed EZ width Curve (AAC). The primary analysis on the estimand follows the Intent-to-Treat (ITT) principle and uses linear model with generalized estimating equation (GEE) to compare the AAC between the intervention and placebo arms while accounting for between-eye correlation. |
| Interim Analysis | When all participants complete their M27 follow-up, interim efficacy analysis will evaluate efficacy based on the primary endpoint to test whether there is significantly less cumulative loss of EZ-width over 27 months in the intervention group compared to the placebo group, using the O'Brien-Fleming alpha spending rule. |
| Study Medication Acquisition and Distribution | Study drug includes the intervention drug of NAC in the form of 600mg effervescent tablet and placebo of identical form. The Study Drug Labeling and Distribution Center repackages, labels and ships the study drug to each Clinical Site. |

#### Ethical considerations

There are potential risks associated with breach of confidentiality, the intervention, and study procedures. Nonetheless, any information that identifies an individual study patient will be retained in a secure and confidential manner at the local clinical site and such information will not be transferred to other trial entities. Therefore, the risk of loss of confidentiality is extremely low. The study procedures are all routine procedures in retina clinical practice. Strategies to protect again potential risks due to the intervention or study procedures include safety monitoring by a physician Medical Monitor, the Study Chairman, and an external, independent Data & Safety Monitoring Committee (DSMC) appointed by the National Eye Institute. Due to the slow progression of RP, the study requires a follow-up period of 45 months, which imposes a burden on participants and Clinical Site personnel. To minimize the burden on all stakeholders, in-clinic visits are limited to time points when a primary outcome measurement would be made and supplemented with virtual visits to assess safety and compliance. Such a design feature will minimize participant travel burden while maintaining rapport to minimize drop outs. Participants may benefit from information obtained from study visits or from some of the study procedures. Future patients who could be participants' family members may benefit from the important information about RP learned in the trial. Because NAC Attack will not restrict enrollment based upon whether a pathogenic mutation has been identified or what the mutation is if one has been identified, the study results will provide critical information that will have a major impact on *all* patients with RP, including patients themselves and their family members. Currently, there are no effective treatments for RP and most patients eventually lose all useful vision. The impending loss of vision in patients with RP is a major source of stress throughout life which can be compounded by fear of passing the disease to children. Finding an efficacious and safe treatment will reduce patient stress and improve self-image. Pre-clinical mechanistic studies and the phase-1 FIGHT RP dose-escalation single arm clinical trial suggest NAC may be a promising pharmacologic option for slowing photoreceptor and visual function loss, but whether this is the case can only be determined by a randomized, placebo-controlled clinical trial. Additionally, NAC Attack will generate data regarding the natural history of RP from patients with a diverse genetic background, allowing a better understanding of the genetic causes of RP and more insights into disease mechanisms. Such knowledge may contribute to the development of future new interventions. Therefore, any potential risks to participants are offset by potential benefits to participants, their family members and all patients with RP. The study benefit-risk ratio is very high.

### Figure 1 Schematic of NAC Attack Study Visits

Each participant will be followed for 45 months after randomization. Study visits including in-clinic visits, tele-visits with site Investigator, and phone calls with site Coordinator.

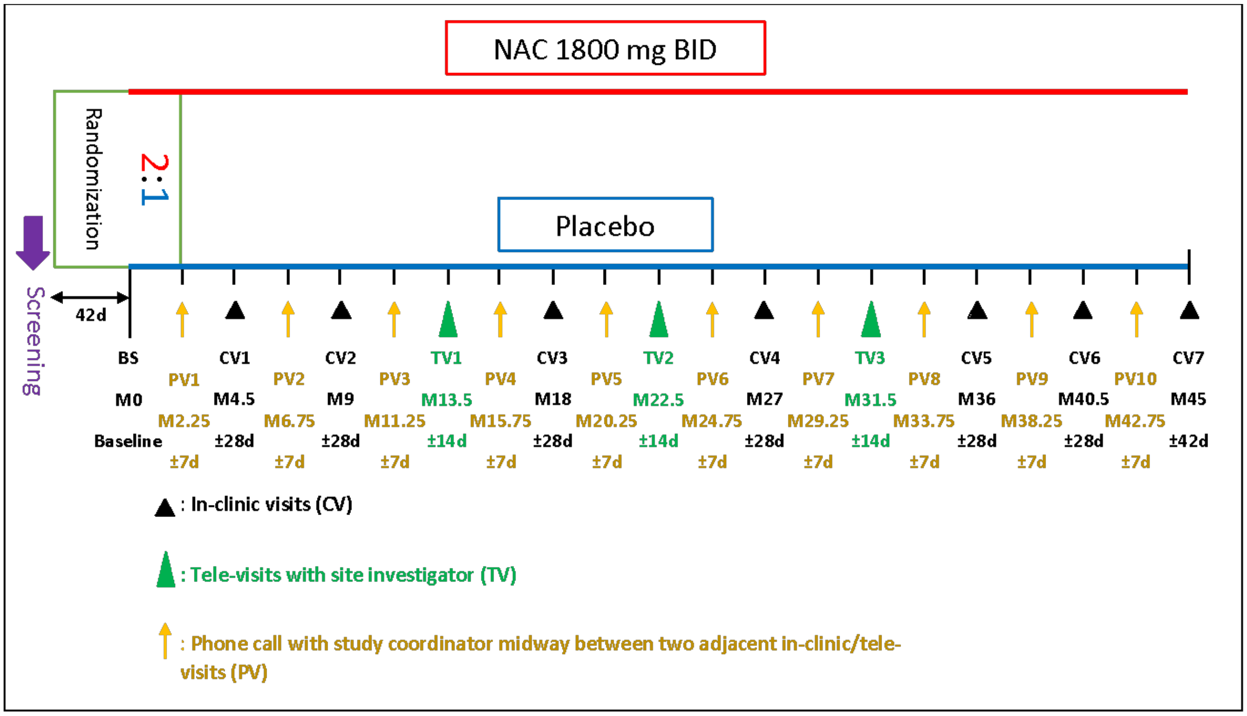

The allowed scheduling windows are:

- Screening visit to Baseline visit: within 42 days.
- For an in-clinic study follow-up visit: ±28 days except for the last visit at M45 which will have a wider scheduling window of ±42 days.
- For a tele-visit with Investigator: ±14 days
- For a phone call with coordinator: ±7 days

#### Table 1. Schedules of activities at study visits.

Study visits include in-clinic visits, tele-visits with investigators, and phone call visits with the site coordinators about every 2.25 months from an in-clinic or tele-visit. An early termination visit should follow activities scheduled for M45.

(SC: Screening. BL: Baseline. M: Month). *: a phone call visit with coordinator. ^: a tele-visit with site investigator.

| | SC | BL | M 2.25 * | M 4.5 | M 6.75 * | M 9 | M 11.25 * | M 13.5 ^ | M 15.75* | M 18 | M 20.25* | M 22.5 ^ | M 24.75 * | M 27 | M 29.25 * | M 31.5 ^ | M 33.75* | M 36 | M 38.25 * | M 40.5 | M 42.75* | M 45 |
|---|---|---|---|---|---|---|---|---|---|---|---|---|---|---|---|---|---|---|---|---|---|---|
| Informed Consent & Eligibility Criteria | X | | | | | | | | | | | | | | | | | | | | | |
| Patient history review for assessing eligibility | X | | | | | | | | | | | | | | | | | | | | | |
| Medical history/concurrent medications review | | X | X | X | X | X | X | X | X | X | X | X | X | X | X | X | X | X | X | X | X | X |
| Review of adverse events | | | X | X | X | X | X | X | X | X | X | X | X | X | X | X | X | X | X | X | X | X |
| Drug dispensing | | X | | | | X | | | | X | | | | X | | | | X | | | | |
| Compliance assessment/turning in both used/unused drug for drug reconciliation | | | | | | X | | | | X | | | | X | | | | X | | | | X |

| | | | | | | | | | | | | | | | | | | | | | | |
|---|---|---|---|---|---|---|---|---|---|---|---|---|---|---|---|---|---|---|---|---|---|---|
| Compliance assessment/turning in only used drug for drug reconciliation | | | | X | | | | | | | | | | | | | | | | | | |
| Vital Signs | X | X | | X | | X | | | | X | | | | X | | | | X | | | | X |
| BCVA | X | X | | X | | X | | | | X | | | | X | | | | X | | X | | X |
| IOP | X | X | | X | | X | | | | X | | | | X | | | | X | | X | | X |
| Slit Lamp Examination | X | X | | X | | X | | | | X | | | | X | | | | X | | | | X |
| Funduscopic Examination | X | X | | X | | X | | | | X | | | | X | | | | X | | | | X |
| MAIA microperimetry | X | X | | X | | X | | | | X | | | | X | | | | X | | X | | X |
| Spectral-domain OCT | X | X | | | | X | | | | X | | | | X | | | | X | | X | | X |
| Ultra-wide Fundus photograph | X | | | | | | | | | | | | | | | | | | | | | |
| Ultrawide fundus autofluorescence | X | | | | | | | | | | | | | X | | | | | | | | X |
| NEI-VFQ-25 | | X | | | | | | | | | | | | X | | | | | | | | X |
| Plasma NAC | | X | | | | X | | | | X | | | | X | | | | X | | | | X |
| Blood chemistry, hematology, liver function | X | | | X | | X | | | | X | | | | X | | | | X | | | | X |
| Urine test of pregnancy& | X | X | | | | | | | | | | | | | | | | | | | | |
| Genetic testing | | | | | | X^ | | | | | | | | | | | | | | | | |

| **SELECTED SITES ONLY** | | | | | | | | | | | | | | | | | | | | | | |
|---|---|---|---|---|---|---|---|---|---|---|---|---|---|---|---|---|---|---|---|---|---|---|
| AOSLO | | X | | | | X | | | | | | | | X | | | | | | | | X |
| FST | | X | | | | | | | | | | | | X | | | | | | | | X |

&: urine test for pregnancy may be done at follow-up visits as necessary.
^: If blood draw for genetic testing is not done at M9, it can be done at another follow-up visit upon communication with the Coordinating Center.

### List of Abbreviations

| Abbreviation | Definition |
|---|---|
| AAC | area above the curve |
| AEs | adverse events |
| ALT | alanine amino transaminase |
| APTC | Antiplatelet Trialists' Collaboration |
| AST | aspartate amino transaminase |
| AOSLO | adaptive optics scanning laser ophthalmoscopy |
| BCVA | best-corrected visual acuity |
| bid | twice a day |
| Co-I | co-investigator |
| COPD | chronic obstructive pulmonary disease |
| CRFs | Case Reporting Forms |
| CRO | clinical research organization |
| CST | central subfield thickness |
| dB | decibels |
| DCC | Data Coordinating Center |
| DSMC | Data and Safety Monitoring Committee |
| eCRF | electronic Case Report Form |
| EDC | electronic data capture |
| ETDRS | Early Treatment Diabetic Retinopathy Study |
| ERG | electroretinogram |
| EU | European Union |

| | |
|---|---|
| EZ | ellipsoid zone |
| FDA | Food and Drug Administration |
| FFA | fundus autofluorescence |
| FIGHT RP | phase 1/2 clinical trial testing NAC in patients with RP |
| FST | Full-field Stimulus Test |
| GEE | generalized estimating equation |
| GI | gastrointestinal |
| HIPPA | Health Insurance Portabiity and Accountability Act |
| IATA | International Air Transport Association |
| IB | Investigator’s Brochure |
| ICH | International Council for Harmonisation |
| ICTR | Johns Hopkins Institute for Clinical and Translational Research |
| IND | Investigation New Drug (Application) |
| IOP | intraocular pressure |
| IRB | Institutional Review Board |
| IWRS | Interactive Web-based Responsive System |
| JHU | Johns Hopkins University |
| mRNA | messenger ribonucleic acid |
| M | month |
| MedDRA | Medical Dictionary for Regulatory Activities |
| MP | microperimetry |
| NAC | n-acetylcysteine |
| NEI | National Eye Institute |

| | |
|---|---|
| NIH | National Institutes of Health |
| PAG | Patient advocacy group |
| PI | principal investigator |
| PK | pharmacokinetic |
| PRPFs | pre-mRNA processing factors |
| PRO | patient-reported outcomes |
| PDE6b | beta subunit of cyclic guanidine monophosphate rod phosphodiesterase gene |
| rd1 | retinal degeneration 1 mouse model of RP |
| rd10 | retinal degeneration 10 mouse model of RP |
| RBCs | red blood cells |
| REDCap | Research Electronic Data Capture |
| RNS | reactive nitrogen species |
| ROS | reactive oxygen species |
| RP | retinitis pigmentosa |
| RHO | rhodopsin gene |
| SAE | serious adverse event |
| SD-OCT | spectral domain-optical coherence tomography |
| SMS | short message service |
| tid | Three time a day |
| ULN | upper limit of normal |
| USA | United States of America |
| UWF | ultra-widefield |
| WBCs | white blood cells |

### 1. Background

#### 1.1. Clinical features and disease progression in retinitis pigmentosa (RP)

RP is a disease in which one of several different mutations differentially causes degeneration of rod photoreceptors while sparing cone photoreceptors. It occurs in roughly 1:4000 individuals and the inheritance pattern may be simplex (only one family member affected) or autosomal recessive (about 60%), autosomal dominant (about 30%), or X-linked recessive (about 10%).[1-3] Disease is usually limited to the eye, but in about 20-30% of RP patients, retinal degeneration occurs as part of a syndrome with associated non-ocular disease. Usher syndrome in which there is hearing loss associated with retinal degeneration, is particularly common accounting for 10-20% of RP cases.[4]

The loss of rod photoreceptors results in poor vision in dim illumination (night blindness), but does not affect most activities of daily life including reading or driving. However, after most rod photoreceptors are eliminated, cone photoreceptors begin to die. The drop out of cones usually begins in the mid-periphery of the retina and extends peripherally and centrally. At this stage, patients have mid-peripheral scotomata that coalesce into a ring which gradually enlarges and patients may complain that things or people in the environment may disappear and pop into view as images pass through the scotomata. As cone cell death continues to progress more centrally, patients become aware of constriction of visual fields. As the visual fields constrict, the level of disability increases but visual acuity is maintained until late. In some patients, visual acuity is reduced by posterior subcapsular cataracts which can be corrected by surgery, or by the occurrence of cystoid spaces in the macula of one or both eyes. The cystoid spaces can worsen or improve spontaneously making it difficult to assess the effect of treatments. Systemic or topical carbonic anhydrase inhibitors have been suggested to provide benefit for cystoid spaces in some patients, but have no apparent effect in many. Eventually visual field constriction progresses to the point that only small central islands of visual field remain (tunnel vision) at which point visual acuity is also reduced. When the central islands are eliminated, there is total blindness.

#### 1.2. Mechanisms of rod photoreceptor degeneration in RP

Mutations that cause RP differentially cause damage and death to rod photoreceptors. This is understandable when the mutation is in a gene that is specifically expressed in rod photoreceptors, such as *rhodopsin (RHO)*. There are a variety of mechanisms by which *RHO* mutations can cause cell death,[5] but a particularly common one is protein misfolding which activates the unfolded protein response which when persistently activated leads to cell death.[6] Since one mutant allele is sufficient to cause cell death, this type of *RHO* mutation results in dominantly inherited RP. Mutations in the *rod-specific β subunit of cGMP phosphodiesterase* gene (*PDE6b*) cause autosomal recessive RP in the *rd1* [7] and *rd10* [8] mouse models of RP and in humans.[9] These mutations cause increased levels of cGMP in rods which has been implicated in rod cell death.[10,11] It is more difficult to understand why mutations in some ubiquitously expressed genes cause

RP. Mutations in genes coding for pre-mRNA processing factors (PRPFs), which are involved in pre-mRNA splicing, can cause dominant RP.[12] This occurs due to haploinsufficiency because rod photoreceptors have a specific splicing program needed for ciliogenesis and hence are less tolerant of reduced expression of PRPFs than other cell types.[13] Thus, the highly specialized structure and abnormally high metabolic demands of rod photoreceptors may make them more susceptible than other cells to damage and death from reduced function of a variety of gene products underlying the surprising number and types of RP disease genes. [14]

##### 1.3. The role of oxidative stress in cone cell death in RP

Rod and cone photoreceptors are the only cell types in the outer retina and rods greatly out number cones with a ratio of 95:5. After rods die from a mutation, oxygen consumption in the outer retina is markedly reduced, but oxygen supply from the choroid is unchanged so that the remaining cones are exposed to very high levels of tissue oxygen.[15] The high tissue oxygen results in generation of superoxide radicals[16] initiating a cascade of production of reactive oxygen species (ROS) and reactive nitrogen species (RNS) that cause progressive damage to cones.[17,18] In several RP models, reduction of oxidative stress with antioxidants decreases cone dysfunction and death.[19,20] N-acetylcysteine (NAC) is particularly effective in reducing oxidative stress in cones in a model of RP and promoting cone survival and maintenance of function.[21] The hostile environment of the outer retina in RP induces metabolic abnormalities in cones that may also contribute to cone cell death,[22,23] but is clear that oxidative stress is a key therapeutic target.[24]

##### 1.4. Background on N-acetylcysteine

NAC is a derivative of L-cysteine that neutralizes ROS and RNS itself, but it is also converted to cysteine which is used to biosynthesize glutathione, a major component of the endogenous antioxidant defense system.[25] When given in a timely manner, intravenous NAC is a life-saving treatment for acetaminophen overdose because it prevents severe oxidative damage from aldehydes formed in the liver during metabolic breakdown of acetaminophen. [26,27] NAC is also approved as a mucolytic because it breaks the disulfide bonds of cross-linked mucus glycoproteins reducing mucus viscosity.[25] It has been hypothesized that NACs antioxidant activity could provide benefit in respiratory diseases leading to clinical trials in patients with chronic obstructive pulmonary disease (COPD) and idiopathic pulmonary fibrosis. Patients with COPD have slow deterioration of lung function punctuated by periods of exacerbations when there is symptomatic deterioration. In the randomized, placebo-controlled BRONCUS trial, 600 mg of NAC given orally once a day did not cause a significant reduction in the rate of decline of forced expiratory volume in 1 second, a measure of lung function, or the number of exacerbations per year.[28] In two subsequent studies a higher dose of NAC, 600 mg twice a day (bid). Compared with placebo, this higher dose of NAC given for 1 year in patients with COPD significantly improved lung function and reduced the frequency of exacerbations in the HIACE trial [29] and significantly reduced exacerbations in the PANTHEON trial [30]. In 182 patients with idiopathic pulmonary fibrosis, the IFIGENIA trial showed that oral NAC 600 mg three times a day (tid) plus standard therapy of prednisone and azathioprine for 1 year significantly improved lung function compared with standard therapy plus placebo,[31] but this was not replicated in a subsequent study.[32,33] While there is not a clear consensus as to whether oral NAC provides definite benefits in COPD and idiopathic pulmonary fibrosis, the many studies in these disease processes have helped to demonstrate that the safety profile of oral NAC is excellent.

##### 1.5. Fight RP Study – A phase I study testing the effect of NAC in patients with RP

The compelling preclinical studies demonstrating that oxidative damage contributes to cone cell death and that antioxidants including NAC promote cone function and survival, along with the excellent safety profile of oral NAC in other diseases, provided rationale for the phase I FIGHT RP study testing the effect of oral NAC in patients with RP (PI: Peter Campochiaro).[34] There were 3 cohorts of 10 patients that received treatment for 6 months and were observed for 3 months after stopping treatment. Patients in cohort 1 received 600 mg NAC bid for 3 months and then tid for 3 months. Patients in cohort 2 received 1200 mg NAC bid for 3 months and then tid for 3 months. Patients in cohort 3 received 1800 mg NAC bid for 3 months and then tid for 3 months. The mean age at baseline was 48±3 years, the mean age at disease onset judged by patient's recollection of the first onset of symptoms was 22, 25, and 30 years, and duration of disease was 28, 23, and 17 years in cohorts 1, 2, and 3, respectively. The mode of inheritance was autosomal dominant in 17%, autosomal recessive in 60%, and simplex in 23%. Genetic testing was not required for study eligibility, but results of genetic testing were available for 53% of patients and a causative mutation was identified in 43%.

There were 44 treatment emergent AEs of which 11 were judged to be drug-related, and 9 of these involved gastrointestinal symptoms. Nausea or stomach upset occurred in 8 participants, diarrhea in 6, heartburn in 1, constipation in 1, and an episode of vomiting in 1. Symptoms occurred during tid dosing in 7 participants and during bid dosing in 4 patients. Symptom severity was moderate in 2 participants in cohort 2 and one subject in cohort 3 soon after dose escalation to tid dosing; dosing was subsequently reduced to bid and the symptoms resolved. The remainder of the events were mild, brief, and resolved spontaneously without any change in dosing.

Good intraocular levels of NAC were obtained with oral dosing. The mean aqueous level was in the range of 100 ng/ml in cohort 1 and 300 ng/ml in cohorts 2 and 3. There was no statistically significant difference in the mean aqueous NAC level during the bid dosing period (week 4, 8, and 12 visits) compared with that during the tid dosing period (week 16, 20, and 24 visits). A pre-dose plasma NAC measurement was obtained at the week 12 and 24 visits, providing a measurement in the midst of treatment and approximately 12 hours after the most recent dose. The mean pre-dose measurement at week 12 and 24 was 302.70 and 392.75 ng/ml in cohort 1, 424.88 and 991.88 ng/ml in cohort 2, and 1025.33 and 1077.78 ng/ml in cohort 3. This indicates that substantial plasma levels can be maintained with bid dosing of 1800 mg NAC.

Mean baseline best-corrected visual acuity (BCVA) was 72 letters or 20/40 in cohort 1 and 74 and 75 letters in cohorts 2 and 3 which is about 20/32. With this good baseline vision, it was surprising to see a gradual steady increase in BCVA during the treatment period in each cohort that was statistically significant by linear mixed effects models. Macular sensitivity was measured with the MAIA microperimeter, which measures sensitivity at 68 loci and calculates the mean macular sensitivity from those 68 measurements. There was a steady increase in mean macular sensitivity during the treatment period in each cohort which was statistically significant in cohort 3. The improvement in mean macular sensitivity between baseline and week 24 was statistically significant in cohorts 2 and 3. There was no significant change from baseline EZ width in any of the cohorts during treatment, which was expected because 6 months is too short a period to detect a change in EZ width in the absence of treatment.

##### 1.6. NAC Attack Study Rationale and Benefit-Risk Assessment

While some of the studies demonstrating that oxidative damage contributes to loss of function and death of cones have been discussed above, there are many more [16,17,19-21,35] including confirmation in an independent laboratory.[36] Antioxidants, including NAC, reduce oxidative damage and promote cone survival and function.[21] Most importantly, as summarized in section 1.5, a phase I study showed that oral NAC is safe in RP patients with a maximum tolerated dose of 1800 mg bid that resulted in good intraocular levels and caused some improvement in cone function during a 6 month treatment period.[34] Assessment of sensitivity changes at individual loci showed that the percentage of loci that declined ≥ 6 dB was significantly less in patients receiving 1800 mg bid/tid of NAC compared with those receiving lower doses which also supports the use of 1800 mg bid.[37] The improvements in parameters of cone function during a 6-month treatment period suggest the possibility that long-term treatment with NAC in patients with RP may promote cone survival, reduce visual field loss, and reduce severe visual disability. Long-term treatment with 600 mg bid or tid NAC has been shown to be safe and well-tolerated in clinical trials in other disease indications. The phase I trial showed good safety and tolerability for 1800 mg bid/tid NAC for 6 months and an extension trial has shown good safety with 1800 mg NAC bid for more than 2 years. Based upon these data the benefit-risk profile is favorable for the proposed phase III trial to test the hypothesis that long-term NAC can promote cone survival and function and prevent severe visual disability in patients with RP.

##### 1.7. Public Health and Scientific Significance

RP is the most prevalent diagnosis among inherited retinal degenerations (IRDs). Currently there is no treatment for RP. Patients gradually lose their visual functions until becoming blind. Recent significant advances in gene therapies would only apply to very specific genotypes in IRDs. A pharmacological approach that promotes cone survival and that does not depend on the knowledge of specific genetic variants, if proven efficacious, will lead to a general therapy benefitting patients with a clinical diagnosis of RP. Thus, NAC Attack has the potential to impact the clinical management of RP. It will also provide clinical evidence (or lack of evidence) elucidating the role of oxidative stress in cone cell death in RP, leading to a better understanding of the molecular mechanism of photoreceptor degeneration in human RP.

### 2. Objectives and Endpoints

This study is a multicenter, randomized, double-masked, parallel and placebo-controlled trial in patients with RP to evaluated the efficacy and safety of 1800 mg bid NAC.

#### 2.1. Efficacy Objectives

##### 2.1.1. Primary Efficacy Objective

The primary efficacy objective is to determine if the progressive loss in EZ width assessed as the cumulative loss of EZ (calculated as the area above the curve, AAC) between baseline and M45 is significantly less in eyes of participants taking NAC 1800 mg bid compared with eyes of participants taking placebo. EZ width will be measured on a spectral domain-optical coherence tomography (SD-OCT) scan through the fovea.

##### 2.1.2. Secondary Efficacy Objective

The secondary efficacy objective is to assess the relative efficacy in eyes of participants taking NAC 1800 mg bid compared with eyes of participants taking placebo on the basis of the following endpoints:

- Change from baseline mean macular sensitivity measured by microperimetry (MP) at M45.
- Change from baseline in BCVA measured by Early Treatment Diabetic Retinopathy Study (ETDRS) protocol at M45.

##### 2.1.3. Exploratory Efficacy Objectives

The exploratory efficacy objective is to assess the relative efficacy in eyes of participants taking NAC 1800 mg bid compared with eyes of participants taking placebo on the basis of the following endpoints.

- Cumulative loss of EZ area assessed as the AAC over the 45-months study follow-up.
- Change from baseline mean macular sensitivity measured by MP at M4.5, M9, M18, M27, M36 and M40.5.
- Change from baseline BCVA at M4.5, M9, M18, M27, M36 and M40.5.
- Change from baseline in cone spacing, regularity, and reflectivity measured by adaptive optics-scanning laser ophthalmoscopy (AOSLO) at M9, M27, and M45 at selected sites.
- Proportion of eyes with ≥ 5 loci improved from baseline by ≥ 6 decibels (dB) at M4.5, M9, M18, M27, M36, M40.5 and M45 by MP.
- Proportion of eyes with ≥ 5 loci decreased from baseline by ≥ 6 dB at M4.5, M9, M18, M27, M36, M40.5 and M45 by MP.
- Exploratory subgroup analysis to assess the treatment efficacy compared to placebo in subgroups defined by sex, race, inheritance mode, genotype, age of symptom onset, smoking status, and by history of oral supplement consumption.
- Change from baseline to M27 and M45 in patient reported outcome assessed using NEI-VFQ 25.

- Change in fundus autofluorescence (FAF) between baseline and M45.
- Change in full-field sensitivity threshold (FST) between baseline and M45 at selected sites.

#### 2.2. Safety Objective

The safety objective of this study is to evaluate the safety and tolerability of oral NAC 1800 mg bid in patients with RP on the basis of the following endpoints:

- Incidence and severity of ocular adverse events (AEs)
- Incidence and severity of non-ocular AEs

#### 2.3. Pharmacokinetic Objectives

- Measure plasma NAC levels at each visit in all participants to evaluate compliance
- Correlate plasma NAC levels with efficacy endpoints

#### 2.4. Pharmacogenomic Objectives

- Examine the relationship between disease causing mutations and progressive decrease in EZ width over time in placebo participants
- Examine the effect of disease variants and modifier variants on treatment effect. Participants will be divided into high-responders and low-responders and their exomes will be compared (a) for patterns in disease genes and (b) in an unbiased fashion regardless of their disease gene to determine if certain disease genes or unbiased-selected genes have higher variant burden in high responders versus low-responders.

### 3. Study Design

#### 3.1 Description of the Study

The NAC Attack Study is a multicenter, randomized, double-masked, placebo-controlled trial in patients with RP to evaluate the efficacy and safety of 1800 mg bid NAC.

##### 3.1.1. Overview of Study Design

A total of 438 participants with RP will be enrolled and randomized at approximately 30 Clinical Sites in the Americas and Europe. Patients will be eligible if both eyes have an RP phenotype consisting of severe loss of rod function followed by progressive constriction of visual field and maintenance of visual acuity. EZ width of an eligible eye is between 1500 and 8000 µm as measured on the horizontal SD-OCT scan through the fovea.

During the screening period, the inclusion and exclusion criteria will be assessed and eligibility to participate in the study will be determined (see Section 4.1 for inclusion and exclusion criteria). Patients who are deemed eligible to participate in the study will be randomized in a 2:1 ratio to one of two arms stratified by Study Clinical Site:

- Intervention of NAC 1800 mg bid
- Placebo

Each participant will be followed for 45 months after randomization. Study visits including in-clinic visits, tele-visits with site Investigator, and phone calls with site Coordinator.

##### 3.1.2. Screening

Potential participants will be identified by reviewing medical records at each clinical site and identifying patients with RP who fit eligibility criteria. The treating physician of the patient will inform the patient of the study and discuss whether he/she would like to be screened for the study. Patients with RP who present to the study sites during the enrollment phase of the study will be informed about the study and offered the opportunity to be screened for the study.

Informed consent must be administered and signed by patients before any study-specific screening procedures are performed. Informed consent can be obtained by an investigator or a study coordinator by reading the consent to the patient and answering all questions. If the patient prefers, he or she can read the consent and then ask questions. If a study coordinator obtains consent and signs the consent form along with the patient, an investigator must then ask the patient if there are any remaining questions and answer them, and this is documented by signing an additional page of the consent by the investigator and the patient. The patient should be given as much time as needed to deliberate, confer with family members, and ask as many questions as necessary before signing the consent.

As part of the screening process:

- SD-OCT will be obtained and submitted to the study OCT Reading Center for eligibility assessment. The SD-OCT images will also be submitted to study Coordinating Center.

- Results of best corrected visual acuity (BCVA), microperimetry, slit lamp and funduscopic examinations, fundus autofluorescence (FAF, ultra-wide field [UW] FAF if available), and fundus photography (FP, UW FP if available) should be obtained and submitted to the Coordinating Center for eligibility assessment by the Study Chair's office.
- Blood should be collected for chemistry, hematology, and liver function tests at the local lab, and the lab results should be submitted to the Coordinating Center as soon as the results become available. If for any reason blood cannot be collected during the screening visit day, blood may be collected within the next 7 days and blood test results be submitted to the Coordinating Center as soon as possible. If blood is not collected at the screening day or within the next 7 days, this patient candidate becomes a screen failure.
- In addition, patient candidate's medical history including genetic test report, FAF, visual fields, MP, and/or Electroretinogram (ERG), if available will be submitted to study Coordinating Center for eligibility assessment by the Study Chair's office.

Patients who do not meet eligibility criteria for a reason that is remedial may be rescreened after an appropriate period of time. At rescreening, a new Study ID number will be assigned to the patient. A patient may be screened up to two times.

##### 3.1.3. Randomization

The randomization schedule will be prepared by the Coordinating Center and incorporated into the study data management system based on the REDCap (Research Electronic Data Capture) platform. Candidate patients deemed eligible by the Coordinating Center from the screening process will return for a baseline visit and after confirmation of eligibility by the enrolling site Investigator, they will be randomized through the Randomization function in REDCap. REDCap will provide a letter code encoding the treatment group which is assigned to the candidate. Once randomized, the candidate patient is called a *participant* of NAC Attack. The cover of each study medication package also has a letter code. The site Coordinator will dispense medication packages that have the same letter code as the letter code to which the participant is assigned through REDCap. The treatment group letter encodes information about the content of the study medication (intervention drug or placebo) but the patient and site personnel are masked with regard to the content.

##### 3.1.4. Treatment and Study Evaluations

After randomization, participants will be given about 10-months supply of study drug (intervention or placebo), with instructions to take 3 effervescent tablets in water twice a day. They will return to the clinic at M4.5 for evaluation and then at M9, M18, M27, M36, M40.5 and M45. At each in-clinic visit, drug reconciliation will occur. At each visit at Baseline, M9, M18, M27, M36, that is, every 9 months, participants will be given another 10-month supply of study drug.

At M13.5, M22.5 and M31.5, i.e. roughly half-way between 9-month in-clinic visits, there will be tele-visits between the participant and the site principal investigator (PI) or a co-investigator (Co-I) to assess for AEs and monitor study drug compliance. Roughly half-way between in-clinic visits and tele-visits, there will be phone calls between a site study coordinator and the participant to assess for AEs, monitor study drug compliance, and maintain rapport. In addition, a short message service (SMS) text-based system set by the Coordinating Center will be used to provide a reminder to take medication daily and through the SMS, once a week, participants will be asked how many doses were missed over the past week.

###### 3.1.5. Safety Monitoring

Section 5 details the procedures related to safety and adverse events (AEs) monitoring in NAC Attack. Medical monitoring in the Study is the responsibility of each Site Investigator and Coordinator, Study Chairman, the Medical Monitor, and the DSMC. A Medical Safety Monitor, who holds an MD, will review reports of SAEs as they occur. Every 3 months the Coordinating Center will provide the Medical Monitor with the number and type of SAEs at each clinical site and will provide statistical and analytical analysis to ascertain the presence of site-specific patterns of safety issues. Every 3 months, the Coordinating Center will also provide the log of AEs that the Study Chair deems to require review by the Medical Monitor.

###### 3.1.6. Data and Safety Monitoring Committee

An independent Data and Safety Monitoring Committee (DSMC) will provide ongoing safety monitoring. Members of the DSMC will follow a charter that outlines roles and responsibilities and will meet approximately every 6 months to evaluate safety data prepared by the Coordinating Center. The Medical Monitor may also, at his/her discretion, instruct the Coordinating Center to notify the full DSMC immediately of an SAE, and may request a meeting or teleconference with the committee prior to its next scheduled meeting. The DSMC will provide recommendations to the Executive Committee as described in the DSMC charter. Further details regarding the roles and responsibilities of the DSMC are provided in the NAC Attack DSMC charter.

##### 3.2. End of Study and Length of Study

The end of study is defined as one year after the date when the last study participant completes the final study visit. This end of study is defined to allow completion of scheduled data analyses, scientific publication of the main study findings and appropriate archiving of study materials.

The length of the study follow-up is 45 months for each participant.

##### 3.3. Rationale for Study Design

As explained in sections 1.1-1.7, there is strong rationale for testing the effect of oral NAC on cone survival and function in patients with RP. There are several features of RP that have a major impact on study design as explained in detail below.

###### 3.3.1. Rationale for Study Patient Population

###### 3.3.1.1. Clinical Diagnosis of RP

Despite the genetic basis of RP, it is a clinical diagnosis. Variants from almost 100 different genes have been linked to RP phenotypes. The variants interact with genetic background to cause a strong differential effect on rod versus cone photoreceptors. In other words, a key feature of RP is that the variant in the genetic background of a particular individual causes rod degeneration with little or no effect on cones. Cones then gradually degenerate independent of the variant, with oxidative damage playing a major role in cone degeneration. The same variant can cause RP in one individual and cone-rod dystrophy or pattern dystrophy in another individual. Thus, while the causative genetic variant is important information that must be gathered if at all possible, it is not essential for assigning a diagnosis of RP, which is based upon clinical characteristics of the disease, including marked loss of rod function with preservation of cone function centrally and relatively slow constriction of visual fields from gradual death of cones in a stereotypical pattern starting in the mid-periphery and extending posteriorly and anteriorly.

###### 3.3.1.2. Disease Stage Eligibility Criteria

When cone cell death has progressed into the central 30 degrees, it can be tracked by SD-OCT, because the EZ in SD-OCT scans corresponds to remaining cones with intact inner and outer segments. Thus, another important feature of the RP phenotype is a distinct, intact EZ centrally that is lost at a distance from the fovea and is relatively symmetrical in the horizontal axis. Documented reduction in EZ width over time strengthens the diagnosis of RP and is a measure of the rate of cone cell death. Therefore, the study population will be patients with an RP phenotype who have an EZ that has well-defined truncation at both the nasal and temporal sides as visualized by SD-OCT using a 25˚X 30˚ window, which has an upper limit of about 8000 µm. This study population of patients with RP phenotype is one in which oxidative damage is a major contributor to cone cell death and is appropriate for testing the effect of NAC on cone survival.

The study population also requires exclusion of late stage patients who have limited preserved EZ on the SD-OCT scan. Considerations include: first, cone density is highest in the fovea and more centrally constricted EZ is known to be associated with slower EZ loss. Thus, in order to observe a change in EZ-width during study follow-up, baseline EZ-width should not be too small. Second, the FIGHT RP study observed that those late stage patients who had a limited measurable EZ-width continued to have functional decline during the NAC treatment period, suggesting the impaired cones in late stage patients may lack the capacity to respond to antioxidative treatment. Third, a mathematical modeling study computationally predicted that under mutation-induced rod degeneration, treatments with antioxidants or trophic factors can significantly delay degeneration of photoreceptors and the earlier the treatment, the greater delay in the onset of blindness in patients with RP.[38] This suggests that there may be a larger NAC treatment effect for early stage versus late stage RP patients. Based upon these considerations, it has been decided that only RP patients with EZ width ≥1500 µm will be eligible for the study.

###### 3.3.1.3. Age Eligibility Criteria

Patients with RP aged 18-65 years will be eligible for NAC Attack. NAC is a regulatory approved drug for use as an antidote for overdose of acetaminophen and prevention of hepatic injury and for use as an adjuvant therapy in managing certain pulmonary conditions (e.g. cystic fibrosis, pneumonia). NAC's safety profile has been shown in studies for these conditions. NAC Attack will exclude RP patients younger than 18 years because there are no NAC safety data for this age group of the population. Many studies in patients ≥ 18 have demonstrated that NAC has an excellent safety profile, but this cannot be generalized to patients < 18 because the effects of NAC during development and periods of rapid growth are unknown. In addition, RP is a slowly progressive disease and there is not a compelling reason to include children who are usually in the rod degeneration phase of the disease with no cone degeneration, or in the early stages of cone degeneration. Also, most pediatric patients would not qualify based upon the criterion of an EZ width ≤ 8000 µm. If NAC is found to be beneficial, it would be reasonable to institute NAC treatment for a pediatric patient after reaching the age of 18 years because NAC is likely to be useful during the cone degeneration phase of the disease when oxidative stress is increased in the outer retina.

RP patients > 65 years at the time of signing the consent are excluded because it is less likely for such patients to have an EZ width ≥ 1500 µm and good central vision. If a patient aged 65 years older has good central vision and EZ width ≥ 1500 µm, it suggests that the patient must have mild disease (may have a genetic background that is protective) and late onset and slow progression. A patient with such slow progression would not be able to show a benefit from an intervention and thus is not appropriate for the trial.

##### 3.3.2. Rationale for Genotyping all Study Participants

Although identification of a causative genetic variant is not essential to identify patients with RP and is not needed for eligibility, it is important information. The rate of rod degeneration is substantially faster in *rd1* mice versus *rd10* mice and the subsequent cone degeneration is also faster in *rd1* mice. The efficacy of antioxidants including NAC with regard to maintaining cone survival and function is greater in *rd10* versus *rd1* mice. We hypothesize that more rapid rod degeneration results in faster onset of oxidative stress in *rd1* mice and this is a greater insult to cones and results in more rapid degeneration that is more difficult to prevent with antioxidants. There is evidence that the rate of rod degeneration influences the rate of cone degeneration in patients with RP as well because patients with X-linked RP who have rapid rod degeneration have more rapid loss of EZ width compared with patients with autosomal dominant RP who have slower rod degeneration.[39] Also, while mutations that result in an RP phenotype have a much greater effect on rods than cones, the difference is not absolute and can vary depending upon the mutation. If one mutation has a greater effect on cone survival than another mutation, it would be expected that NAC would show less benefit with regard to maintaining cone survival and function in patients with the first versus the second mutation. Therefore, it will be important to study the interaction between our efficacy outcome variables and causative genetic variants. We will be obtaining whole genome (exome) sequencing which will allow exploratory pharmacogenomic analyses which will be very valuable.

##### 3.3.3. Rationale for Primary Outcome Measure

The FDA has accepted reduction in the rate of loss of EZ width and EZ area as appropriate outcome measures to assess the efficacy of treatments for RP, because they are measures of cone survival, and over the long term should correlate with loss of visual function. The primary objective of our study is to determine if NAC promotes cone survival in patients with RP and therefore these outcome measures are ideal for our study and we will measure both. EZ width has been selected as the primary outcome measure because it provides greater statistical power and there is greater reproducibility in EZ width measurements compared with EZ area measurements. Independent studies have also reported a correlation coefficient of 0.96 between EZ width and EZ area in patients with RP,[40,41] indicating that they essentially provide the same information. Additionally, EZ-width measurement has stronger structure-function correlations than EZ-area.[42]

The FDA expressed a preference for at least five EZ measurements over time with minimum time between measurements of 9 months and the estimand efficacy evaluation should reflect the separation of curves between the intervention and the placebo groups. Our endpoint of AAC derived by using EZ measurements at baseline, M9, M18, M27, M36, and M45 quantifies the cumulative loss of EZ over 45 months and complies with that request.

##### 3.3.4. Rationale for Secondary Outcome Measures

Outcome measures that provide a clinically meaningful assessment of visual function are important. Constriction of visual fields leads to progressive disability in patients with RP and is therefore a valuable functional outcome measure. Our study population consists of patients with moderately advanced RP who have cone degeneration extending into the central 30˚ and most of these patients have visual field defects within the central 20˚ and show progression over time. We will use MAIA MP to assess central visual fields, because this instrument has the advantage of measuring macular sensitivity over a wide dynamic range at 68 central loci displayed on a fundus image.

In the FIGHT RP study, patients in the third cohort showed significant improvement in mean macular sensitivity during treatment with 1800 mg bid NAC for 3 months followed by 1800 mg tid for 3 months.[34] In the proposed trial, we hypothesize that NAC-treated patients will show an initial small improvement in mean macular sensitivity that may remain stable or may slowly decrease over time. But if there is a decrease, the rate of decrease will be slower than that in placebo-treated patients. This will result in a statistically significant difference in the change in mean macular sensitivity between baseline and M45 in the intervention versus the control group. Therefore, macular sensitivity is an important functional outcome measure.

While assessment of visual field changes with MP is the most appropriate functional outcome measure for our patient population, it has substantially more test-retest variability than measurement of EZ width. This is because measurement of retinal sensitivity requires the patient to push a button when they detect a light stimulus and there are things other than visual function that can influence the responses such as alertness, fatigue, manual dexterity, performance anxiety, and decisiveness. Additionally, posterior subcapsular cataracts are common in patients with RP and tend to progress over time; they can cause reduction in mean macular sensitivity unrelated to cone survival. Similarly, macular cystoid spaces are common in RP and often fluctuate spontaneously which can cause changes in mean macular sensitivity unrelated to cone survival. Furthermore, 10% of patients in FIGHT RP had a high rate of false-positive responses in both eyes during multiple MP tests and simply were unable to provide reliable data. This inability to decide when a light is seen is not related to visual function, but rather seems related to higher order processing and decision-making. Thus, for these reasons, EZ width is a better choice for primary outcome measure, but measurement of macular sensitivity with MP will provide a valuable secondary outcome measure that will complement EZ width measurement.

Visual acuity is spared until late in RP and therefore as was the case in FIGHT RP,[34] it is anticipated that most patients in the current study will have excellent BCVA at baseline. It was surprising to see that despite excellent baseline BCVA, there was a steady, small improvement during the 6-month NAC treatment period in each of the three cohorts that was statistically significant.[34] This indicates that it is useful to measure BCVA, which is also important for safety evaluation of any intervention, but due to a substantial ceiling effect, and the markedly delayed loss of BCVA that occurs in RP, it is appropriate as a secondary, but not a primary outcome measure.

###### 3.3.5. Rationale for Study Duration

The rate of EZ loss in RP is slow and therefore a long follow-up period will be needed to observe a statistically significant and clinically meaningful NAC treatment effect. The FDA recommended measurement of cumulative change in five EZ measurements with minimum time between measurements of 9 months. This requires a total follow-up period of 45 months which determined the study duration.

###### 3.3.6. Rationale for NAC Dose

In the FIGHT RP study, the maximum tolerated dose of NAC was 1800 mg bid and the third cohort showed significant improvement in mean macular sensitivity during treatment with 1800 mg bid NAC for 3 months followed by 1800 mg tid for 3 months and a significant effect was not seen with lower doses.[34] Selection of a dose of 1800 mg bid was also supported by locus-level analysis of macular sensitivity changes which demonstrated that that the percentage of loci that decreased ≥ 6 dB was significantly less in the third cohort.[37]

###### 3.3.7. Rationale for 2:1 Randomization Ratio

The rationale for a 2:1 randomization ratio is to make participation in a placebo-controlled clinical trial more appealing to patients with RP, even though unequal allocation between 2 arms is less statistically efficient compared to a design with equal allocation. There is no available treatment for RP and patients with RP are anxious and eager to try anything that they think might have any chance of providing benefit. In publications and other communications with the scientific community or the public, it has been stressed that while the small short-term improvement in cone function observed in some patients with RP in the FIGHT RP study is promising, it does not answer whether long-term NAC will reduce visual disability and be safe in patients with RP. However, in discussion with RP patients who are seen at our institution and through RP patient advocacy groups, it is our impression that some patients have assumed that NAC provides benefit and some have begun taking supplement formulations of NAC that are available without a prescription. After the need for a placebo-controlled trial to definitively determine if long-term NAC provides benefit and is safe, most RP patients indicate that they would be willing to forsake NAC supplements and participate in a clinical trial, but enthusiasm for participation is increased when it is explained that they would have twice the probability of receiving active drug as receiving placebo.

###### 3.3.8. Rationale for Measurement of NAC Plasma Levels

Measurement of NAC levels in plasma at each visit will provide an objective measure of compliance, particularly for patients in the placebo group. It will also provide a check as to whether a patient was given study drug or placebo in accordance with study group assignment. The timing of the most recent administration of study drug will be captured in the medication log and recorded in the study visit source document. One of the major differences among patients is efficiency of NAC absorption from the GI tract into the circulation resulting in differences in plasma levels. There is moderate correlation between plasma and aqueous NAC levels which will be the basis for an exploratory pharmacodynamic analysis.

###### 3.3.9. Rationale for AOSLO Sub-study

Measurements of EZ width and EZ area provide assessments of remaining cones with intact inner and outer segments, but do not have the resolution to assess individual cone structure and density. AOSLO provides unique ability to monitor cones at single cell resolution over time. As cone degeneration occurs, there is reduction in cone density even in areas where EZ is intact and therefore assessment of cone spacing by AOSLO provides a more sensitive assessment of cone health than SD-OCT derived EZ measurements. This will provide an anatomical assessment of the efficacy of NAC and will help evaluate the feasibility of using AOSLO-derived outcome measures in future interventional studies in RP.

###### 3.3.10 Rationale for FST Sub-study

Full field stimulus testing (FST) provides a global assessment of retinal sensitivity to light. The assessment can be obtained over a wide range of visual function from normal to severely reduced function in patients with advanced retinal degeneration. Unlike perimetry or microperimetry, the light stimulus is not directed at a pre-specified retinal location, but rather measures responses from the most sensitive areas without restricting to any specific locations. Additionally, by using different chromatic stimuli, the relative contribution of rods and cones can be determined. The Espion full field stimulus test uses a ColorDome to present a range of light stimuli to the entire retina and obtain full-field thresholds.

In summary, FST provides a global assessment of retinal function that complements the assessments provided by microperimetry or BCVA. It also uniquely provides measurements of rod function. The light stimuli are not intense and well-within the accepted safety range so the test poses no risk. In patients with RP, there is strong concordance between eyes so only one eye will be selected for testing.

### 4. Materials and Methods

#### 4.1. Study Patient Population

Evaluation of patient candidates for participation in the study is based upon the following eligibility criteria. They must have a diagnosis of RP by the site investigators based upon loss of rod photoreceptor function, followed by progressive loss of cone function but with preserved central cone function. Prior identification of a causative mutation is not required for eligibility, but all genotyping information will be captured and all participants will undergo genomic testing using whole genome (exome) sequencing. Both eyes will be evaluated for eligibility and if both eyes are eligible, data from both will be included.

##### 4.1.1. Inclusion Criteria

###### 4.1.1.1. General Inclusion Criteria

- Ability and willingness to provide informed consent
- Age ≥ 18 and ≤65 years at time of signing Informed Consent Form
- Ability and willingness to comply with the study protocol and to participate in all study visits and assessments in the investigator's judgement
- For candidates of childbearing potential: willingness to use a method of contraception
- Agreement not to take supplements other than vitamin A

###### 4.1.1.2. Ocular Inclusion Criteria

- Both eyes must exhibit the RP phenotype with evidence of loss of night vision, gradual constriction of visual fields, and maintenance of visual acuity;

In addition, an eye must meet the following criteria to be included in the study:

- Gradable EZ on a horizontal SD-OCT scan through the fovea center with width ≤ 8000 µm and ≥1500 µm and with well-defined truncation at both the nasal and temporal sides;

- BCVA ≥ ETDRS letter score of 61 (20/60, [6/18] Snellen equivalent);
- Sufficiently clear ocular media and adequate pupillary dilation to allow good quality images sufficient for analysis and grading by central reading center.

##### 4.1.2. Exclusion Criteria

###### 4.1.2.1. General Exclusion Criteria

- Active cancer within the past 12 months, except for appropriately treated carcinoma in situ of the cervix, non-melanoma skin carcinoma, or prostate cancer with Gleason score ≤ 6 and stable prostate specific antigen for > 12 months
- Renal failure requiring renal transplant, hemodialysis, peritoneal dialysis, or anticipated to require hemodialysis or peritoneal dialysis during the study
- History of thrombocytopenia not due to a reversible cause or other blood dyscrasia
- Uncontrolled blood pressure (defined as systolic > 180 and/or diastolic > 100 mmHg while at rest) at screening. If a patient's initial measurement exceeds these values, a second reading may be taken 30 or more minutes later. If the patient's blood pressure must be controlled by antihypertensive medication, the patient may become eligible if medication is taken continuously for at least 30 days.
- History of other disease, physical examination finding, or clinical laboratory finding giving reasonable suspicion that oral NAC may be contraindicated or that follow up may be jeopardized
- Cerebrovascular accident or myocardial infarction within 6 months of screening
- Patients taking slow-release formulations of nitrates
- Participation in an investigational study that involves treatment with any drug or device within 4 months of screening
- Use of any interventional treatment to the retina such as transcorneal electrical stimulation within 4 months of screening
- Three relatives already enrolled in study
- Pregnant or breast feeding females. Women of childbearing potential who have not had tubal ligation must have a urine pregnancy test at screening.
- Known history of allergy to NAC or any non-medicinal ingredients in the study medication including citric acid, aspartame, sodium bicarbonate, and lemon flavor
- Having taken NAC in any form in the past 4 months
- Phenylketonuria
- Fructose intolerance
- Histamine intolerance
- Glucose-galactose malabsorption
- Sucrase-isomaltase insufficiency
- Any major abnormal findings on blood chemistry, hematology, and renal function lab tests that in the opinion of the Site Investigator and/or the Study Chair makes the candidate not suitable to participate in the trial
- HIV or hepatitis B infection

  Angina that requires administration of sustained delivery of nitrates Being on strict sodium restriction that is deemed by the patient’s general medical doctor to be incompatible with the added sodium intake from study medication.

###### 4.1.2.2. Ocular Exclusion Criteria

- Evidence of cone-rod dystrophy or pattern dystrophy including focal areas of atrophy or pigmentary changes in the central macula
- Cystoid spaces involving the fovea substantially reducing vision
- Glaucoma or other optic nerve disease causing visual field loss or reduced visual acuity
- Intra ocular pressure >27 mm Hg from two measurements. If a patient's initial measurement exceeds 27 mm Hg, a second reading must be taken.
- Any retinal disease other than RP causing reduction in visual field or visual acuity
- Any prior macular laser photocoagulation
- Intraocular surgery within 3 months prior to screening
- High myopia with spherical equivalent refractive error > 8 diopters. If an eye has had cataract surgery or refractive surgery, a pre-operative refractive error spherical equivalent > 8 diopters is an exclusion
- Any concurrent ocular condition that might affect interpretation of results
- History of uveitis in either eye

#### 4.2. Method of Treatment Assignment and Masking

Treatment assignment will be based on randomization with a ratio of 2:1 of intervention versus placebo. Randomization strategy uses permuted block randomization stratified by Clinical Site. A varying block size of 3 or 6 will be used. The Coordinating Center will prepare the treatment allocation schedule for each clinic. The schedules are incorporated into the Randomization form in the study REDCap database. Treatment group allocations are issued by pressing the “Randomization” button available on the Randomization form in REDCap. The button becomes available only after verification of eligibility of a candidate in REDCap. Pressing this button will return a letter code for a participant. This letter code should be used when dispensing study drug to the participant. The letter code encodes allocation group information which is only accessible to designated unmasked study personnel in the Coordinating Center. This process ensures masking of the participants and personnel at the study Clinical Sites.

#### 4.3. Study Medication

The study medication includes the intervention drug and placebo. The intervention drug is N-acetylcysteine (NAC) in the form of 600 mg effervescent tablets (product brand name: Fluimucil). The placebo is also in an effervescent tablet formulation and has identical appearance as the NAC effervescent tablet. Placebo’s contents are the same as the inactive contents in the NAC tablet.

Existing FDA approved use of NAC includes treating acetaminophen (paracetamol) overdose and as a mucolytic for cystic fibrosis and other pulmonary diseases.

##### 4.3.1. Study Medication Storage Condition

Store at 25°C with excursions permitted between 10°C to 30°C. In addition, based on the available relevant stability data, for temperature excursions out of this range, the product properly stored in its original packaging can be used if the highest reached temperature is not more than 40 °C and the sum of the excursion time is less than 45 days.

###### 4.3.2. Study Medication Manufacture and Marketing Authorizations (MA) in Europe

Zambon Switzerland Ltd, Via Industria 13, 6814 Cadempino, Switzerland will provide both NAC (Fluimucil 600 mg effervescent tablets) and matching placebo for the study.

Fluimucil 600 mg effervescent tablets is an authorized product in EU, used in secretolytic therapy for acute and chronic bronchopulmonary diseases that are associated with a disruption of mucus formation and transport. Zambon holds the Marketing Authorisations (MA) for N-Acetylcysteine 600 mg effervescent tablets. The first MA was granted in EU in 1972.

###### 4.3.3. Study Medication Packaging, Labeling and Distribution

The intervention (NAC) and placebo will be produced and packaged in boxes of blister cards (2 tablets/card) at Zambon S.p.A. manufacturing plant in Switzerland and shipped to the NAC Attack Drug Labeling and Distribution Center (DLDC) which is a clinical services vendor that has been contracted for study drug repackaging, labeling and distribution. The shelf-life of NAC from Zambon is 3 years. Zambon will provide no less than 3 lots of study drug over the study period. Should the interim efficacy analysis conclude efficacy, a lot will be manufactured by Zambon allowing participants randomized to the placebo group to receive NAC.

The DLDC follows Good Manufacturing Practices (GMPs) and have internal quality control (QC) procedures regarding inspection and retention sampling of incoming materials, checking on masking of the material, in process inspection, and inspection before dispatches to sites.

The DLDC will provide secondary packaging to repackage 21 blister cards (i.e. 42 tablets, one-week supply of study drug) into a tri-fold package. This tri-fold package becomes the primary packaging for the study drug. A multi-language booklet study medication label following regulatory requirements from all involved regulatory agencies will be affixed to each tri-fold package. The label cover page has a barcode unique to the tri-fold package and encodes the identifying number of this package. The label also is printed with a letter code that can identify content in the package. The linkage information between the letter code and study drug is securely kept and only accessible to DLDC and Coordinating Center personnel who are unmasked (e.g. biostatisticians). The tri-fold packages upon passing QC procedures will be shipped to each Study Site using temperature-controlled shipment.

The first lot of study drug from Zambon is expected to cover the study medication needed during the enrollment period. Since the number of enrollments at each site is unknown as a priori, the first lot will be provided to each Clinical Site via 2 (or more) shipments of a few months apart.

At the time of production of the second or third lot of study drug from Zambon, the number of enrolled and randomized participants at each site will be already known and will be the basis for determining the quantity to ship to each clinical site. Upon repackaging and labeling of each of the lots at the DLDC, all finished tri-fold packages will be shipped to Clinical Sites upon written request and provision of information about the quantity to each site by the Coordinating Center.

Receipt of each shipment of study drug from the DLDC at a site should be documented per the NAC Attack Pharmacy Manual. Storage of study drug at Clinical Sites should follow local Investigative Pharmacy's requirements and should be in a secured and temperature-controlled room.

##### 4.4. Study Treatment Dosage, Administration

There are 2 arms: the intervention arm receiving NAC 1800 mg bid and the placebo control arm. The study drug is in the form of effervescent tablet which should be dissolved in water and taken orally. Participants should be instructed to take 2 doses during a day,12 hours apart, or at least 8 hours apart. If it has been more than 16 hours since the last dose, this means a dose is missed and the participant should skip the missed dose and be instructed to report the number of missed doses in the past week when responding the weekly SMS message survey (Section 4.11).

##### 4.5. Study Treatment Compliance

Compliance will be promoted and assessed by: (1) discussing it with participants during every in-clinic visit, tele-visit, and pre-scheduled phone call with a Study Coordinator, (2) a weekly SMS text message survey asking how many study medication doses were missed in the past week, (3) measurement of plasma NAC levels at each in-clinic visit, and (4) study drug reconciliation by the Study Coordinators. To promote compliance, a text message will be sent each day to remind participants to take study medication.

##### 4.6. Study Medication Dispensing and Accountability

Study medication should be dispensed at Baseline, M9, M18. M27, and M36 visits, i.e. it is dispensed every 9 months.

At Baseline visit, participants will be given a 10-month supply (for the next 9 months period plus 1 month to account for scheduling window) + some overage emergency supply of study drug. They will be instructed to record the date and time of each dose and save all used study medication packages (i.e. the tri-fold packages).

At the M4.5 in-clinic visit, participants should bring all *used* study medication packages and Study Coordinators will perform drug reconciliation and record the results following the NAC Attack Pharmacy Manual. At this visit, participants do not need to bring unused study medication packages and there is no medication dispensing at this visit. The drug reconciliation at this visit allows study coordinators to identify potential study medication adherence issues related to individual participants relatively early in the study.

At each of the M9, M18, M27, M36 and M45 in-clinic visits, participants should bring *all used and unused* study medication packages and Study Coordinators will perform drug reconciliation and record the results following the NAC Attack Pharmacy Manual. Participants will also be given a 10-month supply (for the next 9 months period plus 1 month to account for scheduling window) + some overage emergency supply of study drug. The medication dispensed at each visit to a participant should have the same letter code as the code that the participant was assigned to at the Baseline visit (Section 4.2). Unused and unexpired study medication packages may be redispensed to the same participant.

All study medication dispensing and reconciliation activities should be recorded according to NAC Attack Pharmacy Manual and each study drug accountability activity must be conducted and signed by two study team members at the site.

#### 4.7. Concomitant Therapy

Concomitant therapy consists of any medication (e.g., prescription drugs, over-the-counter drugs, vaccines, herbal or homeopathic remedies, nutritional supplements) used by a participant in addition to protocol-mandated treatment from 30 days prior to screening through the study completion/discontinuation date. Study site investigator or coordinator should collect information about use of any such medications from a participant on the Concurrent Medications and Supplements Log during every encounter with the participant.

##### 4.7.1. Permitted Therapy

- Medications prescribed by the patient’s medical doctor
- Aspirin, other nonsteroidal anti-inflammatory agents, and acetaminophen
- Cough suppressants are permitted but must be administered 2 or more hours before or after study medication as the combination of cough suppressant and study medication can lead to accumulation of secretions and congestion requiring monitoring of secretions and breathing.
- Anti-histamines
- Laxatives
- Any topical ocular medication prescribed by an eye care professional including drops to reduce intraocular pressure, drops for dry eye, antibiotics, and anti-inflammatory drops as indicated
- Over-the-counter medications for intercurrent illness other than antioxidants
- If the patient develops clinically significant cataract that interferes with activities of daily life, the Clinical Site PI should discuss this with the Study Chair prior to considering cataract surgery.
- Antibiotics are permitted for treatment of acute infections but must be administered 2 or more hours before or after study medication.
- Patients who have a history of ulcer disease may take their normal ulcer treating medication and should notify their study investigator if they notice GI upset during the study participation.

- Patients who have a history of ulcer disease may take their normal ulcer treating medication and should notify their study investigator if they notice GI upset during the study participation.

###### 4.7.2. Prohibited Therapy

- Antioxidant supplements including herbal or homeopathic agents

##### 4.8. Study Procedures at Study In-Clinic Visits

###### 4.8.1. Study Assessment Schedule

The schedule of in-clinic visits and activities to be performed during the study are provided in **Table 1**.

###### 4.8.2. Informed Consent and Screening Visit

Written informed consent for participation in the study must be obtained before performing any study-related procedures (including screening evaluations). A physician PI or Co-I at the study Clinical Site may share responsibility for explaining the study with a Study Coordinator, but ultimately it is the physician's responsibility to make sure that the patient candidate understands the study and consents to participate.

If a study team member other than the physician reviews the consent form with the patient and answers initial questions, that team member and the patient should sign the consent, but then the physician must ask the patient if there are any remaining questions or concerns and after they are addressed, the patient and physician must sign an additional part of the consent form. It is important that the patient understands the concept of randomization and the importance of adhering to it. A photocopy of the signed Informed Consent Form will be provided to the patient candidate, and the original copy, regardless whether the patient candidate subsequently is enrolled and randomized, will be maintained at the Clinical Site in the patient's study binder which may be inspected during Coordinating Center's monitoring visits. The DSMC may review consent forms from time to time to assure adherence to minimum standards established by that committee.

All screening evaluations must be completed and all data including lab results and imaging files submitted to the Coordinating Center. The SD-OCT files will also be submitted to the OCT Reading Center. The Study Chair or Vice-Chair will review all data from the screening evaluation of a candidate. The Coordinating Center will combine the eligibility evaluation from the Study Chair's office and that of the OCT Reading Center regarding SD-OCT eligibility to make a final determination of eligibility. This final determination of eligibility will be entered into REDCap by the Coordinating Center within 21 days of screening. Upon such data entry into REDCap, the Clinical Site will receive an email from REDCap notifying the site whether the candidate is eligible or not. A designated Coordinating Center member will also email the result of eligibility determination to the Clinical Site team. The site Coordinator will reply to acknowledge the receipt of the notification within 1 business day. The Coordinator will notify the candidate of the eligibility status and if appropriate schedule the candidate for a baseline visit within 42 days of the screening visit.

The Clinical Site will maintain a screening log to record details of all patients screened, confirm their eligibility, or record reasons for screen failure.

##### 4.8.3. Informed Consent and Screening for the AOSLO Sub-Study

For Clinical Sites that participate in the AOSLO sub-study, in the NAC Attack consent form as described above, patient candidates are given the option to consent to participate in the AOSLO sub-study where if eligible they will receive AOSLO imaging at designated study visits (see **Table 1**). With patient consent, in addition to the NAC Attack screening activities, the enrolling site Investigator completes a screening evaluation form of eligibility for the AOSLO sub-study which will be data entered into REDCap.

After the overall NAC Attack eligibility is determined by the Coordinating Center for a candidate (see above 4.8.2), the Coordinating Center checks the AOSLO screening eligibility for the candidate. The Clinical Site will receive an email noting the AOSLO sub-study eligibility result for the candidate. A candidate can only be eligible for AOSLO sub-study if he/she is eligible for the overall NAC Attack study. If the candidate is eligible for the AOSLO sub-study, the site Coordinator should schedule an AOSLO baseline visit *that is prior to* the NAC Attack baseline visit. This is because AOSLO imaging can be time demanding and is difficult to fit into the NAC Attack Baseline visit schedule. AOSLO baseline imaging should be acquired before study medication initiation, and hence the AOSLO baseline visit should be scheduled before the NAC Attack baseline visit. Scheduling of the NAC Attack baseline visit should always be within the window allowed between screening and baseline (see **Figure 1**).

##### 4.8.4. Patient Identification

Each candidate screened will be assigned a permanent study identification (ID) number and alphabetic code to be used on all study forms, photographs and tests. The patient study ID number is a two-part identifier consisting of the relevant Clinical Site number and a number indicating the patient within the site. Patients will also have a four-letter randomly generated alphabetic code that is not linked to their name.

The identification code(s) assigned to a candidate at the first Screening Visit encounter is used to identify the candidate during additional encounters and is the permanent participant identifier used at the time of enrollment and randomization and for all follow-up visits and procedures. A candidate who fails initial screening such as because of uncontrolled blood pressure may be rescreened after antihypertensive treatment for 30 days to control blood pressure. Such candidate will receive new study identification codes.

At some point in follow-up, a participant may transfer care or study participation from one site to another. In such a case, a Site Transfer of Patient Form is completed and submitted to the Coordinating Center. The participant's identification code(s) are permanent and do not change even when a participant transfers to a different site.

For each participant, the study ID number, alphabetic code, and full name and emergency contacts will be recorded in a patient log and a backup patient log. These patient logs will be kept in a secure and locked place at the local clinical site and will only be accessible to the local site study personnel. These logs should not be entered into the study REDCap database.

###### 4.8.5. Medical History, Demographic Data, and Concomitant Medications

Patient history related to eligibility, including clinically significant diseases, surgeries, cancer history (including prior cancer therapies and procedures), reproductive status, past use of NAC will be recorded at screening. All questions on the Registration and Demographics Form and Patient History for Eligibility Form are answered prior to ophthalmoscopy and photography to ensure that the patient is eligible. If it is determined that a candidate is ineligible based upon the Patient History for Eligibility Form or a procedure performed early during the screening process, this candidate is designated a screen failure and should not undergo the ensuing procedures.

Patient medical history including surgical history, smoking history, and use of alcohol or other recreational substances will be recorded at baseline. In addition, all medications (e.g., prescription drugs, over-the-counter drugs, vaccines, herbal or homeopathic remedies, and nutritional supplements) used by the patient within 2 months prior to baseline will be recorded.

At the time of each follow-up study visit (either an in-clinic visit, a tele-visit, or a phone call with study coordinator), an interval medical history should be obtained and any changes in medications and allergies should be recorded. This information will be recorded in the Concurrent Medication and Supplement Form and entered into the study REDCap database. The investigator should encourage the participant not to take any medication or supplement unless it is prescribed by his/her primary care physician or other specialty physician. Supplement taking may confound the assessment of NAC's effect on RP. More importantly, the participant may be administered a high dose of NAC and the possibility for a dangerous drug interactions or over-dose must be evaluated by the investigator and thus it is critical that the investigator know all medications or supplements that a patient is taking. The Coordinating Center will provide the talking points that the investigator can utilize for this counseling process.

Participants will be instructed to bring all medications or supplements they are taking to in-clinic study visits. Participants should not be taking a supplement unless it is requested by one of their physicians. For any such supplement, the Study Coordinator should take a photo of the bottle or packaging that shows the brand name and the list of ingredients. This information about the supplement then will be recorded in the Concurrent Medication and Supplement Log and be data entered into REDCap. The photo should also be uploaded into the REDCap database as a supporting document for this study visit.

###### 4.8.6. Assessment of Study Drug Compliance and Study Drug Reconciliation

At each study visit (either an in-clinic visit, a tele-visit, or a phone call with study coordinator), the study team should ask patients whether they have missed doses of study drug and if so how many and record the information in the appropriate case reporting form (CRF). Medication compliance will also be assessed in weekly SMS text compliance surveys. If there is suboptimal compliance identified from the SMS survey response, the Clinical Site will be notified by the Coordinating Center who will request a call between the Clinical Site personnel and the participant to discuss compliance issues and provide a plan for improving compliance. Such Coordinating Center requested call between the Clinical Site and a participant should be recorded in an Unscheduled Visit Form. If such a call had already occurred with the participant since the last study visit, the Study Coordinator should ask if the compliance plan has helped and determine whether additional mitigation strategies should be instituted.

At M4.5, participants will only bring used study medication packages for drug reconciliation. At M9, M18, M27, M36, and M45, participants will bring all used and unused study medication packages. Study Coordinators must go through the material, count all used blister packs and remaining unused study medication packages and record this on the Study Medication Reconciliation Form. Unused study medication packages that are not expired may be redispensed according to the study pharmacy manual to the participant as part of the study drug supply for the next 9 months. Expired study drug and unused study drug from already opened study medication packages must be destroyed locally following the Clinical Site's local policy on investigational drug destruction. If a study site does not have the capacity to dispose of unusable study drug, the site should work with the Coordinating Center to have the drug sent to the Coordinating Center for destruction following Johns Hopkins guidelines on drug destruction.

The used medication packages contain participant's recording of the date and time when each dose was taken. The portion of the packaging of each used medication package or a copy of the packaging should be filed in the participant's binder.

###### 4.8.7. Review of AEs

Review of AEs will be done at each study visit during follow-up (either an in-clinic visit, tele-visit, or phone call with study coordinator). Care should be taken not to lead participants into thinking that an AE is expected. Instead, non-leading questions should be asked such as "how are you doing" or how are things going with the medication?"

###### 4.8.8. Vital Signs

At the screening visit, vital signs include measurements of pulse and systolic and diastolic blood pressure with the patient in a seated position after resting for 5 minutes. If there is uncontrolled blood pressure (defined as systolic > 180 and/or diastolic > 100 mmHg), a second reading may be taken 30 or more minutes later. Both measurements must be entered into the Screening Vital Signs Measurements Form. If the second measurement still exceeds the above values, the patient is not eligible and the screening visit is stopped. A patient who fails screening because of hypertension may be rescreened after he/she sees a primary care physician, and achieves good blood pressure control for at least 30 days. Vital signs will also be obtained at each in-clinic follow-up visit

###### 4.8.9. Ocular Assessments

Ocular assessments include the following and will be performed for both eyes except where noted and at specified timepoints according to the schedule of activities in **Table 1**. The first 3 assessments should be done first and follow the order. The remaining assessments can be done in any order. Dilation should always be done *after* BCVA and *before* MP tests.

1. Refraction and BCVA using the ETDRS protocol at a starting distance of 4 meters performed prior to dilation by a study-certified BCVA examiner.

2. Measurement of IOP with a method that remains consistent throughout the study.

3. At visits specified, pupils should be dilated and MP testing done by a study certified MP examiner on both eyes using a MAIA (Macular Integrity Assessment, Centervue, Padova, Italy) microperimeter, a noninvasive confocal microperimetry instrument that measures light sensitivity while using an eye tracker with Scanning Laser Ophthalmoscopy (SLO) for accurate, real-time, compensation for eye movements. During a study visit, MP should be done as early as possible because fatigue negatively impacts test performance. If at the baseline, a participant is unable to conduct the MAIA test reliably (the rate of false positive response to stimuli at the optic nerve>30% is considered unreliable based on the manufacture's Operation Manual 2013), this participant does not need to undergo the MAIA test in follow-up visits.

4. Full field stimulus testing (FST). At selected sites, DiagnosysFST will be done at Baseline visit and selected follow-up visits as specified in **Table 1**. . Only one eye will be tested: the test eye is the study eye if the participant has one eye eligible for NAC Attack, or the better eye in terms of mean macula sensitivity from microperimetry obtained at the Baseline visit if the participant has both eyes as study eyes. The chosen FST eye will be dark adapted for about 30 minutes and receives testing in a dark room. Thresholds with blue light, red light and white light stimuli will be obtained sequentially following the study FST manual. Certification for FST testing is not required.

5. Slit lamp examination. The Ocular Exam Form shows structures to be examined and lists check boxes for findings. If there is a finding not listed, it should be noted in an open text field. It is important to document and grade cataracts, because they can reduce visual function. Standard photographs established by the Age-Related Eye Disease Study (AREDS) cataract grading scheme will be provided by the Coordinating Center and should be pulled up on the screen to assist in grading during the examination.

6. Funduscopic examination. The Ocular Exam Form shows structures to be examined and lists check boxes for findings. If there is a finding not listed, it should be noted in an open text field. Findings of interest for which documentation is requested include

the following. (1) vitreous opacities, Weiss ring, vitreous hemorrhage and inflammation. (2) vertical cup to disc ratio optic nerve pallor, optic nerve head glaucomatous change and edema (3) peripheral retina pigmentary changes, vessel attenuation, retinal detachment, and (4) macular changes including cystoid macular edema, epiretinal membrane, and vitreomacular traction. The SD-OCT scans should be used to help document macular structure (item 4) and determine whether a macular pathology involves the fovea center.

###### 4.8.10. Ocular Imaging

The OCT Reading Center will provide Clinical Sites with the study central OCT Reading Center imaging manual and training materials for SD-OCT. Before any SD-OCT images are obtained, the OCT instrument, site personnel and test images will be certified and validated by the OCT Reading Center as specified in the OCT Reading Center manual. All ocular images will be obtained by certified Clinical Site personnel and submitted to the OCT Reading Center for independent analysis and storage.

Fundus photography and fundus autofluorescence (ultra wide field for both if possible) wil be done at the screening visit and some follow-up visits as specified in **Table 1**. Neither of these imaging tests require certification. Images from both eyes must be submitted to the Coordinating Center within 1 business day.

###### 4.8.11. Laboratory Samples

###### 4.8.11.1. Blood samples for local lab testing

Blood will be drawn for chemistry, hematology, and liver function tests at most in-clinic study visits as specified in Section 4.8.1 Study Assessment Schedule and testing will be done in a local laboratory.

Blood chemistry tests include sodium, potassium, chloride, calcium, carbon dioxide, glucose, creatinine, urea nitrogen/BUN, total bilirubin, direct bilirubin, AST, and ALT. Hematology tests include hemoglobin, hematocrit, platelet count, RBCs, WBCs, mean corpuscular volume, mean corpuscular hemoglobin, red cell distribution width, and differential including neutrophils, lymphocytes, basophils, eosinophils, and monocytes.

###### 4.8.11.2. Blood samples for plasma NAC testing

Blood will be drawn and plasma prepared for plasma NAC by study site personnel at most in-clinic study visits as specified in **Table 1**. The plasma will be stored frozen and will be shipped to Study central laboratory for plasma NAC analysis every few months following requests from the Coordinating Center.

###### 4.8.11.3. Blood sample for genetic testing

Blood will be drawn and placed in tubes that are provided for genetic testing at the baseline visit and immediately shipped to the study central laboratory for genetic testing following instructions from the study central laboratory.

###### 4.8.11.4. Urine sample for pregnancy testing

At screening, candidates of childbearing potential will have a urine sample collected and tested for pregnancy in the clinic. The test result will be checked by 2 study staff members. Candidates with a positive result will be ineligible for trial enrollment. At each follow-up in-clinic visit, participants with childbearing potential will be asked about their pregnancy status. If the status is unsure, a urine pregnancy test will be conducted. The urine test result should always be checked by 2 study staff members. If the pregnancy test is positive, the participant will be instructed to stop the study medication, but will continue all study visits and procedures. The participant may resume study medication after the pregnancy and breastfeeding. Urine test result should always be checked by 2 study staff members.

###### 4.8.12. Patient Reported Outcomes

The NEI-VFQ-25 questionnaire will be self-administered at the baseline, M27, and M45 study visits. The primary mode of administration will be self-administration using paper forms. At baseline, most participants will have good visual acuity and no worse than 20/60 in at least one eye and thus, self-administration is not expected to be a problem and will avoid influence by study team members or family members. A study team member may help clarify the meaning of questions but the participant's companion family members should not provide any input.

A large font will be used to print the questionnaire. If a participant indicates that self-administration is difficult or a study coordinator notes a problem, the study coordinator will ask the participant if he/she would agree to receiving a telephone call from a trained interviewer from NAC Attack Coordinating Center. If so, a telephone call between the participant and the Coordinating Center interviewer can be arranged to administer the questionnaire.

##### 4.9. Study Procedures at Study Phone Calls Visits with Coordinators

Schedules of the phone call visits with Coordinators are shown in **Table 1**. The purpose of the calls is to maintain personal contact with the participant, to remind the participant to take the study medication as scheduled, and to inform the study team of any new health problems or medications. The relevant brief follow-up CRF should be completed to document the phone call visit, medication compliance assessment and AE reported.

##### 4.10. Study Procedures at Study Tele-visits with Site Investigators

Schedules of the tele-visits with site Investigators are shown in **Table 1**. The purpose of the tele-visits is to maintain contact with the participant and keep the participant engaged in the study. The site investigator should remind the participant of the importance of taking the study assigned medication as scheduled and review any symptoms that occurred since the last visit. The Follow-up Tele-visit with Investigator Form should be completed to document the visit, medication compliance assessment and any AEs reported.

##### 4.11. Weekly SMS Messaging for Assessing Compliance

The study will use short messaging system (SMS) messaging based mini-surveys to monitor participant's treatment compliance. The Coordinating Center will program the

SMS system through the REDCap study data collection system. Unless opt-out, the participant will receive an automated SMS message at a certain time of a certain day during his/her participation of the study. The message asks the participant to report the number of doses that was missed during the past week. Participant's responses to the SMS messages will be captured by the study REDCap data system.

#### 4.12. Special Situations

##### 4.12.1. Unscheduled Visits

###### 4.12.1.1. Unscheduled visits due to a safety concern

An unscheduled visit may occur if there are ocular conditions of participants that need to be seen in the clinic. At such a visit, only procedures that are necessary for AE evaluation should be performed. For example, blood draw for plasma NAC and microperimetry do not need to be performed. Blood draw for safety monitoring, BCVA and OCT imaging may need to be performed to facilitate AE evaluation, but the data and files may not need to be submitted to the Coordinating Center or the OCT Reading Center. Only information regarding the AE should be reported following the AE reporting procedures described in Section 5.

###### 4.12.1.2. Unscheduled visits due to a missed study in-clinic visit

An unscheduled visit may also occur if a participant is unable to be seen within the allotted in-clinic visit window. After a missed visit the participant should be seen in the enrolling clinic as soon as possible and be evaluated for any AEs, medication compliance and study medication should be dispensed. All study procedures required at the missed in-clinic visit should be performed at such an unscheduled visit.

##### 4.12.2. Telephone Call Prompted by Lack of Treatment Adherence from the SMS Surveys

The Coordinating Center will generate a summary report of participants SMS survey responses every few weeks. If suboptimal treatment compliance is observed, the Coordinating Center will notify the Study Coordinator who then should initiate a phone call with participant to explore the issue in detail and determine a plan of action to promote compliance or resolve other issues. Such phone call should be documented in the Unscheduled Visit Form.

##### 4.12.3. Pregnancy Events

Candidates of childbearing potential can only be enrolled if they consent to adopt contraceptives during heterosexual intercourse during the study period and to give permission to the study to collect information related to the pregnancy outcome should pregnancy occur. If pregnancy occurs during study follow-up, the participant should stop the study medication. The Study Coordinator should report the event in the Pregnancy Reporting Form and notify the Coordinating Center and local IRB if required. The participant must return all unused study medication to the Site. The Site Coordinator will dispose of the unused study medication appropriately.

The participant should continue study visits and procedures and may resume study medication after delivery and discontinuation of breast feeding. The Coordinator can use the appropriate pregnancy reporting CRF to record such visits.

###### 4.12.4. Follow-up of participants Unable to Return for Scheduled In-Clinic Examinations

Failure to have an in-clinic study visit within the allotted window is a major protocol violation that jeopardizes the study and must be avoided if at all possible. If a scheduled in-clinic visit is missed, Study Coordinator should immediately contact the participant to determine the cause of the missed visit, and set up another visit within the prescribed window. If multiple scheduled visits are missed and it is not possible to have an in-clinic visit within the allowed window, an unscheduled visit for a safety assessment and dispensing of study drug should be scheduled (see Section 4.12.1).

If an in-clinic visit within the allotted window is missed for a participant because the coordinator cannot reach the participant before the window closes, a participant search process should be initiated. Continued effort should be made including no less than 5 phone calls, no less than 5 emails and 1 registered postal mail to the participant, as well as contacting the participant's emergency contacts, family members or friends. The search process and outcome (e.g. contact with the participant was never re-established) should be recorded in the Participant Search Form. If the Study Coordinator discovers that the participant has died, the Study Coordinator should report it using the Adverse Event Reporting Form and enter the form into the REDCap study database.

###### 4.12.5. Changing Site for Participant Follow-Up

During the study participation, a participant may move and it may be more convenient to have study visits at another Clinical Site. If the second Clinical Site team accepts responsibility for follow up, a Clinical Site transfer may be arranged and documented with the Site Transfer of Patient Form. The PI and study coordinator from both Clinical Sites should approve the transfer. The Coordinating Center enters the Participant transfer form in REDCap. A copy of the participant's study binder should be provided by the original Clinical Site to the new Clinical Site. The new Clinical Site will start seeing the participant for his/her next in-clinic study visit. Prior to any procedure at the first in-clinic visit at the new site, the participant must be reconsented at the new site and the signed consent form should be filed into the participant's binder at the new site.

###### 4.12.6. Emergency Unmasking

If a participant has an SAE that is felt to possibly be study medication-related and management of the patient would be facilitated by knowing if the participant was receiving active drug, a site investigator may request unmasking of a participant's treatment group. Such a request should be submitted to the Coordinating Center. The Coordinating Center will forward the request together with relevant participant materials to the Medical Monitor within 24 hours. The Medical Monitor will determine whether unmasking is appropriate and approve it or whether the request is denied within 24 hours. If the request is approved, an unmasked member at the Coordinating Center will provide the participant's treatment group to the Site Investigator, Study Chairman, and Medical

Monitor. This process should be recorded in the Unmasking Form. Communications during this process within the Coordinating Center and between the Coordinating Center and the clinical site, Study Chairman and Medical Monitor will ensure masking remain in place with all other participants.

###### 4.12.7. Participant Death

As soon as clinic personnel become aware that a participant has died, the death event needs to be reported using the Adverse Event Reporting Form. The form should be data entered into REDCap andand sent to the Coordinating Center. No further reminders of remaining visits and telephone contacts will be generated for the deceased participant. As participant death is a serious adverse event (SAE), all reporting should follow the corresponding procedures described in Section 5.

##### 4.13. Procedures Regarding Biological Samples

NAC Attack will require blood sample collection at most in-clinic visits. The schedule of blood draw can be found in **Table 1**. Blood samples will be used for 3 purposes:

- Genetic testing to identify pathogenic variants for RP and whole genome (exome) sequencing.
  - Test Frequency: only once using the blood sample collected at one study visit.
  - Specimen type: whole blood.
  - Lab: Study central laboratory contracted by the Coordinating Center.

- Testing for routine blood chemistry, hematology, and liver function to monitor safety.
  - Test Frequency: at most in-clinic visits per **Table 1**.
  - Specimen type: whole blood.
  - Lab: the Clinical Site's local diagnostic lab.

- Testing for plasma level of NAC.
  - Test Frequency: at most in-clinic visits per **Table 1**.
  - Specimen type: plasma.
  - Lab: Study central laboratory contracted by the Coordinating Center.

###### 4.13.1. Blood Sample for Genetic Testing

The Study central lab for genetic testing will provide an onboarding package for Clinical Sites, which will include a resource guide, requisition forms, specimen collection kits, shipping materials and instructions for returning the kit to the laboratory testing facility. After collection of a blood sample from a participant, the site Study Coordinator will label the specimen collection kit and ship the sample to the testing facility as instructed in the onboarding package. The sample collection and shipment process should be recorded in the appropriate CRF.

The central laboratory will extract DNA from the blood sample for sequencing analysis to identify pathogenic variant(s) related to retinal dystrophy. The DNA sample will be maintained for re-sequencing if the initial sequencing was unsuccessful, and will be destroyed at the sequencing facility at the end of the study. Participants will be provided with a clinical genetic testing report with information regarding whether or not he/she has any genetic variants that have been determined to cause RP.

###### 4.13.2. Blood Sample for Plasma NAC Measurements

For measurement of NAC level, blood samples need to be processed to obtain plasma. The plasma samples will be stored at each Clinical Site and be shipped to the Study central laboratory for measuring plasma NAC levels once every a few months following the shipping schedule provided by the Coordinating Center.

Plasma sample processing procedures are detailed in the study lab manual. In brief, blood is collected in 2ml vacutainer tubes containing EDTA. The vacutainer tube is centrifuged at 1500-1800xg for 10 min at room temperature. After centrifugation, the plasma is removed and divided in half, with each half placed in a barcoded polypropylene tube. The tubes are labeled with study ID, visit number and sample collection date and stored in a -18°C (or lower temperature) freezer.

Every a few months the Coordinating Center will request study sites to ship stored plasma samples to the Study central laboratory for plasma NAC testing. For each participant, one of the two duplicate samples will be shipped and the other sample will remain in storage at the site as a back-up. The back-up sample will be shipped to the Study central laboratory for plasma NAC testing only when necessary (e.g. the original sample sent to the laboratory was damaged during shipping). Shipment procedures follow instructions provided by the Coordinating Center. Upon receiving a shipment request from a site, the shipping courier arranged by the Coordinating Center will provide dry ice and external packaging to the site for the shipment. All plasma shipments need to be prepared by study team members who are IATA certified shippers of biological materials.

The Study central laboratory for plasma NAC will use their validated assay to measure plasma NAC level. Remaining plasma samples will be destroyed within 3-months after the bioanalytical report is provided by the central laboratory and the Coordinating Center confirms the quality of the data. Unused back-up samples at the sites will be destroyed annually.

###### 4.13.3. Blood Sample for Safety Monitoring

Assessment of blood chemistry, hematology, and liver function tests will be performed to monitor safety. These tests are routine blood tests and will be analyzed in local CLIA (Clinical Laboratory Improvement Amendments) certified laboratories. The institutions of the clinical sites often have the capacity to conduct these tests in their hospital laboratories or through their contracted labs. The blood samples at each clinical site should be sent to the local lab on the same day the samples are drawn. The Study Coordinator is responsible to collect the lab results.

##### 4.14. Quality Assurance Methods

###### 4.14.1. Quality Assurance Related to Drug Distribution, Storage and

##### Accountability

The study medication will be supplied to each site by the NAC Attack Drug Labeling and Distribution Center. Receipt of each shipment from the DLDC should be acknowledged by the site within 1 business day. The list of the package numbers of all study medication packages received in the shipment should be submitted to the Coordinating Center with 2 business days. Medication storage condition (Section 4.3.1) should be followed during the shipment receiving and accounting process.

Depending on local policy, a Clinical Site will store the medications following the study medication storage condition described in Section 4.3.1 in a secure room within their clinic or in their local investigative pharmacy. A temperature data recorder should be used in the storage room to track the storage condition (see Section 4.3.1). Every 6 months, the temperature recorder's log must be submitted to the Coordinating Center portal for review. If there is a significant duration with temperature excursion noted, the issue will be brought to the attention of the Study Chairman, Coordinating Center Director and the Site PI and Study Coordinator. A meeting will be scheduled with the Site Coordinator to identity possible reasons and determine and implement measures to prevent any additional temperature excursions.

When medication is dispensed, the Study Coordinator should complete the medication accountability form to record the study medication tri-fold package identification numbers dispensed to the participant and this form should be submitted to the Coordinating Center, following the procedures in the study pharmacy manual.

All drug storage facilities and study treatment records will be made available to the site visitor for inspection during site visits.

##### 4.14.2. General Quality Assurance Measures

The major quality assurance methods of the study are:

- Standard data collection forms and procedures;
- Common protocol for eligibility, examination, and follow-up of all patients in all clinical sites;
- Computerized treatment allocation with eligibility review preceding enrollment;
- Masked assessments at the Clinical Sites;
- Central masked grading of OCTs;
- Direct data entry into the study central database at the Clinical Sites
- Checking data entered into the study database by the Coordinating Center by comparing to the scanned copy of source forms submitted by Clinical Sites.
- Central, computer driven data editing for missing, invalid, and suspect responses;
- Regular reporting on performance of all Clinical Sites;
- On-Site and Virtual monitoring visits to all centers;
- Specific data analyses to identify incorrect or fraudulent data collection processes;
- Certification of clinic staff and of imaging equipment;
- Regular meetings of the Investigative Group to review methods and discuss problems.

The meeting of the Investigative Group is an important component of quality assurance. Such meeting will be held twice per year, for example one in-person meeting during ARVO and one virtual meeting via Zoom. These meetings provide a mechanism of sharing information among NAC Attack investigators and other personnel.

###### 4.14.3. Quality Assurance via the Steering and Quality Assurance Committee

The Steering and Quality Assurance Committee (SQAC) has responsibility for the quality assurance activities required to maintain standardization of procedures and adherence to the NAC Attack protocol. Membership and specific functions may be found in Section 10.8.2. Problems in Clinical Site performance or adherence to the protocol are normally resolved by the Coordinating Center Director and Protocol Monitor working directly with the staff of the Clinical Site. When these efforts fail, the problem is referred to the entire committee.

###### 4.14.4. Quality Assurance via Clinical Site Monitoring

Section 9.4 describes methods of clinical sites monitoring by the Coordinating Center.

###### 4.14.5. Ensuring Participant Study Medication Compliance

Study compliance includes participant acceptance of randomization and adherence to the treatment schedule. Challenges exist considering that many patients are aware of the Phase 1 FIGHT RP trial results and may misinterpret them as suggesting that NAC provides definite benefit. NAC is available as a supplement over-the-counter in pharmacies. Thus, RP patients may consider taking supplement formulations of NAC rather than participating in the NAC Attack Trial. To address these challenges, the Study design has consulted patient advocacy groups (PAGs) for measures to facilitate compliance:

- Given the perceived benefit of the study drug, NAC Attack will use a 2:1 randomization ratio between intervention and placebo. Participants thus have a 66.7% chance of receiving active drug, and this will facilitate acceptance of randomization.
- The Coordinating Center in conjunction with the Study Chairman and the PAGs will provide a script for clinical sites regarding the key points for participant compliance that must be emphasized to patients during recruitment and the informed consenting process.

###### 4.14.6. Clinical Site Measures to Prevent Drop-outs and Missed Visits

Each study visit has an allowed scheduling window (See **Figure 1**). *The Coordinator should try to schedule a visit at the beginning of the scheduling window.* This will leave more room for rescheduling if needed and help prevent a missed visit.

Each Clinical Site must make participant in-clinic visits as efficient as possible by minimizing wait time and providing comfortable waiting and examination facilities. The Coordinator and the PI for each Clinical Site will continually educate the participant as to the nature of the Study, the need for the patient's continued participation and the importance of compliance to assigned study medication, and will answer any questions related to the Study or to the disease. The Coordinating Center in conjunction with the Study Chairman will develop talking points for the Clinical Site investigator and staff to use during each participant encounter to promote study compliance and retention.

Coordinators will contact participants to remind them of a follow-up in-clinic visit within the week of the appointment and remind them to bring all used and unused study medication packages with them if this is required at the visit based on the schedules of activities at study visits (Table 1). Schedules of activities at study visits. The contact can be by telephone or email, but a participant response is necessary. Every effort must be made by the Clinical Site to remain in contact with participants, even if participants do not want to return to be examined or follow the protocol. If rescheduling of visit is needed, the rescheduling should be attempted as soon as the Coordinator learns about the need for rescheduling.

###### 4.14.7. Quality Assurance Methods Ensuring Data Integrity and Quality

Section 7.2.3 describes quality assurance measures for data management.

##### 4.15. Treatment, Patient, Study, and Site Discontinuation

###### 4.15.1. Permanent Study Treatment Discontinuation

Participants must permanently discontinue study treatment if any of the following are experienced. (1) Any medical condition that the investigator and medical monitor determines may jeopardize the patient's safety if he/she continues to receive study treatment (2) Any eventuality that causes the investigator and medical monitor to determine that treatment discontinuation is in the best interest of the patient.

The primary reason for permanent study treatment discontinuation should be documented on the Study Dose Modification Form. Participants who discontinue study treatment prematurely will not be replaced, and unless they withdraw consent, they will continue study follow-up visits as originally scheduled until they finish their M45 follow-up for safety monitoring and to collect study outcome data.

###### 4.15.2. Study Treatment Dose Modification

When managing an AE, Site investigator may consider temporarily reducing the treatment dose for the participant (see Section 5.1.2.1), but the Site Investigator should make every effort to consult the Medical Monitor, the Study Chair, or a Study Vice-Chair before making the recommendation to the participant. Any dose reduction or later increase back to 1800mg bid should be recorded in the appropriate CRFs.

Women who become pregnant during the trial should be informed to stop study medication but continue to attend all study visits. After delivery and after breast feeding is stopped, study medication may be restarted. The dose changes associated with a pregnancy can be recorded in Study Dose Modification Form.

###### 4.15.3. Participant Discontinuation from Study

Participants have the right to voluntarily withdraw from the study at any time for any reason. In addition, the investigator has the right to withdraw a patient from the study for the following reasons: (1) patient withdrawal of consent (2) study termination or site closure (3) any medical condition that the investigator in consultation with the medical monitor determines may jeopardize the patient's safety if he or she continues in the study.

Participants who withdraw from the study should have an early termination visit. The primary reason for withdrawal from the study should be documented on the appropriate CRF. Participants who withdraw from the study will not be replaced.

###### 4.15.4. Study Discontinuation

The National Eye Institute (NEI) reserves the right to curtail, withhold, or terminate support for the study in situations involving: inadequate progress toward meeting study milestones including those related to: availability and regulatory approval of study product as applicable, patient recruitment, follow-up, data reporting, or quality control; a major breach of the study protocol or NEI/NIH policy; a substantive change in the agreed-upon protocol to which the NEI does not agree; statistical evidence that the major study endpoint has been reached ahead of schedule; or, human subject ethical issues that dictate a premature termination. Prior to taking such actions, NEI will consult with and receive recommendations from the DSMC.

###### 4.15.5. Site Discontinuation

The Coordinating Center has the right to close a Clinical Site at any time. Reasons for closing a Clinical Site may include, but are not limited to, the following: (1) excessively slow recruitment (2) poor protocol adherence (3) inaccurate or incomplete data recording (4) non-compliance with the International Council for Harmonization (ICH) guidelines for Good Clinical Practice (5) no study activity (i.e., all participants from the site have completed the study and all obligations have been fulfilled). Before taking such actions, the Coordinating Center will consult with the Study Chairman and the NEI Program Officials.

### 5. Assessment of Safety

Several measures will be taken to ensure the safety of patients participating in this study. Eligibility criteria have been designed to exclude patients at higher risk for treatment related complications. Participants will undergo safety monitoring during the study including assessment of the nature, frequency, and severity of AEs by the site Investigator, the Study Chairman and the Medical Monitor. There is independent safety monitoring from the Medical Monitor and the DSMC. In addition, guidelines for managing AEs, including criteria for treatment interruption or discontinuation, are provided below.

#### 5.1. Overview of Safety Plan

Participants will have a medical history and specific questioning to elicit any AEs at each study visit after baseline. Ocular safety will be assessed by measurement of BCVA, IOP measurement, slit lamp examination and indirect ophthalmoscopy at in-clinic visits as specified in **Table 1**. Systemic safety will be assessed by medical history, vital signs, and blood chemistry, hematology, and liver function tests. All participants will be instructed to contact their Clinical Site at any time if they have any health-related concerns. If warranted, participants will be asked to return to their Clinical Site as soon as possible for an unscheduled safety assessment visit. In addition, tele-visits and study coordinator phone calls will be conducted in between in-clinic visits to inquire about AEs and compliance. A Medical Monitor and the DSMC will monitor safety throughout the trial.

##### 5.1.1. Identified Risks Associated with Oral NAC in Patients with RP

###### 5.1.1.1. Drug-related AEs in the FIGHT RP study

In the FIGHT RP study,[34] patients took the following doses of NAC: (1) 600 mg bid for 3 months followed by 600 mg tid for 3 months in cohort 1 (2) 1200 mg bid for 3 months followed by 1200 mg tid for 3 months in cohort 2 (3) 1800 mg bid for 3 months followed by 1800 mg tid for 3 months. Eight of 30 patients experienced AEs (Table 2, one patient had 2 events and one had 3 events).

**Table 2 Drug-related AEs in the FIGHT RP Study**

| Patient | Patient Reported Event | Dose | Resolution |
|---|---|---|---|
| 1-07 | Occasional flushing | 600mg TID | Spontaneous |
| 1-07 | Vomiting, headache, and flushing | 600mg TID | Spontaneous |
| 1-08 | Stomach aches | 600mg TID | Spontaneous |
| 2-02 | Constipation | 1200mg BID | Spontaneous |
| 2-11 | Nausea and diarrhea after dose escalation to TID | 1200mg TID | Dose de-escalation |
| 2-13 | Nausea and headaches after dose escalation to TID | 1200mg TID | Dose de-escalation |
| 3-05 | Burping and hiccups | 1800mg BID | Spontaneous |

| | | | |
|---|---|---|---|
| 3-06 | Diarrhea after dose escalation to TID | 1800mg TID | Dose de-escalation |
| 3-15 | Heartburn | 1800mg BID | Spontaneous |
| 3-15 | Bloating and abdominal discomfort | 1800mg BID | Spontaneous |
| 3-15 | Tiredness after dose escalation to TID | 1800mg TID | Spontaneous |

Seven of the symptoms occurred during tid dosing and 4 occurred during bid dosing. All symptoms occurring during bid dosing were mild, transient, and resolved spontaneously. Symptom severity was moderate in 2 participants in cohort 2 and one subject in cohort 3 soon after dose escalation to tid dosing; dosing was subsequently reduced to bid and the symptoms resolved. Nine of the 11 events involved the gastrointestinal (GI) system. The other two were flushing and tiredness, and were mild.

###### 5.1.1.2. Gastrointestinal (GI) Symptoms

As noted above 9 of 11 drug-related AEs involved the GI system. Since these symptoms were mild and resolved spontaneously in all patients receiving bid dosing, we do not anticipate GI symptoms being a problem, but all investigators will be made aware of the possibility of GI symptoms and vigilance for them will be requested.

###### 5.1.1.3. Exacerbation of Hypertension

Fluimucil, the effervescent tablets containing NAC, contain sodium bicarbonate which can be a sodium load that has the potential to exacerbate hypertension in susceptible patients. This was not observed in the FIGHT RP study, but is noted in the Investigator's Brochure (IB). All patients will have blood pressure measurements at every visit and patients with known hypertension will be asked to measure their blood pressure at home and report any substantial elevation in systolic or diastolic readings.

###### 5.1.1.4. Caution Using Cough Suppressant in Subjects with Upper Airway Congestion

See Section 4.7.2.

###### 5.1.1.5. Alteration of Laboratory Tests

Simultaneous use of NAC and carbamazepine can lead to sub-therapeutic carbamazepine concentration, and therefore participants on carbamazepine must have carbamazepine levels monitored and its dose adjusted if necessary.

Use of NAC may disturb measurement of ketone bodies in urine and therefore if there is suspicion of ketoacidosis, serum ketones should be measured.

Use of NAC can alter the colorimetric determination of the content of salicylates and therefore this test cannot be relied upon to evaluate for salicylate overdose.

###### 5.1.2. Management of Participants Who Experience AEs

###### 5.1.2.1. Dose Modifications

In the FIGHT RP extension trial where 30 patients have been taking 1800 mg NAC bid for at least 24 months, there was only occasional mild, transient abdominal discomfort in 3 patients that has been largely eliminated by eating prior to dosing and/or avoiding milk products prior to dosing.

In the unlikely case that a NAC Attack study participant experiences a moderate or severe AE that is persistent and has a substantial negative impact on the participant's well-being, the participant will be allowed to decrease the number of effervescent tablets from 3 bid to 2 bid at the discretion of the investigator in consultation with the Medical Monitor, Study Chair or Study Vice-Chair. If the symptoms persist, the investigator in consultation with Medical Monitor, Study Chair or Study Vice-Chair may further reduce the dose to 1 effervescent tablet bid and if symptoms still persist, may stop the study drug upon approval by the Medical Monitor. If the symptoms resolve, the investigator should sequentially increase the dose as tolerated by the participant. The reason for dose reduction should be recorded on appropriate CRF about study medication dose modification and the Adverse Event CRFs. Subsequent dose escalation should also be documented in dose modification CRF.

###### 5.1.2.2. Study Treatment Interruption

Study treatment interruption should be discussed with the medical monitor and the reason for interruption should be recorded on the associated CRF of study medication dose modification or discontinuation, and, if applicable, on the Adverse Event CRF.

###### 5.1.2.3. Recommended Management of Exacerbation of Hypertension

If a patient experiences elevation of systolic (>180 mm Hg) or diastolic (>100 mm Hg) blood pressure, they will be referred to their general medical doctor for evaluation and management.

###### 5.1.3. Emergency Medical Contact

Medical Monitor Contact Information for NAC Attack:

Medical Monitor: Dana Frank, MD.
Telephone Number: 1-410-925-3838


##### 5.2. Overview of Adverse Events Definitions

Safety assessments will consist of monitoring and recording AEs, including SAEs and AEs of special interest, performing protocol specified safety laboratory assessments, measuring protocol-specified vital signs, and conducting other protocol-specified examinations and tests that are deemed critical to the safety evaluation of the study. Certain types of events require immediate reporting to the Coordinating Center, as outlined in Section 5.4.

###### 5.2.1. Adverse Events Definition

According to the ICH guideline for Good Clinical Practice, an AE is any untoward medical occurrence in a clinical investigation subject administered a pharmaceutical product, regardless of causal attribution. An AE can therefore be any of the following. (1) Any unfavorable and unintended sign (including an abnormal laboratory finding), symptom, or disease temporally associated with the use of a medicinal product, whether or not considered related to the medicinal product (2) Any new disease or exacerbation of an existing disease (a worsening in the character, frequency, or severity of a known condition) (see Section 5.3.3.2 for more information) (3) Recurrence of an intermittent medical condition (e.g., headache) not present at baseline (4) Any deterioration in a laboratory value or other clinical test that is associated with symptoms or leads to a change in study treatment or concomitant treatment or discontinuation from study drug (5) AEs that are related to a protocol-mandated intervention, including those that occur prior to assignment of study treatment.

###### 5.2.1.1. Diagnosis versus Signs and Symptoms

A diagnosis (if known) should be recorded as the AE rather than individual signs and symptoms (e.g., record only liver failure or hepatitis rather than jaundice, asterixis, and elevated transaminases). However, if a constellation of signs and/or symptoms cannot be medically characterized as a single diagnosis or syndrome at the time of reporting, each individual event should be recorded on the Adverse Event Reporting Form CRF. If a diagnosis is subsequently established, all previously reported AEs based on signs and symptoms should be nullified and replaced by one AE report based on the single diagnosis, with a starting date that corresponds to the starting date of the first symptom of the eventual diagnosis.

###### 5.2.1.2. AEs that are Secondary to Other Events

In general, AEs that are secondary to other events (e.g., cascade events or clinical sequelae) should be identified by their primary cause, with the exception of severe or serious secondary events. A medically significant secondary AE that is separated in time from the initiating event should be recorded as an independent event on the Adverse Event CRF. For example:

- If vomiting results in mild dehydration with no additional treatment in a healthy adult, only vomiting should be reported on the CRF.
- If vomiting results in severe dehydration, both events should be reported separately on the CRF.
- If a severe GI hemorrhage leads to renal failure, both events should be reported separately on the CRF.
- If dizziness leads to a fall and consequent fracture, all three events should be reported separately on the CRF.
- If neutropenia is accompanied by an infection, both events should be reported separately on the CRF.

All AEs should be recorded separately on the Adverse Event Reporting Form CRF if it is unclear as to whether the events are associated.

###### 5.2.1.3. Persistent or Recurrent AEs

A persistent AE is one that extends continuously, without resolution, between patient evaluation timepoints. Such events should only be recorded once on the Adverse Event Reporting Form CRF. The initial severity (intensity or grade) of the event will be recorded at the time the event is first reported. If a persistent AE becomes more severe, the most extreme severity should also be recorded on the CRF. If the event becomes serious, it should be reported to the Coordinating Center immediately (i.e., no more than 24 hours after learning that the event became serious; see Section 5.4 for reporting instructions). The Adverse Event Reporting Form should be updated by changing the event from "non-serious" to "serious," providing the date that the event became serious, and completing all data fields related to serious AEs.

A recurrent AE is one that resolves between patient evaluation timepoints and subsequently recurs. Each recurrence of an AE should be recorded as a separate event on the Adverse Event Reporting Form CRF.

###### 5.2.1.4. Abnormal Laboratory Values

Not every laboratory abnormality qualifies as an AE. A laboratory test result must be reported as an AE if it meets any of the following criteria: (1) Is accompanied by clinical symptoms, (2) results in a change in study treatment (e.g., dosage modification, treatment interruption, or treatment discontinuation), (3) results in a medical intervention (e.g., potassium supplementation for hypokalemia) or a change in concomitant therapy, or (4) Is clinically significant in the investigator's judgment.

It is the investigator's responsibility to review all laboratory findings. Medical and scientific judgment should be exercised in deciding whether an isolated laboratory abnormality should be classified as an AE.

If a clinically significant laboratory abnormality is a sign of a disease or syndrome (e.g., alkaline phosphatase and bilirubin 5x upper limit of normal (ULN) associated with cholestasis), only the diagnosis (i.e., cholestasis) should be recorded on the Adverse Event Reporting Form CRF.

If a clinically significant laboratory abnormality is not a sign of a disease or syndrome, the abnormality itself should be recorded on the Adverse Event Reporting Form, along with a descriptor indicating whether the test result is above or below the normal range (e.g., "elevated potassium," as opposed to "abnormal potassium"). If the laboratory abnormality can be characterized by a precise clinical term per standard definitions, the clinical term should be recorded as the AE. For example, an elevated serum potassium level of 7.0 mEq/L should be recorded as "hyperkalemia."

Observations of the same clinically significant laboratory abnormality from visit to visit should only be recorded once on the Adverse Event Reporting Form.

###### 5.2.1.5. Abnormal Vital Sign Values

Not every vital sign abnormality qualifies as an AE. A vital sign result must be reported as an AE if it meets any of the following criteria: (1) is accompanied by clinical symptoms, (2) results in a change in study treatment (e.g., dosage modification, treatment interruption, or treatment discontinuation), (3) results in a medical intervention or a change in concomitant therapy, or (4) is clinically significant in the investigator's judgment.

It is the investigator's responsibility to review all vital sign findings. Medical and scientific judgment should be exercised in deciding whether an isolated vital sign abnormality should be classified as an AE.

If a clinically significant vital sign abnormality is a sign of a disease or syndrome (e.g., high blood pressure), only the diagnosis (i.e., hypertension) should be recorded on the Adverse Event CRF.

Observations of the same clinically significant vital sign abnormality from visit to visit should only be recorded once on the Adverse Event CRF.

###### 5.2.1.6. Abnormal Liver Function Tests

The finding of an elevated ALT or AST (> 3x ULN) in combination with either an elevated total bilirubin (> 2x ULN) or clinical jaundice in the absence of cholestasis or other causes of hyperbilirubinemia is considered to be an indicator of severe liver injury (as defined by Hy's Law). Therefore, investigators must report as an AE the occurrence of either of the following: (1) treatment-emergent ALT or AST > 3x ULN in combination with total bilirubin > 2x ULN, or (2) treatment-emergent ALT or AST > 3x ULN in combination with clinical jaundice. The most appropriate diagnosis or (if a diagnosis cannot be established) the abnormal laboratory values should be recorded on the Adverse Event CRF (see Section 5.4.1) and reported to the Coordinating Center immediately (i.e., no more than 24 hours after learning of the event) as an SAE.

###### 5.2.1.7. Deaths

All deaths that occur during the protocol-specified AE reporting period (see Section 5.3.1), regardless of relationship to study drug, must be recorded on the Adverse Event CRF and immediately reported to the IRB and the Coordinating Center (see Section 5.4.1).

Death should be considered an outcome and not a distinct event. The event or condition that caused or contributed to the fatal outcome should be recorded as the single medical concept on the Adverse Event CRF. Generally, only one such event should be reported. If the cause of death is unknown and cannot be ascertained at the time of reporting, "unexplained death" should be recorded on the Adverse Event CRF. If the cause of death later becomes available (e.g., after autopsy), "unexplained death" should be replaced by the established cause of death. The term "sudden death" should not be used unless combined with the presumed cause of death (e.g., "sudden cardiac death"). If the death is attributed to progression of diabetes mellitus, diabetes mellitus progression" should be recorded on the Adverse Event Reporting Form.

###### 5.2.1.8. Preexisting Medical Conditions

A preexisting medical condition is one that is present at the baseline visit for this study. Such conditions should be recorded on the appropriate CRF. A preexisting medical condition should be recorded as an AE only if the frequency, severity, or character of the condition worsens during the study. When recording such events on the Adverse Event CRF, it is important to convey the concept that the preexisting condition has changed by including applicable descriptors (e.g., "more frequent headaches").

###### 5.2.1.9. Hospitalization or Prolonged Hospitalization

Any AE that results in hospitalization (i.e., inpatient admission to a hospital) or prolonged hospitalization should be documented and reported as a SAE (per the definition of SAE in Section 5.2.2), except as outlined below.

An event that leads to hospitalization under the following circumstances should not be reported as an AE or a SAE. (1) Hospitalization for a preexisting condition, provided that all of the following criteria are met: The hospitalization was planned prior to the study or was scheduled during the study when elective surgery became necessary because of the expected normal progression of the disease. (2) The patient has not experienced an AE.

An event that leads to hospitalization because of patient requirement for outpatient care outside of normal outpatient clinic operating hours is not considered to be a SAE, but should be reported as an AE instead.

##### 5.2.2. Serious Adverse Events (SAE) Definition (Immediately Reportable to Coordinating Center and to the IRB)

An AE should be classified as SAE if:

- It resulted in death (i.e., the AE caused or led to death).
- It was life threatening (i.e., the AE placed the participant at immediate risk of death; this does not apply to an AE that might have caused death if it had been more severe).
- It required or prolonged inpatient hospitalization (i.e., the AE required at least a 24-hour inpatient hospitalization or prolonged a hospitalization beyond the expected length of stay; this does not apply to hospitalizations for elective medical/surgical procedures, scheduled treatments, or routine check-ups).
- It was disabling (i.e., the AE resulted in a substantial disruption of the participant’s ability to carry out normal life functions).
- It resulted in a congenital anomaly/birth defect (i.e., an adverse outcome in a child or fetus of a participant exposed to the molecule or study drug prior to conception or during pregnancy).
- It does not meet any of the above serious criteria but may jeopardize the participant or may require medical or surgical intervention to prevent one of the outcomes listed above.

The definition of a SAE focuses on the “outcome” of the event, and the SAE may involve only one, or possibly more, of the above criteria. Any adverse event that does not meet any of the criteria for an SAE should be regarded as a non-serious adverse events.

###### 5.2.3. Adverse Events of Special Interest Definition

An AE should be classified as an AE of Special Interest if:

- It is considered as potentially causally related to the study medication, i.e. it is assessed as “Possible”, “Probable” or “Definite” during attribution of causality of the event.
- It is considered SIGHT-THREATENING, as defined by the following criteria:
  - o The event causes a decrease in visual acuity of >30 letters (compared with the last assessment of visual acuity) lasting >24 hours.
  - o The event causes a decrease in visual acuity to the level of light perception or worse lasting >24 hours.
  - o The event requires medical or surgical intervention (e.g., conventional surgery, vitreous tap or biopsy with intravitreal injection of antibiotics, or laser or retinal cryopexy with gas) to prevent permanent loss of sight.
  - o The event is associated with severe intraocular inflammation (i.e., 4+ anterior chamber cell/flare or 4+ vitritis).

##### 5.3. Methods for Assessing and Recording Adverse Events

The investigator is responsible for ensuring that all AEs (see Section 5.2.1 for definition) are recorded on the Adverse Event Reporting Form and reported to the Coordinating Center in accordance with instructions provided in this section and in Sections 5.4-5.5.

###### 5.3.1. AE Reporting Period

Investigators will seek information on AEs at each patient contact. All AEs, whether reported by the patient or noted by study personnel, will be recorded in the patient's medical record and on the Adverse Event Reporting Form CRF.

The reporting period for a participant during which AEs must be reported is the period from enrollment to the end of study follow-up. Any unresolved AE must be followed by the Investigator until the events has resolved, the participant is lost to follow-up, or the AE is otherwise explained. The study site should follow up with a participant by phone if there are any ongoing active AEs or SAEs at the time of the participant’s final M45 study visit. If an AE or SAE that is felt unlikely related to study drug is still active 30 days after the final study visit, it may be listed as unresolved and the participant’s study record may be closed out. If an AE or SAE felt to possibly or probably be related to study drug is still active at the final study visit, the study site should follow up with phone calls with the participant until there is resolution or improvement in the AE or SAE. If after prolonged follow up, there is no change in the AE or SAE, it may be graded as stable and the participant’s study record may be closed out.

###### 5.3.2. Eliciting AE Information

A consistent methodology of non-directive questioning should be adopted for eliciting AE information at all participant evaluation time points. Examples of non-directive questions include the following.

"How have you felt since your last clinic visit?"
"Have you had any new or changed health problems since you were last here?"

##### 5.3.3. Assessment of AEs

Assessments of AEs include coding of the type of an AE, grading the severity of the AE, and attributing the causality of AEs.

###### 5.3.3.1. Coding Systemic and Ocular Adverse Events at Clinical Sites

All AEs will be coded using the Common Terminology Criteria for AEs (CTCAE) developed by the National Cancer Institute and used beyond oncology. The CTCAE version 5.0 provides definitions for a large subset of AE terms and a grading (severity) scale for each AE. CTCAEv5.0 and its associated grading criteria are very specific, providing an AE term (MedDRA name and number) and grade that precisely describes the event. The NAC Attack REDCap data collection system has built in the CTCAE 5.0 database. The Site Coordinator documents the medical name of the AE on the paper Adverse Event Reporting Form, and when entering it in the REDCap field, a drop-down list will be automatically provided by REDCap for the Coordinator to choose the applicable CTCAE term and grade.

###### 5.3.3.2. Grading Severity of Adverse Events by Site Investigators

All events must be graded for severity by the Site Investigator, using a 5-point scale as in Table 3:

**Table 3 Grading Severity of Adverse Events**

| |
|---|
| 1 = Mild: Awareness of sign or symptom, but easily tolerated. |
| 2 = Moderate: Interference with normal daily activities. |
| 3 = Severe: Inability to perform normal daily activities. |
| 4 = Life threatening or disabling: Immediate risk of death or disablement. |
| 5 = Death |

Note, the terms “serious” and “severe” are not synonymous. The term “severe” is often used to describe the intensity (severity) of a specific event (e.g., a mild, moderate, or severe myocardial infarction); the event itself, however, may be of relatively minor medical significance (such as a severe headache). This is not the same as “serious”, which is based on patient or event outcome or action criteria, usually associated with events that pose a threat to a patient’s life or functioning. Seriousness (not severity) serves as a guide for defining regulatory reporting obligations. When recording AEs and SAEs, severity and seriousness must be independently assessed.

###### 5.3.3.3. Attributing the Causality of Adverse Events at Clinical Sites

Investigators should use their knowledge of the participant, the circumstances surrounding the event, and an evaluation of any potential alternative causes to assess whether an AE is considered to be related to the study drug. The guidance in **Table 4** should be used when attributing the causality of an AE. Because there still can be uncertainty as to whether an SAE is related to study drug or not, a 5-point scale as in **Table 5** will be used to record the result from the attribution of causality of an AE and should be reported on the Adverse Event Report Form.

**Table 4 Guidance for Attribution of Causality of Adverse Events**

| Is the adverse event suspected to be caused by the study drug on the basis of facts, evidence, science-based rationales, and clinical judgment? | |
|---|---|
| Yes | 1) the onset is temporally related to the administration of the study drug and the AE is not explainable by the participant's concomitant conditions or therapies, ***And/or,*** 2) the AE follows a pattern of response to the study drug, ***And/or,*** 3) the AE lessens or resolves when the study drug dose or frequency is reduced or the study drug is discontinued, and the AE reoccurs when study drug is reintroduced (when applicable). |
| No | An AE will be considered related, unless it fulfills the criteria specified below. Evidence exists that the AE has an etiology other than the study drug (e.g., preexisting medical condition, underlying disease, intercurrent illness, or concomitant medication); Or, the AE has no plausible temporal relationship to administration of the study drug (e.g., cancer diagnosed 2 days after first dose of study drug). |

**Table 5 Categories for recording the result from the attribution of causality of an AE.**

| Category | Relation to study drug |
|---|---|
| Unrelated | The AE is clearly not related to the investigational agents. |
| Unlikely | The AE is doubtfully related to the investigational agents. |
| Possible | The AE may be related to the investigational agents. |
| Probable | The AE is likely related to the investigational agents. |
| Definite | The AE is clearly related to the investigational agents. |

###### 5.3.3.4. Assessment of Adverse Events by the Study Chair

The Coordinating Center will maintain a log of all AEs that contains the coding, severity grading, and causality provided by site investigators along with a brief description of the event and any follow-up. Once a month the Coordinating Center will send the updated log to the Study Chair's Office. The Study Chair and his team will review the AE log to determine if AE descriptions are consistent with site grading of severity and causality and determine if any additional information is needed from the site and if so a query will be generated. The Study Chair will look for patterns, repetitions, and other potential concerns. AEs with any grading disagreements or other concerns will be recorded by the Coordinating Center. The Coordinating Center will send all relevant information of such AEs for assessment by the Medical Safety Monitor every 3 months.

###### 5.3.3.5. Assessment of Adverse Events by the Medical Monitor

The Medical Safety Monitor will evaluate all AEs for which there is concern regarding site grading of severity and causality and make final determinations that will be sent to the Study Chair and Coordinating Center for addition to the AE log. The monitor will also evaluate any concerns, patterns, and repetitions raised by the Study Chair and provide feedback to the Study Chair, Coordinating Center, and DSMC. In cases when the evaluation of the Medical Monitor regarding causality differs from the Investigator's, the Monitor's assessment will prevail with regard to filing regulatory reports.

##### 5.3.4. Procedures for Clinical Sites Recording AEs

Investigators should use correct medical terminology/concepts when recording AEs on the Adverse Event Reporting Form CRF and avoid colloquialisms and abbreviations. Only one AE term should be recorded in the event field on the Adverse Event Reporting Form CRF. During each in-clinic or tele-visit or phone call with participants, Investigators and/or Study Coordinators will assess the occurrence, status change and resolution of AEs and SAEs by examination (if in-clinic visit) and by questioning the participant.

Complete reporting information should include the following:

- Specific condition or event and direction of change
- Grade/severity
- Event type
- Dates of onset and (if applicable) resolution
- Outcome
- Whether event necessitated a change in study treatment
- Abnormal laboratory value (for SAEs only)
- Attribution to study drug (for SAEs only)

To improve the quality and precision of acquired AE data, investigators should observe the following guidelines:

- If possible, use recognized medical terms when recording an AE. Do not use colloquialisms and/or abbreviations.
- If known, record the diagnosis (i.e., disease or syndrome) rather than the component signs and symptoms of the AE (e.g., record congestive heart failure rather than dyspnea, rales, and cyanosis). However, signs and symptoms that are considered unrelated to an encountered syndrome or disease should be recorded as individual (e.g., if congestive heart failure and severe headache are observed at the same time, each event should be recorded as an individual AE).
- Any AE occurring secondary to another process should be identified by the primary cause. A "primary" AE, if clearly identifiable, generally represents the most accurate clinical term

The Clinical Site Investigator is responsible for ensuring that all AEs that are observed during the study are recorded on the NAC Attack Adverse Event Form and in the participant's clinical record. The data in the AE Log should be entered into the REDCap database within 48 hours of the visit or call with the participant. The REDCap database AE form will perform an automatic data check during data entry to ensure that all required reporting elements have been entered into the system.

##### 5.4. Immediate Reporting Requirements of SAEs and AEs of Special Interest

Certain events require immediate reporting to allow the Coordinating Center to take appropriate measures to address potential new risks in a clinical trial. The investigator must report such events to the Coordinating Center immediately; under no circumstances should reporting take place more than 24 hours after the investigator learns of the event. The following is a list of events that the investigator must report to the Coordinating Center within 24 hours after learning of the event, regardless of relationship to study drug. Their reporting requirements are described in this section.

- SAEs (definition see Sections 5.2.2) and AEs of Special Interest (see Section 5.2.3)
- Pregnancies

###### 5.4.1. Clinical Sites Recording and Reporting SAE to the Coordinating Center

The flow of reporting of SAE from the originating clinical site is summarized in Figure 2 All SAEs should be reported on the Adverse Event Reporting Form which should be emailed to the Coordinating Center and also be data entered into the REDCap study database within 24 hours of awareness of the event. Any new record entered in the REDCap Adverse Event Reporting Form will trigger an automatic email notification sent to the Coordinating Center. The Coordinating Center will send a reminder to the study site coordinator to report the event to the local IRB if the event meets the IRB's reporting requirements.

When an SAE is associated with a hospital stay, the Study Coordinator must ask the

***Figure 2 Summary of the flow of SAE and AE of Special Interest reporting.***

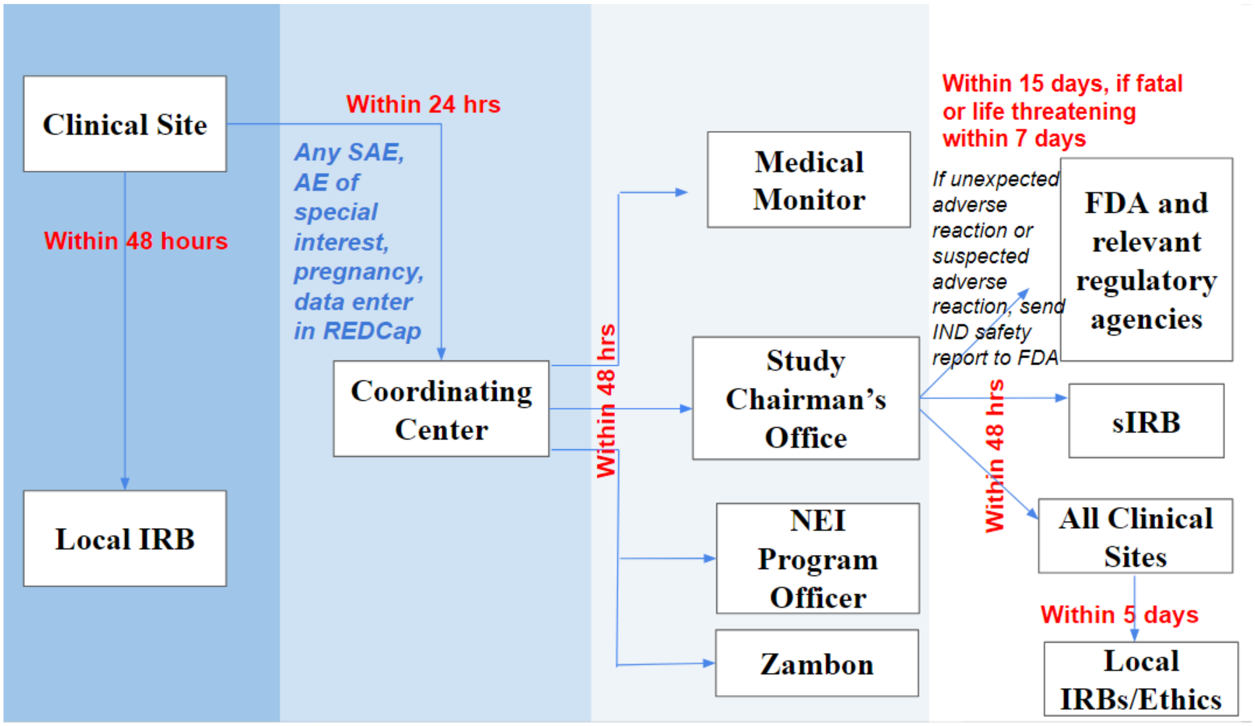

participant to sign a release to obtain the hospital discharge summary. Upon receipt, a de-identified copy of the report must be sent to the Coordinating Center and the original filed in the participant's NAC Attack file.

##### 5.4.2. Reporting Events that Occur Prior to Study Drug Initiation

After informed consent has been obtained but prior to initiation of study drug/placebo, only SAEs caused by a protocol-mandated procedure should be reported. The paper Adverse Event Reporting Form provided to investigators should be completed and submitted to the Coordinating Center immediately (i.e., no more than 24 hours after learning of the event).

##### 5.4.3. Reporting Events That Occur After Study Drug Initiation

After initiation of active drug/placebo, SAEs will be reported until the final study visit. For patients who terminate early from the study, all AEs will be collected up to the early termination visit.

All SAEs should be reported on the appropriate AE reporting form and be data entered into the REDCap study database within 24 hours of awareness of the event. Any new SAE record entered in the REDCap will trigger an automatic email notification sent to the Coordinating Center.

##### 5.4.4. Reporting Requirements for Pregnancies

###### 5.4.4.1. Pregnancies

Participants of childbearing potential will be instructed to immediately inform the investigator if they become pregnant during the study or within 28 days after the last dose of study drug/placebo. A paper Pregnancy Reporting Form should be completed and submitted to the Coordinating Center immediately (i.e., no more than 24 hours after learning of the pregnancy) by entering the form in the REDCap study database which will trigger an automatic email notification sent to the Coordinating Center.

*Pregnancy should NOT be recorded on the Adverse Event Reporting Form.* The investigator should discontinue study drug of the participant as soon as learning about the pregnancy. Monitoring of the patient should continue until conclusion of the pregnancy. Any SAEs associated with the pregnancy (e.g., an event in the fetus, an event in the mother during or after the pregnancy, or a congenital anomaly/birth defect in the child) should be reported on the Adverse Event Reporting Form. In addition, the investigator will submit a Pregnancy Reporting Form when updated information on the course and outcome of the pregnancy becomes available.

###### 5.4.4.2. Congenital Anomalies/Birth Defects and Abortions

Any congenital anomaly/birth defect in a child born to a female participant exposed to study drug should be classified as a SAE, recorded on the Adverse Event CRF, and reported to the Coordinating Center immediately (i.e., no more than 24 hours after learning of the event; see Section 5.4.1). Any abortion should be reported in the same fashion.

##### 5.4.5. Clinical Sites Reporting SAEs to Local IRB

Should an SAE occur at a participating Clinical Site, the Clinical Site must inform its local IRB within 48 hours of the investigator becoming aware of the event if required by the local IRB.

The Study Coordinator is reminded on the Adverse Event Reporting Form that if an SAE meets the requirements for local IRB notification, the SAE should be reported to the local IRB as soon as possible. The Coordinating Center Protocol Monitor will contact the Study Coordinator on a daily basis until documentation is submitted indicating that local IRB notification has been made or such notification is not required. During site visits to the clinical sites, the Protocol Monitor will ensure that documentation exists to confirm that the local IRB was notified of all reportable SAEs that occurred at the site.

###### 5.4.6. Study Chairman Reporting to the FDA: IND Safety Reports

The NAC Attack Study Chairman as the investigational new drug (IND) holder will notify the FDA in a written IND safety report of "any adverse experience associated with the use of the drug that is both serious and unexpected".

Below is abstracted from the FDA "IND Application Reporting: Safety Reports" (https://www.fda.gov/drugs/investigational-new-drug-ind-application/ind-application-reporting-safety-reports, accessed on June 06 2022)

**Adverse event** means any untoward medical occurrence associated with the use of a drug in humans, whether or not considered drug related.

**Suspected adverse reaction** means any adverse event for which there is a reasonable possibility that the drug caused the adverse event. For the purposes of IND safety reporting, 'reasonable possibility' means there is evidence to suggest a causal relationship between the drug and the adverse event. A suspected adverse reaction implies a lesser degree of certainty about causality than an adverse reaction.

**Adverse reaction** means any adverse event caused by a drug. Adverse reactions are a subset of all suspected adverse reactions where there is reason to conclude that the drug caused the event.

**Unexpected** adverse event or suspected adverse reaction refers to an event or reaction that is not listed in the investigator's brochure or is not listed at the specificity or severity that has been observed; or, if an investigator's brochure is not required or available, is not consistent with the risk information described in the general investigational plan or elsewhere in the current IND application.

**Serious** adverse event or suspected adverse reaction refers to an event or reaction that, in the view of either the investigator or sponsor, results in any of the following outcomes:

- death,
- a life-threatening adverse event,
- in-patient hospitalization or prolongation of existing hospitalization,
- a persistent or significant incapacity or substantial disruption of the ability to conduct normal life functions, or

- a congenital anomaly or birth defect.

**Life-threatening** adverse event or suspected adverse reaction is considered "life-threatening" if, in the view of the investigator or sponsor, its occurrence places the patient or subject at immediate risk of death. It does not include an adverse event or suspected adverse reaction that, had it occurred in a more severe form, might have caused death.

Important medical events that may not result in death, be life-threatening, or require hospitalization may be considered serious when, based upon appropriate medical judgment, they may jeopardize the patient or research subject and may require medical or surgical intervention to prevent one of the outcomes listed as serious.

Unexpected AE or suspected adverse reaction refers to an event or reaction that is not listed in the investigator's brochure or is not listed at the specificity or severity that has been observed.

###### Mandatory Safety Reporting

- ***Initial reporting:*** IND application sponsor must report any suspected adverse reaction or adverse reaction to study treatment that is both serious and unexpected.

  Unexpected serious suspected adverse reactions and observations from animal studies suggesting significant risk to human subjects must be reported to FDA as soon as possible but no later than within **15 calendar days** following the sponsor's initial receipt of the information.

  Unexpected fatal or life-threatening suspected adverse reactions represent especially important safety information and must be reported to FDA as soon as possible but no later than **7 calendar days** following the sponsor's initial receipt of the information.

- ***Follow-up reporting:*** Any relevant additional information obtained by the sponsor that pertains to a previously submitted IND safety report must be submitted as a Follow-up IND Safety Report. Such report should be submitted without delay, as soon as the information is available but no later than 15 calendar days after the sponsor receives the information.

All IND safety reports must be submitted on Form 3500A. FDA recommends that sponsors submit safety reports electronically.

###### 5.4.7. Study Chairman Reporting to Non-US Regulatory Agencies

In addition to Study Chairman's following the above timelines of submitting IND report to the FDA, the following timelines specific to a relevant non-US regulatory agency will also be followed by Study Chairman:

- Canada: the safety report follows the FDA timelines above. Additionally, within eight (8) days after having informed Health Canada of the IND report, Study Chairman submits a report that includes an assessment of the importance and implication of any findings.

- Mexico: the safety report of serious unexpected adverse reaction or suspected adverse reaction must be reported within a maximum of seven (7) calendar days (severe cases from abroad should only be included in the final study safety report, if the study has a research center in Mexico); Two (2) or more serious cases, in the same place with the same drug and the same batch, must be reported immediately, and no later than 48 hours.
- United Kingdom: the safety report to the Medicines and Healthcare products Regulatory Agency follows the FDA timelines above. Any additional relevant information should be sent within eight (8) days of the initial report.
- European Union countries: for Suspected unexpected serious adverse reactions (SUSARs), they should be reported to EudraVigilance instead of separately to each involved EU country. The reporting timelines are the same as the FDA timelines above. In addition, in the case of a suspected unexpected serious adverse reaction which was initially considered to be non-fatal or non-life threatening but which turns out to be fatal or life-threatening, as soon as possible and in any event not later than seven days after the sponsor became aware of the reaction being fatal or life-threatening.
- Switzerland: report all serious (expected or unexpected) and all non-serious + unexpected adverse drug reactions (ADRs) occurring in Switzerland. In addition, ADR(s) for which an unexpected increase in frequency is observed or clusters of expected or unexpected ADR(s) as well as unexpected rises of serious cases of misuse or abuse of a medicine should be reported. Timelines are: 15 days for all serious ADR(s); 15 days for clusters of expected or unexpected ADR(s) ; 60 days for non-serious unexpected ADR(s). Additionally, SUSARs resulting in death: Initial notification within 7 days, follow-up report within 8 days after initial notification (even if no influence on causality assessment).

###### 5.4.8. Coordinating Center and Clinical Sites Reporting IND Safety Reports to the IRBs

The Study Chairman's office should submit any IND Safety Report of any adverse experience associated with the use of the drug that is both serious and unexpected to the sIRB following the submission guidelines of the study sIRB.

The Coordinating Center should also send the IND Safety Report to all participating Clinical Sites within 48 hours. Each Clinical Site is responsible for copying the IND report and submitting the copy to their local IRB within 5 days (or shorter if the local IRB requires a shorter reporting period). The original report and dated documentation of IRB submission (via cover letter) must be maintained at the clinical site. During site visits to the clinical sites, the Protocol Monitor will ensure that documentation exists to confirm that the local IRB was notified of all reportable SAEs.

###### 5.4.9. Coordinating Center Reporting SAEs to the DSMC

The Director of the Coordinating Center will inform the full DSMC in writing of all SAE reports at semi-annual committee meetings. The Medical Monitor may also, at his/her discretion, instruct the Coordinating Center to notify the full DSMC immediately of an SAE, and may request a meeting or teleconference of the committee prior to its next scheduled meeting.

The following information will be provided to the DSMC by the Coordinating Center:
• Clinical Site
• Patient ID Number
• Description of event
• Date of onset
• Current status
• Severity of the event
• Whether study treatment was discontinued
• Medical Monitor's assessment of association between SAE and study drugs
• Reason the event is classified as serious

#### 5.5. Follow-up of Participants After AEs

##### 5.5.1. Investigator Follow-up

The investigator should follow each AE until the event has resolved to baseline grade or better, the event is assessed as stable by the investigator, the patient is lost to follow-up, or the participant withdraws consent. The study site should follow up with a participant by phone if there are any ongoing active AEs or SAEs at the time of the participant's final M45 study visit. If an AE or SAE that is felt unlikely related to study drug is still active 30 days after the final study visit, it may be listed as unresolved and the participant's study record may be closed out. If an AE or SAE felt to possibly or probably be related to study drug is still active at the final study visit, the study site should follow up with phone calls with the participant until there is resolution or improvement in the AE or SAE. If after prolonged follow up, there is no change in the AE or SAE, it may be graded as stable and the participant's study record may be closed out.

During the study period, resolution of AEs (with dates) should be documented on the Adverse Event Reporting Form CRF and in the patient's medical record to facilitate source data verification.

All pregnancies reported during the study should be followed until pregnancy outcome.

##### 5.5.2. Coordinating Center Follow-up

For SAEs and pregnancies, the Coordinating Center may follow up with the clinical sites by telephone, fax, email, and/or a monitoring visit to obtain additional case details and outcome information (e.g., from hospital discharge summaries, consultant reports, autopsy reports) in order to collect information for the Study Chair and Medical Monitor to perform an independent medical assessment of the reported case.

#### 5.6. Annual Report to the FDA and Relevant Foreign Regulatory Agencies

##### 5.6.1. Annual Report to the FDA

Every year, within 60 days of the anniversary date of the IND (January 14 2017), the Study Chairman and the PI of the Coordinating Center will submit to the FDA a report that includes a status report for the study as well as annual summary that includes:
- Tables of the most frequent and serious SAEs by body system
- Summary of all IND Safety Reports
- A list of deceased participants and causes of death
- Drops-out due to AEs
- (If relevant) a description of new understanding of the study drugs' actions

###### 5.6.2. Annual Report to non-US Regulatory Agencies

Each non-US regulatory agency relevant to NAC Attack has its own annual reporting requirement. The Study Chair's Office and the Coordinating Center will work with the local country's chief investigator to ensure prompt reporting the relevant regulatory agency.

### 6. Statistical Considerations and Analysis Plan

#### 6.1. NAC Attack Primary Outcome: the ICH E9(R1) estimand framework

The ICH E9(R1) addendum aims "to improve the planning, design, analysis and interpretation of clinical trials" by presenting a structured framework, the estimand framework, to link trial objectives with trial design and tools for estimation and hypothesis testing.[43] Estimand refers to "A precise description of the treatment effect reflecting the clinical question posed by the trial objective. It summarizes at a population-level what the outcomes would be in the same patients under different treatment conditions being compared."[43] At time of developing of the study design, the estimand framework required clear delineations on 4 attributes of the trial: **A: Population; B: Variable; C: Intercurrent event;** and **D: Population-level summary for the variable**. Each attribute applied in NAC Attack is described in the following sections.

##### 6.1.1. Estimand-framework Attribute A: NAC Attack Study Population

The NAC Attack study population is patients with RP and enrolled in NAC Attack.
It is important to note that RP is a clinical diagnosis. About 30-40% of patients diagnosed with RP do not have an identifiable RP-causing pathogenic variant. NAC Attack enrollment criteria do not require that patients have a known RP-causing pathogenic variant identified. This is because the proposed mechanism of NAC's slowing cone degeneration in RP is reduction of the progressive oxidative damage thereby reducing loss of function and preventing death of cones. This mechanism is independent of the genetic variant responsible for rod cell death. Therefore, the NAC Attack study population, which are not limited to RP patients with known genotype, will allow the evaluation of a pharmacological approach that, if proven efficacious, will lead to a general therapy benefitting RP patients with any pathogenic genetic variant and for RP patients in whom the pathogenic variant is unknown.

##### 6.1.2. Estimand-framework Attribute B: NAC Attack Primary Endpoint

###### 6.1.2.1. Primary outcome: considerations of OCT based anatomic measure versus visual function measures

The primary outcome measure of NAC Attack is the width of the EZ on a horizontal OCT scan through the foveal center. Regulatory agencies prefer functional over anatomic or structural endpoints.[44] However, RP is a slow progressing disease and using functional outcomes in RP treatment trials may be impractical and expensive as many years would be needed to show efficacy. In RP, progressive visual field loss is a defining feature of the disease, but the rate of visual field loss is slow in the disease natural history.[45] Based on the natural history data of visual field loss in RP, we have calculated that a trial with an outcome measure based on visual field testing would require a large and impractical sample size (>1000) to identify an efficacy of 30% of slower loss of visual field comparing a treatment group to the placebo group. The BCVA has been the gold standard endpoint in ophthalmic trials because of its strong correlation with ability to function in activities of daily life,[46,47] but in RP, BCVA is excellent until late in the disease and is expected to show no substantial decline for decades in the RP patient population that we are interested in studying.[45]

Macular sensitivity measured by MP may provide a useful functional outcome in inherited retinal diseases.[44] However, in RP, it is noted that some patients may not be able to perform MP test reliably as identified by a consistently high false-positive response rate.[48,49]This is because measurement of macular sensitivity requires the patient to push a button when they detect a light stimulus and there are factors other than visual function that can influence the responses such as alertness, fatigue, manual dexterity, performance anxiety, and decisiveness. In FIGHT RP, 10% of patients had a high rate of false-positive responses in both eyes in multiple MP tests and were unable to provide reliable data. This inability to decide when a light is seen is not related to visual function, but rather seems related to higher order processing and decision-making. Therefore, using a microperimetry based parameter as a primary outcome measure would restrict patient enrollment criteria.

Additionally, visual function measurements are subject to influences of confounding pathologies in addition to cone function and survival. Patients with RP often develop PSC and cystoid macula edema (CME) which have a negative impact on visual function. Different clinical sites may have different practices in managing cataract and CME. Thus, factors such as progression of lens opacity, cataract surgery, and fluctuations in CME may confound the estimation of the effect of the intervention on visual function measurements of BCVA or macular sensitivity outcomes.

The EZ represents the ellipsoid region of the inner segments of photoreceptors and is recognized as a well-defined bright line on a SD-OCT scan. The EZ width is measured as the horizontal extent of the EZ on a SD-OCT scan through the foveal center and provides a measurement of remaining photoreceptors with intact inner and outer segments in the horizontal meridian through the fovea. Thus, the EZ-width is an anatomic measure that reflects the integrity of photoreceptors. It is selected as the primary outcome measure for NAC Attack with the following considerations:

- EZ width correlates with visual function outcomes and has been accepted by the FDA as a surrogate endpoint in the context of geographic atrophy due to AMD.[44]
- EZ width can be reliably measured in patients with RP.[50] In the FIGHT-RP study, the intra class correlation coefficient was 0.98 baseline EZ width measurements graded for two baseline images by different graders.
- Natural history studies in patients with RP have shown that loss of EZ width is detectable during one to a few years of follow-up.[51-53]
- The FDA has agreed that assessment of loss of EZ is an appropriate outcome measure for evaluating treatment effect in RP.

###### 6.1.2.2. Primary outcome: considerations of EZ-width versus EZ-area

Recently, consideration has been given to the area of preserved EZ as an outcome measure for RP.[54] The EZ area can be measured on en-face SD-OCT images and is a measurement of the area in the posterior retina where there are remaining photoreceptors with intact inner and outer segments junction.

We considered the following aspects about the measures of EZ-width and EZ-area with respect to their suitability as NAC Attack's primary outcome measure.

- Reliability of both measurements is high.
  Based on the literature, the test-retest repeatability was comparable, and the grade-regrade reproducibility intraclass correlation coefficient was >0.99 for both measures.[42]

- Correlation between the EZ-width and EZ area measures is high.
  In two independent studies, both measures were obtained using the same images. The correlation coefficient between the two measures was 0.96 in both studies. [54 42] This suggests that the two measures are highly linearly correlated, and thus provide a similar amount of information of data.

- The data distribution of EZ-width makes it statistically more powerful and thus requires a smaller sample size.

  Tee et al. (2019) [42] provides key statistics for EZ-width and EZ-area measurements that were obtained on the same set of images of 56 eyes. These statistics provide a fair comparison of the distributions of the two measures: the annual change of EZ-width had a mean ± standard deviation (SD) of 233.55±189.13 µm; the annual change of EZ-area had a mean ± SD of 0.67 ± 0.95 $\mu m^2$. Therefore, the coefficient of variation of the change of EZ-width (0.89) is lower than the coefficient of variation of the change of EZ-area (1.42). Due to the lower coefficient of variation, a smaller sample size will be required to detect the same level of difference between the intervention and control groups in the rate of loss in EZ width as compared to the rate of loss in EZ area. Alternatively, given a fixed sample size, the power to detect the efficacy will be much larger using the loss of EZ measured by EZ-width as compared to using EZ-area.

- EZ-width measurement has stronger structure-function correlations than EZ-area.

- Using EZ-area as a primary outcome measure will further restrict enrollment criterion.
  The study uses a fixed OCT scanning window. To fully capture the progression of EZ loss during follow-up, one eligibility criterion is that impairment of EZ should be already observable at enrollment. This means that if the EZ-area measurement was used, it would require observable EZ impairment on all the B-scans, whereas if the foveal-centered EZ-width measurement was used, it would only require observable EZ impairment on the B-scan through the fovea. Reading centers have informed us that about 10-15% eyes that have the fovea EZ-width within the OCT scan window would have a portion of reserved EZ-area extends outside the OCT scan window and thus would not be eligible if using the EZ-area as the primary outcome measure.

In summary, EZ-area reflect the preservation of the EZ of the entire macula, whereas foveal-centered EZ-width is a measurement of the EZ on the horizontal meridian through the foveal center. However, numerically, these two variables are highly correlated, using EZ-width is statistically more efficient and incurs less stringent enrollment criteria. EZ-width on the horizontal fovea scan therefore is determined to be the primary outcome measure in NAC Attack.

###### 6.1.2.3. NAC Attack Primary Endpoint Derived from the Primary Outcome of EZ-width

The primary outcome of NAC Attack is the EZ-width on the fovea horizontal OCT scan. Because it is unknown whether the loss of EZ-width over 45 months would follow a linear trajectory, the rate of EZ width loss per year would not be an appropriate parameter as "rate" implies linear change over time. An alternative variable is the change of EZ-width from the baseline to the M45 follow-up. However, the FDA recommended that the primary outcome measure should be assessed at a minimum of 5 follow-up time points with 2 adjacent time points at least 9 months apart and that the efficacy should reflect separation of EZ loss curves for the intervention versus the placebo groups. Therefore, we decided to use the derived variable of Area Above the Curve (AAC) which can be interpreted as the cumulative loss of EZ over the M45 of follow-up. Figure 3 illustrates the AAC.

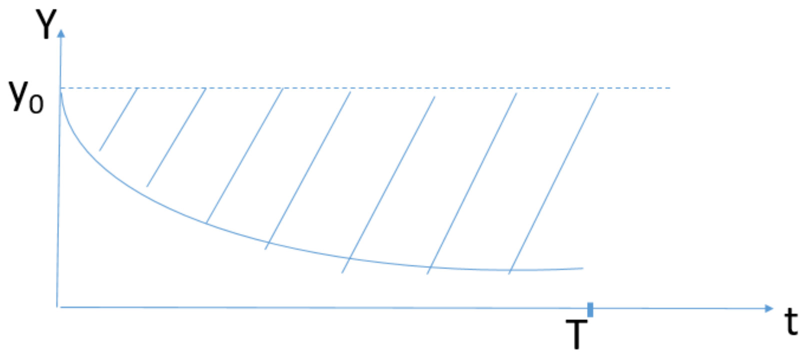

*Figure 4 Illustration of the concept of Area Above the Curve (AAC). The X-axis is follow-up time, the Y-axis is the measurement of EZ-width. $y_0$ is baseline EZ-width. If there is no loss of EZ during the follow-up, the measurement of EZ width would be the dotted line. The real line curve is the trajectory of EZ-width during follow-up. The shaded area above the curve, i.e. the AAC is the cumulative loss of EZ-width during the follow-up up to time T. Note, AAC is a derived variable specific to a pre-determined time period T.*

The outcome measure is repeatedly measured at multiple visits. The AAC statistic is essentially a summary statistic that uses a scalar to summarize the repeated outcome measurements for the same eye. A similar statistic, the Area Under the Curve (AUC), has been noted in the statistical literature,[55 56] and was used as estimands in other ophthalmic trials. [57,58] The AAC is the complement (as compared to the baseline value) of AUC. Statistically, AAC and AUC are both linear combinations of the repeated outcome measurements and thus have similar statistical properties. We choose to use AAC because it is clinically more interpretable in the context of NAC Attack's study: in RP natural history, EZ-width declines over time. The clinical hypothesis for testing the NAC treatment is it will slow the course of loss of EZ-width and thus slow the loss of visual function. The loss of EZ at each time point can be quantified by the difference between baseline and the follow-up measurements, and the AAC statistic quantifies the cumulative losses over all follow-up times.

Operationally, to estimate the AAC, the area can be non-parametrically estimated as the sum of trapezoids (Figure 4). Let $y_j$ be the outcome variable measured at visit $j$, $j = 0,1,2,\cdots,K$. The interval length, denoted as $t$, is pre-specified and equally spaced. The AAC then can be estimated empirically as $\left(\sum_{j=1}^{K-1}(y_o - y_j) + \frac{1}{2}(y_0 - y_K)\right)\cdot t$. $t$ is fixed and equal over all intervals by design. In practice, there is an allowed scheduling window for each follow-up visit, thus $t$ can differ by a few days. However, the EZ-loss is a slow process, and thus $t$ (in the unit of month) can be treated as fixed. Therefore, without loss of generalizability, AAC is operationally estimated as $\left(\sum_{j=1}^{K-1}(y_o - y_j) + \frac{1}{2}(y_0 - y_K)\right)$. It is the cumulative loss of EZ-width during the study follow-up period.

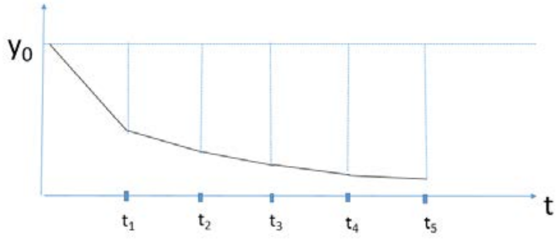

*Figure 5 Estimating the Area Above Curve with outcome measurements observed at pre-specified follow-up visits. The AAC can be empirically and non-parametrically estimated as the sum of all the trapezoids determined by the measurements at all visits.*

Note, in RP's natural history, EZ width decreases over time. Under the intervention condition with NAC, it is unexpected to have a gain in EZ-width, but should this happen at an early visit, the AAC statistic can still be calculated as (the area of loss- the area of gain), i.e. ($B$ - $A$) as shown in Figure 5, where the area of $B$ is

$$B = \frac{t}{2}\cdot\left[(y_0 - y_2) + 2(y_0 - y_3) + 2(y_0 - y_4) + (y_0 - y_5)\right] + [\frac{t}{2}\cdot(y_0 - y_2)\cdot(1 - \frac{(y_1 - y_0)}{(y_1 - y_2)})]$$

And the area of $A$ is

$$A = \frac{t}{2}\cdot(y_1 - y_0) + (y_1 - y_0)\cdot\frac{(y_1 - y_0)}{(y_1 - y_2)}\cdot\frac{t}{2}$$

Therefore, for the situation in Figure 5,

$$\begin{aligned} AAC = B - A = &\left[\frac{t}{2}\cdot\left[(y_0 - y_2) + 2(y_0 - y_3) + 2(y_0 - y_4) + (y_0 - y_5)\right] + \frac{t}{2}\cdot(y_0 - y_2)\cdot(1 - \frac{(y_1 - y_0)}{(y_1 - y_2)})\right] \\ &-\left[\frac{t}{2}\cdot(y_1 - y_0) + (y_1 - y_0)\cdot\frac{(y_1 - y_0)}{(y_1 - y_2)}\cdot\frac{t}{2}\right] \\ \propto &\left[\left[(y_0 - y_2) + 2(y_0 - y_3) + 2(y_0 - y_4) + (y_0 - y_5)\right] + (y_0 - y_2)\cdot(1 - \frac{(y_1 - y_0)}{(y_1 - y_2)})\right] \\ &-\left[(y_1 - y_0) + \frac{(y_1 - y_0)^2}{(y_1 - y_2)}\right] \end{aligned}$$

*Figure 6 Estimating the Area Above Curve if there is an increase in EZ-width measurement*

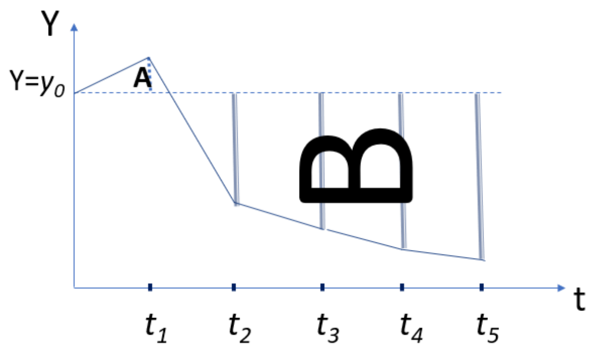

##### 6.1.3. Estimand-framework Attribute C: Possible Intercurrent Events in NAC Attack

The third attribute of an “estimand” is “intercurrent events” which refer to “Events occurring after treatment initiation that affect either the interpretation or the existence of the measurements associated with the clinical question of interest.” The Addendum recommends to clearly describe the envisioned intercurrent events and the planned strategies to account for the intercurrent events at the design stage. Different strategies can reflect different questions being answered, some of which may not be of regulatory interest. Therefore, it is important to specify the specific strategy to be used for each intercurrent event in the protocol.

Possible intercurrent events in NAC Attack:

- Participant taking his/her assigned medication but not at the prescribed level or frequency, i.e. sub-optimal medication adherence. This may include participant missing one or more doses or participant taking the assigned medication at a lower than prescribed level. NAC Attack requires chronic dosing of 1800mg/BID per day (either the active intervention or placebo control) during the 45 months follow-up period. The study will use 3 methods to monitor adherence: participant self-report via regularly scheduled mini-surveys using SMS messaging, medication reconciliation via pill counting by clinical site staff at each in-clinic visit, and biological measurement of plasma NAC level using the blood sample collected at each in-clinic visit. However, no method can measure adherence for each dose of each day. Therefore, the intercurrent event due to lack of adherence cannot be accurately and precisely measured. Moreover, in practice, a treatment can only work if patients adhere to its prescribed dose appropriately. Thus, to account for the adherence associated intercurrent event, the primary analysis will use the “Treatment policy strategy” where the observed values for the primary and secondary outcome measures will be used irrespective of the three adherence measures. The “Treatment policy strategy” reflects the well-known ICH E9 Intention-To-Treat (ITT) principle. [43]

  On the other hand, it is of great clinical interest to estimate the treatment effect if the prescribed dosage can be consistently adhered to during the study follow-up. Such clinical interest is also practically meaningful. Therefore, the treatment effect under the “Hypothetical strategy” is also of interest. Intensive efforts will be made to keep participants engaged and to promote adherence during the study. Supplementary analysis will also be conducted to estimate the treatment effect under hypothetical strategy by taking into account of adherence, i.e. using the Per-Protocol Set. [59]

- In addition to the aforementioned suboptimal adherence, another type of participant incompliance that will be detrimental to the ability of estimating treatment effect is participants randomized to the control arm self-taking NAC supplement. NAC is an antioxidant supplement available over the counter and can be purchased from online stores. If participants randomized to the placebo group arm take NAC rather than the assigned placebo, their data will dilute any estimate of the treatment effect. The measurements of plasma NAC using samples obtained during in-clinic visits can help identify incompliance of control arm participants. If NAC is identified using any in-clinic visit sample for a control arm participant, that participant will be deemed as incompliant for the trial.

  To account for intercurrent event of this kind of incompliance in the control group, we plan to use the "Hypothetical strategy" which is the scenario that such incompliance would not occur. That is, if incompliance of a control arm participant is detected through plasma NAC level at a visit, the participant's observed data on the primary and secondary outcome measures at this and subsequent visits will not be used. Instead, the primary analysis will use multiple imputation to impute the participant measurements at the M45 visit (See Section 6.1.5.1). Calculation of the AAC for the primary outcome variable will follow Scenario A in Section 6.1.5.1. The imputation in this case will be conducted because of the participant's self-uptake of a non-study NAC medication. Such a decision is expected to be independent of the primary and secondary outcome measures which are based on objective imaging or visual function examinations. Thus, the Missing-at-Random assumption for using multiple imputations to impute the M45 visit data should be reasonable. Imputing data however will increase the variability in the outcome measurements thus reducing study power.

  Sensitivity analysis will also be conducted by excluding these incompliant control participants in the analysis dataset (i.e. Per-protocol Analysis Set). [59] Excluding the incompliant control participants may introduce unbalance between the intervention arm and the remaining control arm participants thus introducing bias.

  In summary, potential incompliance of control arm participants because of the availability of NAC outside of the trial context will cause protocol deviation and is particularly detrimental to the evaluation of treatment effect by introducing bias and reducing power. To minimize the potential occurrence of such problem, we have involved patient advocacy groups (PAGs) at the NAC Attack's planning stage and developed a plan to minimize study incompliance. Throughout NAC Attack, the PAGs will be a critical bridge between the trial investigators and the patients' community.

- Intercurrent event due to pregnancy. Given that RP is an inherited condition that can impact people of all ages, women of childbearing potential will be enrolled into the study. Female participants are told to use contraceptive measures during heterogeneous intercourse during the study, but should pregnancy occur during the follow-up, the participant will be instructed to stop the study medication. Such participants will continue study visits and procedures and will resume their study medication after pregnancy and breastfeeding.

We expect pregnancy event to be rare. Primary analysis accounting for this type of intercurrent event will use the “Hypothetical strategy” which is the scenario that pregnancy had not occurred and the participant had stayed on study drug. That is, the participant’s observed data on the primary and secondary outcome measures during pregnancy and breasfeeding (i.e. when study drug is paused) will not be used. Instead, the primary analysis will use the participant’s outcome data when she has resumed study medication for at least 3 months. If at the M45 visit, the participant has not resumed study medication for 3 months yet, multiple imputation will be used to impute the participant measurements at this visit (See Section 6.1.5.1).

Supplementary analysis will also be conducted to estimate the treatment effect under “Treatment policy strategy” by using pregnant participants’ outcome data measured during their pregnancy and breastfeeding (i.e. when study medication was paused).

Additionally, sensitivity analysis will be conducted by excluding the participants who experience pregnancy during the study follow-up.

- For intercurrent event due to study treatment discontinuation for managing AEs, the “Treatment policy strategy” i.e. ICH E9 Intention-To-Treat (ITT) principle will be used for data analysis.

In addition to the above foreseeable situations leading to intercurrent events, unpredictable situations may arise. Before the data are unmasked, a summary of all situations that lead to protocol deviations will be discussed. Strategies to deal with unpredicted situations will be determined and documented in the protocol amendment.

###### 6.1.4. Estimand Framework Attribute D: Population-Level Summary for the Primary Endpoint of Area Under the Curve

The difference between the AAC in the intervention group and the AAC in the control group over the 45 months follow-up period is the population-level summary for the variable for the primary outcome in NAC Attack.

###### 6.1.5. Missing Data

In NAC Attack’s trial design, we adopt multiple approaches to promote participants retention and compliance and reduce the chance of missing data. These include frequent interactions with participants though Coordinator’s phone calls every 2.25 months, televisits with Ophthalmologists every 4.5 months, SMS based daily medication reminders and a weekly mini-survey of medication adherence, and engagement of PAGs to help educate patients regarding the necessity of a randomized placebo controlled clinical trial before NAC can be taken as a long-term medication. However, loss to follow-up and intercurrent events will occur resulting in missing data. Missing data can arise at the participant-visit level and are differentiated into 2 scenarios:

> Scenario A: a participant remains in the study but misses one or more intermediate in-clinic visits but the last visit at the M45 follow-up is observed. In this case, the AAC statistic over M45 will be calculated as the sum of the trapezoids determined by data from the observed visits. This implies that the outcome measure at the missed visit is imputed by linearly interpolating the outcome measures at the 2 adjacent observed visits.
>
> Scenario B: a participant misses the last visit at the M45 follow-up. In this case, the AAC over M45 is inestimable. For this reason, the last visit is of particular importance, and this is the reason why we have designed to allow a wider scheduling window for the M45 visit. We also institute an in-clinic visit at M40.5 where OCT imaging, MP and BCVA exam should be conducted. If the M45 OCT of an eye is unavailable, the OCT at M40.5 will be graded and measurement from the M40.5 OCT will be used to as the primary outcome measure at the M45 visit; if M40.5 OCT is also unavailable, the Full-Analysis Set will use multiple imputation to impute the missing values at the M45 visit. [60]

###### 6.1.5.1. Multiple Imputations for Generating the Full-Analysis Set

SAS Proc MI will be used for multiple imputations. To develop the imputation model, auxiliary variables will be identified by comparing participants who are observed at the M45 visit to those who miss the last follow-up visit. The comparison will include all variables collected at baseline as well as the indicator for treatment group. Auxiliary variables will also include baseline variables that are known to be associated with the outcome measure and include baseline variables that will be adjusted in the analytical model (i.e. the model for estimating treatment effect). [61] Since participants may miss an intermediate in-clinic visit at varying times during the follow-up, the imputation model will marginally model the outcome measure (i.e. EZ-width) as a function of follow-up time (denoted as $t$) and the other aforementioned auxiliary variables.

It is likely a linear model with respect to $t$ for the imputation model is not ideal. Therefore, multiple variables linear model and non-linear models (including quadratic model and exponential model) with respect to *t* will be fit and used to predict the observed values at the intermediate visits for those participants who miss the M45 visit. The model that has the smallest prediction error will be used as the imputation model to impute the missing outcome measure at the M45 visit for these participants. The fully conditional method using the predictive mean matching [62] may be used to generate the imputed values.

It has been recognized that a small number of imputations may be inadequate for inferential goals of confidence interval and p-value estimations. [61,63] Considering the Relative Efficiency Estimates [61,64], we will generate 50 imputation datasets.

The analysis model (Section 6.5) will be applied on the Full-Analysis-Set from each imputation. The final results will be obtained following Rubin's rule. [60]

###### 6.1.5.2. Sensitivity Analysis for Missing Data

Multiple imputation is a common method to deal with missing data and requires the assumption of Missing at Random (MAR). Sensitivity analysis will be conducted to examine the robustness of the primary analysis results.

Sensitivity analysis will use the tipping point analysis with the imputed datasets from the multiple imputations.[64,65] Briefly, from pulling the multiple imputation datasets, the mean difference between the intervention and the control arms at the M45 visit can be estimated, and this estimate is denoted as $\delta_{m45}$.

$R, R \geq 0$ is a sensitivity analysis tuning parameter. $R = 0$ suggests no deviation from the originally imputed value, $R = 1$ suggests the intervention arm participant would be imputed by the same value for the outcome measure at M45 visit as the mean of the control arm participants. For a range of $R$ values, the analysis to estimate the treatment effect can be conducted by pulling the estimation results from the multiple imputed datasets where $\hat{y}*_{m45}$ replaces $y*_{m45}$ for the intervention arm participants who miss the M45 visit. The tipping point $R$ value is the value that the treatment effect is not statistically significant anymore; and the corresponding $\hat{y}*_{m45}$ can be obtained. The biological plausibility of $\hat{y}*_{m45}$ at the tipping point will be discussed in consideration of the knowledge of RP's disease natural history.

#### 6.2. NAC Attack Secondary Outcome Measures

The ICH E9 (R1) specifies that estimands for secondary trial objectives (i.e. related to secondary variables) that may support regulatory decisions should also be defined and specified explicitly. NAC Attack secondary outcome measures include macular mean sensitivity (MMS) obtained from the MAIA microperimetry test and BCVA.

Outcome measures that provide a clinically meaningful assessment of visual function are important. Constriction of visual fields leads to progressive disability in patients in RP and is therefore a valuable functional outcome measure. Our study population consists of patients with moderately advanced RP who have photoreceptor degeneration extending into the central 30˚ and most of these patients have visual field defects within the central 20 ˚ or 30˚ and show progression over time. We will use MAIA MP to assess central visual fields. This instrument has the advantage of measuring retinal sensitivity over a wide dynamic range at 68 central loci distributed in the central 20˚ of the field. In the FIGHT RP study patients in the third cohort showed significant improvement in mean macular sensitivity (MMS) during treatment with 1800 mg BID NAC for 3 months followed by 1800 mg TID for 3 months. [48] In the current trial, we hypothesize that NAC-treated patients will show an initial small improvement in MMS that may remain stable or may slowly decrease over time, but if it decreases, the decrease will be smaller than that in placebo-treated patients during the 45 months follow-up period.

Visual acuity is spared until late in RP. As was the case in FIGHT RP, [48] it is anticipated that most NAC Attack participants will have excellent BCVA at baseline. In FIGHT RP, it was surprising to see that despite excellent baseline BCVA, there was a steady, small and statistically significant improvement during the 6-month NAC treatment period in each of the three cohorts.[48] This indicates that even the baseline BCVA is expected to be excellent, it is useful to measure BCVA over time in NAC Attack, which is also important for safety evaluation of any intervention.

##### 6.2.1. NAC Attack Study Populations for the Secondary Outcome Measures

###### 6.2.1.1. Population for Best Corrected Visual Acuity

The “population” for BCVA is patients with RP and enrolled in NAC Attack.

###### 6.2.1.2. Population for Macular Mean Sensitivity

The “population” for MMS is patients with RP, enrolled in NAC Attack and who can complete the MAIA test reliably (the false positive rate at the optic nerve is >30% per the recommendation from the manufacture’s Operation Manual 2013 on reliability of a test based on false positive responses).

##### 6.2.2. The Variables for the Secondary Outcome Measures

The variables for the secondary outcome measures are the change in BCVA from baseline to M45 visit* and the change in MMS from baseline to M45 visit*, respectively. (*Note: to improve statistical power, BCVA and MMS measured at the baseline visit will be used; but for the M45 visit, the average of BCVA values measured at the M40.5 and the M45 visits and the average of MMS values measured at the M40.5 and the M45 visits will be used to calculate the changes from baseline to M45 visit in BCVA and MMS, respectively).

Based on FIGHT-RP study observations, MMS and BCVA may improve during the first months of medication in the intervention arm. The improvement may reach a ceiling as improvement is unlikely unlimited during a period of 45 months. It is of clinical interest to understand whether visual functions can be better maintained in the intervention arm than the control arm over a long period of treatment. Therefore, the change between baseline and the last visit at M45 is determined to be the variable for both the BCVA and MMS. This decision received agreement from the FDA during the EOP meeting for the IND.

##### 6.2.3. Intercurrent Events for the Secondary Outcome Measures

The intercurrent events described in Section 6.1.3 for the primary outcome measure also apply to BCVA and MMS.

For MMS based on MAIA MP, it is expected that participants who can complete the test reliably at baseline will also be able to complete the test at follow-ups. Participant who are unable to provide reliable data from the MAIA MP test at Baseline visit do not need to undergo MAIM MP in his/her follow-up visits. For a participant who provides reliable MAIA MP data at Baseline, it is very unlikely that he/she cannot do the test in follow-up reliably based on the experiences from the Study Chair and the NAC Attack planning advisors, and the participants should always be administered the MAIA test at a follow-up in-clinic visit, irrespective whether the participant's MAIA test result is reliable at the previous follow-up visit. In the unlikely situation that the test result is unreliable for one or both eyes of a participant (false positive response at the optic nerve>30% per the manufacture recommendation) at the M45 visit and M40.5 visit, the MMS measurement for this visit will be unavailable. The "Principal stratum" strategy will be applied where the target population (Section 6.1.3) for the outcome of MMS is taken to be the "principal stratum" in which MMS can be reliably measured.

###### 6.2.4. Population-Level Summary for the Variables of the Secondary Outcome Measures

For each of the secondary outcome measures (BCVA and MMS), the variable is the change between baseline and the M45 visit. The difference in the change between the intervention group and the control group is the population-level summary statistic for the variable for each of the secondary outcomes.

###### 6.2.5. Missing Data

The plan of handling missing data described in Section 6.1.5 applies here for the secondary outcomes. In brief, if the M45 measurement is unavailable, the measurement at M40.5 will be used to impute the measurement for M45; If measurements at both visits are available, the average of the two visits will be used in place of M45 to increase power; and if measurements at both visits are missing, multiple imputation will be used to impute the missing values at the M45 visit. Sensitivity analyses using the tipping point analysis approach will be performed.

##### 6.3. NAC Attack Sample Size Calculations

The sample size is determined for the treatment effect as measured by the primary outcome measure of EZ-width on the OCT fovea scan. The variable for assessing the treatment effect is the AAC during the M45 follow-up. The summary statistic for the treatment effect is the difference in the AAC between the intervention arm and the control arm.

The AAC variable calculated as $\left(\sum_{j=1}^{K-1}(y_o - y_j) + \frac{1}{2}(y_0 - y_K)\right)$ is essentially a linear combination of the repeated measurements for an eye. Thus, for each study eye, the repeated measurements are reduced to a scalar statistic- the AAC. The framework of generalized estimating equation (GEE) with linear model can be used to estimate the difference between the intervention and control arms while accounting for the between-eye correlation on the AAC variable.

###### 6.3.1. Sample Size Calculation Assumptions

Assumptions on the target effect size, the standard deviation of the AAC statistic, the correlation coefficient between AACs of the two eyes within an individual, and the proportion of participants that will contribute to one study eye. PASS 2019 the GEE Test for Two Groups (Continuous Outcome) was used to do the sample size calculations.

###### 6.3.1.1. Sample Size Calculation Assumptions: Target Effect Size

The scientific objective is to determine whether the treatment with NAC 1800mg BID can delay the progression of disease by at least one third compared to the placebo control group. Considering that EZ-width is an anatomic measure and it is difficult to determine a minimum clinical effect in terms of the AAC measurement, the target effect is expressed as Cohen's D[65]. Specifically, assuming the standard deviations (SD) of the intervention and the control groups are the same, the target effect size is to detect $(\mu_i - \mu_c) \approx \frac{1}{3} * SD_c$, where $\mu_c$ is the mean of AAC in the control group, $\mu_i$ is the mean of AAC in the intervention group, and $SD_c$ is the SD of the AAC in the control group. This means that the cumulative loss of the EZ-width in the intervention group is 1/3 lower than the cumulative loss of EZ-width in the control group, where the 1/3 is relative to the standard deviation of the cumulative EZ loss in the control group, i.e. a Cohen's D[66] effect size of 1/3.

###### 6.3.1.2. Sample Size Calculation Assumptions: Between-eye Correlation on AUC

Based on data from our group (Iftikhar et al. [52]), the between-eye correlation on the changes of EZ-width is 0.76. Thus, a plausible range of between-eye correlation on AAC is 0.7-08.

###### 6.3.1.3. Sample Size Calculation Assumptions: Proportion of Participants with Only One Eye Eligible

As an inherited disease, symmetry between the right and left eyes is generally high. Thus, participants are expected to have both eyes eligible except a small proportion of participants who may have another disease process in one eye that makes the eye ineligible. We estimate the proportion of participants with bilateral enrollment to be between 85-95%. A value 86% was used in the sample size calculations.

###### 6.3.2. Sample Size Estimation

Sample size estimation was conducted using PASS 2019 the GEE Test for Two Groups (Continuous Outcome) module. Two-sided test was used. The type-I rate alpha in the calculations was taken to be 0.0494 to account for the interim analysis (Section 6.5.7) (the O'Brien-Fleming method is used for the alpha-spending).

###### 6.3.2.1. Sample Sizes Estimated Under Different Scenarios

**Table 6** presents the sample sizes estimated under different scenarios determined by the values of the necessary parameters. The results show a range of sample size required.

**Table 6 Total Sample Size Estimations for a variety of scenarios based on the plausible ranges of the parameters.** Allocation ratio is 2:1.

| **Power** | **Rho (between-eye correlation)** | **Assumed effect size (Cohen's D ranges from 25-35%, i.e. the intervention delays progression by 25-35% compared to the control)** | | |
|---|---|---|---|---|
| | | **25%** | **30%** | **35%** |
| **80%** | **0.7** | 561 | 390 | 287 |
| | **0.8** | 594 | 417 | 304 |
| **90%** | **0.7** | 751 | 522 | 383 |
| | **0.8** | 795 | 552 | 406 |

###### 6.3.2.2. Final Sample Size Determination

The final sample size determination will need to consider cost, resource availability and enrollment feasibility. Considering that the ITT effect might be lower than the biological effect (i.e. when there is perfect adherence, no protocol violation or loss to follow-up), the assumptions for the final sample size are: the target effect size is Cohen's D=29%, between-eye correlation of AAC is 0.7, alpha=0.049, 86% of participants have both eyes enrolled, and 80% power. The resulting sample size is 417. This sample size can yield 85% and 90% power if the effect size is Cohen's D of 31% and 34%, respectively.

Furthermore, a small proportion of participants might come from the same pedigree and their data might be correlated. However, it is known that the phenotypic presentations can be highly heterogeneous in patients from the same family, [66,67] thus the level of correlation between such participants is expected to be very low. Nevertheless, to account for potential enrollment from the same pedigree and loss to follow-up at M45, **the final total sample size is determined to be 438.**

To support intuitive understanding of the 29% effect size in Cohen's D, we reviewed the literature on RP natural history and interpret the effect size in terms of the difference in rates of EZ-width loss. A few retrospective chart review studies report the annual rate of EZ-width loss based on patients with longitudinal follow-up of variable lengths and are summarized in the table below (**Table 7**). The rate of EZ loss is known to be associated with baseline EZ where the larger the baseline EZ-width, the faster loss of the EZ. [42,65,68-70]

**Table 7 EZ-width rates of loss reported in the literature.**

| **RP genotype** | **Number** | **EZ-width Mean (SD)** | **Duration of follow-up** | **Mean (SD) of annual** | **Reference** |
|---|---|---|---|---|---|

| | of patients | at baseline | (years) | rate of EZ decline (µm/year) | |
|---|---|---|---|---|---|
| ***PDE6A* or *PDE6B*** | 7 | NA | 4 ± 2.6 | 91 (64) | Takahashi et.al.[71] |
| **Mixed genotypes** | 27 | 2346 (1204) | 4.8±1.4 | 76 (SD not reported) | Colombo et.al.[70] |
| **Mixed genotypes** | 106 | 1957 (1140) | 1-4 | 151 (96) | Iftikhar et al.[52] |
| ***RPGR*** | 35 | 1964 (1542) | 1-4 | 234 (189) | Tee, J. et.al [42] |
| **X-linked** | 28 | 3571 (2370) | 2 | 248 (SD not reported) | Birch et.al.[69] |
| **Mixed** | 81 | NA | 3.1 ± 1.7 | 140 (153) | Cabral, et.al.[65] |

In NAC Attack, the interval between 2 adjacent in-clinic visits is 9 months, and assuming the rate of change in EZ-width is constant, then the cumulative EZ loss, i.e. the area above the EZ-trajectory curve, *AAC* = $\left(\sum_{j=1}^{K-1}(y_0 - y_j) + \frac{1}{2}(y_0 - y_K)\right) \approx \frac{75}{8}\Delta$, where $\Delta$ is the annual rate of change. Considering NAC Attack will enroll mixed genotypes of RP patients, thus the disease progression in average would be slower than progression in patients with *RPGR* or other X-linked inheritance and faster than patients with autosomal dominant genotypes.

Based on Table 7, a plausible range for $\Delta$, is 80-200µm/year. The SD ($\Delta$) can range from 80~160 µm. Thus, a plausible value of SD of the *AAC* from a sample of mixed genotype can be taken as 1125 µm (i.e. $\frac{75}{8}\times 120$). With an effective sample size of 413 participants, the minimum detectable difference between the intervention group and the control group in terms of the mean annual rate of EZ-loss would be ~ 35µm/year.

##### 6.3.3. Power Calculations for the Secondary Outcomes

The final sample size determined by the primary outcome is 438. The minimum detectable difference (MDD) is estimated for the secondary outcomes of BCVA and MMS from MP, respectively.

The estimand for BCVA or MMS is the change from the baseline to the M45 visit.

###### 6.3.3.1. Minimum Detectable Difference between Intervention and Control for BCVA

Assumptions:

- A key parameter to calculate MDD is the SD of the change in BCVA. Based on the possible distributions of visual acuity of cross-sectional samples reported in recent literature (**Table 8**), the correlation between visual acuity measured at 2 time points about 4 years apart is 0.81 (unreported using data published in Iftikhar et al. [52]). Given SD (VA change) ≈ $SD(VA)\sqrt{(2-2*0.8)}$ , a plausible range of the SD of the change in BCVA is determined to be 7-13 letters.

**Table 8 Distributions of visual acuity in RP reported in the literature.**

| RP genotype | Number of patients | Mean (SD) of Cross-sectional Visual Acuity | Reference |
|---|---|---|---|
| ***PDE6B*** | 15 | 0.64 (0.9) LogMAR | Kim et.al.[72] |
| **USH2A and ARRP (autosomal recessive)** | 127 | 80 (NA) letters | Birch et.al[49] |
| **Mixed genotypes** | 39 | 70 (18) letters | Iftikhar et al.[52] |
| ***RPGR*** | 47 | 0.46 (0.35) LogMAR | Tee, J. et.al[73] |
| ***Mixed genotypes*** | 30 | 74 (11) letters | Campochiaro, et.al. [48] |

- The between-eye correlation coefficient on the change of BCVA is assumed to be 0.7 to 0.8.
- Two-sided test using the GEE model, alpha=0.05, 86% of the sample is bilateral enrollment. 80% power.
- Effective sample size is 413 after considering the potential small correlation between participants from the same pedigree.

Table 9 summarizes the MDDs under various scenarios based on the assumptions of key parameters. With an effective sample size of 413 and alpha=0.05, if the SD of the change in BCVA during a M45 period is 11 letters and the between-eye correlation is 0.7, then the study will have 80% power to detect a difference of 3.2 or more letters between the intervention and control arms on the change in BCVA. Alternatively speaking, to detect a difference of 3 letters in the loss of BCVA during the M45 follow-up between the intervention and control arms, the study power is 74.6%. To detect a difference of 5 letters in the loss of BCVA between the intervention and control arms, the power is 99.2%.

**Table 9 Minimum Detectable Difference (MDD) between the intervention and control (letters) on the change of BCVA.**

| Assumed SD of Change of BCVA (letters) | Rho (between-eye correlation) | MDD (letters) |
|---|---|---|
| **7** | **0.7**<br>**0.8** | 2.04<br>2.10 |
| **9** | **0.7**<br>**0.8** | 2.62<br>2.70 |
| **11** | **0.7**<br>**0.8** | 3.20<br>3.30 |
| **13** | **0.7**<br>**0.8** | 3.79<br>3.90 |

###### 6.3.3.2. Minimum Detectable Difference between Intervention and Control for Macular Mean Sensitivity

The MMS is obtained from the MAIA MP test using the "Expert" mode testing 68 loci distributed on the central 20° of the retina. It is estimated that a small proportion of participants will be unable to perform the MP test reliably based on the rate of false positive response to the light stimuli at the optic nerve. Also considering the potential enrollment of participants from the same pedigree, the effective sample size for the change in MMS during the M45 of follow-up is assumed to be 90% of the enrolled sample size i.e. ~N=393.

Assumptions:

- A key parameter to calculate MDD is the SD of the change of MMS. The MMS in decibel (dB) estimated using different MP instruments (e.g. MAIA vs. Nidek MP-1 or MP-3 microperimeters) are not numerically the same, thus only the distribution of MMS in the FIGHT-RP study (which used MAIA and the 68-loci test mode) was referenced.
  The Mean/SD of MMS of FIGHT-RP baseline was 8.4/6.7 dB of 29 patients (57 study eyes). Assuming the correlation of MMS between 2 time points on the same eye is in the range of $\rho_{y_0, y_{m45}} = 0.6 \text{ to } 0.8$, then
  $\text{SD(Change of MMS)} = \sqrt{(2 - 2 * \rho_{y_0, y_{m45}})} * \text{SD(MMS)}$ is in the range of 4.24 to 5.99 dB.
- The between-eye correlation coefficient on the change of MMS is assumed to be 0.7 to 0.8.
- Two-sided test using the GEE model, alpha=0.05, 86% of the sample is bilateral enrollment. 80% power.
- Effective sample size is 393 after considering the potential small correlation between participants from the same pedigree and that some participants will be unable to perform MP test reliably.

**Table 10** summarizes the MDDs under various scenarios based on the assumptions of key parameters. With an effective sample size of 393 and alpha=0.05, if the SD of the change of MMS during a M45 period is 5.5dB and the between-eye correlation is 0.7, then the study will have 80% power to detect a difference of 1.64 or more dBs between the intervention and control arms on the change of MMS. Alternatively, to detect a difference of 1dB in the loss of MMS during the M45 of follow-up between the intervention and control arms, the study power is 40%. To detect a difference of 2dB on the loss of MMS between intervention and control arms, the power is 93%.

**Table 10 Minimum Detectable Difference (MDD) between the intervention and control (dB) on the change of MMS.**

| Assumed SD of Change of MMS (dB) | Rho (between-eye correlation) | MDD (dB) |
|---|---|---|
| **4** | **0.7** | 1.20 |
| | **0.8** | 1.23 |
| **4.5** | **0.7** | 1.34 |

| | | |
|---|---|---|
| | **0.8** | 1.38 |
| **5** | **0.7** | 1.49 |
| | **0.8** | 1.54 |
| **5.5** | **0.7** | 1.64 |
| | **0.8** | 1.69 |
| **6** | **0.7** | 1.79 |
| | **0.8** | 1.84 |

#### 6.4. Plan for Sample Size Re-estimation

Sample size calculations are always based on assumptions of relevant parameters. The parameters needed for NAC Attack sample size determination are listed in Section 6.3.1) and the assumptions were made based on relevant information from the literature. How well these assumptions apply to the NAC Attack study populations is unknown as a priori. To ensure enough study power regarding the primary outcome measure, during the enrollment period, the study sample size will be re-estimated based on refined parameters estimated using 1) the baseline data of available NAC Attack study participants, 2) longitudinal data from the extension study of our phase-1 FIGHT RP study (PI: Peter Campochiaro), and 3) longitudinal data from the natural history study of RP currently ongoing at the Wilmer Eye Institute (PI: Peter Campochiaro).

The sample size re-estimation will be based on masked data only. This will ensure the study type-I error is not inflated. The re-estimation also needs to be conducted relatively early during the enrollment period, allowing time to arrange necessary logistics before closure of the originally planned enrollment period, should study sample size be increased. The specific plan is described below.

Once 100 participants are enrolled and randomized, their baseline data and data from the extension study of our phase-1 FIGHT RP and our natural history study will be used to refine the following parameters:

- The proportion of participants with one eye enrolled. This parameter and its 95% confidence interval (CI) will be estimated using the 100 NAC Attack participants. The upper limit of the 95%CI will be used for subsequent sample size estimation. Using PASS 2019 Confidence Intervals for One Proportion procedure and Exact (Clopper-Pearson) method, an available sample of 100 will allow over 99% confidence to estimate the upper limit to be between 92-96% if the point estimate is 84%-88% and the width of the CI is 20%.

- The between-eye correlation of the AAC of EZ-width. Because the AAC requires data of the longitudinal follow-ups, this parameter cannot be directly estimated when only baseline data are available. But we expect the between-eye correlation of the AAC to be close to and slightly smaller than the between-eye correlation of cross-sectional EZ-measurements: using a dataset of EZ-width from a previous retrospective chart review of RP patients at Wilmer (unreported using data published in Iftikhar et al.[74]), the cross-sectional between-eye correlation $\rho_0$ was about 0.90 and the between-eye correlation of the change of EZ-width over 3-6 years was about 0.79, Thus in the sample size reestimation, the range of the lower limit of the 95%CI and the point estimate of the between-eye correlation of cross-sectional EZ-measurements using the sample of 100 participants will be considered as a plausible range for the between-eye correlation of the AAC.

- At the time of sample size re-estimation, the M4.5 follow-up may be available for a small subset of participants. The rate of missed visit estimated from this subset may provide some insight on risk of missing data in the study. Additionally, the follow-up rate in the ongoing extension study of the FIGHT RP study participants will be considered for the assumption of rate of missing data in the sample size re-estimation. Approximately 27 FIGHT RP participants consented to participate the extension study where they have been receiving NAC 1800mg/bid since 2020 and will have been followed for ~24-30 months at the time of the sample size re-estimation.

- As summarized in Table 7, the mean and SD of the baseline EZ-width and the distribution of inheritance mode of the 100 enrolled participants can be informative for projecting the rate of EZ-width loss. Such knowledge together with information from the longitudinal data from the FIGHT RP extension study and the natural history study can inform the absolute effect size in loss of EZ-width and how it is related to the target effect size parameter of Cohen's D statistic for AAC. The natural history study at Wilmer was designed in 2020 to facilitate the design of NAC Attack by generating data to estimate parameters necessary for sample size calculation. The eligibility criteria of the natural history study were similar to NAC Attack criteria, allowing the preparation of a pool of potentially eligible candidates for NAC Attack screening. Enrollment of the natural history study started at the end of 2020. By the time NAC Attack starts enrollment, we expect to have ~20 participants of the natural history study each with a follow-up of 9-18 months. The OCT imaging protocol of the natural history study is similar to that of NAC Attack, and NAC Attack OCT Reading Center will provide grading to the natural history study images to generate the EZ-width measurements. The longitudinal data on EZ-width from the natural history and FIGHT RP extension studies may be used to infer the mean/SD of AAC like statistic as compared to the mean/SD of cross-sectional baseline EZ measurement, and thus may inform the specification of target effect size in the sample size re-estimation for NAC Attack. These longitudinal data will also provide information on the between-eye correlation of the change of EZ-width which can inform the between-eye correlation of the AAC.

The re-estimated sample size table under refined assumptions and 80-90% power and the rate of enrollment, will be presented to the DSMC for the decision regarding the final study sample size. Additionally, the correlation coefficients between each pair of the outcome variables will be estimated using data of the 100 participants. The disjunctive power of the hypothesis testing of the primary and secondary outcome measures using Hochberg method to adjust for multiplicity will be estimated following the methods in Section 6.5.3.2 and presented to the DSMC.

#### 6.5. Data Analysis Plan

##### 6.5.1. Analysis Plan for the Primary Endpoint

###### 6.5.1.1. Principles for Primary Analysis

The estimand for the primary outcome measure of EZ-width is the cumulative loss of EZ-width over the 45 months of follow-up, i.e. the AAC of the EZ-width trajectory (See Section 6.1.2.3). The EZ-width will be assessed at Baseline, M9, M18, M27, M36 and M45, i.e. every 9-months (the actual length of an interval for a participant may not be numerically exactly 9 months because of the allowable scheduling window but t=0, 9, 18, 27,36, 45 months will be used). If M45 visit is missed, the M40.5 OCT will be graded and used to replace EZ-width measurement at M45.

The primary analysis will follow the Intent-to-treat principle and use the Full-Analysis-Set including all randomized participants. Construction of the Full-Analysis-Set handles missing data (if the EZ-width measurement for the last study visit at M45 is unavailable) and intercurrent events and is described in detail in Section 6.1.5. Briefly, Rubin's multiple imputation will be used to impute the missing M45 measurement and draw inference. Missingness may occur because of loss to follow-up, early withdrawal or pregnancy event during the study follow-up. The intercurrent event of incompliance of the control group participants taking a supplement formulation of NAC will be identified from plasma NAC measurements. Such intercurrent even will be handled via the "Hypothetical strategy" where data for the incompliant control group participants will be imputed using multiple imputations (Sections 6.1.3 and 6.1.5.1).

Sensitivity analysis will be conducted to examine the robustness of the primary analysis results. The sensitivity analysis will be conducted using the tipping point analysis based on multiple imputations.[75,76] Details of the sensitivity analysis are described in Section 6.1.5.2).

AAC is essentially an eye-level summary measure of the repeated measurements over time. Most participants will have both eyes enrolled as study eyes, and thus analysis needs to account for the clustered data nature (the cluster size is 2 for participants with bilateral eyes enrollment and is 1 for participants with unilateral eye enrollment). The statistical modeling framework will be linear regression with GEE to account for the between-eye correlation. SAS PROC GENMOD will be used for the analysis. The "Compound Symmetry" option will be used for modeling the working correlation matrix (dimension is 2x2) and the empirical standard error estimates will be used. Wald-test will be used for hypothesis testing.

To improve precision and efficiency and following the FDA's "Adjusting for Covariates in Randomized Clinical Trials for Drugs and Biologics with Continuous Outcomes Guidance for Industry" (April 2019, available at https://www.fda.gov/regulatory-information/search-fda-guidance-documents/adjusting-covariates-randomized-clinical-trials-drugs-and-biologics-continuous-outcomes-guidance), the primary analysis model will adjust for baseline covariates that introduce imbalance by chance between the groups and prognostic factors known to be strongly associated with disease progression. Because the study does not use an equal allocation, the baseline covariates that have chance imbalance must be determined after study unmasking. Person level baseline covariates (e.g. age, sex, inheritance mode) will be compared between the two groups using two sample t-test or $\chi^2$ test. For eye-level baseline covariates, GEE models (linear model for continuous variable and logistic regression for categorical variable) will be used to compare between the groups. Any baseline covariate having a p-value $\leq 0.1$ for comparing the intervention and control groups will be included in the primary analysis model. In particular, the baseline covariates to be compared between the 2 groups will include the baseline BCVA but not baseline MMS because there may be a small proportion of participants who may be unable to conduct the MP test reliably thus resulting missing MMS values. Additionally, the variable inheritance mode, categorized into 3 groups (group 1: autosomal dominant; group 2: X-linked recessive, group 3: autosomal recessive, simplex, or indeterminate), is known to strongly predict disease progression (if there are too few X-linked participants, this group may be combined with group 3). The baseline EZ-width is also known to be strongly associated with the rate of subsequent loss of EZ-width. Thus baseline EZ-width and inheritance mode will also be included in the primary analysis model to improve precision and efficiency of the estimation.[76]

Whether the primary analysis will adjust for the effect of clinical site will be determined by findings from simulations to be conducted by the Coordinating Center statisticians during start-up and recruitment phases. In the case of independent data, adjusting center effect using a random effects model with a main random center effect could gain efficiency compared to unadjusted analysis.[77,78] However, it was noted that in practice the gain of efficiency is low.[78] We have correlated eye-level data and the performance of a mixed effects model including a random effect for center and also accounting for the between-eye correlation has not been reported in the Statistics literature and this will be studied during start-up and recruitment phases.

###### 6.5.1.2. Supplementary Analysis

Supplementary analysis will be conducted by taking into account of medication adherence and control group participants incompliance. The Per-Protocol Set[59] (PPS) will be used in such analysis. As noted in the ICH E9 R(1) Addendum, the PPS based results may be subject to bias, for reasons such as that medication adherence pattern is different between the treatment arms or that control group participants of certain characteristics that are related to disease progression may be more likely to self-taking a supplement formulation of NAC. If the results from the primary and per-protocol analyses differ substantially, further exploratory analyses will be performed to evaluate the factors that have contributed to the differences.

To estimate the “hypothetical strategy” estimand which provides an estimate of the biological effect of the treatment and account for the intercurrent event of suboptimal adherence to study medication dosing or frequency (Section 6.1.3 intercurrent event), additional analysis with the PPS dataset will apply causal inference methods such as propensity score based matching or inverse probability weighting methods. [79]

###### 6.5.2. Analysis Plan for the Secondary Outcomes

NAC Attack has 2 secondary outcome measures: BCVA and MMS from MAIA MP (Section 6.2). These 2 measures are visual function outcomes. The variables for the secondary outcomes are the change in BCVA and change in MMS from baseline to the M45 visit.

###### 6.5.2.1. Analysis Plan for the Secondary Outcome of BCVA

The principles of analysis for the primary outcome measure (Section 6.5.1) apply to the analyses for each of the secondary outcome measures. Briefly, for the change in BCVA between baseline and M45, it is an eye level measure. Thus, linear model with GEE will be used, and the primary analysis for this secondary endpoint will use the Full-Analysis-Set to retain randomization. Missing data and the intercurrent event of control group participants taking NAC supplement formulation will be handled via multiple imputations. The primary analysis will adjust for baseline covariates that show imbalance between the intervention and control groups. In particular, the baseline EZ-width, baseline BCVA itself, and inheritance mode will be considered when identifying baseline covariates that have imbalance between the 2 groups. Baseline MMS will not be included considering that a small portion of participants may not be able to conduct the MP test reliably and that MMS is highly correlated with EZ-width (unpublished from the FIGHT RP data).

Sensitivity analyses will use the tipping point analysis (Section 6.1.5.2). Supplementary analysis will include analysis based on the Per-Protocol Set.

###### 6.5.2.2. Analysis Plan for the Secondary Outcome of MMS

For the secondary endpoint of change in MMS between baseline and M45, the principles are similar to those for the change in BCVA. The caveat is that the target population for MMS is the participants who can reliably complete the MP tests at both the baseline and M45. This target population is essentially a subgroup of all participants in the trial. We anticipate that only a small proportion of participants (<5%) cannot perform the MP test reliably. For these participants, no more MP test will be administered during follow-up. Among participants who can complete the micrometry test reliably at baseline, we expect it would be rare that these participants will be unable to do the test reliably during follow-up.

The change in MMS is an eye level measure. Thus, linear model with GEE will be used and the primary analysis for this secondary endpoint will use the Full-Analysis-Set for the target population. Missing data and the intercurrent event of control group participants self-seeking NAC supplement formulation will be handled via multiple imputations. The primary analysis will adjust for baseline covariates that show imbalance between the intervention and control groups. In particular, the baseline EZ-width, baseline BCVA itself, baseline MMS, and inheritance mode will be considered when identifying baseline covariates that have imbalance between the 2 groups. (unpublished from the FIGHT RP data).

Sensitivity analysis will use the tipping point analysis. Supplementary analysis will include analysis based on the Per-Protocol Set among the target population for the secondary endpoint of change in MMS between baseline and M45.

###### 6.5.3. Hypothesis Testing with the Primary and Secondary Outcomes , Multiplicity and Disjunctive Power

The primary outcome measure is EZ-width which is an anatomic parameter reflecting the integrity of photoreceptors. It is selected as the primary outcome measure for sample size considerations and because of the limitations of visual function outcomes in RP (see Section 6.1.2). The secondary outcomes of BCVA and MMS are both visual function outcomes and are clinically important. EZ-width has been shown to be correlated with visual functions in RP.[42,52,53,73] It has also been shown that there can be sensitivity response outside of the reflective band of the EZ.[80,81] Therefore, the 3 endpoints are correlated but provide assessments of different aspect of the disease process. Thus, the hypotheses testing of these 3 endpoints are deemed of equal scientific importance and will not be tested sequentially following a pre-specified order (normally determined by their levels of importance).

Following the FDA "Multiple Endpoints in Clinical Trials Clinical Trials Guidance for Industry" (Draft Guidance, January 2017, available at [https://www.fda.gov/regulatory-information/search-fda-guidance-documents/multiple-endpoints-clinical-trials-guidance-industry](https://www.fda.gov/regulatory-information/search-fda-guidance-documents/multiple-endpoints-clinical-trials-guidance-industry)), multiplicity needs to be accounted for to allow findings of significant treatment effects at the individual endpoint level while controlling the Type I error rate ($\alpha$) of the trial. The Single-step procedure of Bonferroni method is convenient but is conservative with multiple endpoints that are positively correlated.

Positive correlations are expected for the tests statistics of the 3 endpoints for NAC Attack given the known structure-function relationships of photoreceptor integrity and visual functions and the known positive correlation between the 2 visual function outcomes [42,53 73] (in the FIGHT-RP baseline data, the correlation coefficients between EZ-width and BCVA, between EZ-width and MMS, and between BCVA and MMS were 0.38, 0.73, and 0.41, respectively). When 2 endpoints have a positive correlation, the corresponding test statistics will be also positively correlated.[82] Therefore, assumptions are met for appropriately using stepwise semiparametric procedures with a data-driven hypothesis ordering. Specifically, the step-up Hochberg procedure will be used. The Hochberg procedure is uniformly more powerful than the nonparametric step-down Holm procedure. The procedure controls the Familywise Error Rate in the strong sense under the assumption that the test statistics associated with the null hypotheses of the individual endpoints follow a multivariate normal distribution with positive pairwise correlations.[82] This assumption is reasonable here because the test statistics for the individual endpoints in NAC Attack all come from a z-statistic based on linear models with GEE, and they are positively correlated based on the biological or clinical nature of the endpoints. Moreover, properties of the Hochberg procedure hold with common imputation methods for handling missing data.[82]

###### 6.5.3.1. Applying Hochberg Procedure in Hypothesis Testing of the Primary and Secondary Endpoints in NAC Attack

The Null hypotheses are denoted as:

| | | |
|---|---|---|
| $H_1$: No difference in the cumulative loss of EZ-width during the 45-months follow-up between the intervention and placebo control groups. | $H_2$: No difference in the change in BCVA from the baseline to the 45-months visit between the intervention and placebo control groups. | $H_3$: No difference in the change in MMS from the baseline to the 45-months visit between the intervention and placebo control groups. |

Let $p_1$, $p_2$ $p_3$ denote the p-values from the individual primary analysis models for the outcomes on EZ-width, BCVA and MMS, respectively. After ordering these 3 p-values, let $p_{(1)}$, $p_{(2)}$, $p_{(3)}$ denote the ordered p-values, i.e. $p_{(1)} \le p_{(2)} \le p_{(3)}$; and $H_{(1)}, H_{(2)}, H_{(3)}$ the corresponding ordered null hypotheses.

Based on the O'Brien-Fleming alpha-spending for the final analysis, the $\alpha = 0.0494$ when there is one interim analysis (See Section 6.5.7). The Hochberg procedure determines when to reject $H_1$, $H_2$ and/or $H_3$ based on the following algorithm.

- If $p_{(3)} \le \alpha$, we reject all null hypotheses and claim significant differences between the intervention and control on all endpoints;

  Otherwise, continue to $p_{(2)}$.

  - If $p_{(2)} \le \frac{\alpha}{2}$, we reject $H_{(1)}$ and $H_{(2)}$ but cannot reject $H_{(3)}$, i.e. there are significant differences in the endpoints corresponding to $H_{(1)}$ and $H_{(2)}$ but no significant difference in the endpoint corresponding to $H_{(3)}$;

    Otherwise, continue to $p_{(1)}$.

  - If $p_{(1)} \le \frac{\alpha}{3}$, we reject $H_{(1)}$ only, i.e. there is significant difference in the endpoint corresponding to $H_{(1)}$ but no significant difference in either of the endpoints corresponding to $H_{(2)}$ and $H_{(3)}$;
    Otherwise, we cannot reject any null hypothesis, i.e. no significant difference between the intervention and control in any of the endpoints.

The Hochberg procedure evaluates efficacy on the primary and secondary endpoints while controlling the familywise error rate. The simultaneous confidence intervals from multistep procedures however are not readily available in the statistical literature. As pointed out by the FDA "Multiple Endpoints in Clinical Trials Clinical Trials Guidance for Industry" (Draft Guidance), "Multistep procedures are generally more efficient in that they better preserve the power of the tests, but do not readily provide adjusted confidence intervals."

###### 6.5.3.2. Disjunctive Power of the Hypothesis Testing of the Primary and Secondary Endpoints

Disjunctive power is the power to detect at least one effect from all the outcome measures in the hypothesis testing.[83] The study sample size was determined based on power and sample size calculations of the primary outcome measure and considering resources and feasibility. Here we conducted simulations to assess the disjunctive power of the study hypothesis testing plan given the effective study sample size of 417 (Section 6.3.2.2)

**Simulation Methods**

The parameters used for data generation referenced parameters used in the final sample size calculation and are listed below. Multivariate normally distributed data were generated where each subject if both eyes were eligible would have two measurements (left and right eyes) for each of the 3 outcomes measures.

Parameter assumptions:

- Sample size N=417
- 86% of the subjects had both eyes eligible.
- Group allocation ratio =2:1
- Alpha=0.0494
- Treatment effect for the primary outcome measure in Cohen'D=30%, i.e. beta1=0.3.
- Inter-eye correlation coefficient for the same outcome variable $\rho_{lr}$=0.7
- Correlation coefficient between any pair of outcomes of the left (right) eyes $\rho$ ranged between 0.2 and 0.8.
- The inter-eye and between pair of outcomes= $\rho * \rho_{lr}$
- Treatment effect (in Cohen'D) for each of the secondary outcome measures ranged from 10% to 50%, i.e. beta2= 0.1~0.5, beta3=0.1~0.5.

Data Generation

Under each parameter setting, B=1000 simulated datasets were generated in R 3.6.3. Each simulated dataset was generated as multivariate normal with mean $\boldsymbol{\mu} = (beta_1, beta_1, beta_2, beta_2, beta_3, beta_3) \cdot x$ where $x = 0$ for a subject in the control group and $x = 1$ for a subject in the treatment group. The variance-covariance matrix had diagonals as 1 and off-diagonals as the correlation coefficients (inter-eye correlation for the same outcome variable, between outcomes correlation of the left/right eyes, or the inter-eye and between- outcomes correlations) as appropriate.

Disjunctive Power Estimation

With each simulated data, linear models with generalize estimating equation were used to estimate the unadjusted p-values for the treatment effects of the 3 outcome variables.

The R function “geeglm” with exchangeable correlation structure was used for the estimation of treatment effects.

Hochberg method was subsequently used to obtain the multiplicity adjusted p-value using the R function “p.adjust”. With the B=1000 simulations, Disjunctive power was estimated as the proportion of simulations where at least one outcome variable had adjusted p-value<=alpha.

**Disjunctive Power Results**
Figure 6 shows the disjunctive power estimated from the simulation. Each plot is for a given effect size of one secondary outcome (beta2=0.1 to 0.5). Within each plot, each line is for a given effect size of the other secondary outcome (beta3=0.1 to 0.5) and it shows the disjunctive power as a function of $\rho$ the between-outcome correlation.

The disjunctive power as expected is a function of the effect sizes of the secondary outcomes. When both secondary outcomes had a small treatment effect (Cohen’s Ds=10%), the disjunctive power is lower than 80% but can be controlled at ~70%. When

*Figure 7 Disjunctive power of the planned hypothesis testing of the primary and secondary outcomes using Hochberg procedure.*

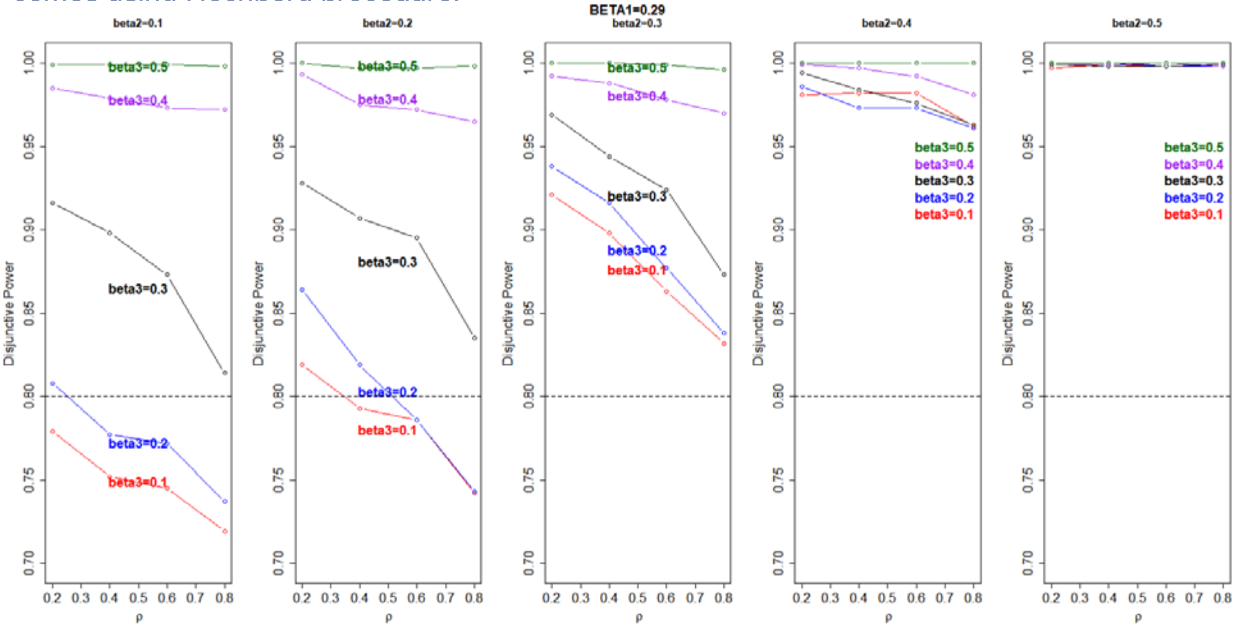

at least one of the secondary outcomes had a treatment effect of 20%, the disjunctive power is often greater than 80% except when the outcome variables were highly correlated with $\rho \geq 0.6$, the power was about 75-80%. When the secondary outcomes had larger treatment effects (≥30%), the disjunctive power is often greater than 85%.

In practice, the level of treatment effect is unknown until the study is completed, and thus the exact disjunctive power can only be estimated for a range of treatment effect size. The assumed range (Cohen’s D 10%-50%) for the effect size of the secondary outcomes is biologically plausible and can be interpreted as that the loss in MMS or BCVA was 10% to 50% less in the NAC group compared to the control group.

###### 6.5.4. Pre-planned Subgroup Analysis

The mechanism by which NAC treatment is hypothesized to slow progression in RP is to preserve cone survival and function by reducing oxidative stress in the outer retina. Rods die due to genetic mutations in RP. Death of rods reduces oxygen consumption, leading to elevated tissue-level of oxygen in the outer retina. The oxidative stress then contributes to cone degeneration and loss of function. This mechanism does not depend on the specific genetic variant responsible for rod cell death, and thus the treatment has the potential to benefit patients with a clinical diagnosis of RP including those in whom no pathogenic variant has been identified.

NAC Attack does not restrict to enrollment of patients with known RP related pathogenic variants and the patient population thus can be genetically heterogeneous. However, while genetic variants that cause RP differentially affect rods much more than cones, the magnitude of the differential effect can differ by different genetic variants. For example, mutations in genes that are expressed in rods, but not cones, have no direct deleterious effect on cones. In contrast, some mutations are in genes that are expressed in both rods and cones. They cause greater damage to rods but may also cause milder damage in cones. In such patients, mutation-induced damage contributes to cone death in combination with oxidative damage resulted from rod death. Therefore, it is reasonable to hypothesize that these patients may experience less benefit from NAC compared with patients with a mutation in a rod-specific gene. Thus, we hypothesize that beneficial effect of NAC may vary depending upon genetic variant causing RP.

Certain covariates such as smoking and iron supplementation which may promote oxidative stress or interfere with the oxidative process may also interfere with the antioxidant effects of NAC and hence reduce therapeutic effect. [84] Subgroup analyses therefore will provide a better understanding of patient characteristics and subpopulations regarding the response to NAC and antioxidants.

The subgroup analysis will adhere to the relevant scientific and statistical principles.[85] Based on potential regulatory requirement and biological plausibility, we prospectively define exploratory subgroup analysis. [85] The treatment effects by the following baseline covariates will be examined to assess the consistency of treatment effects.

- Sex.
- Race.
- Inheritance mode, categorized as (1) autosomal dominant, (2) X-linked recessive, or (3) autosomal recessive, simplex, or indeterminate.
- Age of symptom onset
- Smoking status
- Oral mineral supplement consumption, categorized as yes vs. no.
- Genotype, defined by multiple methods as described in Section 6.6.
- The binary classifier of whether a participant has RP-causing pathogenic variant

identified from the whole genome (exome) -sequencing (WES). The WES data will be obtained from a single genetic sequencing vendor (see Section 10.7.4) and the genetic data will be analyzed by Dr. Robert Hufnagel's lab (10.7.4). Participants will be categorized as whether there is any RP-causing pathogenic variant identified from WES.

Each subgroup defining covariate will be examined individually. Approaches including forest plots, test of the treatment and covariate interaction effect and interaction-to-overall effects ratio.[85,86]

###### 6.5.5. Analysis Plan for the Exploratory Outcomes

Exploratory objectives play a general supportive role and can be presented in product label for descriptive purposes (i.e. no statements supported by statistical inference), thus they bear no multiplicity adjustment consideration and p-values and confidence interval may not be used.[87] Their analyses focus therefore will be descriptive. The following exploratory outcomes are continuous and will be summarized by mean, standard deviation, range, median and interquartile range by treatment group.

- Change from baseline mean macular sensitivity measured by MP at M4.5, M9, M18, M27, M36, and M40.5.
- Change from baseline in BCVA measured ETDRS protocol at M4.5, M9, M18, M27, M36, and M40.5.
- Change from baseline in cone density, spacing, mosaic regularity, and reflectivity measured by (AOSLO) at M9, M27, and M45 at selected sites.
- Change between baseline and M45 in patient reported outcome assessed using NEI VFQ-25.

The following exploratory outcomes are categorical and will be summarized by proportions by treatment group.

- Proportion of eyes with ≥ 5 loci improved from baseline by ≥ 6 decibels (dB) at M4.5, M9, M18, M27, M36, M40.5 and M45 by MP
- Proportion of eyes with ≥ 5 loci decreased from baseline by ≥ 6 decibels (dB) at M4.5, M9, M18, M27, M36, M40.5 and M45 by MP

###### 6.5.6. Safety Analysis Plan

AEs will be categorized as systemic versus ocular. The events will be tabulated by treatment group. The frequency of the event occurring at least once will be calculated. Rates of AEs will be compared between treatment groups using Barnard's unconditional exact test. The total number of AEs per participant may also be compared using Poisson distribution based models between treatment groups.

Based on observations in the FIGHT-RP study, most drug-related AEs involved the gastrointestinal (GI) system. The effervescent tablets containing contain a sodium load and therefore have the potential to exacerbate hypertension in susceptible patients. Thus, NAC Systemic AE will focus on:

- GI symptoms such as vomiting, stomach aches, constipation, nausea and diarrhea, burping and hiccups, bloating and abdominal discomfort.

- Exacerbation of hypertension
- Other systemic symptoms such as headache and fatigue

Ocular AEs were not observed in FIGHT RP study. Any ocular AE will be recorded, and the frequency of all ocular AE will be calculated and compared between the groups. The following serious systemic AEs will be assessed:

- Primary:
  - Death
  - Serious SAE (at least one)
  - Hospitalization (at least one)
  - Cardiovascular/cerebrovascular events according to Antiplatelet Trialists' Collaboration (excerpted from BMJ Jan 8, 1994):
    - Non-fatal myocardial infarction
    - Non-fatal stroke (counted only if symptoms lasted at least 24 hours)
    - Death attributed to cardiac, cerebral, hemorrhagic, embolic, other vascular (does not need to be ischemic in origin), or unknown cause
    - At least one event (non-fatal myocardial infarction, non-fatal stroke, or death attributed to potential vascular or unknown cause)
      APTC Notes: Transient ischemic attacks, angina, and possible myocardial infarction or stroke are not counted. 'Nonfatal' myocardial infarction or stroke required that the participant was alive at the end of the study. If not, only the death is counted.
- Secondary:
  - Frequency of at least one event per participant in each Medical Dictionary for Regulatory Activities (MedDRA) system organ class

An additional tabulation will be made for AEs possibly related to study treatment.

###### 6.5.7. Interim Analysis

Although consideration may be given to stop the study early because of an apparent beneficial treatment effect, early termination for efficacy will be considered with caution because of the degree of uncertainty with regard to the long-term benefit of treatment even if a short-term benefit seems apparent prior to the completion of the study.

NAC Attack will have one interim analysis and the final analysis. The interim analysis will be conducted after all participants complete their 27-months follow-up and will use all data up to the 27-months visit. The decision of an interim analysis at 27-months is based on 2 considerations: first, RP progression is slow in the disease natural history (patients lose vision over decades). Thus it is thought that at least 2 years would be needed to have a likelihood to observe some treatment effect that can suggest long-term efficacy of NAC on delaying disease progression. Second, the study treatment NAC has known a safety profile for its FDA approved usages (such as treating acetaminophen overdose) which is short-term use. There are no data on the long-term safety of NAC. Therefore, it is important to have long-term use safety data.

The focus of the interim analysis is to evaluate safety and efficacy. The interim analysis will not seek to re-estimate sample size or change study population because of the logistical complexity in an international multicenter trial. The interim analysis will also not test for futility because given the great unmet need in RP management and the slow progressing nature of the disease, any type II error will be consequential as it will be undesirable to discard a promising treatment with good safety profile by erroneously stopping the trial early. [88]

The efficacy evaluation at the interim analysis will use the O'Brien-Fleming alpha spending rule[88] and the decision rule will be based only on the efficacy of the primary objective of significant reduction in the cumulative loss of the EZ-width (i.e. AAC over the 27-months) in the intervention group compared to the placebo group. The efficacy decision will not be based on the secondary endpoints related to BCVA or MMS. This means that the trial will only stop early if efficacy is demonstrated on the primary endpoint related to EZ-width, irrespective of the effects of the treatment on the secondary endpoints related to BCVA and MMS. The rationale is that in the FIGHT-RP phase-1 trial, a small improvement in BCVA and MMS was observed during the 6-months NAC treatment period. However, visual functions cannot improve unlimitedly in a linear fashion and will reach a plateau. There is no knowledge on the potential trajectory of BCVA or MMS after they reach the plateau while under continuing NAC treatment. Should NAC be proven efficacious for delaying progression in RP, it will be a life-long medication and will have economic implications. Therefore, it is both clinically and economically important to understand whether the effects of NAC on visual functions can be maintained over a long and sustained period of time. On the other hand, if efficacy is observed for the anatomic measure of EZ-width, it means that the treatment promotes survival of cone photoreceptors. Progressive death of rod and cone photoreceptors is the etiological pathway leading to vision loss in RP. Thus, if an effect on preserving cone photoreceptors is observed, slower loss of visual functions is expected and can be maintained.

Therefore, in the DSMC charter, the interim efficacy analysis is based on the hypothesis testing of the primary endpoint of the cumulative loss of the EZ-width over 27-months (AAC) is significantly different between the intervention and control group. The significance level uses O'Brian-Fleming boundary of 0.0054.[87]

If efficacy is determined, the randomized trial will stop but NAC Attack will continue to follow all participants until all participants complete their M45 visits. Each control group participant will be provided with NAC at no cost for another 12 months. As soon as interim efficacy is determined, the study Drug Manufacturer and Drug Labeling and Distribution Center will be notified to prepare for the batch allowing for the control group participants to receive NAC for 12 months.

###### 6.5.8. Additional Analyses

The following will be tabulated according to treatment group:

- Baseline demographic and clinical characteristics
- Visit completion rate at each visit (phone call with Coordinator, tele-visit with Ophthalmologist, or in-clinic visit), at each in-clinic visit.

- Plasma NAC level summary at each in-clinic visit for the intervention group; the percent of plasma NAC detected for the placebo control group.
- Participant level in-clinic visit completion rate over the study period.
- Study medication adherence assessed by medication reconciliation via pill counting at each in-clinic visit.
- Participant level medication adherence assessed by pill counting summarized over the study period.
- Participant level medication adherence assessed by weekly SMS text messaging based mini survey of adherence summarized over the study period.

###### 6.5.8.1. Analysis Plan for the Pharmacokinetic Objectives

The Pharmacokinetic Objectives are:

- Measure plasma NAC levels at each visit in all participants to evaluate compliance.
- Correlate plasma NAC levels with efficacy endpoints.

For participants randomized to the intervention group, at each in-clinic visit, their plasma NAC levels will be summarized. In addition, at an individual participant level, the trajectory of NAC will be graphically examined. Mixed effects models may be used to examine whether there is any trend with within-individual fluctuations of NAC levels over study follow-up while adjusting for time since the last dose.

Linear regression models with GEE will be used to evaluate whether efficacy endpoints are associated with the level of NAC in plasma. For each of the outcome measures of EZ-width, BCVA and MMS from MAIA MP and at each follow-up visit, the change from baseline to the follow-up will be the dependent variable, and the plasma NAC measurement is the exposure variable. The baseline value of the efficacy outcome measure itself may also be included in each regression to account for the potential “regression to the mean” effect when using the change as the dependent variable.

For participant randomized to the control group, their NAC level should be 0. For each follow-up visit, the proportion of control group participants having NAC level detected will be estimated.

###### 6.5.8.2. Pharmacogenomic Objectives

The Pharmacogenomic Objectives are:

- Examine the relationship between disease causing mutations and the effect of NAC on primary, secondary, and exploratory outcome measures.
- Examine the relationship between disease causing mutations and progressive decrease in EZ width over time as assessed by area over the curve in placebo subjects.

RP is a clinical diagnosis. Over 270 genes have been reported to be associated with Mendelian forms of inherited retinal diseases (RetNet; accessed on 08/28/2020). There can also be different pathogenic variants associated with the identification of a disease-causing gene. NAC Attack enrollment does not restrict to specific genotypes of RP. The genetic profiles of the NAC Attack study participants based on whole genome (exome) sequencing findings are expected to be highly heterogeneous. Thus, before examining any genotype-phenotype relationship in the placebo participants or examining pharmacogenomics of NAC in the intervention arm, it is necessary to determine how genotypes may be defined via genomic analysis. Section 6.6 describes the genomic data analysis plan focusing on the pharmacogenomics of NAC in RP.

To examine the relationship between genetics and disease progression in EZ-width in placebo subjects, genotypes will be defined in various ways as described in Section 6.6, including by known genes the variants of which cause RP, by molecular pathways or functional groups, or by exome-wide variant association analysis on gene burden, etc. To compare disease progression by genotypes in the placebo group, mixed effects models may be used to examine the trajectories of EZ-width loss over time by genotype. The cumulative loss of EZ-width (the AAC) over the study period can be compared by genotype using ANCOVA method adjusting for baseline EZ-width and other covariates (such as inheritance mode, age of symptom onset, etc.).

###### 6.5.8.3. Additional Exploratory Analysis

NAC Attack will generate rich data allowing a greater understanding of the natural history of RP and a deep exploration of the potential treatment mechanism of antioxidant in preserving cones and visual functions in RP. Therefore, many analyses in addition to the analyses for safety and efficacy evaluations will be performed. Some examples are listed below:

- To examine the longitudinal trajectory of each outcome measure.

  The longitudinal data of each outcome measure will be plotted over time to visually examine the pattern of longitudinal change by treatment group. Regression modeling will also be conducted to explore longitudinal trajectory with each outcome measure, including:

  - Linear or non-linear mixed effects model (MEM) for the longitudinal EZ-width measurement. If a non-linear trajectory is observed, piece-wise linear MEM or growth curve models may be considered. The MEM framework can account for the between-eye correlation and longitudinal correlation from repeated measurements.[89]

  - Linear or non-linear MEM for the longitudinal MMS or BCVA measurements.

  - For the exploratory outcomes of NEI-VFQ and the outcomes based on AOSLO, a linear or a piece-wise linear MEM may be built for each of the variables.

  - For the outcomes from AOSLO on cone measurements, at each visit and for each eye, there can be multiple measurements from different regions of interest (ROIs)

- To assess cross-sectional structure-function correlations.

The MEM regression framework will be used to correlate cross-sectional structural measurements (EZ width, EZ area) with visual function outcomes (BCVA, MMS). Similarly, cone measurements from AOSLO will be correlated with EZ measurements and with visual function outcomes.

- To assess longitudinal structure-function correlations.

First, MEM regression will be used to assess whether and to what degree the change of a structural or anatomic measurement is associated with the change of a function outcome during a period.
Second, MEM regression will be used to assess whether the change in a structural or anatomic measurement during an earlier and shorter period predicts the loss of visual function during a longer time.

- To examine genotype-phenotype relationships (see Section 6.6).

#### 6.6. Genomic Data Analysis Plan

##### 6.6.1. Rationale of Studying Pharmacogenomics of NAC

NAC is a precursor of cysteine, and involved in glutathione (GSH) metabolism, which is a non-enzymatic antioxidant essential for cellular protection against oxidative stress. NAC can also act as an antioxidant itself by scavenging free radicals and/or decreasing the quantity of oxidized lipids and proteins.[90] In a recent study, biochemical basis of retinal regeneration by NAC were described to be a consequence of reducing oxidized proteins which depended on MAPK (pErk1/2) pathway. [91]

Pharmacogenomics of NAC have not been well studied. However, it has been reported that genetic polymorphism in *TOLLIP* gene may impact the NAC response in individuals with idiopathic pulmonary fibrosis ([92]). Likewise, several studies suggest that genetic polymorphisms like *glutathione-S-transferase* (GST M1 null, and GST M1 and T1 double null) isoforms and *EPHX1* (p.Tyr113His and p.His139Arg) polymorphisms, may be associated with the response to NAC treatment of acute respiratory distress syndrome (ARDS)/acute lung injury (ALI) and COPD, respectively.[93,94]

##### 6.6.2. Plans for Studying Pharmacogenomics of NAC

Over 270 genes have been reported to be associated with Mendelian forms of inherited retinal diseases (RetNet; accessed on 08/28/2020). These genes can be grouped and analyzed based on their functions (e.g. visual transduction related, cilia related), shared pathway relationships (e.g. apoptosis, autophagy), or expression in a specific cell type (e.g. photoreceptor cells specific; retinal pigment epithelium (RPE) specific). Further, genes which may be involved in NAC response can be the genes involved in drug metabolism and oxidative stress related genes.

We hypothesize that there is a genetic basis for the response to NAC treatment, which results in high responders and low responders. This could relate to NAC function and metabolism, the underlying cause of disease, or an unknown mechanism. Genetic analysis comparing high and low responders will be performed by the following methods:

1. Comparison of known SNPs related to NAC response – *GST* (M1, P1, T1 isoforms), *EPHX1* (p.Tyr113His and p.His139Arg), *TOLLIP* (rs3750920 TT genotype).
2. Comparing variant burden in causal retinal degeneration genes grouped individually and by pathway, functional group, or cell-type expression as described above.
3. Unbiased genome-wide rare and common variant association analysis approaches, including gene burden.

###### 6.6.3. Analysis Pipeline for Studying Pharmacogenomics of NAC

Briefly, the DNA samples will be sent to the Study central laboratory for genetic testing where whole genome (exome) sequencing will be performed. The raw data (e.g. FASTQ files) will be uploaded by the central laboratory on a secure web portal. The Study Genetic Analysis Lab physically located at the National Institutes of Health (NIH) will download the raw data securely onto NIH HPC drive. The raw data will be processed by an in-house NGS processing pipeline (https://github.com/Bin-Guan/NGS_genotype_calling & https://github.com/Bin-Guan/variant_prioritization). In brief, raw reads will be aligned to hg19 human reference genome by using Burrows Wheeler Alignment tool (BWA), duplicates will be marked by Picard tool and quality control analysis will be done by using MultiQC and Picard tool. Then single nucleotide variants (SNVs) and small insertions/deletions (InDels) will be called separately by GATK and/or FreeBayes and will be merged in a single variant call file (VCF). Then by using Variant Effect Predictor (VEP), InterVar, ANNOVAR and Vcfanno, many publicly available as well as some in-house annotations will be added to each variant. Some of the popular and frequently used annotations include healthy population allele frequencies (gnomAD, ESP, 1000 genomes), in-silico pathogenicity predictions (CADD, Revel, PolyPhen, SIFT), disease population databases (HGMD, ClinVar). Then by using Vcf2Db tool, the annotated variants from cases as well as controls will be consolidated in a single GEMINI database (https://gemini.readthedocs.io/en/latest/ ). Then association testing will be performed which will include Fisher's exact test (R Studio), c-alpha test (GEMINI database), and SKAT-O test (R Studio) to identify statistically significant variants associated with NAC response. This will be followed by generation of publication grade graphical visualizations for better understanding association testing. All the data including raw data, BAM and/or CRAM files, VCF files, GEMINI database and QC reports, plots, graphs, tables etc. will be stored on secure storage system which will occupy about 8-10 terabytes for the sequencing data of the NAC Attack participants.

### 7. Data Collection and Management

#### 7.1. Categories of Patient Data Collected in NAC Attack

Herein "data" broadly refer to any information about patients enrolled in NAC Attack. *The data can be categorized into three types.*

- Data collected using study forms and involving personal identifying information such as patient names.
- Data collected using study forms, not involving personal identifying information, and entered into the REDCap study database.
- Data contained in electronic files that are not managed through REDCap, such as imaging and test files, imaging gradings and lab results, etc.

#### 7.2. Clinical Site Management of Patient Data Containing Personal Identifying Information

Such data refer to study forms/logs that collect patient related information involving patient names and their emergency contacts. These include the informed consent form and emergency contact form. These forms are filed into study binders and securely stored at clinical sites. Data on these study forms do not enter into the study REDCap database and are only accessible to study team members at the clinical site where a patient is enrolled.

During the study participation, a participant may move and consent to have study visits at another clinical site or may need to receive AOSLO imaging at a nearby imaging site. Data transfer related to the participant changing site is described in Section 4.12.5.

#### 7.3. Coordinating Center Management of Patient Case Reporting Forms (CRFs) Data Through REDCap

All data entry will be performed by Study Coordinators or certified clinic staff under the supervision of Study Coordinators using paper study CRFs. Only de-identified (pseudonymized) data will be collected on these forms. Patient's Study ID and visit date are recorded on every page of all CRFs to ensure the identification of the data on the forms.

After a patient visit is completed, data on the CFRs should be entered into the study REDCap database within the same day, or when necessary, data entry to REDCap should be completed by the next business day. A scanned copy of the source CRFs should be submitted to the Coordinating Center cloud-based portal.

The Coordinating Center is responsible for designing the paper CRFs and building and maintaining the REDCap data collection interface and database corresponding to the paper CRFs.

##### 7.3.1. The REDCap Data Collection System

REDCap (Research Electronic Data Capture) https://projectredcap.org/software/ is a secure web application for building and managing online databases. NAC Attack uses REDCap managed by the Johns Hopkins Institute for Clinical and Translational Research (ICTR) which is a REDCap consortium partner. Johns Hopkins ICTR is funded by the NIH Clinical and Translational Sciences Award program and serves the entire Johns Hopkins University medical research community. The Coordinating Center will develop the REDCap project database for NAC Attack with support from the ICTR staff. ICTR also provides continuing maintenance support on the functioning of REDCap projects.

REDCap provides secure and web-based data input for users across multiple sites with authentication and data logging. Important features include audit trails for tracking data manipulation and user activity, automated export procedures for data download to common statistical packages, reporting tools and user-friendly data entry interface. REDCap also provides the capability of sending SMS text messages to survey participants through a third-party service (Twilio), and the survey responses can be integrated with the project database for the same participants. REDCap can be installed in a variety of environments for regulatory compliance with standards including HIPAA (Health Insurance Portability and Accountability Act), FDA regulation (21 CFR Part 11 Electronic Records Electronic Signatures), FISMA (Federal Information Security Management Act), and international standards.

The Johns Hopkins Institute for Clinical and Translational Research (ICTR) is a REDCap consortium partner. ICTR is funded by the NIH Clinical and Translational Sciences Award (CTSA) program and serves the entire Johns Hopkins University medical research community. The Coordinating Center will develop the REDCap project for NAC Attack with support from the ICTR staff. ICTR also provides continuing maintenance support on the functioning of REDCap projects.

Study sites will receive training and have access to a manual for REDCap data entry.

###### 7.3.2. Source CRF Documentation

NAC Attack will use paper CRFs to collect de-identified data at the Clinical Sites. These forms will be data entered into REDCap Study database developed and administered by the Coordinating Center through JHU ICTR. Data Access Groups will be used to ensure that clinics will only have access to the data at their own clinic. REDCap automatically tracks events such as record creation and data value changes. Study site staff will not be given permissions to delete their own records.

All paper CRFs should be completed by designated, trained site staff. CRFs should be reviewed and electronically signed and dated by the investigator or a designee at the time of data locks for the interim efficacy analysis and the final analysis.

At the end of the study, the site investigator will receive patient data for his or her site in a readable format that must be kept with the study records. Acknowledgement of receipt of the data is required.

The paper CRFs will be kept at the Clinical Site in the patient's binder stored in a locked file cabinet or room. The paper forms may be used during clinic monitoring visits to verify data entry accuracy.

###### 7.3.3. Data Quality Assurance

During data entry, built-in data check functions in REDCap form can automatically check for valid codes, legitimate ranges and dates and remind data entry staff if invalid data values are entered. The source forms should also be scanned into a PDF copy and submitted to the Coordinating Center portal.

Form revisions and additions will be accommodated by having Coordinating Center data management staff modify or create the appropriate data definition. Each study REDCap form will build-in a field to capture the version changes of the form over time.

Central laboratory data and central reading centers data will be submitted directly to the Coordinating Center, using the Coordinating Center's standard procedures to securely handle and process the electronic transfer of these data. The paper CRFs will be kept at the Clinical Site in the patient's binder stored in a locked file cabinet or room. The paper forms will be used during clinical site monitoring visits to verify data collection accuracy.

Specific quality assurance features related to data management are:

- Standard data collection forms and procedures;
- Training and certification in data collection and data entry;
- Central concurrent processing of data by the REDCap data collection system to detect problems early and provide immediate feedback to the clinical sites;
- Data checking by the REDCap data collection system for missing, invalid, and suspect responses;
- Data checking by Coordinating Center Research Assistants to compare REDCap data entry to scanned source CRFs;
- Data monitoring within the Coordinating Center of critical forms to compare the REDCap data entry to scanned source CRFs. If there is any data entry error identified and such error was not identified from the above data checking step by the Research Assistant, all forms that were checked by the Research Assistants will go through the data monitoring process in order to identify data entry errors potentially missed by the Research Assistants.
- Regular reporting on performance of all clinical sites , including parameters on enrollment, screen failure rate, protocol deviations;
- Checking a random 10% sample of original data collection forms against medical records during site monitoring visits.
- Explicit instructions about new/revised questions with each distribution of new data collection form masters and clear instructions to discard all previous versions and copies.

##### 7.4. Management of Patient Data Not Collected Through CRF

This type of patient data can be further summarized by where the data are generated, including:

- Data (e.g. electronic files such as imaging files) from the clinical sites
- Data (e.g. image grading) from the OCT Reading Center
- Data from the AOSLO Reading Center (for participants enrolled in the AOSLO sub-study)
- Data from the Study central laboratories, including:
  - Data from the Study central lab of plasma NAC testing
  - Data from the study central lab for genetic testing
  - Data from the genetic data analysis lab headed by Co-investigator Robert Hufnagel.

The sections below summarize the management of these data. All participant data will be identified by Study ID. Personal identifying information such as patient name, date of birth, phone number or address will not be present in these data. All data transfers will be through HIPAA compliant portals for secure file sharing and storage (Section 7.4.1).

###### 7.4.1. Coordinating Center Cloud-based Data Files Management System

The Coordinating Center will use JHOneDrive to allow clinics to upload files to a HIPAA-compliant cloud-based server using a web-based interface.

JHOneDrive is administered by the Johns Hopkins University (JHU) Information Technology (IT) department. It is the individual cloud storage component of the Office 365 product suite that allows users to store and share documents and files from any device with an internet connection. In addition to unlimited storage space per user, JHOneDrive also allows users to share documents with colleagues within or outside of JHU. OneDrive meets all HIPAA and FERPA compliance standards for secure file sharing and storage. Data Access Groups will be used by the staff at the clinical sites, the Coordinating Center, the OCT and AOSLO Reading Centers, central genetic testing lab, the central bioanalytics lab for NAC testing and at the Genetic Analysis Collaborator Dr. Hufnagel's lab. Each site will only have access to the data from their own site. The Coordinating Center will be able to monitor and revoke access of any individual at any time.

###### 7.4.2. OCT Reading Center Data Transmission System

The NAC Attack OCT Reading Center (OCTRC) is the Duke Reading Center located in the Duke Eye Center at Duke University, Durham, NC, USA (Section 10.6.1). The OCTRC will provide the NAC Attack Clinical Sites detailed written procedures for transmitting images to the OCTRC through the Data Transmission Site (DTS). The DTS is the OCTRCs FDA regulation (21 CFR Part 11) compliant portal in accordance with the study protocol.

For the a few clinical sites participating in the AOSLO sub-study, AOSLO montage files transfer to the AOSLO Reading Center will follow relevant procedures of the AOSLO Reading Center through 21 CFR Part 11 compliant cloud-based portal.

###### 7.4.3. Clinical Sites Submission of Data Not Collected Through CRF

In addition to data collected on CRFs and managed through REDCap, there are raw files generated through ocular imaging and tests, such as OCT image files and MAIA microperimetry test results. **Table 11** summarizes the sources of such data including the associated study procedures and study visits, and the submission and transfer of these data. Procedures for submissions of raw imaging and test files to the Coordinating Center, OCT Reading Center and AOSLO Reading Center should follow the individual manuals from these centers.

**Table 11 Summary of transfer and management of data files from clinical sites that are not collected through case reporting forms. The schedule of activities can be found in** ***Table 1*****.**

| **Patient Data (non-CRF based)** | **At Which Study Visits** | **Submission to Where and Timeline** |
|---|---|---|
| Patient candidate's histori-cal medical imaging and/or reports such as below. In-formation about patient names and date of birth should be removed<br>- Genetic testing report<br>- Visual field report<br>- Microperimetry result<br>- ERG<br>- Fundus autofluores-cence imaging files<br>- OCT imaging files | Screening visit only | Coordinating Center Only |
| Patient candidate's imag-ing files from study imag-ing at the screening visit<br>- fundus photo (ul-trawide)<br>- Fundus fluorescence (ultrawide) | Screening visit only for fun-dus photo<br>Screening visit and M27 and M45 for FAF | Coordinating Center Only |
| Patient candidate's screen-ing visit imaging files<br>- OCT | Screening visit only | Coordinating Center and OCT Reading Center |
| Participant OCT imaging files | Baseline, M9, M18, M27, M36 M40.5, and M45 | OCT Reading Center Only |
| Site's local lab results for blood chemistry, hematol-ogy, liver function | Screening visit, M9, M18, M27, M36 and M45 | Coordinating Center Only |
| Microperimetry test results and .txt raw data files | Screening visit, baseline, M4.5, M9, M18, M27, M36 M40.5, and M45 | Coordinating Center Only |
| Scanned PDF copy of the source CRFs | All study visits (in-clinic, tele-visits, phone calls with | Coordinating Center Only |

| | | |
|---|---|---|
| | coordinators) | |
| FST test results | Baseline visit, M27 and M45 | Coordinating Center Only |
| AOSLO montage files (at selective sites) | Baseline visit, M9, M27 and M45 | AOSLO Reading Center Only |

###### 7.4.4. Management of Data from the Reading Centers and Study Central Labs

Data from the OCT Reading Center include datasets about patient candidate's OCT related eligibility assessment and OCT imaging grading results. These data will be transferred from the OCT Reading Center in the format of .csv files to the Coordinating Center cloud-based portal regularly. In addition, the study OCT imaging files will be transferred to the Coordinating Center within 1 year of the final study visit for the last participant enrolled.

Data from the AOSLO Reading Center include datasets regarding AOSLO sub-study participants' AOSLO imaging grading results. These data will be transferred from the AOSLO Reading Center in the format of .csv files to the Coordinating Center JHOneDrive cloud-based portal regularly.

Data from the study central labs include datasets of plasma NAC levels and genetic analysis results from the central lab for genetic testing and from the genetic analysis lab. These data will be submitted by the individual labs to the Coordinating Center cloud-based portal regularly.

##### 7.5. Retention of Records at Clinical Sites

Records and documents pertaining to the conduct of this study, including CRFs, Informed Consent Forms, laboratory test results, images, and medication inventory records, must be retained by the site Principal Investigator for at least 5 years after completion or discontinuation of the study or for the length of time required by relevant local health authorities or institutions, whichever is longer. After that period of time, the documents may be destroyed, subject to local regulations.

Written notification should be provided to the Coordinating Center prior to transferring any records to another party or moving them to another location.

### 8. Ethical Considerations

##### 8.1. Compliance with Laws and Regulations

This study will be conducted in full conformance with the ICH E6 guideline for Good Clinical Practice and the principles of the Declaration of Helsinki, or the laws and regulations of the country in which the research is conducted, whichever affords the greater protection to the individual. The study will comply with the requirements of the ICH E2A guideline (Clinical Safety Data Management: Definitions and Standards for Expedited Reporting). Studies conducted in the United States or under a U.S. Investigational New Drug (IND) Application will comply with U.S. FDA regulations and applicable local, state, and federal laws. The study will also comply to foreign regulatory agencies the European Union or European Economic Area will comply with the E.U. Clinical Trial Regulation (No 536/2014).

##### 8.2. Informed Consent

For study sites in the United States, the consent form contains two parts: Part I also called master consent contains information about the study; Part II contains information of the study site specific to each patient interested in participating. The master consent form approved but unstamped by the study single IRB (s-IRB) together with the Part II Site-Specific Consent Information (SSCI) will be provided to study sites in the United States; and each US site will develop Part II the SSCI locally to ensure the consent form fulfills local requirements. The SSCI will be merged with the approved master consent form, stamped and approved for each participating stie. When applicable to a US site, the master consent form will be provided in a certified translation in Spanish.

For study sites outside of the US, the master consent form will be provided to each site for translation to their local language (if applicable) and also adaptation to local consent template. The translated copy should be reviewed and approved by the Study Chair's office before submission for local ethical review.

The Study Chair or his designee must review and approve any local part of the Informed Consent Forms or any alternate consent forms proposed by the site (collectively, the "Consent Forms") before IRB submission. The final IRB–approved Consent Forms must be provided to the Coordinating Center for regulatory authority submission purposes according to local requirements.

The Informed Consent Form will contain separate sections for any optional procedures. The investigator or authorized designee will explain to each patient the objectives, methods, and potential risks associated with each optional procedure. Patients will be told that they are free to refuse to participate and may withdraw their consent at any time for any reason.

The Consent Forms must be signed and dated by the patient before his or her participation in the study. The case history or clinical records for each patient shall document the informed consent process and that written informed consent was obtained prior to participation in the study.

The Consent Forms should be revised whenever there are changes to study procedures or when new information becomes available that may affect the willingness of the patient to participate. The final revised IRB-approved Consent Forms must be provided to the Coordinating Center for regulatory authority submission purposes.

If the Consent Forms are revised through an amendment while a patient is participating in the study, the patient must re-consent by signing the most current version of the Consent Forms or the addendum, in accordance with applicable laws and IRB policy. For any updated or revised Consent Forms, the case history or clinical records for each patient shall document the informed consent process and that written informed consent was obtained using the updated/revised Consent Forms for continued participation in the study.

A copy of each signed Consent Form must be provided to the patient. All signed and dated Consent Forms must remain in each patient's study file or in the site file and must be available for verification by study monitors at any time.

For sites in the United States, each Consent Form may also include patient authorization to allow use and disclosure of personal health information in compliance with the U.S. Health Insurance Portability and Accountability Act (HIPAA) of 1996. If the site utilizes a separate Authorization Form for patient authorization for use and disclosure of personal health information under the HIPAA regulations, the review, approval, and other processes outlined above apply except that IRB review and approval may not be required per study site policies.

##### 8.3. Institutional Review Boards

The trial follows NIH's Single IRB policy and uses a single IRB (sIRB) for clinical sites in the US. Sites outside the US use their local IRBs (ethical committees ). The study Chairman, with assistance from the Coordinating Center, takes the responsibility of obtaining and maintaining sIRB upon trial's initiation. Documentation of sIRB and non-US sites' local IRBs approvals and copies of approved consent forms will be kept on file with the Coordinating Center.

This protocol, the Informed Consent Forms, any information to be given to the patient, and relevant supporting information must be submitted to the local IRB by the PI and reviewed and approved by the IRB before the study is initiated. In addition, any patient recruitment materials must be approved by the IRB.

The Study Chairman is responsible for providing written summaries of the status of the study to the sIRB annually. The Coordinating Center will assist in drafting study status summaries. The PI at each non-US site is responsible for providing written summaries of the status of the study to the local IRB annually or more frequently in accordance with the requirements, policies, and procedures established by the local IRB. Investigators are also responsible for promptly informing the IRB of any protocol amendments.

In addition to the requirements for reporting all AEs to the Coordinating Center, investigators must comply with requirements for reporting SAEs to the local regulatory authority and IRBs. Investigators may receive written IND safety reports or other safety-related communications from the Coordinating Center. Site investigators are responsible for ensuring that such reports are reviewed and processed in accordance with local regulatory authority requirements and the policies and procedures established by their local IRBs, and archived in the site's study file.

##### 8.4. Confidentiality

The Study maintains confidentiality standards by coding each patient enrolled in the study through assignment of a unique study identification number. This means that patient names are only known to the study team members of the clinical site where the patient is enrolled. Patient names or medical record numbers will not be included in data sets, images, test results or specimens transmitted to the Coordinating Center, the Reading Centers or study laboratories. Patient medical information obtained by this study is confidential and may be disclosed to third parties only as permitted by the Informed Consent Form (or separate authorization for use and disclosure of personal health information) signed by the patient, unless permitted or required by law.

Medical information as specified in the Consent Form may be given to a patient's personal physician or other appropriate medical personnel responsible for the patient's welfare, for treatment purposes.

Given the complexity and exploratory nature of exploratory biomarker analyses, data derived from these analyses will generally not be provided to Study Investigators or patients unless required by law. The aggregate results of any conducted research will be available in accordance with study policies and procedures on study results publication.

Data generated by this study must be available for inspection upon request by representatives of national and local health authorities, Coordinating Center protocol monitors, and the IRB for each Clinical Study Site, as appropriate.

Study data may be shared with researchers, government agencies, companies, or other groups that are not participating in this study to advance science and public health, or for analysis, development, and commercialization of products to treat and diagnose disease. The study data sharing will follow the NIH’s Data Sharing Policy published in the NIH Guide on February 26, 2003 (https://grants.nih.gov/grants/policy/data_sharing/data_sharing_guidance.htm). In accord with the NIH guidelines, a summary, de-identified NAC Attack data set will be made available through submission of the dataset to a government or other health research database. We will share individual-participant level data (IPD) together with their associated data dictionaries. The data will be made available 1 year after publication of the primary findings of the study, in a de-identified format. In addition to the IPD data set, the researchers will share the trial protocol including the statistical analysis plan, data collections forms, and analytic codes used in the main reports of the trial. The rights and privacy of people who participated in the Study will be protected at all times by stripping all identifiers including study IDs that could lead to disclosing the identity of individual research participants from the data. This commitment to privacy-protected data sharing will be incorporated in all levels of data sharing activities. In addition, redacted clinical study reports, retinal images, and other summary reports may be provided to researchers upon approval of their requests by the study leadership. The requesting researchers will be required to sign a data use agreement before they are given access to study reports or images.

#### 8.5. Benefit - Risk Considerations

##### 8.5.1. Potential Risks

There are potential risks associated with breach of confidentiality, the intervention, and study procedures.

There will be extremely low, if any, risk of breach of participant privacy or confidentiality. Any private identifying information of a participant will only be kept in a locked cabinet or a locked room at the enrolling clinical site and will not be sent to the Coordinating Center or any other trial entity. Any medical records of a study participant obtained outside of the Study procedures will be anonymized by site personnel and only identified by study ID before transferring to the Coordinating Center. In the event the Coordinating Center becomes aware of a study patient’s name or medical record number, the Coordinating Center will immediately delete the patient’s personal identifying information and request that the relevant clinical site resends the patient materials after the materials are appropriated anonymized.

All imaging files submitted to the Study OCT Reading Center and AOSLO Reading Center must be de-identified and only identified by study ID. If an image submitted mistakenly contains patient identifying information, the receiving Reading Center will immediately destroy it and request that an appropriately de-identified image be resent.

The risks of taking the active study drug, oral NAC treatment at 1800 mg bid are expected to be minimal. NAC is a generic medication approved by the FDA for acetaminophen overdose and for use as an adjuvant therapy in managing certain pulmonary conditions. Its safety profile has been well established [95]. Adverse events associated with oral NAC may occur including nausea, vomiting, diarrhea, flatus and gastroesophageal reflux. In the phase-1 FIGHT RP study[48], no ocular adverse events occurred in the RP patient participants.

All the vision related testing procedures in NAC Attack are routine clinical procedures in retina clinics, including best-corrected visual acuity testing, microperimetry, SD-OCT, fundus photographs, and fundus autofluorescence imaging. Vision-related quality of life surveys are common in ocular clinical trials. These are all non-invasive procedures that pose little or no risk.

Blood will be drawn for a variety of tests according to the schedule in **Table 1**. Venipuncture is a common procedure and there is no risk of exposure to transmissible diseases if the blood is drawn using aseptic conditions. Only trained phlebotomists at NAC Attack clinical sites will collect blood samples from study patients. A patient may feel lightheaded or faint, or experience pain or discomfort at the site of puncture during the procedure. There may be possible bruising or swelling at the puncture site, and rarely there may be an infection. To minimize burden due to blood draw, blood will only be drawn every 4.5 or 9-months (**Table 1**).

###### 8.5.2. Strategies for Protection of Study Subjects

All information that identifies individual study patients will be retained in the local clinical site. The linkages between personal identifiers and coded study identifiers will be maintained in a secure and confidential manner at each clinical center. None of the trial central resource centers will receive data or materials that contain personal identifiers. Therefore, the risk of loss of confidentiality due to trial participation is extremely low.

Strategies to protect again potential risks due to the intervention or study procedures include safety monitoring by a physician Medical Monitor, the Study Chairman, and an external, independent Data & Safety Monitoring Committee (DSMC) appointed by the National Eye Institute. Detailed safety monitoring procedures are in Section 5.

Due to the slow progression of RP, the study requires a follow-up period of 45 months, which imposes a burden on participants and Clinical Site personnel. To minimize the burden on all stakeholders, in-clinic visits are limited to time points when a primary outcome measurement are needed and supplemented with virtual visits to assess safety and compliance. Such a design feature will minimize participant travel burden while maintaining rapport to minimize drop outs.

###### 8.5.3. Potential Benefits of NAC Attack to Trial Participants and Others

Participants may benefit from information obtained from study visits or from some of the study procedures. RP patients may be aware of the FIGHT RP phase-1 trial results and may take NAC as a dietary supplement. However, while the safety of up to one year of oral NAC at doses of 600 mg or 1200 mg per day is well established, the long-term safety of 1800 mg twice a day is not known. The only prudent way to take such high doses of NAC is within the setting of a clinical trial in which study participants will be frequently examined by study investigators to assess safety.

Future patients which could be participants' family members may benefit from the important information about RP learned in the trial. Because NAC Attack will not restrict enrollment by the knowledge of genetic mutation of a participant, the study results will provide critical information that will have a major impact on *all* patients with RP, including patients themselves and their family members.

###### 8.5.4. Importance of the Knowledge to be Gained

Currently, there are no effective treatments for RP and most patients eventually lose all useful vision. Although pre-clinical mechanistic studies and the phase-1 FIGHT RP dose ranging clinical trial suggest NAC may be a promising pharmacologic option for slowing cone degeneration, this can only be definitively determined by a randomized, placebo-controlled clinical trial with a sufficient sample size. NAC Attack will determine if NAC provides long-term benefits in patients with RP. In addition, although NAC's safety profiles have been well established and NAC is an FDA approved drug, the approved usages and safety data are for short-term use of the drug. NAC Attack will provide long-term safety data which are important, because if NAC is proven efficacious for RP, it will be a life-long treatment.

Moreover, currently impending loss of vision in patients with RP is a major source of stress throughout life which can be compounded by fear of passing the disease to children. In addition to possibly providing direct benefits on visual function, NAC could provide added benefits with regard to reducing stress and improving self-image. Therefore, a randomized placebo-controlled trial is warranted, and the benefit-risk ratio is very high.

In addition to the knowledge of the treatment effect and safety of NAC, NAC Attack will generate valuable data regarding the natural history of RP from patients of diverse genetic background. The data will also allow a better understanding of the genetic causes of RP and provide insights into the mechanisms of disease pathogenesis, in particular the role of oxidative stress in RP. Thus, the research findings from NAC Attack may also contribute to the development of future new interventions.

##### 8.6. Financial Disclosure

Investigators will provide the Coordinating Center with sufficient, accurate financial information in accordance with local regulations to allow the Coordinating Center to submit complete and accurate financial certification or disclosure statements to the appropriate health authorities. Investigators are responsible for providing information on financial interests annually during the course of the study and for 1 year after completion of the study (see definition of "end of study" in Section 3.2).

##### 8.7. Participant Care After Study Participation

At present standard care for RP is observation. Most NAC Attack participants will be enrolled by participating study retina specialist and retina clinic that was managing them before NAC Attack and after completion of the trial the same retina specialist or retina clinic will continue to care for the patient. Participants referred to a NAC Attack study clinical site from another retina specialist will resume standard care with the referring retina specialist. Current standard care for RP is observation but depending upon the results of NAC Attack, treatment with N-acetylcysteine could be incorporated into standard care and if so, instructions will be provided to retina specialists who provide care for former NAC Attack participants.

### 9. Study Documentation, Monitoring, and Administration

#### 9.1. Study Documentation

The investigator must maintain adequate and accurate records to enable the conduct of the study to be fully documented, including, but not limited to, the protocol, protocol amendments, Informed Consent Forms, and documentation of IRB and regulatory approval. In addition, at the end of the study, the investigator will receive the patient data, including an audit trail containing a complete record of all changes to data.

#### 9.2. Protocol Deviations

The Investigator should document and explain any protocol deviations. The Investigator should promptly report any deviations that might have an impact on patient safety and data integrity to the Coordinating Center and to the IRB in accordance with established IRB policies and procedures. Protocol deviations may also be identified during Coordinating Center's site monitoring visits or quarterly conference call with clinical sites.

The Coordinating Center will review all protocol deviations and assess whether any represent a serious breach of Good Clinical Practice guidelines and require reporting to health authorities.

#### 9.3. Management of Study Quality Through Training and Certification

The Coordinating Center will implement a NAC Attack Training and Certification Program to ensure standardized training on all protocols and data collection procedures, along with certification of NAC Attack personnel. The NAC Attack Steering and Quality Assurance Committee oversees the implementation of the training program. The Coordinating Center is responsible for finalizing certification requirements, developing the on-line system for completing knowledge assessments, providing customized feedback, issuing certification certificates, and maintaining the electronic and paper roster of all certified study personnel.

All members of the Investigative Group must complete a General Knowledge Assessment about NAC Attack that requires knowledge of basic design facts (e.g. duration and frequency of participant follow-up, the primary and secondary outcome measures), AE and IRB reporting guidelines and timelines. In addition, individuals are required to complete role-specific assessments.

Knowledge assessments must be completed by the original NAC Attack Study Group members and by all new personnel at the time of hiring. The Coordinating Center Director and Protocol Monitor are responsible for preparing certification materials and reviewing the certification materials completed by Study Coordinators, BCVA Examiners and MAIA Microperimetry Examiners, respectively, and contacting the respondents if there are areas of misunderstanding. Trained Readers at the OCT Reading Center are responsible for reviewing certification materials submitted by OCT Technicians. The study Chairman or his designees at the Coordinating Center or Reading Centers is responsible for reviewing materials submitted by PIs and Co-Is for NAC Attack certification. The

Coordinating Center maintains a log of all people who have successfully completed certification requirements.

Prior to becoming eligible for NAC Attack certification, all investigators and clinical site staff must have training in protecting the rights and welfare of human subjects involved in clinical research. Current certificates documenting the successful completion of a Human Subjects Training program compliant to the relevant regulations (e.g. HIPAA for clinical sites in the US) must be submitted to the Coordinating Center by all members of the investigative group prior to NAC Attack certification.

NAC Attack Study personnel must be certified before performing any study procedures. All people seeking certification will receive a checklist of certification requirements that must be met for each study-specific role prior to any interaction with study patients. Checklists and materials needed to complete certification requirements will be made available by the Coordinating Center.

Maintaining certification in some NAC Attack roles is contingent on performing the procedure within a specified time period. If study personnel have not performed the procedure for which certification was issued within the specified time, the person will be decertified for the role and the Principal Investigator and Study Coordinator for the site will be notified by the Coordinating Center.

#### 9.4. Site Monitoring

##### 9.4.1. Monitoring Visits to Clinical Site

Periodic site visits by an independent observer are necessary to ensure that there is standardization of procedures, that clinic personnel have been trained adequately, that the clinic facilities meet standards, and that patients and their data are being managed as specified in the protocol. The site visitor also provides assistance in solving logistical problems by conveying efficient, accurate solutions used in one clinical site to other clinical sites. All sites will be visited by a Coordinating Center protocol monitor within a few months of the initiation of patient recruitment and will then be visited on a staggered schedule about 2-3 years later when their first enrolled participants are around their 27months follow-up. In addition to periodic random visits or virtual visits, there is a risk-based monitoring plan. A monitoring visit will be triggered when a site has multiple protocol deviations, problems with data reporting, or incongruities between database entries and source documents. In between on-site visits, there may be virtual visits through Zoom meetings. Clinical Sites may be visited more frequently if the SMQAC deems it necessary due to problematic performance or clinic staff turnover. A written summary prepared by the site monitor will be sent to the site PI and Study Coordinator, and members of the Site Monitoring and Quality Assurance Committee. A copy of the report is also maintained in the Coordinating Center library of NAC Attack documentation.

##### 9.4.2. Quarterly Conference Calls with Clinical Sites

A teleconference is scheduled once every 3 months (unless a site visit has recently occurred) between the Coordinating Center staff and Study Coordinator to make sure that changes (if any) in NAC Attack personnel, facilities, equipment, site's communication with local IRB and relevant regulatory agency have been communicated and that progress is being made in any problem areas of performance. The status of certifications and re-certification requirements are reviewed. The Study Coordinators should bring any problems, either within the Clinical Site, or with the Coordinating Center or OCT Reading Center, to the attention of the Coordinating Center staff.

###### 9.4.3. Study Medication Storage Monitoring

Section 4.14.1 describes monitoring and quality assurance related to drug distribution, storage and accountability.

##### 9.5. Dissemination of Data

Regardless of the outcome of the trial, the Study Chair and Coordinating Center Director are dedicated to openly providing information on the trial to healthcare professionals and to the public, at scientific congresses, in clinical trial registries of the U.S. National Institutes of Health and the European Medicines Agency, and in peer-reviewed journals. The Study Chair and Coordinating Center Director will comply with all requirements for publication of study results. Study data may be shared with others who are not participating in this study, and redacted clinical study reports and other summary reports will be provided upon request.

Study publications are defined as those that use data, documents, or other information collected during the course of the Study. Publication of study results will be governed by the policies and procedures developed by the Executive Committee (Section 11.2). The Executive Committee reviews all written reports prepared for publication. A subcommittee of the Executive Committee ensures that the preparation of the results for abstracts for meetings or manuscripts complies with NIH policies and guidelines, and appropriate analysis and conclusions are reached.

##### 9.6. Protocol Amendments

All significant changes to the NAC Attack protocol must be approved by the NAC Attack Executive Committee and DSMC before implementation of any changes. In some circumstances, approval from NEI also may be required. If the DSMC recommends a protocol modification, the Study Chair and PI of the Coordinating Center, in consultation with the NEI, will be responsible for implementing the change. If the Study Chair, PI of the Coordinating Center, and/or NEI do not concur with the DSMC's recommendation, it will be their collective responsibility to work with the DSMC Chair to resolve differences and find a mutually acceptable resolution. All DSMC recommendations for protocol modifications must be accompanied by adequate rationale.

When a change in protocol is implemented, a protocol memorandum is issued to clinical site staff; pertinent information posted on the NAC Attack website is updated to reflect the change. All NAC Attack clinical site staff will be required to acknowledge receipt of the protocol memorandum that describes the change and date of implementation and understanding of its contents by signing and dating a form that is sent to the Coordinating Center. During site visits, the Protocol Monitor will review whether all NAC Attack protocols, forms and other documents are up to date.

The Study Chair ensures that FDA, the s-IRB and local IRBS, and other relevant non-US regulatory agencies and ethical review committees are notified of any protocol changes on a timely manner.

Approval must be obtained from the IRB and regulatory authorities (as locally required) before implementation of any changes, except for changes necessary to eliminate an immediate hazard to patients or changes that involve logistical or administrative aspects only (e.g., change in Medical Monitor or contact information).

### 10. Study Administrative Structure

The organization structure of the NAC Attack trial can be described in two ways:

- Study sites and other stakeholders
- Standing committees

#### 10.1. Overview of Study Sites and Other Stakeholders

The study sites include the Clinical Sites, Coordinating Center, Chairman's Office, and 2 resource centers: the OCT Reading Center (OCTRC) and the AOSLO (Adaptive Optics Scanning Laser Ophthalmology) Reading Center. The investigators and personnel at these sites have responsibilities that begin with initiation of trial activities, continue throughout participant treatment and follow-up data collection, and end only after trial findings have been analyzed and disseminated.

In brief about the responsibilities of clinical sites, at a minimum, each Clinical Site includes an ophthalmologist who is designated the site principal investigator (PI) and a Site Coordinator who is responsible for the local day-to-day trial activities. The roles and personnel of the Study Chair's office, the Coordinating Center and of the 2 resource centers are summarized below. Other resource centers have more limited responsibilities. Details of responsibilities are described elsewhere, as indicated.

Other Stakeholders that provide resources or services are the National Eye Institute (NEI, funder of the study), the Drug Labeling and Distribution Center (DLDC), and central laboratories with specific capabilities and roles (see Section 10.7).

#### 10.2. Overview of Standing Committees

The Study Chairman and Coordinating Center Director comprise the Study Leadership. The standing committees of the trial (Section 10.8) include the *Executive Committee*, the *Steering and Quality Assurance Committee (SQAC), the Investigative Group*, and *the Data and Safety Monitoring Committee (DSMC)*. Subcommittees to the SQAC may be appointed from time to time for specific purposes. Medical monitoring may be assigned to either a subcommittee of the SQAC or of the DSMC.

#### 10.3. Study Chairman's Office

The study Chairman's Office is responsible for maintaining the trial organization as a group of collaborating investigators and personnel and for interactions between the Investigative Group and the medical community.

The Study Chairman is Peter A. Campochiaro, MD, who conducted the research that led to the design of the randomized trial and who worked closely with the Director of the Coordinating Center to develop the trial protocol and procedures and the funding applications submitted to the National Eye Institute (NEI). The Chairman's Office is housed within the Retina Division of the Wilmer Eye Institute, Baltimore, Maryland. The primary role of the Chairman's Office is to provide clinical and scientific leadership for the trial from initiation through publication of key findings. The study Chairman is an *ex officio* member of all standing committees.

##### 10.4. Coordinating Center

Director of the Coordinating Center for NAC Attack is Xiangrong Kong, PhD, the statistician who worked closely with the Chairman to publish research findings that led to planning of the NAC Attack randomized trial. She was PI of the R34 planning grant awarded by the NEI to support development of the trial protocol and methods for its implementation. She is the PI of the NEI award to support the activities of the Coordinating Center. The Coordinating Center PI is an *ex officio* member of all standing committees. She may designate other Coordinating Center personnel to serve in her place on one or more committees.

A key role of the Coordinating Center is to assure that all participating sites adhere to the trial protocol so that the findings from the trial are scientifically valid. The Coordinating Center has responsibility for equitable disbursement of study financial resources to Clinical Sites and for monitoring distribution of study drugs to Clinical Sites and/or trial participants. In addition to providing expertise in clinical trials research methods as part of the scientific leadership of the trial, the Coordinating Center is responsible for assembling and maintaining a current, accurate, and complete trial database and preparing and disseminating reports based on the accumulated data, monitoring the performance of all Clinical Sites, and participating in dissemination of trial findings by statistical analysis of trial data and contributions to manuscripts to present findings. The Coordinating Center has a key role in each of the organizational requirements of the trial.

The NAC Attack Coordinating Center is located in the Wilmer Eye Institute, Baltimore, Maryland, USA. It is administratively distinct and physically separated from the Chairman's Office. The proximity of the Chairman's Office and Coordinating Center facilitates communication and interaction of trial personnel at these two study centers.

##### 10.5. Clinical Sites

Each Clinical Site includes one or more investigators and one or more study coordinators. A single investigator is designated the lead PI and is responsible for conduct of the study at the Clinical Site. There may be other co-investigators (Co-Is) that share responsibility managing study patients at the site. Either the PI or one or more Co-I must be an ophthalmologist who is responsible for informed consenting, eye examinations, dispensing study medication, and managing and reporting AEs and SAEs.

The following roles are handled by study coordinators: measurement of BCVA, microperimetry examinations, scheduling and overseeing patient visits, sample management (obtaining, storing, and shipping), data entry, and uploading images. SD-OCT, fundus photography, and fundus autofluorescence is performed by a site photographer or technician. Certain site personnel will be certified for study activities by the Coordinating Center.

##### 10.6. Study Resource Centers

###### 10.6.1. OCT Image Reading Center

The OCT Image Reading Center (OCTRC) is located in the Duke Eye Center at the Duke University Hospital, Durham, NC, USA. It is directed by Glenn Jaffe, MD and Sina

Farsiu, PhD. The primary role of the OCT Image Reading Center is to assure that OCT images of study eyes are of good quality, to provide the necessary measurements and interpretations required to judge the strict eligibility of each participant enrolled, and to produce the baseline and follow-up data for analysis of OCT based primary and exploratory outcomes for the trial. Thus, the directors and personnel of this center provide training of OCT Technicians and monitor their performance. The data from evaluation, measurements, and interpretation of OCT images are transmitted to the Coordinating Center for integration with other trial data for each participant.

###### 10.6.2. AOSLO Image Reading Center

The AOSLO Image Reading Center is a virtual reading center and co-directed by Jacque Duncan, MD at the Retinal Degenerations Clinic of the University of California San Francisco, San Francisco, CA and Joseph Carroll, PhD at the Eye Institute of Medical College of Wisconsin, Milwaukee, WI, USA. The role of the AOSLO Image Reading Center is to provide measurements and interpretations of the exploratory outcome measures of cones based on AOSLO imaging obtained at participating centers of the NAC Attack AOSLO sub-study. The directors and their personnel will provide training of AOSLO Technicians and monitor their performance. The data from evaluation, measurements, and interpretation of OCT images are transmitted to the central database at the Coordinating Center for integration with other trial data for each relevant participant.

##### 10.7. Other Trial Stakeholders

###### 10.7.1. National Eye Institute

The funding sponsor for the NAC Attack Trial is the NEI, part of the U.S. National Institutes of Health (NIH). An NEI Program Director, serving as Project Coordinator, will have substantial programmatic involvement that is above and beyond the normal stewardship role in awards, as described below. The appropriate NEI extramural Program Director from the Division of Extramural Science Programs whose name appears on the Notice of Grant Award (NoA) will:

- Nominate members of an independent DSMC.
- Assist the Study Chairman and Coordinating Center Director in the identification of additional participating clinics, if necessary, to enhance patient recruitment.
- Assist the Study Leadership Committee in site visits and routine performance monitoring of the entire study including matters of quality control within and among various components, and in the determination of inadequate patient recruitment or failure to comply with the protocol on the part of individual clinics or study core centers. The NEI Program Director will attend and participate in study meetings as appropriate.
- Assist the Editorial/Writing Committee in the preparation and review of study results for publication.
- Assist the DSMC as an expert resource in their evaluation of safety, efficacy, quality and progress on an ongoing basis, as applicable. Serving as a steward of federal funds, the NEI Program Official will assist but not direct deliberations and

decisions of the Committee, e.g., proceeding from one phase of the study to the next; implementing protocol changes, evaluating study progress and quality including patient recruitment, overall clinical and resource center performance; monitoring study timeliness and progress toward meeting milestones, approving ancillary studies, planning data analysis; releasing unmasked data; announcing study findings; determining the timing of release of any interim or final reports; and, reviewing primary outcome manuscript(s) prior to journal submission.

The NEI reserves the right to curtail, withhold, or terminate support for the study, for an individual award, or support for a participating consortium, in situations involving: inadequate progress toward meeting study milestones including those related to: availability and regulatory approval of study product as applicable, patient recruitment, follow-up, data reporting, or quality control; a major breach of the study protocol or NEI/NIH policy; a substantive change in the agreed-upon protocol to which the NEI does not agree; statistical evidence that the major study endpoint has been reached ahead of schedule; or, human subject ethical issues that dictate a premature termination. Prior to taking such actions, NEI will consult with and receive recommendations from the DSMC. Additionally, the NEI Program Director will be responsible for the normal scientific and programmatic stewardship of the award and will be named in the award notice.

###### 10.7.2. Drug Manufacturer

The Drug Manufacturer is Zambon Switzerland Ltd, Via Industria 13, 6814 Cadempino, Switzerland who will manufacture 600 mg effervescent tablets of NAC (Fluimucil) and identically appearing placebo tablets for the trial. Fluimucil and placebo will be produced in multiple lots to ensure that unexpired drug will be available throughout the duration of the trial.

###### 10.7.3. Drug Labeling and Distribution Center

The Drug Labeling and Distribution Center (DLDC) will provide secondary packaging of NAC and placebo tablets. The packages will be labelled with codes to ensure masking. The DLDC will ship the processed medication packages to each site using temperature-controlled shipment.

###### 10.7.4. Other Centers/Laboratories/Collaborators

Other centers that will provide specific services or that have specific roles in the NAC Attack Trial are:

- Central laboratory responsible for assaying NAC in plasma.
- Central laboratory for whole genome (exome) sequencing.
- Genetic data analysis collaborator Robert B. Hufnagel, MD, PhD, Staff Clinician, Ophthalmic Genetics and Visual Function Branch, National Eye Institute, National Institutes of Health, Bethesda, MD, USA.

##### 10.8. Standing Committees

###### 10.8.1. Executive Committee

Many issues arise day to day during the conduct of a multicenter randomized clinical trial that require rapid resolution and others that are logistical and do not require advice or consideration by the larger Steering and Quality Assurance Committee. These issues are handled by the Executive Committee made up of the Study Leadership including:

- Study Chairman
- Coordinating Center Director
- NEI Representative(s)

And

- Additional members of the Steering and Quality Assurance Committee may be asked to participate in meetings of the Executive Committee as necessary to make informed decisions that are based on all relevant information.

###### 10.8.2. Steering and Quality Assurance Committee

The Steering and Quality Assurance Committee (SQAC) is a group of trial investigators, clinical coordinators and other stakeholders that serves as the primary source of advice to the Study Chairman and the Coordinating Center PI regarding scientific, policy, administrative and logistical issues encountered during the conduct of the trial. They periodically review performance of Clinical Sites and resource centers and provide feedbacks and suggestions to the Executive Committee. Members of the SQAC are appointed by the Study Chairman and Coordinating Center Director.

The members of the SQAC are:

- Study Chairman
- Study Vice-Chairmen
- Coordinating Center Director who also serves as study Statistician
- Protocol Monitors(s) from the Coordinating Center
- PIs and Project Manager of the Reading Centers
- NEI Representatives
- Representative from the Drug Labeling and Distribution Center
- Two to four Clinical Site Investigators who serve 1 or 2-year terms
- Genetic Data Analysis Center Collaborator
- Two to four Study Coordinator representative who serve 1 or 2-year terms
- Representatives from other entities as appropriate.

###### 10.8.3. Investigative Group

The Investigative Group consists of all trial personnel at all participating Clinical Sites, Study Chairman’s office, Coordinating Center, resource centers, and other entities. The responsibility of the Investigative Group is to implement the trial protocol. The Investigative Group meets at least annually, either in full or in selected subgroups of centers or trial roles.

###### 10.8.4. Data and Safety Monitoring Committee (DSMC)

The DSMC is responsible for monitoring the ethical conduct of the NAC Attack Trial and for monitoring the accumulating trial data for evidence of adverse and beneficial effects of NAC, the study drug under evaluation. The DSMC provide recommendations to study leadership and the NEI. The members of the DSMC are the only people who see accumulating data regarding study outcomes by drug versus placebo until they have made a decision regarding drug effects relative to placebo (inactive treatment). The DSMC operates under a charter negotiated among the DSMC members, the Executive Committee, and the NEI Officials. The DSMC decides when findings from the trial regarding primary and secondary outcomes defined in the study protocol may be released to the trial investigators, to trial participants, and to the medical community. The DSMC also oversees the informed consent process developed for the trial.

### 11. Study Policies: Publicity, Publication, Ancillary Studies, and Prompt Reporting to IRB

#### 11.1. Publicity Policy

All publicity and press releases on behalf of NAC Attack are to have prior approval of the Executive Committee. NAC Attack investigators who are approached by the press for information concerning NAC Attack should refer these inquiries to the Study Chairman. It is recognized that when information is sought from an individual investigator by the local press in his or her own community, it is sometimes necessary or desirable for the investigator to handle the request him/herself. In such an event, the participating investigator who gives information should speak as an individual and not as the official representative of NAC Attack. This fact should be made clear to the press; however, the information given should be accurate and reflect the general policy and views of the group.

During the recruitment phase of the study, announcements (pre-approved by the NAC Attack Executive Committee and the IRB) may be placed in local media (newspaper, radio, television, online). The Coordinating Center also prepares for each clinical site a set of slides to present at local professional society meetings to aid in recruitment and to enhance study visibility. On a national level, study publicity will be increased by NAC Attack's study webpage hosted by the Coordinating Center, by postings on the NEI and ClinicalTrials.gov web sites, and by mailings to RP patient groups.

#### 11.2. Publication Policy

NAC Attack publications are defined as those that use data, documents, or other information collected during the course of the Study. Publication of the results of NAC Attack trial will be governed by the policies and procedures developed by the Executive Committee. The Executive Committee reviews all written reports prepared for publication.
A subcommittee of the Executive Committee ensures that the preparation of the results for abstracts for meetings or manuscripts complies with NIH policies and guidelines, and appropriate analysis and conclusions are reached.

##### 11.2.1. Authorship

All reports from NAC Attack will list the members of the writing committee as authors followed by "for the NAC Attack Study Group". Additionally, all NAC attack personnel, past and present, may be listed with the approval of the site PI.

##### 11.2.2. Manuscript Writing Teams

The NAC Attack Executive Committee will determine potential manuscript topics based on interim analyses and study hypotheses and will invite site investigators and other relevant personnel to join the writing committee for each manuscript. Writing committee members will be further determined based upon level of contribution to the NAC Attack trial including site enrollment and performance (retention and data quality) and committee work. All reports from NAC Attack will also list "for the NAC Attack Study Group" in the authorship.

###### 11.2.3. Manuscript Pre-Submission Review

Manuscripts prepared for publication must be sent to the NAC Attack Chairman and the Coordinating Center Director for review and approval by the Executive Committee. If approved by the Executive Committee, the manuscript is then sent to the DSMC for review and approval.

Oral presentations of more than local scope must be approved in advance by the Executive Committee. Abstracts to be printed must be approved by the Executive Committee. The DSMC, at their initial meeting, may also decide to mandate their review of oral presentations and abstracts in advance. No unpublished study results may be used for oral presentations, local or otherwise, unless the Executive Committee grants a specific exception. The above restrictions do not apply to local presentations on the design of the NAC Attack Trial, provided these presentations contain no unpublished Study results. Such presentations are encouraged to stimulate recruitment.

Copies of Study manuscripts will be sent to all PIs as well as members of the Executive Committee and the DSMC for information before publication. Reprints of publications will be mailed to members of the DSMC and to each center for distribution among the staff.

Manuscripts emanating from ancillary studies (Section 11.6) must be sent to the Executive Committee for review before submission for publication.

##### 11.3. Site's Local Use of Data from NAC Attack Participants

Site investigators are welcome to analyze data collected under NAC Attack for participants enrolled at their own site. However, any publication or presentation beyond a local audience of such data and analysis must be labeled as preliminary and not representative of the trial as a whole and must be approved by the NAC Attack Executive Committee beforehand.

##### 11.4. Acknowledgements

Each publication must acknowledge support from NEI.

##### 11.5. Clinical Trials Registration and Results Submission

In accord with NIH policy, NAC Attack will be registered at ClinicalTrials.gov no later than 21 calendar days after the enrollment of the first participant, and the trial results information will be submitted to ClinicalTrials.gov no later than one year after NAC Attack primary completion date.

##### 11.6. Ancillary Studies

Individual investigators who wish to carry out ancillary studies are encouraged to do so as long as they do not impose an additional burden on trial participants and do not divert financial and other resources intended for the conduct of the trial. Ancillary studies may greatly enhance the value of the NAC Attack Study and ensure the continued interest of many investigators. However, to protect the integrity of the NAC Attack Trial, ancillary studies must be reviewed and approved by the Executive Committee, subcommittee of the Steering and Quality Assurance Committee and DSMC before their initiation, whether or not they involve the Coordinating Center personnel, access to the trial database, or personnel at (other) resource centers.

###### 11.6.1. Definition of a NAC Attack Ancillary Study

An ancillary study is a research study that requires either:

- Supplementary observations or procedures to be performed upon all or a subgroup of NAC Attack participants according to a set protocol, or,
- Additional effort or activity by the Coordinating Center, OCT or AOSLO Reading Center, or Study Chairman's Office beyond the current scope of NAC Attack.

###### 11.6.2. Reasons for Requirement of Approval

Everyone concerned with NAC Attack is entitled to assurance that no ancillary study will complicate the interpretation of the NAC Attack results; adversely affect patient cooperation; jeopardize the public image of NAC Attack; or create a serious diversion of NAC Attack resources locally or at the Resource Centers.

###### 11.6.3. Preparation of Request for Approval of a NAC Attack Ancillary Study

The request for approval of an ancillary study should be in narrative form. It should contain, whenever relevant, the following items:

- Description of the objectives, methods, and significance of the study.
- Full details concerning any procedures to be carried out on any NAC Attack participants.
- Descriptions of any observations to be made or procedures to be performed on a study participant outside of the Clinical Site, and the extent to which the ancillary study will require extra clinic visits by the participant or will lengthen the participant's usual clinic visits.
- Description of any new grading to be conducted with any imaging modality captured in NAC Attack
- Data that required from the trial database maintained by the Coordinating Center
- Responsible ancillary study personnel
- Provisions for funding ancillary study activities, including data analysis

###### 11.6.4. Publication of Ancillary NAC Attack Results

All manuscripts or presentations for scientific meetings based on ancillary study data must be reviewed and approved by the NAC Attack Executive Committee before publication or presentation. Such review will pertain to the expected impact on NAC Attack Study objectives and not to scientific merit alone. Appropriate acknowledgment of NAC Attack resources used—whether data, participants, or NAC Attack investigators—should be included.

##### 11.7. Prompt Reporting to Coordinating Center and IRB

Required by Johns Hopkins University School of Medicine’s organizational policy, the following events (Section 11.7.1) should be reported promptly to the Johns Hopkins sIRB which oversees the study Coordinating Center. All NAC Attack Clinical Sites (both US and non-US based sites) and Resource Centers should report these events to the Coordinating Center within *5 working days* after the sites becoming aware of a reportable event unless the event is an SAE for which the timeline of immediate reporting in Section 5.2.2 should be followed. The Study Chair's Office with assistance of the Coordinating Center then will report to the sIRB within 5 working days upon receiving the event report from the Clinical Sites. If required by the Clinical Site's local policy, the Clinical Site should also report to such events to their local IRBs.

The Study Chair's Office with assistance of the Coordinating Center will also report any reportable event reported by other trial stakeholders such as the Drug Manufacture, the DLDC, and Study Central Laboratories to the sIRB within 10 working days upon Coordinating Center receiving notice of the event.

##### 11.7.1. Reportable Events Requiring Prompt Reporting to IRB

**I. UPIRSO:**

All potential "unanticipated problems involving risks to subjects or others"("UPIRSO").An event is considered an UPIRSO when it meets all of the following criteria:

(1) It is unexpected (in terms of nature, severity, or frequency) given (a) the research procedures that are described in the protocol-related documents, such as the IRB-approved research protocol and informed consent document; and (b) the characteristics of the population being studied;

Unexpected events could be either medical or non-medical events.

(2) It is related or possibly related to participation in the research ( i.e. there is a reasonable possibility that the incident, experience, or outcome may have been caused by the procedures involved in the research);

and,

(3) It places subjects or others [e.g. study team members or relatives of a subject] at a greater risk of harm (including physical, psychological, economic, or social harm) than was previously known or recognized

**II. POTENTIAL SERIOUS OR CONTINUING NON-COMPLIANCE:**

Non-Compliance is defined as the failure to follow the research protocol, federal, state, or local laws or regulations governing human subjects research, institutional policies, or the requirements or determinations of the IRB. Only incidents that may qualify as serious or continuing non-compliance must be promptly reported:

(1) Serious Non-compliance is defined as non-compliance that either (a) significantly

harms or poses an increased risk of significant harm to subjects or others, or (b) significantly compromises the rights and welfare of the subjects or the integrity of the Organization's human research protection program.

(2) Continuing Non-compliance is defined by the Organization as a pattern of non-compliance that significantly compromises the scientific integrity of the study or the rights and welfare of the subjects or the integrity of the Organization's human research protection program. When applying this definition, particular consideration may be given by the IRB to activity that recurs after a previous report has been evaluated by the IRB and corrective action has been instituted.

**III. OTHER EVENTS THAT REQUIRE PROMPT REPORTING:**

1) Unexpected adverse device effects (UADEs):

UADEs are defined as: "any serious adverse effect on health or safety or any life-threatening problem or death cause by, or associated with, a device, if that effect, problem, or death was not previously identified in nature, severity, or degree of incidence in the investigational plan or application (including a supplementary plan or application), or any other unanticipated serious problem associated with a device that relates to the rights, safety, or welfare of subjects." (21 CFR 812.3(s)).

2) Potential Breaches of Confidentiality: Any unauthorized disclosure of subject's personally identifiable information.

Please Note: Potential breaches of confidentiality that involve protected health information (PHI) must also be reported promptly to the HIPAA Privacy Officer.

3) Incarceration of a participant in a study not approved by the IRB to involve prisoners and the study team plans to continue study activities with prisoners while incarcerated.

4) Unresolved Subject Complaints: Complaints of subjects when the complaint indicates unexpected risks or cannot be resolved by the research team.

Events that do not meet the above criteria should be summarized and reported to the IRB at the time of continuing review.

##### 11.8. Data and Resource Sharing

The study will follow the NIH's Data Sharing Policy. The "Final NIH Statement on Sharing Research Data" was published in the NIH Guide on February 26, 2003 (https://grants.nih.gov/grants/policy/data_sharing/data_sharing_guidance.htm). In accord with the NIH guidelines, a summary, de-identified NAC Attack data set will be made available through Vivli which is a recently formed Global Clinical Research Data Sharing Platform https://vivli.org/ of which the Johns Hopkins Medicine is a member.

The final dataset will include demographic, visual functions test results, ocular imaging gradings, vision related QOL, and lab assay data on drug levels. We will share individual-participant level data (IPD) together with their associated data dictionaries. The data will be made available 1 year after completion of the study (symbolized by the publication of the primary findings of the study), in a de-identified format. The deidentification will include steps including replacing Study IDs with random codes and random perturbation of all study dates. In addition to the IPD data set, the researchers will share the trial protocol including the statistical analysis plan, data collections forms, and analytic codes used in the main reports of the trial.

The rights and privacy of people who participated in the Study will be protected at all times by stripping all identifiers that could lead to disclosing the identity of individual research participants from the data. This commitment to privacy-protected data sharing will be incorporated in all levels of database design.

Vivli is a non-profit clinical research data sharing platform that has been created to meet the needs of researchers who use and produce clinical research data worldwide. In order to access IPD arising from this project, users must complete the Vivli data request form and sign the Vivli Data Use Agreement, which limits subsequent use to the terms of the approved request and requires that users maintain data security, and refrain from any attempts to re-identify research participants or engage in any unauthorized uses of the data. In order to get access to the data, the user must submit a valid scientific question, include a statistical analysis plan, and complete all required fields on the Vivli data request form. Vivli will review the data request for completeness. Anyone who has submitted an approved data request and signed a data use agreement on Vivli will be given access to the data. Vivli will then make the data available, without cost, to users. Vivli will maintain storage and access of the data for as long as it maintains scientific utility.

The full SAS databases (not de-identified) associated with NAC Attack will be kept on secured computer systems maintained by the Study Chair and the Director of the Coordinating Center. Researchers may request limited access data sets and will need to enter into a data sharing agreement. The request must be made in written format providing information on: the purpose of the data request, the research questions to be studied, the plan of data analysis, procedures of data management that will protect the privacy of NAC Attack trial participants, key personnel who will have access to the limited data sets, and a description of data dissemination plan. Access to the NAC Attack database will be similar to these guidelines. Researchers requesting limited access data sets will bear the cost of their preparation.

##### 11.8.1. Genomic Data Sharing

NAC Attack will generate genetic sequencing data of all participants. Sharing of the genetic data will follow NIH's GDS Policy (NOT-OD-14-124, Effective Date January 25 2015, https://grants.nih.gov/grants/guide/notice-files/NOT-OD-14-124.html). Within 12 months after the study closure, genomic data will be submitted to dbGap as Controlled-access data. Database of Genotype and Phenotype (dbGaP) is an NIH maintained database to archive and distribute the results of studies that have investigated the interaction of genotype and phenotype in Humans.

NAC Attack genomic data would be made available for secondary research only after interested researchers have obtained approval from NIH to use the requested data for a particular project. Before data submission, the genomic data will be de-identified, and any identifiers protected by the Health Insurance Portability and Accountability Act (HIPAA) Privacy Rule will be removed. Since RP is a rare disease, all information that may allow identification of a study participant (e.g. the clinical site where the participant is recruited from) will be removed from the genomic data. Study IDs will be replaced by random unique codes. The key of the codes to Study IDs will be kept in the study Coordinating Center and will be submitted in the IPD data set uploaded in Vivli. This will allow researchers to retrieve clinical data to match with the genomic data of the same individuals.

###### 11.8.2. OCT Image Sharing

NAC Attack will generate retinal OCT images of all participants. Retinal images are protected health information under HIPPA. Sharing of the images will follow a plan of controlled public access to the images. Interested researchers should submit a request to the Executive Committee with descriptions of the intended use of the images. Once a request is approved by the Executive Committee, the researcher will be required to sign a data use agreement with the Johns Hopkins University Office of Research Administration and then will be provided with access to the images. The Study IDs and any information that may allow identification of a study participant will be removed from the images and replaced by random codes. The key of the codes to Study IDs will be kept in the study Coordinating Center and will be submitted in the IPD data set uploaded in Vivli. This will allow researchers to retrieve clinical data to match with the data of the same individuals.

###### 11.8.3. AOSLO Image and Data Sharing

###### 11.8.3.1. Protocol Sharing Plan

The AOSLO Reading Center will make available all of the image acquisition, processing, and analysis protocols. In addition, the training and certification procedures and test images used for certification will be made freely available as a downloadable package. These materials will be available on a website hosted by the affiliating institutions of the AOSLO PIs (Medical College of Wisconsin or University of California-San Francisco), or publishing the protocol in an open access journal.

###### 11.8.3.2. Image Sharing Plan

Prior to sharing, AOSLO images and data files (e.g., raw cone coordinates, cone metrics) will be de-identified by random codes. The key of the codes to Study IDs will be kept in the AOSLO Reading Center and will be submitted in the IPD data set uploaded in Vivli. These images can be used by other researchers to develop novel image processing and analysis tools, including machine learning or AI-based algorithms. The AOSLO dataset will represent one of the largest standardized AOSLO data sets openly available to the vision research community, so we will endeavor to share as openly as possible.

To facilitate this, the AOSLO Reading Center PIs have an existing IRB-approved research bank in the Advanced Ocular Imaging Program at the Medical College of Wisconsin. It provides a path for individuals in the vision community to gain access to a wealth of high-resolution imaging data that, until now, has been largely sequestered in individual labs/institutions. As part of the consent process, we will ask participants for permission to deposit their AOSLO images into this bank.

Having data and images in the bank allows for future, unspecified use after the trial is completed. The data will be made available 1 year after completion of the study to interested parties who shall obtain any regulatory or ethical approvals (from the appropriate IRB or research ethics committee) required by law or institutional policy before beginning any analysis. The parties shall comply with all applicable state/provincial, and local laws, regulations, codes and guidelines, including those regarding the handling, analyzing and reporting of analyses of data. Only after the interested parties submit their IRB approvals to the bank, they will be given access to the data and images in the bank.

### References

1. Bunker CH, Berson EL, Bromley WC, Hayes RP, Roderick TH. Prevalence of retinitis pigmentosa in Maine. *Am J Ophthalmol.* 1984;97:357-365.
2. Haim M. Epidemiology of retinitis pigmentosa in Denmark. *Acta Ophalmol Scand Suppl.* 2002;233:1-34.
3. Rivolta C, Sharon D, DeAngelis MM, Dryja TP. Retinitis pigmentosa and allied diseases: numerous diseases, genes, and inheritance patterns. *Hum Mol Genet.* 2002;11:1219-1227.
4. Boughman JA, Vernon M, Shaver KA. Usher syndrome: definition and estimated of prevalence from two high-risk populations. *J Chronic Dis.* 1983;36:595-603.
5. Athanasiou D, Agula M, Bellingham J, et al. The molecular and cellular basis of rhodopsin retinitis pigmentosa reveals potential strategies for therapy. *Prog Ret Eye Res.* 2018;62:1-23.
6. Cao SS, Kaufman RJ. Unfolded protein response. *Curr Biol.* 2012;22:R622-R626.
7. Bowes C, Li T, Danciger M, Baxter LC, Applebury ML, Farber DB. Retinal degeneration in the rd mouse caused by a defect in the beta subunit of rod cGMP-phosphodiesterase. *Nature.* 1990;347:677-680.
8. Chang B, Hawes NL, Pardue MT, et al. Two mouse retinal degerations caused by missense mutations in the beta-subunit of rod cGmP phosphodiestase gene. *Vision Res.* 2007;47(5):624-633.
9. McLaughlin ME, Sandberg MA, Berson EL, Dryja TP. Recessive mutations in the gene encoding the beta subunit of rod phosphodiesterase in patients with retinitis pigmenta. *Nat Genet.* 1993;4(2):130-134.
10. Farber DB, Lolley RN. Cyclic guanosine monophosphate: elevation in degenerating photoreceptor cells of the C3H mose retina. *Science.* 1974;186:449-451.
11. Iribarne M, Masai I. Neurotoxicity of cGMP in the vertegrate retina: from the initial research on rd mutant mice to zebrafish genetic approaches. *J Neurogen.* 2017;31:88-101.
12. Vithana EN, Abu-Safieh L, Allen MJ, et al. A human homolog of yeast pre-mRNA splicing gene, PRP31, underlies autosomal dominant retinitis pigmentosa on chromosome 19q13.4 (RP11). *Mol Cell.* 2001;8(2):375-381.
13. Buskin A, Zhu L, Chichagova V, et al. Disrupted alternative splicing for genes implicated in splicing and ciliogenesis causes PRPF31 retinitis pigmentosa. *Nat Commun.* 2018;9:4234.
14. Daiger SP, Sullivan LS, Bowne SJ. Genes and mutations causing retinitis pigmentosa. *Clin Genet.* 2013;84:132-141.
15. Yu DY, Cringle SJ, Su EN, Yu PK. Intraretinal oxygen levels before and after photoreceptor loss in the RCS rat. *Invest Ophthalmol Vis Sci.* 2000;41:3999-4006.
16. Usui S, Oveson BC, Lee SY, et al. NADPH oxidase plays a central role in cone cell death in retinitis pigmentosa *J Neurochem* 2009;110 1028-1037.
17. Shen J, Yan X, Dong A, et al. Oxidative damage is a potential cause of cone cell death in retinitis pigmentosa. *J Cell Physiol.* 2005;203(3):457-464.
18. Komeima K, Usui S, Shen J, Rogers BS, Campochiaro PA. Blockade of neuronal nitric oxide synthase reduces cone cell death in a model of retinitis pigmentosa. *Free Radic Biol Med.* 2008;45:905-912.
19. Komeima K, Rogers BS, Lu L, Campochiaro PA. Antioxidants reduce cone cell death in a model of retinitis pigmentosa. *Proc Natl Acad Sci USA.* 2006;103(38):11300-11305.
20. Komeima K, Rogers BS, Campochiaro PA. Antioxidants slow photoreceptor cell death in mouse models of retinitis pigmentosa. *J Cell Physiol.* 2007;213(3):809-815.

21. Lee SY, Usui S, Zafar AB, et al. N-acetylcysteine promotes long term survival of cones in a model of retinitis pigmentosa. *j Cell Physiol.* 2011;226:1843-1849

22. Ait-Ali N, Fridlich R, Millet-Puel G, et al. Rod-derived cone viability factor promotes cone survival by stimulating aerobic glycolysis. *Cell.* 2015;161:817-832.
23. Petit L, Ma S, Cipi J, et al. Aerobic glycolysis is essential for normal rod function and controls secondary cone death in retinitis pigmentosa. *Cell Rep.* 2018;23(9):2629-2642.
24. Campochiaro PA, Mir TA. The mechanism of cone cell death in retinitis pigmentosa. *Prog Retin Eye Res.* 2018;62(1):24-37.
25. Aldini G, Altomare A, Baron G, et al. N-Acetylcysteine as an antioxidant and disulphide breaking agent: the reasons why. *Free Radic Res.* 2018;52(7):751-762.
26. Prescott LF, Park J, Ballantyne A, Adriaenssens P, Proudfoot AT. Treatment of paracetamol (acetaminophen) poisoning with N-acetylcysteine. *Lancet.* 1977;2:432-434.
27. Smilkstein MJ, Knapp GL, Kulig KW, Rumack BH. Efficacy of oral N-acetylcysteine in the treatment of acetominophen overdose. Analysis of the national multicenter study (1976-1985). *N Eng J Med.* 1988;319:1557-1562.
28. Decramer M, Rutten-van Molken M, Dekhuijzen PN, et al. Effects of N-acetylcysteine on outcomes in chronic obstructive pulmonary disease (Bronchitis Randomized on NAC Cost-Utility Study, BRONCUS): a randomised placebo-controlled trial. *Lancet.* 2005;365(9470):1552-1560.
29. Tse HN, Raiteri L, Wong KY, et al. High-dose N-acetylcysteinine in stable COPD: the 1-year, double-blind, randomized, placebo-controlled HIACE study. *Chest.* 2013;144(1):106-118.
30. Zheng JP, Wen FQ, Bai CX, et al. Twice daily N-acetylcysteine 600 mg for exacerbations of chronic obstructive pulmonary disease (PANTHEON): a randomized, double-blind placebo-controlled trial. *Lancet.* 2014;2(3):187-194.
31. Demedts M, Behr J, Buhl R, et al. High-dose acetylcysteine in idiopathic pulmonary fibrosis. *N Engl J Med.* 2005;353(21):2229-2242.
32. Raghu G, Anstrom KJ, King TEJ, Lasky JA, Martinez FJ, Network IPFCR. Prednisone, Azathoprine, and N-acetylcysteine for pulmonary fibrosis. *N Engl J Med.* 2012;366(21):1968-1977.
33. Martinez FJ, de Andrade JA, Anstrom KJ, King TEJ, Raghu G, Network IPFCR. Randomized trial of acetycysteine in idipathic pulmoary fibrosis. *N Engl J Med.* 2014;370(22):2093-2101.
34. Campochiaro PA, Iftikhar M, Hafiz G, et al. Oral N-acetylcysteine improves cone function in retinitis pigmentosa patients in phase 1 trial. *J Clin Invest.* 2019;pii: 132990. doi: 10.1172/JCI132990.
35. Usui S, Komeima K, Lee SY, et al. Increased expression of catalase and superoxide dismutase 2 reduces cone cell death in retinitis pigmentosa. *Molec Ther.* 2009;17:778-786.
36. Xiong W, MacColl Garfinkel AE, Benowitz LI, Cepko CL. NRF2 promotes neuronal survival in neurodegeneration and acute nerve damage. *J Clin Invest.* 2015;125(4):1433-1445.
37. Kong X, Hafiz G, Wehling D, Akhlaq A, Campochiaro PA. Locus level changes in macular sensitivity in patient with retinitis pigmentosa treated with oral N-acetylcysteine. *Am J Ophthalmol.* 2020;Aug 11;S0002-9394(20)30421-9. doi: 10.1016/j.ajo.2020.08.002.
38. Roberts PA, Gaffney EA, Whiteley JP, Luthert PJ, Foss AJE, Byrne HM. Predictive Mathematical Models for the Spread and Treatment of Hyperoxia-induced Photoreceptor Degeneration in Retinitis Pigmentosa. *Invest Ophthalmol Vis Sci.* 2018;59(3):1238-1249.

39. Cai CX, Locke KG, Ramachandran R, Birch DG, Hood DC. A comparison of progressive loss of the ellipsoid zone (EZ) band in autosomal dominant and x-linked retinitis pigmentosa. *Invest Ophthalmol Vis Sci.* 2014;55(11):7417-7422.
40. Hariri AH, Zhang HY, Ho AC, et al. Quantifiation of the ellipsoid zone changes in retinitis pigmentosa using en face spectral domain-optical coherence tomography. *JAMA Ophthalmol.* 2016;134(6):628-635.
41. Tee JJL, Yang Y, Kalitzeos A, Webster aR, Bainbridge J, Michaelides M. Natural history study of retinal structure, progression, and symmetry using ellipzoid zone metrics in RPGR-associated retinopathy. *Am J Ophthalmol.* 2019;198(2):111-123.
42. Tee JJL, Yang Y, Kalitzeos A, Webster A, Bainbridge J, Michaelides M. Natural History Study of Retinal Structure, Progression, and Symmetry Using Ellipzoid Zone Metrics in RPGR-Associated Retinopathy. *Am J Ophthalmol.* 2019;198:111-123.
43. FDA. E9(R1) Statistical Principles for Clinical Trials: Addendum: Estimands and Sensitivity Analysis in Clinical Trials. https://www.fda.gov/regulatory-information/search-fda-guidance-documents/e9r1-statistical-principles-clinical-trials-addendum-estimands-and-sensitivity-analysis-clinical. 2019.
44. Csaky K, Ferris F, 3rd, Chew EY, Nair P, Cheetham JK, Duncan JL. Report From the NEI/FDA Endpoints Workshop on Age-Related Macular Degeneration and Inherited Retinal Diseases. *Invest Ophthalmol Vis Sci.* 2017;58(9):3456-3463.
45. O'Neal T, Luther E. Retinitis Pigmentosa. [Updated 2019 Apr 10]. In: Treasure Island (FL): StatPearls Publishing; 2019: https://www.ncbi.nlm.nih.gov/books/NBK519518/.
46. Sadda SR, Chakravarthy U, Birch DG, Staurenghi G, Henry EC, Brittain C. Clinical Endpoints for the Study of Geographic Atrophy Secondary to Age-Related Macular Degeneration. *Retina.* 2016;36(10):1806-1822.
47. Linder B, Dill H, Hirmer A, et al. Systemic splicing factor deficiency causes tissue-specific defects: a zebrafish model for retinitis pigmentosa. *Hum Mol Genet.* 2011;20(2):368-377.
48. Campochiaro PA, Iftikhar M, Hafiz G, et al. Oral N-acetylcysteine improves cone function in retinitis pigmentosa patients in phase I trial. *J Clin Invest.* 2020;130(3):1527-1541.
49. Birch DG, Cheng P, Duncan JL, et al. The RUSH2A Study: Best-Corrected Visual Acuity, Full-Field Electroretinography Amplitudes, and Full-Field Stimulus Thresholds at Baseline. *Transl Vis Sci Technol.* 2020;9(11):9.
50. Ramachandran R, C XC, Lee D, et al. Reliability of a Manual Procedure for Marking the EZ Endpoint Location in Patients with Retinitis Pigmentosa. *Transl Vis Sci Technol.* 2016;5(3):6.
51. Sujirakul T, Lin MK, Duong J, Wei Y, Lopez-Pintado S, Tsang SH. Multimodal Imaging of Central Retinal Disease Progression in a 2-Year Mean Follow-up of Retinitis Pigmentosa. *Am J Ophthalmol.* 2015;160(4):786-798 e784.
52. Iftikhar M, Kherani S, Kaur R, et al. Progression of Retinitis Pigmentosa as Measured on Microperimetry: The PREP-1 Study. *Ophthalmol Retina.* 2018;2(5):502-507.
53. Tee JJL, Carroll J, Webster AR, Michaelides M. Quantitative Analysis of Retinal Structure Using Spectral-Domain Optical Coherence Tomography in RPGR-Associated Retinopathy. *Am J Ophthalmol.* 2017;178:18-26.
54. Hariri AH, Zhang HY, Ho A, et al. Quantification of Ellipsoid Zone Changes in Retinitis Pigmentosa Using en Face Spectral Domain-Optical Coherence Tomography. *JAMA Ophthalmol.* 2016;134(6):628-635.
55. Wilding GE, Chandrasekhar R, Hutson AD. A new linear model-based approach for inferences about the mean area under the curve. *Stat Med.* 2012;31(28):3563-3578.
56. Dawson JD. Sample size calculations based on slopes and other summary statistics. *Biometrics.* 1998;54(1):323-330.

57. Writing Committee for the Diabetic Retinopathy Clinical Research N, Gross JG, Glassman AR, et al. Panretinal Photocoagulation vs Intravitreous Ranibizumab for Proliferative Diabetic Retinopathy: A Randomized Clinical Trial. *JAMA.* 2015;314(20):2137-2146.
58. DRCR. N. Anti-VEGF vs. Prompt Vitrectomy for VH From PDR - Full Text View - ClinicalTrials.gov. https://clinicaltrials.gov/ct2/show/NCT02858076.
59. FDA. Guidance for Industry E9 Statistical Principles for Clinical Trials. http://www.fda.gov/cder/guidance/index.htmorhttp://www.fda.gov/cber/guidelines.htm; . 1998.
60. Little R, Rubin D. *Statistical Analysis with Missing Data.* Wiley; 2002.
61. *SAS. SAS/STAT ® 14.1 User's Guide The MI Procedure. .* 2015.
62. Horton NJ, Lipsitz SR. Multiple imputation in practice: Comparison of software packages for regression models with missing variables. *American Statistician.* 2001;55(3):244-254.
63. von Hippel PT. How to Impute Interactions, Squares and Other Transformed Variables. *Sociological Methodology 2009, Vol 39.* 2009;39:265-291.
64. Campion WM. Multiple Imputation for Nonresponse in Surveys - Rubin,Db. *Journal of Marketing Research.* 1989;26(4):485-486.
65. Cabral T, Sengillo JD, Duong JK, et al. Retrospective Analysis of Structural Disease Progression in Retinitis Pigmentosa Utilizing Multimodal Imaging. *Sci Rep.* 2017;7(1):10347.
66. Hollander SA, Alsaleh N, Ruzhnikov M, et al. Variable clinical course of identical twin neonates with Alstrom syndrome presenting coincidentally with dilated cardiomyopathy. *Am J Med Genet A.* 2017;173(6):1687-1689.
67. Alshamrani AA, Raddadi O, Schatz P, et al. Severe retinitis pigmentosa phenotype associated with novel CNGB1 variants. *Am J Ophthalmol Case Rep.* 2020;19:100780.
68. Iftikhar M, Usmani B, Sanyal A, et al. Progression of retinitis pigmentosa on multimodal imaging: The PREP-1 study. *Clin Exp Ophthalmol.* 2018.
69. Birch DG, Locke KG, Wen Y, Locke KI, Hoffman DR, Hood DC. Spectral-domain optical coherence tomography measures of outer segment layer progression in patients with X-linked retinitis pigmentosa. *JAMA Ophthalmol.* 2013;131(9):1143-1150.
70. Colombo L, Montesano G, Sala B, et al. Comparison of 5-year progression of retinitis pigmentosa involving the posterior pole among siblings by means of SD-OCT: a retrospective study. *Bmc Ophthalmology.* 2018;18.
71. Takahashi VKL, Takiuti JT, Jauregui R, Lima LH, Tsang SH. Structural disease progression in PDE6-associated autosomal recessive retinitis pigmentosa. *Ophthalmic Genet.* 2018;39(5):610-614.
72. Kim YN, Song JS, Oh SH, et al. Clinical characteristics and disease progression of retinitis pigmentosa associated with PDE6B mutations in Korean patients. *Sci Rep.* 2020;10(1):19540.
73. Tee JJL, Yang Y, Kalitzeos A, et al. Characterization of Visual Function, Interocular Variability and Progression Using Static Perimetry-Derived Metrics in RPGR-Associated Retinopathy. *Invest Ophthalmol Vis Sci.* 2018;59(6):2422-2436.
74. Iftikhar M, Lemus M, Usmani B, et al. Classification of disease severity in retinitis pigmentosa. *Br J Ophthalmol.* 2019;103(11):1595-1599.
75. Yang Y. Sensitivity Analysis in Multiple Imputation for Missing Data. . 2014.
76. Permutt T. Sensitivity analysis for missing data in regulatory submissions. *Statistics in Medicine.* 2016;35(17):2876-2879.
77. Kahan BC, Morris TP. Analysis of multicentre trials with continuous outcomes: when and how should we account for centre effects? *Statistics in Medicine.* 2013;32(7):1136-1149.
78. Senn S. A note regarding 'random effects'. *Statistics in Medicine.* 2014;33(16):2876-2877.

79. Austin PC. An Introduction to Propensity Score Methods for Reducing the Effects of Confounding in Observational Studies. *Multivariate Behavioral Research.* 2011;46(3):399-424.
80. Birch DG, Fish GE. Rod ERGs in retinitis pigmentosa and cone-rod degeneration. *Invest Ophthalmol Vis Sci.* 1987;28(1):140-150.
81. Birch DG, Wen Y, Locke K, Hood DC. Rod sensitivity, cone sensitivity, and photoreceptor layer thickness in retinal degenerative diseases. *Invest Ophthalmol Vis Sci.* 2011;52(10):7141-7147.
82. Dmitrienko A, D'Agostino R, Sr. Traditional multiplicity adjustment methods in clinical trials. *Stat Med.* 2013;32(29):5172-5218.
83. Vickerstaff V, Omar RZ, Ambler G. Methods to adjust for multiple comparisons in the analysis and sample size calculation of randomised controlled trials with multiple primary outcomes. *BMC Med Res Methodol.* 2019;19(1):129.
84. Papi A, Zheng J, Criner GJ, Fabbri LM, Calverley PMA. Impact of smoking status and concomitant medications on the effect of high-dose N-acetylcysteine on chronic obstructive pulmonary disease exacerbations: A post-hoc analysis of the PANTHEON study. *Respir Med.* 2019;147:37-43.
85. Dmitrienko A, Muysers C, Fritsch A, Lipkovich I. General guidance on exploratory and confirmatory subgroup analysis in late-stage clinical trials. *Journal of Biopharmaceutical Statistics.* 2016;26(1):71-98.
86. Wang SJ, Hung HMJ. A Regulatory Perspective on Essential Considerations in Design and Analysis of Subgroups When Correctly Classified. *Journal of Biopharmaceutical Statistics.* 2014;24(1):19-41.
87. Dmitrienko A, D'Agostino RB. Multiplicity Considerations in Clinical Trials. *New England Journal of Medicine.* 2018;378(22):2115-2122.
88. Piantadosi S. *Clinical trials : a methodologic perspective.* 2005.
89. Ying GS, Maguire MG, Glynn RJ, Rosner B. Tutorial on Biostatistics: Longitudinal Analysis of Correlated Continuous Eye Data. *Ophthalmic Epidemiol.* 2021;28(1):3-20.
90. Zafarullah M, Li WQ, Sylvester J, Ahmad M. Molecular mechanisms of N-acetylcysteine actions. *Cell Mol Life Sci.* 2003;60(1):6-20.
91. Echeverri-Ruiz N, Haynes T, Landers J, et al. A biochemical basis for induction of retina regeneration by antioxidants. *Dev Biol.* 2018;433(2):394-403.
92. Oldham JM, Ma SF, Martinez FJ, et al. TOLLIP, MUC5B, and the Response to N-Acetylcysteine among Individuals with Idiopathic Pulmonary Fibrosis. *Am J Respir Crit Care Med.* 2015;192(12):1475-1482.
93. Moradi M, Mojtahedzadeh M, Mandegari A, et al. The role of glutathione-S-transferase polymorphisms on clinical outcome of ALI/ARDS patient treated with N-acetylcysteine. *Respir Med.* 2009;103(3):434-441.
94. Zhang JQ, Zhang JQ, Liu H, et al. Effect of N-acetylcysteine in COPD patients with different microsomal epoxide hydrolase genotypes. *Int J Chron Obstruct Pulmon Dis.* 2015;10:917-923.
95. Salamon S, Kramar B, Marolt TP, Poljsak B, Milisav I. Medical and Dietary Uses of N-Acetylcysteine. *Antioxidants (Basel).* 2019;8(5).

### NAC Attack

**PROTOCOL ADDENDUM TO Protocol Version 2.0-Version Date 2023-07-13**

**Date: 2024-01-01**

**RE: Protocol 4.12.6 Emergency Unmasking.** This section is changed to below:

Site investigator is responsible for the medical care of trial participants. Under emergency situation when site investigator feels a participant has an SAE that necessities immediate unmasking of the participant's treatment group to manage the event, site investigator may contact the Coordinating Center to receive the unmasked treatment group information. Coordinating Center unmasked staff will provide the information to site investigator in a timely manner. This process should be recorded in the Unmasking Form. Communication during this process between the Coordinating Center and the clinical site will ensure masking remain in place for all other participants.

### NAC Attack

#### PROTOCOL ADDENDUM TO Protocol Version 2.0-Version Date 2023-07-13

#### Date: 2024-10-01

#### RE: Protocol Addendum to be added as 6.1.2.4

Although EZ loss is expected to be a degenerative process, it is unknown whether EZ width at a follow-up visit may be greater than that at baseline due to measurement error or due to intervention effect. The AAC is the cumulative loss of EZ width overtime. Its calculation needs to consider if there was gain of EZ width during follow-up (i.e. negative loss). Section 6.1.2.3 derives the AAC calculation if $y_1$ is increased compared to $y_0$. Here we provide an algorithm that derives the general AAC calculation without assuming whether a follow-up EZ-width measurement is greater than $y_0$.

Let $k = 1, \cdots K$ index follow-up visits.

For $k = 1$, the area of loss between baseline $t_0$ and $t_1$ $area[1] = 1/2 \cdot | y_0 - y_1 | \cdot t \cdot Sign(y_0 - y_1)$.

For $k > 1$ and if $(y_0 - y_{k-1}) \cdot (y_0 - y_k) \geq 0$, the area between $t_{k-1}$ and $t_k$ is a trapezoid. It is either a loss or gain depending on whether or not $y_{k-1}$ and $y_k$ are greater than $y_0$. The area of loss, $area[k] = 1/2 \cdot (| y_0 - y_{k-1} | + | y_0 - y_k |) \cdot t \cdot Sign(y_0 - y_k)$.

For $k > 1$ and if $(y_0 - y_{k-1}) \cdot (y_0 - y_k) < 0$, this means either $y_{k-1}$ or $y_k$ was greater than $y_0$. Thus there is one area of gain of EZ width between $t_{k-1}$ and $t_k$. The overall area of loss between $t_{k-1}$ and $t_k$,

$$area[k] = \frac{t}{2} \cdot \frac{| y_0 - y_{k-1} |^2 \cdot sign(y_0 - y_{k-1})}{| y_0 - y_{k-1} | + | y_0 - y_k |} + \frac{t}{2} \frac{| y_0 - y_k |^2 \cdot sign(y_0 - y_k)}{| y_0 - y_{k-1} | + | y_0 - y_k |}.$$

Then the AAC at the last visit $K$ is $\sum_{k=1}^{K} area[k]$.

### NAC Attack

#### PROTOCOL ADDENDUM TO Protocol Version 2.0-Version Date 2023-07-13

#### Date: 2025-02-20

#### RE: Protocol Addendum to be added as 6.4.1 and 6.4.2 to Protocol Version 2.0

##### 6.4.1. Final Study Sample Size after Sample Size Re-estimation

Protocol Section 6.4 describes Plan for Sample Size Re-estimation to determine the final study sample size. In brief, after the study has enrolled 100 participants, masked data would be used to refine the parameter assumptions used in sample size estimation. The sample size re-estimation and rate of enrollment would be reviewed by the study Data and Safety Monitoring Committee (DSMC) for decision on the final study sample size.

The NAC Attack DSMC held its first in-person meeting on January 13 2025 to review study progress and data. Enrollment data accrued as of December 29 2024 had surpassed the 100 participants required in the plan for sample size re-estimation specified in Protocol Section 6.4 and were used to refine the parameter assumptions for sample size estimation.

During the meeting, the DSMC reviewed the following information:

- Parameter values assumed for the original SS estimation versus values estimated based on the study data accrued as of December 29 2024 (Table 1)
- Study enrollment progress since starting enrollment in November 2023
- Feasibility of completing sample size target and projected timeline of completion
- Study medication availability, logistics status and downstream plan

**Based on the review, the DSMC, in consultation with representatives from the funder (the National Eye Institute of the U.S. National Institutes of Health) and the Study Leadership, recommended the study final sample size to be N=483, an increase of 45 from the original sample size of 438.**

Table 1 below summarizes the parameter assumptions in the sample size calculation: the estimates for Parameters 1-3 using accrued study data suggest that the original assumptions of these parameters are reasonable. For example, the between-eye correlation coefficient of the primary outcome variable was assumed to be 0.7. This is the upper limit of the 95% confidence interval (CI) of the parameter estimate based on the accrued data of ~70 eyes for which the primary outcome measurements were available. Similarly, the 86% of bilateral enrollment assumed originally is in the 95% CI of the parameter estimate using the 295 participants randomized by December 29 2024. Therefore, the original *effective sample size* of 417 (Protocol 6.3.2.2), i.e. before accounting for loss to follow-up (LFUP) remains reasonable.

The original sample size further assumed an additional 21 (Protocol 6.3.2.2), i.e. ~5% participants would be loss to follow-up (LFUP) (~ a rate of LFUP of 0.11/100 person-months), thus a total sample size of 438. However, as of December 29 2024, 6 participants had withdrawn from the study (out of 295 participants accruing 1830.33 person-months). The observed rate of LFUP in the study by Dec 29

2024 was ~0.33/100 person-months, about two times higher than the assumed rate.

Therefore, the DSMC considered it sensible to refine the assumption of this parameter to add 40-45 subjects. Further considering enrollment feasibility and logistics capacity, the final study sample size was decided to be 483.

*Table 1 Original parameter assumptions vs. parameter estimates based on study data accrued before the DSMC meeting on January 13 2025. alpha=0.049. Power = 80%.*

| | Parameter | Original Assumptions | Estimates from current trial data based on data available as of December 29 2024 |
|---|---|---|---|
| **1.** | **Cohen's D (Effect size as % difference between arms over standard deviation of difference)** | *29% (Protocol 6.3.1.2)* | N/A as this parameter is a target effect and determined by relevance to clinical management of the disease. |
| **2.** | **Between-eye correlation coefficient of AAC** | *0.7 (Protocol 6.3.1.2)* | 0.44 (95%CI 0.09-**0.70**) (estimated using baseline and Month 9 (M9) data of ~70 eyes available) |
| **3.** | **% of Bilateral enrollment** | *86% (Protocol 6.3.1.3)* | 85.4% (80.9%-89%)<br>(estimated based on 295 participants randomized) |
| **4.** | **Number of loss of follow-up (LFUP) by M45** | N=21 (Protocol 6.3.2.2)<br><br>i.e. if constant rate of LFUP: 0.107/100 person-months | Actual rate of LFUP by Dec. 29 2024: 0.328/100 pm ~2 times higher than assumed. |

##### 6.4.2. Study Disjunctive Power After Sample Size Re-estimation

Following Protocol 6.4, after sample size re-estimation, the correlation coefficients between each pair of the outcome variables should be estimated and the disjunctive power of hypothesis testing of the primary and secondary outcome measures using Hochberg method would be estimated following the method in Section 6.5.3.2.

The parameter assumptions in Section 6.5.3.2 largely remain to hold. For example, the effective sample size N=417 is not changed after sample size re-estimation. The specific parameter of "Correlation coefficient between any pair of outcomes of the left (right) eyes" was originally assumed to be 0.2 to 0.8. Table 2 below summarizes pairwise correlation coefficients of the primary and secondary outcome variables estimated using data accrued as of December 29 2024. Table 2 suggests that the pairwise correlation between outcome measures would be unlikely to be too high (e.g. greater than 0.60) but may be weakly negative. Thus based on Table 2, disjunctive power is further estimated by expanding the range of this parameter to -0.4 to 0.2. Estimated disjunctive power results are presented in *Figure 1*: The results are very similar to the original estimation results in Section 6.5.3.2. If at least one of the secondary outcomes has a treatment effect of 20%, the disjunctive power is greater than 80%.

*Table 2 Correlation coefficients between each pair of primary and secondary outcome variables estimated using data available as of Dec 29 2024. These estimates inform the estimation of the disjunctive power of the hypothesis testing of the primary and secondary outcome measures using Hochberg method to account for multiplicity.*

| **Outcome variable** | **With outcome variable** | **Available No. of study eyes as of Dec 29 2024** OD (Right eyes) | **Correlation coefficient (95% confidence interval)** OD (Right eyes) | **Available No. of study eyes as of Dec 29 2024** OS (Left eyes) | **Correlation coefficient (95% confidence interval)** OS (Left eyes) |
|---|---|---|---|---|---|
| EZ-width AAC (estimated using baseline and M9 data of ~70 eyes available by Dec 29 2024) | MMS change between baseline and M9 | 32 | -0.21 (-0.52, 0.15) | 36 | 0.07 (-0.27, 0.39) |
| Macular mean sensitivity (MMS) change between baseline and M9 | BCVA change between baseline and M9 | 55 | -0.06 (-0.21, 0.32) | 55 | -0.13 (-0.38, 0.14) |
| Best corrected visual acuity (BCVA) change between baseline and M9 | EZ-width AAC | 33 | 0.02 (-0.32, 0.36) | 37 | 0.26 (-0.07, 0.54) |

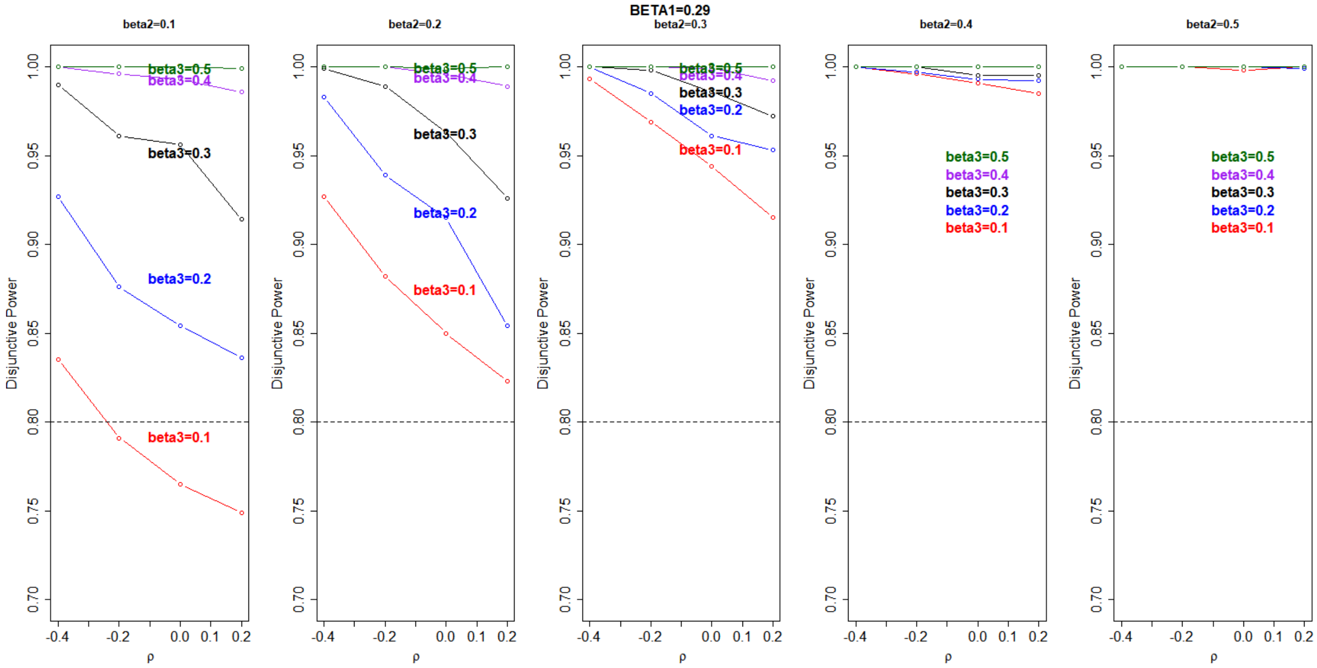

*Figure 1 Disjunctive power of the planned hypothesis testing of the primary and secondary out-comes using Hochberg procedure, expanding the assumed range of the correlation coefficient between any pair of outcomes of the left (right) eyes to -0.4 to 0.2 now. The other parameter assumptions remain as those in the original estimation in Section 6.5.3.2.*
