## Supplemental 3 list of sites for "Protocol for NAC Attack, a phase-3, multicenter randomized, parallel, double masked, placebo controlled trial evaluating the efficacy and safety of oral N-acetylcysteine (NAC) in patients with retinitis pigmentosa"

**Supplemental Material -3 List of NAC Attack clinical and AOSLO imaging sites.** There are 31 clinical sites enrolling NAC Attack participants.

*: the site also participates in the AOSLO sub-study where NAC Attack participants who consented for the AOSLO sub-study receives AOSLO imaging at the site.

^: the site participates in the AOSLO sub-study where NAC Attack participants who consented for AOSLO receive AOSLO imaging at a nearby study AOSLO imaging center.

ǂ: the site does not enroll trial participants but provides AOSLO Imaging to a nearby study clinical site.

| Country | State/Province | Institution |
|---|---|---|
| Austria | Styria | Medical University of Graz, Department of Ophthalmology |
| Canada | Quebec | McGill University, The Research Institute of the McGill University Health Center |
| Germany | Baden-Württemberg | University of Tübingen, Department für Augenheilkunde |
| Netherlands | Gelderland | Radboud University, Radboud University Medical Centre |
| Netherlands | North Holland | University of Amsterdam, Amsterdam Medical Center |
| Switzerland | Basel | Universitätsspital Basel, Eye Clinic |
| United Kingdom | England | University College London, Moorfields Eye Hospital* |
| United States | California | University of California - Davis, Department of Ophthalmology & Vision Science |
| United States | California | University of Southern California, Keck School of Medicine |
| United States | California | University of California - San Francisco, Department of Ophthalmology * |
| United States | California | Stanford University, Byers Eye Institute |
| United States | Florida | Vitreo Retinal Associates |
| United States | Florida | University of Florida – Jacksonville, UF Health Jacksonville |
| United States | Florida | University of Miami, Bascom Palmer Eye Institute |
| United States | Georgia | Emory University, Emory Eye Center |
| United States | Illinois | Northwestern University, Feinberg School of Medicine |
| United States | Illinois | University of Illinois at Chicago, College of Medicine |
| United States | Iowa | University of Iowa, Carver College of Medicine |
| United States | Maryland | Johns Hopkins University, Wilmer Eye Institute |
| United States | Massachusetts | Harvard University, Mass. Eye and Ear |
| United States | Michigan | University of Michigan, Kellogg Eye Center |
| United States | Minnesota | University of Minnesota, Department of Ophthalmology and Visual Neurosciences |
| United States | Minnesota | Mayo Clinic, Department of Ophthalmology |
| United States | Oklahoma | University of Oklahoma, Dean McGee Eye Institute |
| United States | Pennsylvania | University of Pennsylvania, Scheie Eye Institute * |
| United States | Tennessee | Vanderbilt University, Vanderbilt Eye Institute |
| United States | Texas | Retina Foundation of the Southwest ^ |
| United States | Utah | University of Utah, Moran Eye Center |
| United States | Washington | University of Washington, Department of Ophthalmology * |
| United States | Wisconsin | University of Wisconsin – Madison, McPherson Eye Research Institute ^ |
| United States | Wisconsin | Medical College of Wisconsin, The Eye Institute * |
| United States | Texas | University of Houston, College of Optometry ǂ |
