## Supplemental 4 List of Study Group 7.21.2025 for "Protocol for NAC Attack, a phase-3, multicenter randomized, parallel, double masked, placebo controlled trial evaluating the efficacy and safety of oral N-acetylcysteine (NAC) in patients with retinitis pigmentosa"

July 02 2025

### STUDY CHAIR'S OFFICE

**Wilmer Eye Institute, Johns Hopkins University School of Medicine**

**600 N. Wolfe St. Maumenee 815**
**Baltimore, MD, 21287, United States.**

Peter Campochiaro, MD. Study Chair
Gulnar Hafiz, Coordinator
Muhammad Jehanzeb Khan, Research Fellow, Technician – images, MAIA, FST

Zainab Rustam, Research Fellow, Technician – images, MAIA, FST

Dagmar Wehling, Coordinator
Imad Shaikh, Coordinator

### COORDINATING CENTER

**Wilmer Eye Institute, Johns Hopkins University School of Medicine**

**600 N. Wolfe St. Woods 155**
**Baltimore, MD, 21287, United States.**

Xiangrong Kong, PhD. Director, Senior Biostatistician of Coordinating Center
Kurt Dreger, Informatics Director of Coordinating Center
Folahan Ibukun, MBBS, MPH, Research Fellow, Associate Director of Coordinating Center
Bhavishya Pancharathi, Project Coordinator
Chrisanthe Papapavlou, Project Coordinator

Yanzhao Wang, Project Coordinator

Barbara Hawkins, PhD. Co-Director of Coordinating Center
Sheila West, PhD. Co-Director of Coordinating Center

### MEDICAL MONITOR

Dana Frank, MD.

### OCT READING CENTER

**Duke Eye Center**
**Duke University School of Medicine**
**2351 Erwin Road**
**Durham, NC. 27710, United States.**

| | |
|---|---|
| Glenn J. Jaffe | Co-PI |
| Sina Farsiu | Co-PI |
| April Flora | Project Manager |
| Kevin Choy | Graduate Student developing AI algorithm for EZ area |
| John Choong | Senior Reader |
| Chris Forbes | Senior Reader |
| Murad Maksumov | Senior Reader |
| Sumru Onal | Senior Reader |
| Hoang Nguyen | Senior Reader |

### AOSLO READING CENTER (ARC)

**University of California, San Francisco**
**Department of Ophthalmology School of Medicine 675 Nelson Rising Lane, #54A San Francisco CA 94158**

| | |
|---|---|
| Jacque Duncan, MD | AOSLO Site Co-PI/ARC Co-Director |
| Jessica Wong, BS, MS | AOSLO reading center grader, AOSLO imaging expert |

**Medical College of Wisconsin**
**Eye Institute 925 N. 87th St.**
**Milwaukee, WI 53226**

| | |
|---|---|
| Joseph Carroll, PhD | AOSLO Site PI/ARC Co-Director |
| Joseph Kreis, BA | ARC Research Associate & Senior Grader |
| Morgan Connaughton, MS | AOSLO Reading Center Data Management |
| Brian Higgins, BS | AOSLO Imaging and Data Analysis |

### CLINICAL SITES

**<u>Dean McGee Eye Institute at University of Oklahoma Health Sciences Center 608</u>**
**Stanton L. Young Blvd.**
**Oklahoma City, OK 73104**

| | |
|---|---|
| Lea Bennett | PI |
| Grace Trigler | Coordinator |

**<u>Emory University</u>**
**Department of Ophthalmology**
**1365B Clifton Road NE, Suite B2400**
**Atlanta, GA 30322**

| | |
|---|---|
| Nieraj Jain | PI |
| Judith Tribe | Study Coordinator |
| Donna Leef | Backup Coordinator |
| Daniel Starostenko | Ophthalmic Photographer |

July 02 2025


**<u>Harvard Medical School</u>**
**Massachusetts Eye and Ear**
**Department of Ophthalmology**
**Harvard Medical School**
**325 Cambridge Street, Floor 7**
**Boston, Massachusetts, 02114 USA**


| | |
|---|---|
| Rachel M. Huckfeldt, MD, PhD | PI |
| Jason Comander, MD, PhD | Sub-I |
| Ahmad Al-Moujahed, MD, PhD | Sub-I |
| Kirill Zaslavsky, MD, PhD | Sub-I |
| Yuki Wiland | Project Manager |
| Christopher F. Barile | Study Coordinator |
| Fabian Perez, MS | Study Coordinator |
| Howard Chang, MS | Study Coordinator |
| Mirjana Nordmann, PhD | Program Manager |


**<u>Johns Hopkins University</u>**
**The Wilmer Eye Institute**
**Johns Hopkins University School of Medicine 600**
**N. Wolfe Street**
**Baltimore, MD 21210**


| | |
|---|---|
| Peter A. Campochiaro | PI |
| Dagmar Wehling | Coordinator |
| Gulnar Hafiz | Coordinator |
| Imad Shaikh | Coordinator |
| Muhammad Jehanzeb Khan | Technician obtaining images, MAIA, and FST |
| Zainab Rustam | Technician obtaining images, MAIA, FST |
| Maria Chairez Miranda | Study medication reconciliation |
| Mandeep Singh | Co-investigator |
| Ishrat Ahmed | Co-investigator |
| Mira Sachdeva | Co-investigator |
| Tin Yan Alvin Liu | Co-investigator |


**<u>Mayo Clinic</u>**
**200 First St SW**
**Rochester MN 55905**


| | |
|---|---|
| Raymond Iezzi, MD | PI |
| Brittni Scruggs, MD, PhD | Sub-I |
| Fadi Shaya | Clinical Research Coordinator (CRC) |
| Suzanne Wernimont | CRC |
| Tessa Holtegaard | CRC |
| Laura Taylor | CRC |

July 02 2025

| | |
|---|---|
| Kelsey Howard | CRC |

**McGill University**

**McGill University Health Center**

**1001 Decarie Blvd**

**Montreal, Quebec, Canada H4A 3J1**

| | |
|---|---|
| Robert K. Koenekoop | Principal Investigator |
| Joanie Gonthier | Clinical Research Coordinator |
| Daphné Doucet | Clinical Research Coordinator |
| Christine Gannon | Clinical Research Coordinator |
| Luan Tran | Clinical Research Supervisor |
| Kerry McKenna | Clinical Research Supervisor |
| Glenda Vargas | Ophthalmology Technician |
| Goreth Xethalis | Ophthalmology Technician |
| Mona Hijazi | Orthoptist |
| Connie Pham | Ophthalmology Technician |

**Medical College of Wisconsin**

**Eye Institute**

**925 N 87th Street**

**Milwaukee, WI 53226**

| | |
|---|---|
| Thomas Connor, MD | PI |
| Katie McKenney, MS | Primary Coordinator |
| Phyllis Summerfelt | Backup Coordinator |
| Christopher Langlo, MD, PhD | Sub-I |
| Joon-Bom Kim, MD | Sub-I |
| Erin Breggeman, BS | Coordinator |

**Medical University of Graz**

**Department of Ophthalmology**

**Auenbruggerplat 4**

**8036 Graz**

July 02 2025

| | |
|---|---|
| Rupert Strauss | PI |
| Laura Posch-Pertl | Sub -I |
| Tamara Pichler-Seitlinger | Study Coordinator |
| Ulrike Geweßler | Study Coordinator |
| Kathrin Vollnhofer | Sub -I |
| Marina Casazza | Sub -I |

**Northwestern University**
**259 East Erie Street, Suite 1500**
**Chicago, IL 60611**

| | |
|---|---|
| Safa Rahmani MD MS | PI |
| Fred Collision OD | Sub-PI |

**Radboud University**
**Stichting Radboud Universitair Medisch Centrum**
**Geert Grooteplein 10**
**P.O. Box 9101**

| | |
|---|---|
| Suzanne Yzer, PhD, MD | PI |
| Milan Phan, MD | Sub-PI |

**Retina Foundation of the Southwest**
**9600 North Central Expressway, Suite 200**
**Dallas, TX 75231**

| | |
|---|---|
| David Birch, PhD | PI |
| Kaylie Jones, MS | Coordinator |

**Stanford University**
**Byers Eye Institute at Stanford 2452**
**Watson Ct.**
**Palo Alto, CA 94303**

| | |
|---|---|
| Vinit B. Mahajan | PI |
| Theodore Leng, MD, FACS | Sub-PI |
| Lorraine Almeda | CRC |
| Kenny Trang | Research Photographer |
| Aarushi Kumar | Back up CRC |

**UC Davis Health Eye Center 4860**
**Y Street**
**Sacramento, CA 95817**

**6500 HB Nijmegen**

| | |
|---|---|
| Franca Hartgers, PhD | Study coordinator |

July 02 2025

| | |
|---|---|
| Paul Sieving, MD PhD | PI |
| Denise Macias | CRC |
| Michael Lien | CRC |
| Gregory Kizer | Imaging Technician |
| Megan Hughes-Salaber | Imaging Technician |
| Ziana John D'Souza | Research Assistant |

**University College London**
**Moorfields Eye Hospital**
**162  City Road EC1V 2PD**

| | |
|---|---|
| Michel Michaelides | PI |
| Nancy Aychoua | Sub-PI |

**University Hospital Basel**
**University Hospital Basel Eye Clinic**
**Mittlere Strasse 91**
**4031  Basel, Switzerland**

| | |
|---|---|
| Josep Callizo, MD | Principal Investigator (2025.07.01- present) |
| Kristina Pfau, MD | Principal Investigator (2024-05-06-2025-06-30) |
| Maximilian Pfau, MD | Sub-Investigator |
| Chrysoula Gabrani | Sub-Investigator |
| Ursula Hall | Study Coordinator |
| Daniela Hauenstein | Study Coordinator |
| Nils Schaerer | Study Coordinator |
| Petra Rossouw | Study Coordinator |
| Prof. Nicolas Feltgen | Clinic Director at the University Hospital Basel, Eye Clinic, Co- Director IOB |

**University of Amsterdam** **Amsterdam UMC**
**loc. VUmc, De Boelelaan 1117**
**1081  HV Amsterdam, the Netherlands**

| | |
|---|---|
| Camiel J.F. Boon, MD, PhD | PI |
| Roselie M.H. Diederen, MD, PhD | Sub-investigator |

**University of California San Francisco**
**Department of Ophthalmology**
**UCSF Wayne and Gladys Valley Center for Vision  490 Illinois Street**
**San Francisco, CA 94143 -4081**

| | |
|---|---|
| Jacque L. Duncan, MD | Principal Investigator |
| Erika Reyes, OD | Study Optometrist and technician |
| Beth Cutrer, PhD | Clinical research coordinator supervisor |

| | |
|---|---|
| Nicholas Bobbitt, BS, MS | Clinical research coordinator |
| Julia Wessman, BA | Assistant Clinical Research Coordinator |

**<u>University of Florida – Jacksonville</u>**
**College of Medicine, Jacksonville, FL**

| | |
|---|---|
| Sandeep Grover, MD | PI |
| Ghulam Hamdani, MBBS | Research Manager |
| Shivani Kochhar, MD | Coordinator |
| Carla Thomas | Coordinator |
| Bharani Krishna Mynampati, PHD | Coordinator |
| Diti Patel, MD | Coordinator |
| Cheryl Shoup | Coordinator |

**<u>University of Houston * AOSLO imaging only</u>**
**College of Optometry**
**J. Davis Armistead Building**
**4401 Martin Luther King Boulevard**
**Houston, TX 77204-2020**

| | |
|---|---|
| Jason Porter, PhD | Site PI |
| Alexander W. Schill, PhD | Site Co-I |
| Kaitlyn A. Sapoznik, OD, PhD | Site Co-I |

**<u>University of Illinois at Chicago</u>**
**Department of Ophthalmology and Visual Sciences**
**1855 W Taylor St**
**Chicago IL 60612**

| | |
|---|---|
| Jennifer Lim, MD/FARVO | PI |
| Robert Hyde, MD/PhD | Sub-I |
| Jianrong Sheng | Primary Study Coordinator |
| Patrizia Chavero | Study coordinator |
| Scott Wang | Study coordinator |

**<u>University of Iowa</u>**
**Health Care Department of Ophthalmology**
**200 Hawkins Drive**
**Iowa City, Iowa 52242**

| | |
|---|---|
| Arlene V. Drack, MD | PI |
| Ian Han, MD | Sub-I |
| Alina Dumitrescu, MD | Sub-I |
| Giulia Del Valle | Sub-I |
| Mary McCormick | Coordinator |
| Lacy Flanagan | Coordinator |
| Jennifer Stark | Visual Acuity |
| Jody Troyer | Imaging |

| | |
|---|---|
| Meghan Menzel | Imaging |
| Megan Smith | FST |
| Wanda Pfeifer | FST |
| Paul Swihart | Microperimetry |
| Cathrine Jones | Microperimetry |
| Jill Oulman | Microperimetry |

**University of Miami**
**Bascom Palmer Eye Institute**
**900 NW 17th Street**
**Miami, FL 33136**

| | |
|---|---|
| Byron L. Lam, MD | PI |
| Carlos E. Mendoza-Santiesteban | Sub-I |
| Tamara Juvier-Riesgo | Research Coordinator |
| Adriana Padilla Drada | Research Coordinator |
| Nisha K. Phogat | Research Coordinator |
| Patrick Valera Fernandez | Research Coordinator |

**University of Michigan Kellogg Eye Center**
**1000 Wall Street**
**Ann Arbor, MI 48105**

| | |
|---|---|
| Abigail Fahim, MD, PhD | PI |
| Callie Gordon, COA, CCRP | Primary Study Coordinator |
| Courtney Snyder, COA, BHSA | Backup Study Coordinator |
| Sonya Cosby, CRA | Ophthalmic Imaging Specialist |

**University of Minnesota**
**516 Delaware St. SE, 9th Floor**
**Eye Clinic Ophthalmology and Visual Neurosciences**
**Minneapolis, MN 55455**

| | |
|---|---|
| Meaghyn Kramer | Clinical Research Coordinator |
| Sandra Montezuma, MD | Principal investigator |
| Christian Curran, MD | Sub Investigator |
| Richard Sather lll, MD | Study Team Member |
| Christian Larson, OD | Study Team Member |

**University of Pennsylvania**
**Center for Hereditary Retinal Degenerations (CHRD)**
**Scheie Eye Institute**
**Department of Ophthalmology**
**University of Pennsylvania 51**
**N 39th St.**
**Philadelphia, PA 19104**

| | |
|---|---|
| Alexandra V. Garafalo | Lead coordinator |
| Artur V. Cideciyan, PhD | PI |
| Tomas S. Aleman, MD | Co-Investigator |
| Vivian Wu | Coordinator |
| Iryna Viarbitskaya | Coordinator |
| Mariejel Weber | Coordinator |
| Robert Russell | Coordinator |
| Kelsey M. Parchinski | Photographer |
| Arlene J. Santos | Technician |
| Rebecca J. Kim | Technician |
| Andrew Billek | Photographer |
| Alexander Sumaroka, PhD | Photographer |

**University of Southern California**
**USC Keck School of Medicine Ophthalmology**
**Department 1450 San Pablo Street 4th Floor**
**Los Angeles, CA. 90033**

| | |
|---|---|
| Hossein Ameri, MD | Site PI |
| Mariana Edwards | Research Coordinator |
| Kimberly Rodriguez | Research Coordinator |
| Hossein Ameri MD | Principal Investigator |
| Mosies Tellez | Imager |
| Leobardo Cortez | Imager |
| Emilio Ortega | Imager |
| Bernarda Duenas | Clinical Lab Technician |
| Charlene Chiu | Pharmacist |
| Akiko Maloney | Pharmacist |
| Daniel Barajas | Imager |
| Magaly Gastelum | Clinical Lab Technician |
| Paul Chung | Pharmacist |
| Travis Rock | Pharmacy Technician |
| Ramon Moreno | Pharmacy Technician |
| Sun Young Lee | |

**University of Tübingen**
**Department for Ophthalmology**
**University Eye Clinic,**
**Eberhard Karls University Tübingen,**
**Tübingen, Germany**

| | |
|---|---|
| Laura Kuehlewein, MD | Site PI |
| Katarina Stingl, MD | Sub-I |
| Tobias Peters, MD | Clinical Trial Center Director |

July 02 2025

| | |
|---|---|
| Melanie Ziegler, PhD | Clinical Trial Center |

**<u>University of Utah</u> Moran Eye Center**
**65 Mario Capecchi Dr.**
**Salt Lake City, UT 84132**

| | |
|---|---|
| Paul Bernstein, MD, PhD | Principal Investigator |
| Kimberley Wegner, BS | Lead Study Coordinator |
| Karen Gutierrez, COA | VAE/microperimetry |
| Marcela Pasaye | Lead Ophthalmic Imager |

**<u>University of Washington</u>**
**Karalis Johnson Retina Center**
**750 Republican Street 1st Floor**
**Seattle, WA 98109**

| | |
|---|---|
| Jennifer R Chao | Lead PI |
| Alyssa C Bonnell | Sub-I |
| Yewlin E Chee | Sub-I |
| Christopher R Fortenbach | Sub-I |
| Cecilia S Lee | Sub-I |
| Debarshi Mustafi | Sub-I |
| Lisa C Olmos de koo | Sub-I |
| Russell N Van Gelder | Sub-I |
| Palash Bharadwaj | Study Coordinator |
| Akanksha Singh | Lead Study coordinator |
| Ramkumar Sabesan | Sub-I |
| Benjamin J Wendel | Study Coordinator |
| Simona Vuletic | Study coordinator |
| Teng Liu | Study coordinator |
| Jennifer Huey | Study coordinator |

**<u>University of Wisconsin Madison</u> 2880 University Ave**
**Madison, WI 53705**

| | |
|---|---|
| Kimberly Stepien, MD | PI |
| Nicole Boone, BS | Study Coordinator |
| Nickie Stangel, CCRC | Study Coordinator |

**<u>Vanderbilt Eye Institute</u>**
**2311 Pierce Ave Nashville, TN 37232**

| | |
|---|---|
| Lindsay Veach | Study coordinator |
| Milam Brantley | Principal investigator |
| Sapna Gangaputra | Sub-investigator |

July 02 2025

**<u>Vitreo Retinal Associates, P.A.</u>**
**4340 W Newberry Rd, Suite 202**
**Gainesville FL 32607**

| | |
|---|---|
| Christine N. Kay, MD | PI |
| Siva Iyer, MD | Sub-PI |
| Jing Zhang | CRC |
| Hailey Ahrens | CRC |
| Brianna Nguyen | CRC |
| Zadie Dulaney | CRC |
